# Emergence and spread of SARS-CoV-2 lineages B.1.1.7 and P.1 in Italy

**DOI:** 10.1101/2021.03.24.21254277

**Authors:** Francesca Di Giallonardo, Ilaria Puglia, Valentina Curini, Cesare Cammà, Iolanda Mangone, Paolo Calistri, Joanna C.A. Cobbin, Edward C. Holmes, Alessio Lorusso

## Abstract

Italy’s second wave of SARS-CoV-2 has hit hard, with more than 3 million cases and over 100,000 deaths, representing an almost ten-fold increase on the numbers reported by August 2020. Herein, we present the analysis of 6,515 SARS-CoV-2 sequences sampled in Italy between 29^th^ January 2020 and 1^st^ March 2021 and show how different lineages emerged multiple times independently despite lockdown restrictions. Virus lineage B.1.177 became the dominant variant in November 2020, when cases peaked at 40,000 a day, but since January 2021 this is being replaced by the B.1.1.7 ‘variant of concern’. In addition, we report a sudden increase in another documented variant of concern – lineage P.1 – from December 2020 onwards, most likely caused by a single introduction into Italy. We again highlight how international importations drive the emergence of new lineages and that genome sequencing should remain a top priority for ongoing surveillance in Italy.

## Introduction

COVID-19, caused by infection with severe respiratory coronavirus 2 (SARS-CoV-2), has had a devastating global impact, with an estimated 116 million cases and 2.5 million deaths^1^. Of these reported infections, more than 78% have occurred during the so-called ‘second wave’ of the epidemic between September 2020 – March 2021, greatly outnumbering the 25 million cases and ∼800k deaths reported globally prior to 31 August 2020^2^. This second wave of infections is also characterised by the emergence of numerous new lineages of SARS-CoV-2, three of which are commonly referred to global ‘variants of concern’ – B.1.1.7, B.1.351, and P.1^3^. The B.1.1.7 lineage was first detected in the United Kingdom in late September 2020^4^ and is characterised by 17 polymorphisms across the genome including the N501Y amino acid mutation and a two amino acid deletion at positions 69 and 70 of the spike protein^5^. The B.1.351 lineage was first reported in South Africa and shares similar substitutions to B.1.1.7 including N501Y but not the 69/70 deletion^6^, while the P.1 lineage was first identified in Brazil and shares two substitutions with B.1.351 – E484K and N501Y in the spike protein^4^. All of these lineages have seemingly replaced previous circulating variants in their geographic regions and have spread to other countries in Europe, the Americas, and Asia^7-9^. There is also considerable concern about the possibility of reinfection with these new lineages due to reduced cross-protective immunity^10-12^, which may also have implications for vaccine efficacy^13,14^. Thus, increased surveillance to document and understand the spread of different SARS-CoV-2 lineages is of upmost importance.

The first cases of SARS-CoV-2 in Italy were reported in January 2020 and the country was hit hard with a peak of >6,500 cases a day in late March 2020. Italy was also the first European country to impose a nation-wide lockdown from 9^th^ March to the 18^th^ May 2020. In retrospect, this first wave was of relatively low magnitude compared to the huge number of infections reported between September 2020 and March 2021. Indeed, during this second wave, Italy experienced a maximum of ∼40,000 cases a day in mid-November 2020 (Fig. 1a), and an estimated 3.1 million cases and ∼100,000 deaths have been reported to date (8^th^ March 2021)^15^. Beginning in October 2020, a colour-coded system was established to restrict the mobility of residents in each administrative region in Italy, depicting increasing the levels of restriction from yellow to red. In addition, in attempt to reduce viral transmission during the holiday season, further restrictions to social mobility and on possible sources of infection (schools, restaurants, etc.) were put in place for the whole of Italy between 21^st^ December 2020 and the 6^th^ January 2021. On 20^th^ December 2020, the Italian Minister of Health signed an order prohibiting all air flights from/to United Kingdom. On 16^th^ January 2021 the same restriction was applied to the air flight from/to Brazil.

**Fig. 1.**
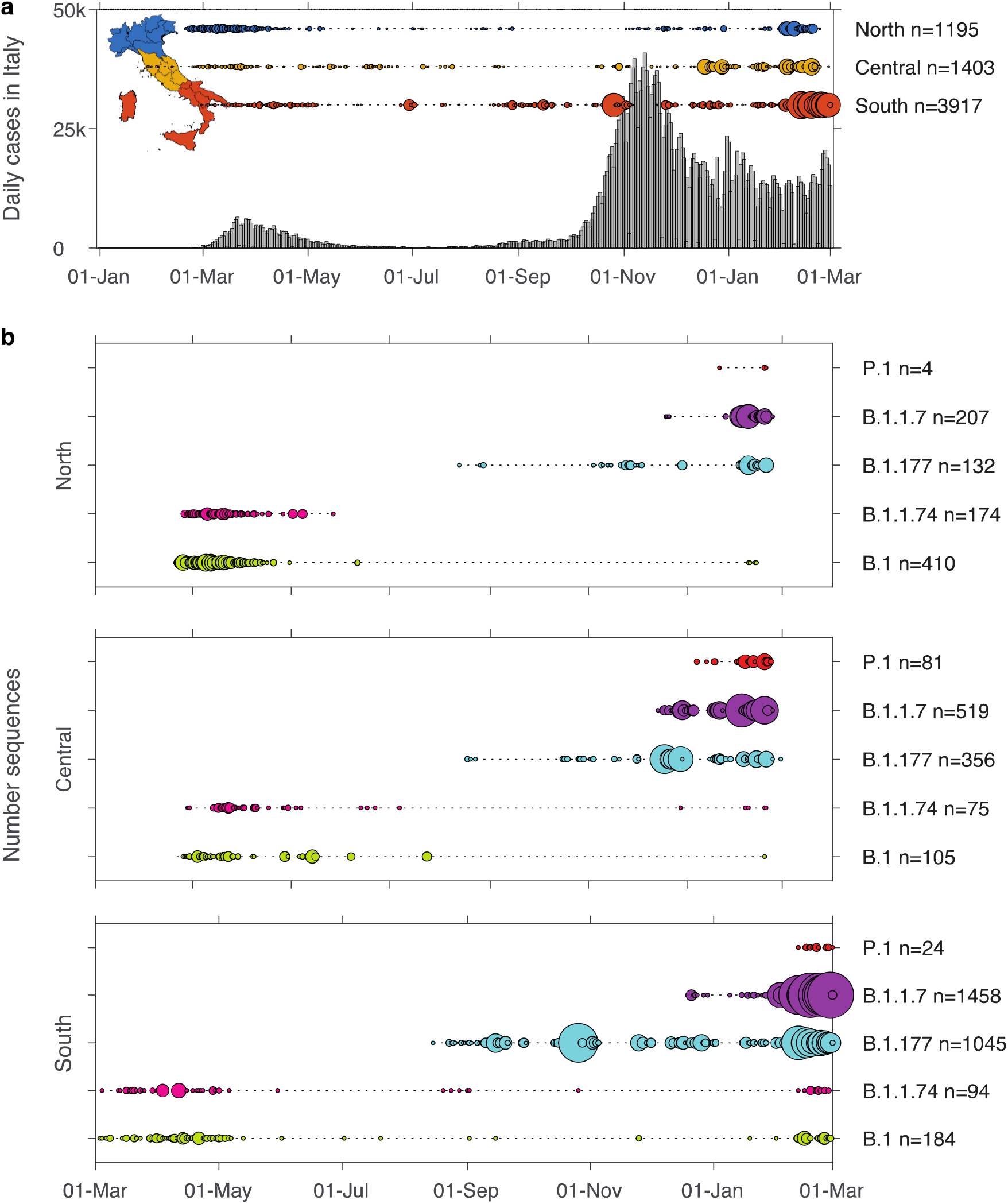
Number of SARS-CoV-2 genomes sampled over time in Italy. **a** Bars show the number of cases reported in Italy between 1^st^ January 2020 – 1^st^ March 2021. Horizontally aligned circles show the number of sequences sampled for each day in North (blue), Central (yellow), South (red) Italy. The size of the circle is equivalent to the total number of sequences (max = 256). Italian regions are coloured according to their macro areas: blue = North (Valle d’Aosta, Piemonte, Liguria, Lombardia, Emilia-Romagna, Veneto, Friuli-Venezia-Giulia, and Trentino-Alto Adige), yellow = Central (Lazio, Marche, Toscana, Umbria, and Abruzzo), red = South (Puglia, Basilicata, Calabria, Campania, Molise, Sicilia, and Sardegna). **b** Number of sequences sampled in Italy for the lineages B.1. B.1.1.74, B.1.177, B.1.1.7, and P.1 for North (top), Central (middle), and South (bottom) Italy. The total number of sequences per day (circles) and the time range between first and most recent sequence (dotted line) is shown (max = 146).

We previously showed that virus importation associated with travel, followed by local transmission, were key drivers of viral spread during the first wave of SARS-CoV-2 in Italy^16^. After a year into the pandemic, lockdown restrictions have proven effective in reducing virus transmission and limiting geographic spread^17^, although international travel is still a major source for the introduction of new lineages into Italy^18^. Here we show how the B.1.1.7 and P.1 variants entered Italy before the implementation of border closures and that these two variants have subsequently spread rapidly across the country.

## Results

### Emerging lineages in Italy

Our analysis included 6,515 Italian SARS-CoV-2 sequences covering a time span between 29^th^ January 2020 to 1^st^ March 2021. These comprised 103 different lineages, with B.1 (11%), B.1.1.74 (5%), B.1.1.7 (34%), and B.1.177 (24%) the most commonly identified. Sequence sampling was not equal between North, Central, and South Italy, as a large proportion of SARS-CoV-2 sequence data from North Italy was only available for the beginning of the pandemic (March – May 2020) and again the most recent months (January – March 2021) (Fig.1a). Despite this, the time range of data availability overlaps between these three macro areas, minimising the effect of sampling bias.

Lineages B.1 and B.1.1.74 both appeared in Italy during early 2020 and were replaced by lineage B.1.177 during the second half of 2020 (Fig. 1b). Lineage B.1 was the first reported in all three macro areas on 20^th^ February, 24^th^ February, and 3^rd^ March in North, Central, and South Italy, respectively. A similarly narrow time period of introduction was observed for lineage B.1.1.74 which first appeared on 24^th^ February, 27^th^ February, 4^th^ March 2020 in North, Central, and South Italy, respectively. This latter lineage persisted in Central and South Italy until February 2021, although at low levels, while no sequences from this lineage were sampled in North Italy after 27^th^ May 2020. Subsequently, lineage B.1.177 emerged in August in all three macro areas becoming the dominant variant in the second wave, with the earliest documentation in North Italy (13^th^ August), followed by South Italy on 15^th^ August and Central Italy on August 18^th^.

Although these data show that different lineages of SARS-CoV-2 were first reported within a short time period throughout the country, we also identified region-specific sampling biases with unusual abundances of sequences obtained on individual days (Supplementary Data S1). These sampling ‘peaks’ are most likely the result of targeted sampling after an outbreak. For example, 193 sequences were sampled on 26^th^ October 2020 alone, of which 104 were B.1.177 all sampled in the region Campania in South Italy. Also, in this region, 42 B.1.1.187 sequences were sampled on 29^th^ June 2020 alone, and which represents 51% of all sequence data available for this lineage in Italy. Notably, according to GISAID, this lineage has only been sampled in the UK (n=14) and Japan (n=1), and South Italy (n=81).

### Lineages of global concern - B.1.1.7, P.1, and B.1.351

At the time of writing (8^th^ March 2021), a total of 2,184 B.1.1.7, 109 P.1, and 8 B.1.351 sequences sampled in Italy were available for analysis and had been identified in all three Italian macro areas. The B.1.1.7 lineage appeared in all three areas within a five-day period, first in Central Italy (14^th^ December) and five days later in both the North and South (19^th^ December for both lineages) (Fig. 1b). The first P.1 sequences were identified on 7^th^ January 2021 in Central Italy in travellers returning from Brazil ^19^. This lineage was subsequently found on 21^st^ January and 12^th^ February in the North and South Italy, respectively.

Notably, the Italian B.1.1.7 sequences were distributed across the global phylogeny indicating multiple independent introductions into the country (Fig. 2). In addition, we observed numerous Italian-specific transmission clades (i.e. extended transmission chains), although most (n=73, 78%) were area specific, the largest containing 104 infections all sampled in South Italy (Fig. 2). Of the remaining clades (n=22), 82% contained infections from two and 18% from all three macro areas. For 73% (n=16) of these, the clade in question comprised ≥80% of infections from a single area only and only a small number of infections from a second or third macro area. For example, one clade with 64 sequence comprised 63 from Central Italy, one from South Italy and none from North Italy. Similarly, another clade contained 76 infections from North Italy and one from Central Italy. In addition, we identified six clades with more inter-area mixing, including one example consisting of 35 infections (10 North, 12 Central, 13 South). Notably, this is the only example of prolonged transmission between macro areas for this time period (Fig. 2).

**Fig. 2.**
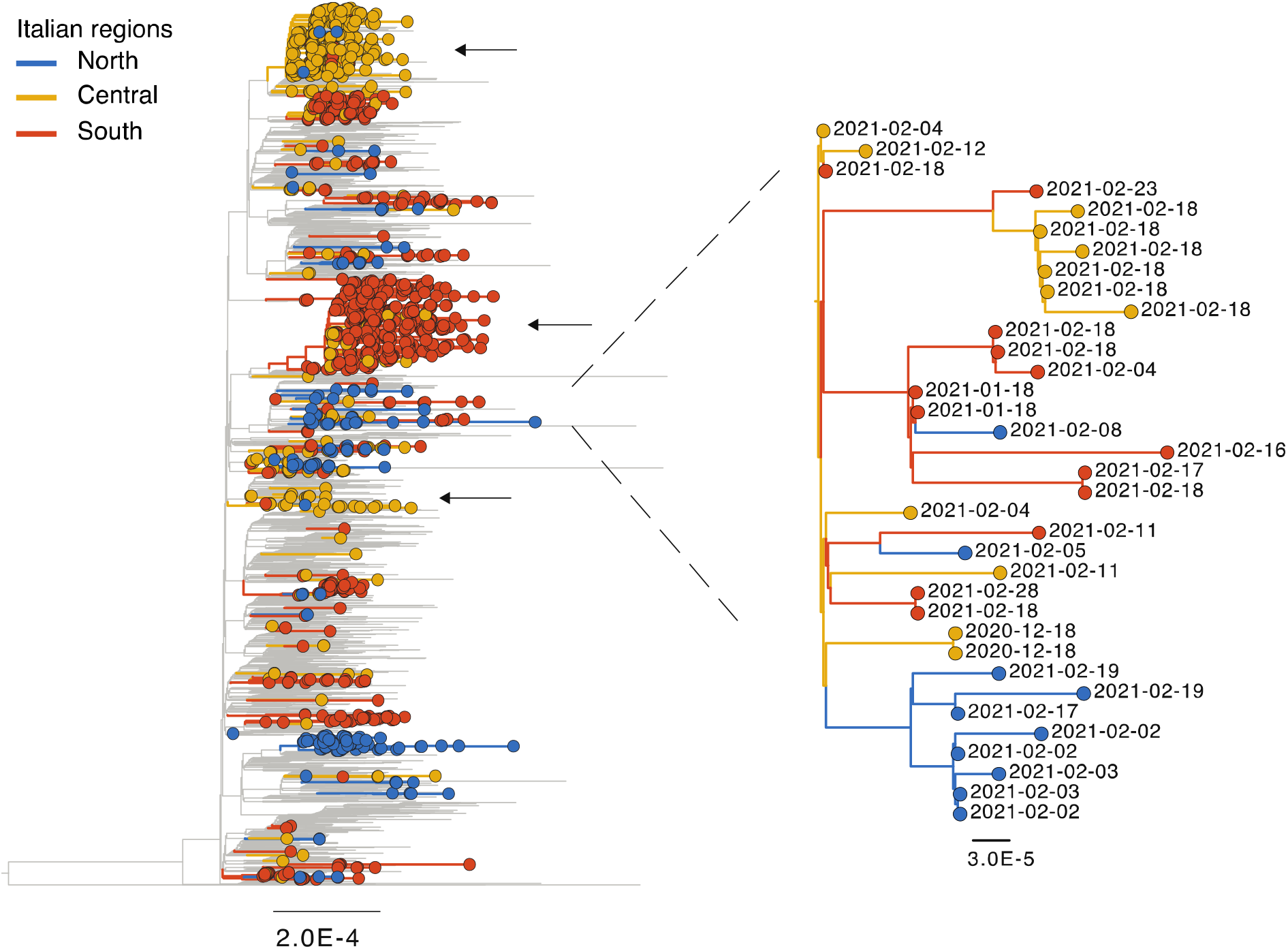
Genomic epidemiology of SARS-CoV-2 lineage B.1.1.7. Maximum likelihood phylogeny of SARS-CoV-2 full genome sequences. Due to the large amount of data for B.1.1.7 (>100,000 sequences), a random subset of 3,000 B.1.1.7 sequences sampled globally were incorporated (2,900 after data cleaning). Branches are coloured according to the macro area: grey = global, blue = North Italy, yellow = Central Italy, red = South Italy. Branch length is scaled according to the number of nucleotide substitutions per site. Lineage B.1.351 and P.1 were used as an outgroup (B.1.351=EPI_ISL_660190, P.1=EPI_ISL_833137). Arrows indicate examples of large Italian clades with limited inter-region transmission. One clade with increased inter-region transmission is shown enlarged on the right.

Three infections of the P.1 lineage were identified in international travellers on 18^th^ January in Central Italy. Fortunately, these did not lead to ongoing local transmission (Fig. 3). However, infections of P.1 were also identified on 7^th^ January, and which formed a distinct clade (clade I) within the P.1 global phylogeny distinguished by a single T->C synonymous mutation in ORF1ab (nt 13,577 / aa 4,526 Ref EPI_ISL_833137). In contrast to B.1.1.7, almost all of Italian P.1 sequences (n=104) fell within this single node which also contained 10 sequences from Germany (Fig. 3). Within this clade I, 61 Italian sequences (clade II) contained an additional distinct synonymous mutation in the spike gene (spike nt 3,357 / aa 1,118 Ref EPI_ISL_833137), and 43 of these Italian sequences (clade III) also contained a non-synonymous mutation in the spike leading to an S813N mutation (spike nt 2,437 / aa 813 Ref EPI_ISL_833137). This S813N mutation is located between two fusion peptides and not in the receptor binding domain where the E484K and N501Y mutations are located (Supplementary Data S2). P.1. viruses with the mutation were first sampled on 13^th^ January 2021 in Central Italy and the most recent occurrence was found in an infection sampled on 1^st^ March. To date, the mutation has only been found in infections from Central and South Italy. Of note, one additional sequence (EPI_ISL_1169907, Central Italy, 19-Feb) outside of both clade II and III also contained this amino acid substitution (Fig. 3).

**Fig. 3.**
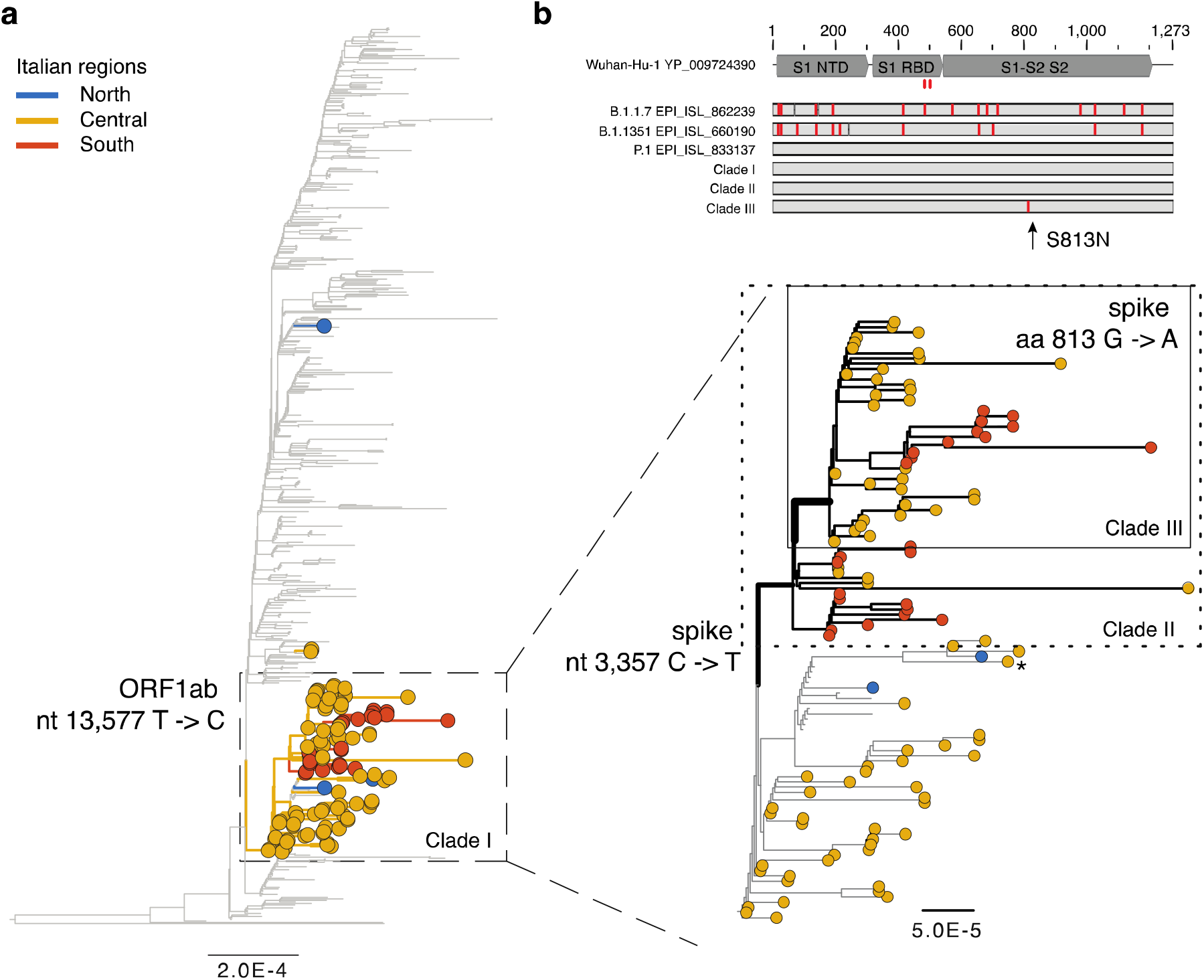
Genomic epidemiology of SARS-CoV-2 lineage P.1. **a** Maximum likelihood phylogeny of SARS-CoV-2 full genome sequences (n=503 global, n=111 Italian). Lineage B.1.351 was used as an outgroup (EPI_ISL_660190). Branch lengths are scaled according to the number of nucleotide substitutions per site and are coloured according to geographic macro area: grey = global, blue = North Italy, yellow = Central Italy, red = South Italy. The node containing an T to C mutation in ORF1ab is marked with a box. **b (bottom)** Enlargement of this node and additional clades and their nucleotide or amino acid substitutions are indicated. Note, a single sequence containing also the S813N mutation but falling outside the main clade is marked with an asterisk. Branches without a tip circle represent sequences from Germany. **(top)** Schematic view of the spike protein alignment for different reference sequences as well as the clade containing the S813N substitution. The protein structural regions are marked: S1 NTD = N-terminal domain of the S1 subunit, S1 RBD = receptor binding domain of S1, S1-S2 S2 = S1/S2 cleavage region and S2 fusion subunit. A detailed view of the protein alignment can be found in Supplementary Data S2.

Finally, five of the eight B.1.351 sequences were scattered across the global phylogeny, confirming that these are independent infections associated with returned travellers (30^th^ January and 11^th^ February in North Italy n=2, 23^rd^ February in Central Italy n=1, 19^th^ and 26^th^ February in South Italy n=2, Supplementary Data S3). However, we also found B.1.351 sequences from North Italy sampled on 11^th^ and 13^th^ February 2021 who formed a phylogenetic cluster, suggestive of some local transmission.

## Discussion

We document the complex patterns of virus transmission and lineage turnover during the second wave of SARS-CoV-2 in Italy, with an analysis based on >6,500 sequences sampled from January 2020 – March 2021. First, we observed the replacement of B.1 and B.1.1.74 by B.1.177, the latter being the most sampled lineage during the second wave. Second, and more notably, we observed the more recent appearance of the emerging ‘variants of concern’ B.1.1.7 and P.1, with the former being the most sampled lineage during January and February 2021.

Our phylogenetic analysis confirms extensive virus transmission within North, Central, and South Italy, but only limited transmission between these regions, consistent with lockdown restrictions imposed in December 2020 that prohibited travel between regions. However, the limited sequence data available makes it challenging to accurately identify the directionality of transmission and lineage spread across the country. Nevertheless, our data shows how different lineages appeared almost simultaneously in North, Central, and South Italy (i.e., within a five-day time frame), suggesting a complex pattern of multiple introductions into the different macro areas.

We focused our analysis on lineages B.1.1.7, B.1.351, and P.1 as these have been the subject of considerable discussion and concern. In the case of B.1.351, we only found one instance of local transmission in mid-February 2021, with no evidence for additional ongoing transmission. In contrast, we found strong evidence for local ongoing transmission of both the B.1.1.7 and P.1 lineages, with the former potentially replacing the previous dominant strain B.1.177. Of note, from 8^th^ March 2021 onwards, B.1.1.7 represented on average 50% of all sequences sampled. This is compatible with observations that B.1.1.7 has enhanced transmissibility^5^ and has become the dominant variant in numerous other European countries, e.g. Denmark^20^.

After Brazil, Italy contains the second highest number of P.1 infections reported to date^3^. This variant was first reported in Manaus (Brazil), which was hit hard by the first wave of SARS-CoV-2 with an estimated 76% of the population infected^21^. Thus, the surge of P.1 in this area strongly implies the possibility of re-infection^9^. Our molecular epidemiological analysis provided strong evidence for a single introduction of P.1 into Italy, followed by extensive local transmission. As expected, our phylogenetic analysis depicts importation of the P.1 variant into Italy, although from a yet unknown source, followed by possible onward spread from Italy to Germany. Importantly, within the Italian network we also found a unique, clade defining amino acid substitution - S813N. Although the function of this mutation is currently unknown, it is likely unrelated to virus binding and hence does not represent an immune escape variant although this should be assessed further^13,22^. No clade defining amino acid substitution was found for the B.1.1.7 lineage in spike gene of Italian sequences.

In conclusion, we depict the rapid emergence and replacement of new lineages in Italy despite imposed lockdown restriction, suggesting that the disease management policies employed were insufficient to halt the spread of emerging variants. Notably, both B.1.1.7 and P.1 spread across Italy before international border closers were imposed for the UK and Brazil. This highlights the importance of a rapid and inclusive vaccine roll-out on a global scale.

## Methods

### Sequence data

All Italian sequences of SARS-CoV-2 available on the GISAID EpiCov™ database were downloaded (8^th^ March 2021, acknowledgment Table S1). Sequences without a complete date of collection were removed. As per our previous study, cases were grouped according to geographical non-administrative macro areas in North (n=1,195), Central (n= 1,403), and South Italy (n= 3,917)^16^. SARS-CoV-2 lineages were assigned to each sequence using the Pangolin COVID-19 Lineage Assigner tool v2.0.7 (github.com/cov-lineages/pangolin). All available global sequence data for B.1.351 and P.1 was also downloaded from the GISAID EpiCov™ database. Due to the large amount of data available for B.1.1.7 (>170,000), a random subset of 3,000 global sequence data was used in combination with all available Italian sequence data (this was extracted from the global SARS-CoV-2 tree^23^).

### Phylogenetic analysis

Separate nucleotide sequence alignments were constructed for the B.1.1.7, P.1, and B.1.351 lineages. A representative sequence for each lineage was used as a reference and as an outgroup for tree rooting (B.1.1.7=EPI_ISL_862239, B.1.351=EPI_ISL_660190, P.1=EPI_ISL_833137). Three separate alignments were performed using MAFFT implementing the L-INS-I algorithm and manually inspected for accuracy using Geneious Prime^(r)^ 2021.1.1 (https://www.geneious.com)^24^. Full genome sequences with >5% ambiguity and no exact sampling date were removed. The final data sets comprised: P.1 n=614 (Italy n=111), B.1.351 n=2,317 (Italy n=8), and B.1.1.7 n=4,361 (Italy n=1,461). A maximum likelihood tree was estimated for each lineage using IQ-TREE implementing the following: Hasegawa-Kishino-Yano nucleotide substitution model with a gamma distributed rate variation among sites (HKY+Γ) and an ultrafast bootstrap method (1000 repetitions)^25,26^.

## Data Availability

Data used in this study was obtained from the GISAID EpiCovTM database.

## Author Contributions

The project was conceptualized by F.D.G, E.C.H. and A.L. The manuscript was written by F.D.G, E.C.H. and A.L. Sequence data was generated by I.P., V.C., M.M., P.C., A.L. Analyses were performed by F.D.G, J.C.A.C. Results were visualized by F.D.G.

## Acknowledgments

We are thankful to all individuals who submit data to GISAID. We thank the Pango team for their efforts in maintaining a freely available tool for lineage classification. The authors would like to acknowledge all doctors, nurses, technicians, medical staff, administrators, food and cleaning service workers, pharmacists, and all other members of the COVID-19 diagnostic group at IZSAM.

## Supplementary Information

Fig. S1: Changing frequency of SARS-CoV-2 lineages sampled over time in Italy. Fig. S2: Amino acid alignment of the SARS-CoV-2 spike protein. Fig. S3: Molecular epidemiology of global SARS-CoV-2 lineage B.1.351. Table S1: GISAID EpiCov™ acknowledgment.

## References

1 WHO. Weekly operational update on COVID-19 - 8 March 2021. (2021).

2 WHO. Weekly epidemiological update - 31 August 2020. (2020).

3 Rambaut, A. et al. A dynamic nomenclature proposal for SARS-CoV-2 lineages to assist genomic epidemiology. Nat. Microbiol. 5, 1403-1407 (2020). doi:10.1038/s41564-020-0770-5

4 CDC. Science brief: emerging SARS-CoV-2 variants, https://www.cdc.gov/coronavirus/2019-ncov/more/science-and-research/scientific-brief-emerging-variants.html. Accessed 1 Mar 2021

5 Davies, N. G. et al. Estimated transmissibility and impact of SARS-CoV-2 lineage B.1.1.7 in England. Science (2021). doi:10.1126/science.abg3055

6 Tegally, H. et al. Emergence and rapid spread of a new severe acute respiratory syndrome-related coronavirus 2 (SARS-CoV-2) lineage with multiple spike mutations in South Africa. medRxiv (2020). doi:10.1101/2020.12.21.20248640

7 O’Toole, A. et al. Tracking the international spread of SARS-CoV-2 lineages B.1.1.7 and B.1.351/501Y-V2, https://virological.org/t/tracking-the-international-spread-of-sars-cov-2-lineages-b-1-1-7-and-b-1-351-501y-v2/592. Accessed 1 Mar 2021

8 Walensky, R.P., Walke, H.T. & Fauci, A.S. SARS-CoV-2 variants of concern in the United States challenges and opportunities. JAMA (2021). doi:10.1001/jama.2021.2294

9 Sabino, E. C. et al. Resurgence of COVID-19 in Manaus, Brazil, despite high seroprevalence. Lancet 397, 452–455 (2021). doi:10.1016/S0140-6736(21)00183-5

10 Resende, P. C. et al. Spike E484K mutation in the first SARS-CoV-2 reinfection case confirmed in Brazil, 2020, https://virological.org/t/spike-e484k-mutation-in-the-first-sars-cov-2-reinfectioncase-confirmed-in-brazil-2020/584. Accessed 1 Mar 2021

11 Naveca, F. et al. SARS-CoV-2 reinfection by the new Variant of Concern (VOC) P.1 in Amazonas, Brazil, https://virological.org/t/sars-cov-2-reinfection-by-the-new-variant-of-concern-voc-p-1-in-amazonas-brazil/596. Accessed 1 Mar 2021

12 Wibmer, C. K. et al. SARS-CoV-2 501Y.V2 escapes neutralization by South African COVID-19 donor plasma. (2021). doi:10.1101/2021.01.18.427166

13 Williams, T. C. & Burgers, W. A. SARS-CoV-2 evolution and vaccines: cause for concern? Lancet Respir. Med. (2021). doi:10.1016/S2213-2600(21)00075-8

14 Xie, X. et al. Neutralization of SARS-CoV-2 spike 69/70 deletion, E484K and N501Y variants by BNT162b2 vaccine-elicited sera. Nat. Med. (2021). doi:10.1038/s41591-021-01270-4

15 Statistiche coronavirus. Statistiche coronavirus in Italia, https://statistichecoronavirus.it/coronavirus-italia/. Accessed 8 Mar 2021

16 Di Giallonardo, F. et al. Genomic epidemiology of the first save of SARS-CoV-2 in Italy. Viruses 12 (2020). doi:10.3390/v12121438

17 Schlosser, F. et al. COVID-19 lockdown induces disease-mitigating structural changes in mobility networks. Proc. Natl. Acad. Sci. U. S. A. 117, 32883–32890 (2020). doi:10.1073/pnas.2012326117

18 Swadi, T. et al. Genomic evidence of in-flight transmission of SARS-CoV-2 despite predeparture testing. Emerg. Infect. Dis. 27, 687–693 (2021). doi:10.3201/eid2703.204714

19 Delli Compagni, E. et al. Genome sequence of three SARS-CoV-2 P.1 strains iIdentified from patients returning from Brazil to Italy. Microbiol. Resour. Announc. (2021). doi:accepted for publication

20 Danish Covid-19 Genome Consortium. Genomic overview of SARS-CoV-2 in Denmark, https://www.covid19genomics.dk/statistics. Accessed 15 Mar 2021

21 Buss, L. F. et al. Three-quarters attack rate of SARS-CoV-2 in the Brazilian Amazon during a largely unmitigated epidemic. Science 371, 288–292 (2021). doi:10.1126/science.abe9728

22 Wise, J. Covid-19: The E484K mutation and the risks it poses. BMJ 372, 359 (2021). doi:10.1136/bmj.n359

23 Lanfear, R. A global phylogeny of hCoV-19 sequences from GISAID, https://www.gisaid.org/. Accessed 10 Mar 2020

24 Kuraku, S., Zmasek, C. M., Nishimura, O. & Katoh, K. aLeaves facilitates on-demand exploration of metazoan gene family trees on MAFFT sequence alignment server with enhanced interactivity. Nucleic Acids Res. 41, W22–28 (2013). doi:10.1093/nar/gkt389

25 Trifinopoulos, J., Nguyen, L. T., von Haeseler, A. & Minh, B. Q. W-IQ-TREE: a fast online phylogenetic tool for maximum likelihood analysis. Nucleic Acids Res. 44, W232–235 (2016). doi:10.1093/nar/gkw256

26 Nguyen, L. T., Schmidt, H. A., von Haeseler, A. & Minh, B. Q. IQ-TREE: a fast and effective stochastic algorithm for estimating maximum-likelihood phylogenies. Mol. Biol. Evol. 32, 268–274 (2015). doi:10.1093/molbev/msu300

