## Supplementary Material for "Emergence and spread of SARS-CoV-2 lineages B.1.1.7 and P.1 in Italy"

**Supplementary Figure S1. Changing frequency of SARS-CoV-2 lineages sampled over time in Italy.**

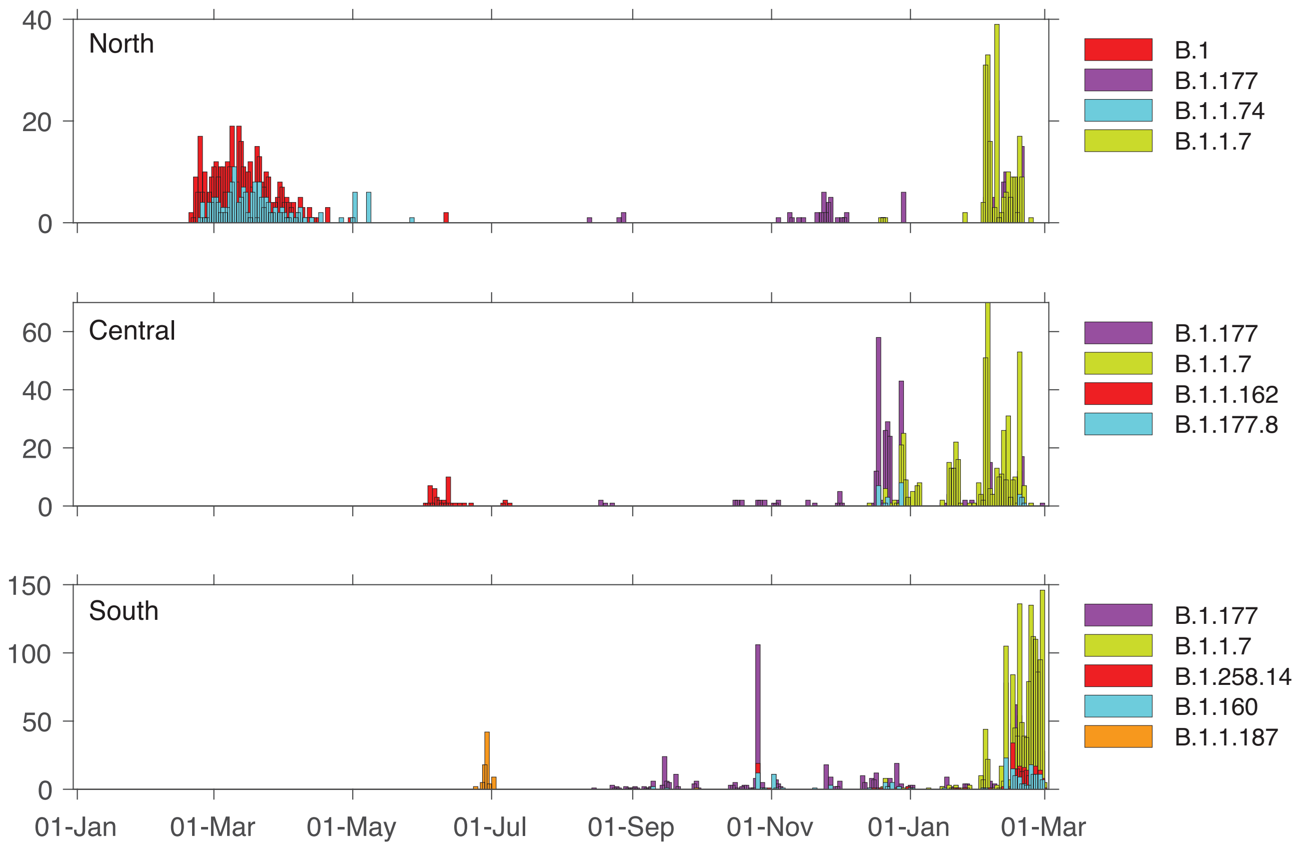


Bars show the Number of cases reported between 01 January 2020 – 1 March 2021 in North (top), Central (middle), and South Italy (bottom).

**Supplementary Figure S2. Amino acid alignment of the SARS-CoV-2 spike protein.**

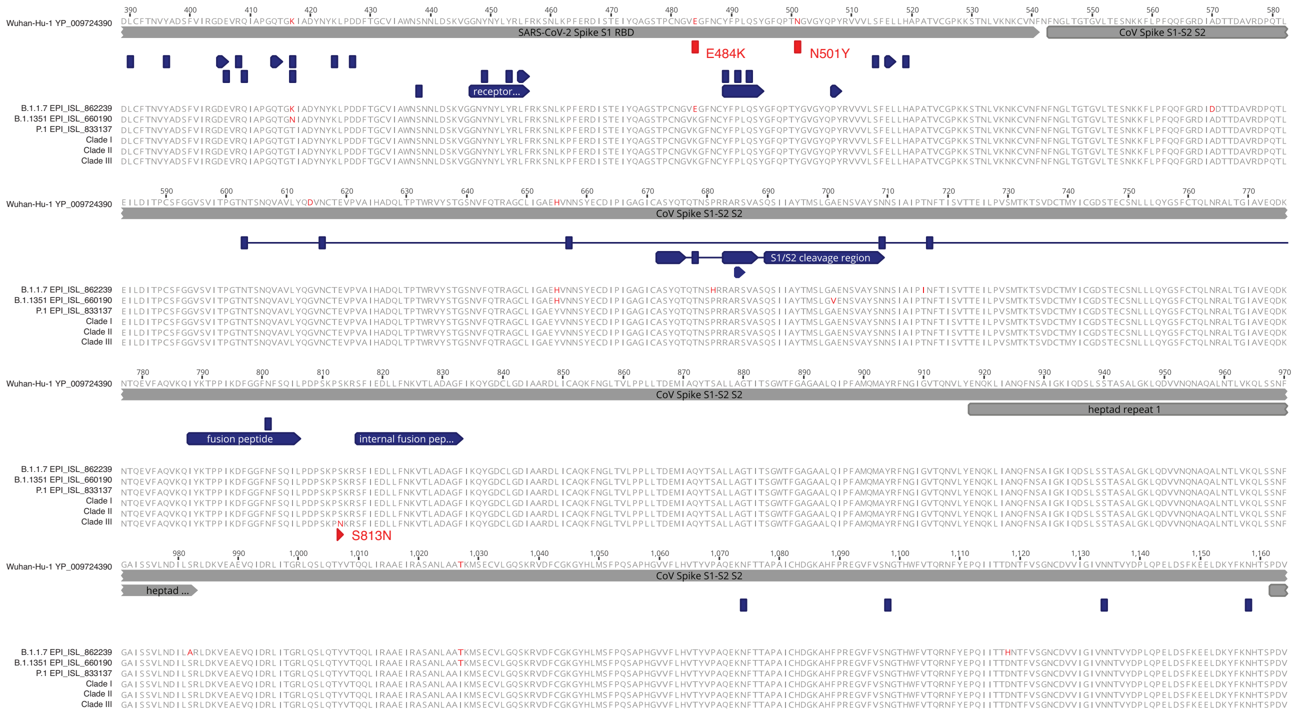


Amino acid alignment of the partial spike protein sequence is shown for reference sequences of different SARS-CoV-2 lineage variants as well as the consensus sequence of clade I – III of P.1 (Fig. 3). Functional features are annotated and amino acid differences are marked in red. The E484K, N501Y and S813N amino acid substitutions are indicated.

**Supplementary Figure S3. Genomic epidemiology of global SARS-CoV-2 lineage B.1.351.**

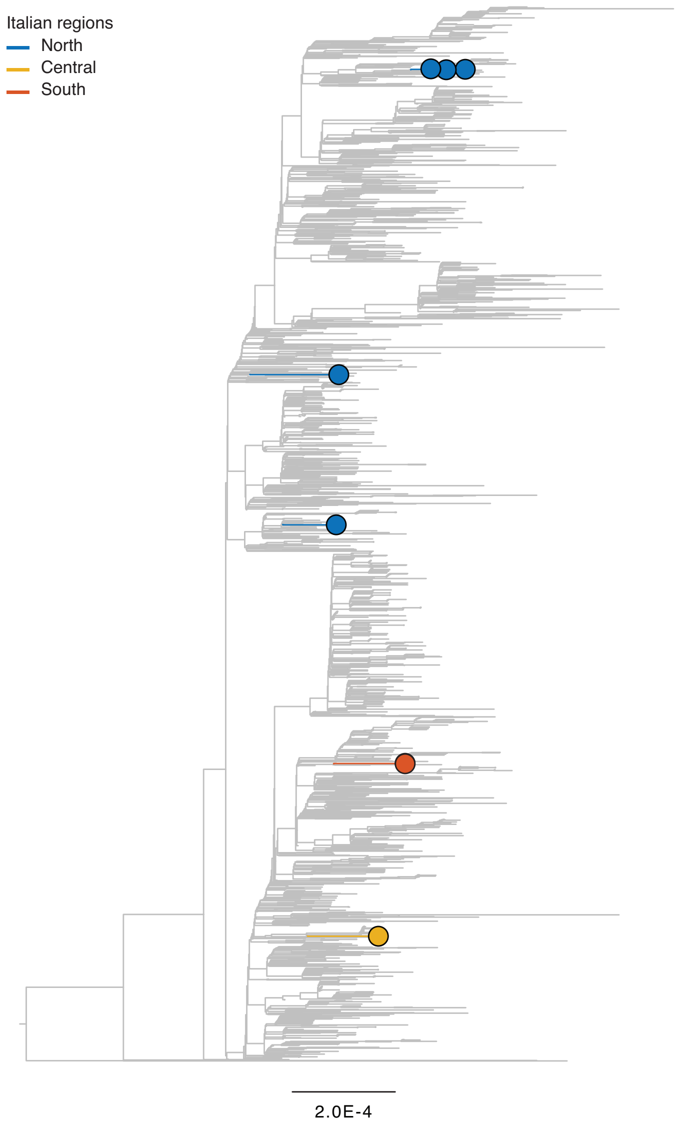


Maximum likelihood phylogeny of SARS-CoV-2 full genome sequences (n=2,309 global, n=8 Italian). Branch lengths show the number of nucleotide substitutions per site and branches are coloured according to geographic region of sampling: global = grey, blue = North Italy, yellow = Central Italy, red = South Italy. Global data is shown in grey.
