## Supplementary Table 1 for "Emergence and spread of SARS-CoV-2 lineages B.1.1.7 and P.1 in Italy"

We gratefully acknowledge the following Authors from the Originating laboratories responsible for obtaining the specimens, as well as the Submitting laboratories where the genome data were generated and shared via GISAID, on which this research is based.

All Submitters of data may be contacted directly via www.gisaid.org

Authors are sorted alphabetically.

| Accession ID | Originating Laboratory | Submitting Laboratory | Authors |
|---|---|---|---|
| EPI_ISL_1007642 | University of Bari Biomedical Sciences and Human Oncology | University of Bari Biomedical Sciences and Human Oncology | Chironna M., Sallustio A., Loconsole D., Accogli M. |
| EPI_ISL_1008663 | Istituto Zooprofilattico Sperimentale della Puglia e della Basilicata | Istituto Zooprofilattico Sperimentale della Puglia e della Basilicata | Parisi A., Bianco A., Capozzi L., Del Sambro L., Simone D., Chironna M., Loconsole D., Sallustio A., Giannico A. |
| EPI_ISL_1008664 | Ospedale Santa Caterina Novella | Istituto Zooprofilattico Sperimentale della Puglia e della Basilicata | Parisi A., Bianco A., Capozzi L., Del Sambro L., Simone D., Chironna M., Loconsole D., Sallustio A., Ridolfi D. |
| EPI_ISL_1008665, EPI_ISL_1008666, EPI_ISL_1008667, EPI_ISL_1008668 | Istituto Zooprofilattico Sperimentale della Puglia e della Basilicata | Istituto Zooprofilattico Sperimentale della Puglia e della Basilicata | Parisi A., Bianco A., Capozzi L., Del Sambro L., Simone D., Chironna M., Loconsole D., Sallustio A., Ridolfi D. |
| EPI_ISL_1008669, EPI_ISL_1008670, EPI_ISL_1008671, EPI_ISL_1008672, EPI_ISL_1008673, EPI_ISL_1008674, EPI_ISL_1008675, EPI_ISL_1008676, EPI_ISL_1008677 | Istituto Zooprofilattico Sperimentale della Puglia e della Basilicata | Istituto Zooprofilattico Sperimentale della Puglia e della Basilicata | Parisi A., Bianco A., Capozzi L., Del Sambro L., Simone D., Chironna M., Loconsole D., Sallustio A., Giannico A. |
| EPI_ISL_1008678 | Presidio di Brindisi Di Summa - Perrino | Istituto Zooprofilattico Sperimentale della Puglia e della Basilicata | Parisi A., Bianco A., Capozzi L., Del Sambro L., Simone D., Chironna M., Loconsole D., Sallustio A., Ridolfi D. |
| EPI_ISL_1008679, EPI_ISL_1008680, EPI_ISL_1008681, EPI_ISL_1008682, EPI_ISL_1008683, EPI_ISL_1008684, EPI_ISL_1008685, EPI_ISL_1008686, EPI_ISL_1008687 | Istituto Zooprofilattico Sperimentale della Puglia e della Basilicata | Istituto Zooprofilattico Sperimentale della Puglia e della Basilicata | Parisi A., Bianco A., Capozzi L., Del Sambro L., Simone D., Chironna M., Loconsole D., Sallustio A., Ridolfi D. |
| EPI_ISL_1008688, EPI_ISL_1008689, EPI_ISL_1008690, EPI_ISL_1008691, EPI_ISL_1008692, EPI_ISL_1008693, EPI_ISL_1008694, EPI_ISL_1008695, EPI_ISL_1008696, EPI_ISL_1008697, EPI_ISL_1008698, EPI_ISL_1008699 | | | |
| see above | Istituto Zooprofilattico Sperimentale della Puglia e della Basilicata | Istituto Zooprofilattico Sperimentale della Puglia e della Basilicata | Parisi A., Bianco A., Capozzi L., Del Sambro L., Simone D., Chironna M., Loconsole D., Sallustio A., Giannico A. |
| EPI_ISL_1008700 | Ospedale Santa Caterina Novella | Istituto Zooprofilattico Sperimentale della Puglia e della Basilicata | Parisi A., Bianco A., Capozzi L., Del Sambro L., Simone D., Chironna M., Loconsole D., Sallustio A., Ridolfi D. |
| EPI_ISL_1008701, EPI_ISL_1008702 | Istituto Zooprofilattico Sperimentale della Puglia e della Basilicata | Istituto Zooprofilattico Sperimentale della Puglia e della Basilicata | Parisi A., Bianco A., Capozzi L., Del Sambro L., Simone D., Chironna M., Loconsole D., Sallustio A., Giannico A. |
| EPI_ISL_1008703, EPI_ISL_1008704, EPI_ISL_1008705 | Presidio di Brindisi Di Summa - Perrino | Istituto Zooprofilattico Sperimentale della Puglia e della Basilicata | Parisi A., Bianco A., Capozzi L., Del Sambro L., Simone D., Chironna M., Loconsole D., Sallustio A., Ridolfi D. |
| EPI_ISL_1008706 | Ospedale Santissima Annunziata | Istituto Zooprofilattico Sperimentale della Puglia e della Basilicata | Parisi A., Bianco A., Capozzi L., Del Sambro L., Simone D., Chironna M., Loconsole D., Sallustio A., Ridolfi D. |
| EPI_ISL_1008707, EPI_ISL_1008708, EPI_ISL_1008709, EPI_ISL_1008710, EPI_ISL_1008711, EPI_ISL_1008712 | Presidio di Brindisi Di Summa - Perrino | Istituto Zooprofilattico Sperimentale della Puglia e della Basilicata | Parisi A., Bianco A., Capozzi L., Del Sambro L., Simone D., Chironna M., Loconsole D., Sallustio A., Ridolfi D. |
| EPI_ISL_1009018 | Istituto Zooprofilattico Sperimentale Lazio e Toscana "M. Aleandri" | INMI Lazzaro Spallanzani IRCCS | CEM Gruber, B Bartolini, E Giombini, M Rueca, O Butera, F Messina, MT Scicluna, G Manna, A Cersini, A Di Caro, MR Capobianchi |
| EPI_ISL_1009024 | Fondazione Policlinico Universitario "A. Gemelli" IRCCS | INMI Lazzaro Spallanzani IRCCS | E Giombini, M. Rueca, B Bartolini, O Butera, C.E.M Gruber, F Messina, P Cattani,M Sanguinetti, A Di Caro, MR Capobianchi |
| EPI_ISL_1009025 | Fondazione Policlinico Universitario "A. Gemelli" IRCCS | INMI Lazzaro Spallanzani IRCCS | B Bartolini, O Butera, C.E.M Gruber, M Rueca, F Messina, E Giombini, P Cattani,M Sanguinetti, MR Capobianchi, A Di Caro |
| EPI_ISL_1009026 | Fondazione Policlinico Universitario "A. Gemelli" IRCCS | INMI Lazzaro Spallanzani IRCCS | F Messina, C.E.M Gruber, B Bartolini, E Giombini, M Rueca, O Butera, P Cattani,M Sanguinetti, A Di Caro, MR Capobianchi |
| EPI_ISL_1009027 | Fondazione Policlinico Universitario "A. Gemelli" IRCCS | INMI Lazzaro Spallanzani IRCCS | M Rueca, O Butera, F Messina, CEM Gruber, B Bartolini, E Giombini, P Cattani,M Sanguinetti, A Di Caro, MR Capobianchi |
| EPI_ISL_1009028 | AOU Policlinico Umberto I; Sapienza Università di Roma | INMI Lazzaro Spallanzani IRCCS | CEM Gruber, B Bartolini, E Giombini, M Rueca, O Butera, F Messina, G Antonelli, O Turriziani, A Di Caro, MR Capobianchi |
| EPI_ISL_1009029 | AOU Policlinico Umberto I; Sapienza Università di Roma | INMI Lazzaro Spallanzani IRCCS | B Bartolini, O Butera, C.E.M Gruber, M Rueca, F Messina, E Giombini, G Antonelli, O Turriziani, MR Capobianchi, A Di Caro |
| EPI_ISL_1009030 | AOU Policlinico Umberto I; Sapienza Università di Roma | INMI Lazzaro Spallanzani IRCCS | M Rueca, O Butera, F Messina, CEM Gruber, B Bartolini, E Giombini, G Antonelli, O Turriziani, A Di Caro, MR Capobianchi |
| EPI_ISL_1009031 | AOU Policlinico Umberto I; Sapienza Università di Roma | INMI Lazzaro Spallanzani IRCCS | E Giombini, M. Rueca, B Bartolini, O Butera, C.E.M Gruber, F Messina, G Antonelli, O Turriziani, MR Capobianchi, A Di Caro |
| EPI_ISL_1012923, EPI_ISL_1012924 | Department of Infectious Diseases, Istituto Superiore di Sanità, Rome, Italy; ASST Sette Laghi, Varese, Italy | Istituto Superiore di Sanità (ISS) | Paola Stefanelli, Angela Di Martino, Alessandra Lo Presti, Stefano Fiore, Fabrizio Maggi, Federica Novazzi, Andreina Baj, Angelo Genoni, Manuela Marra, Maria Carollo, Marco Crescenzi |
| EPI_ISL_1013095 | Azienda Sanitaria dell'Alto Adige Laboratorio Aziendale di Microbiologia e Virologia | Istituto di Genomica Applicata | Elisabetta Pagani, Irene Bianconi, Elisabetta Giacobazzi, Elisa Masi, Stefanie Wieser, Irena Jurman, Vera Vendramin, Eleonora Paparelli, Davide Scaglione, Michele Morgante |
| EPI_ISL_1013531, EPI_ISL_1013532, EPI_ISL_1013557, EPI_ISL_1013558, EPI_ISL_1013559, EPI_ISL_1013560, EPI_ISL_1013561, EPI_ISL_1013562, EPI_ISL_1013563, EPI_ISL_1013564, EPI_ISL_1013565 | | | |
| see above | 1.AO Universitaria 'S. Giovanni di Dio e Ruggi D'Aragona, Scuola Medica Salernitana' Hospital / 2.UOC di Virologia e Microbiologia, Università della Campania 'L. Vanvitelli' / 3.AO Universitaria 'Federico II' Napoli Hospital / 4.AORN 'San Giuseppe Moscati' Avellino Hospital / 5.AO 'San Pio - presidio G. Rummo' Benevento Hospital / 6.AO 'Sant'Anna e San Sebastiano' Caserta Hospital / 7.PO 'Maria Santissima Addolorata' Eboli Hospital / 8.Biogem Istituto di Ricerche Genetiche | 1. Genome Research Center for Health (CRGS) / 2. Laboratory of Molecular Medicine and Genomics(LMMGe) / 3. Center for Research in Pure and Applied Mathematics (CRMPA) | Giorgio Giurato, Francesca Rizzo, Alessandro Weisz, Gianluigi Franci, Giovanni Nassa, Pasquale Pagliano, Roberta Tarallo, Elena Alexandrova, Ylenia D'Agostino, Carlo Ferravante, Jessica Lamberti, Viola Melone, Domenico Memoli, Valeria Mirici Cappa, Domenico Palumbo, Giovanni Pecoraro, Assunta Sellitto, Oriana Strianese, Ilaria Terenzi, Giuseppe Fenza, Aniello Gentile, Antonello Saccomanno, Sonia Amabile, Teresa Rocco, Annamaria Salvati, Emilia Vaccaro, Massimiliano Galdiero, Michele Cennamo, Giuseppe Portella, Maria Grazia Foti, Mariarosaria Ingino, Maria Landi, Maurizio Fumi, Vincenzo Rocco, Rita Greco, Vittoria Letizia, Arnolfo Petruzziello, Maddalena Schioppa, Gregorio Goffredi, Francesca Marciano, Michele Caraglia, Alessia Cossu, Marianna Scrima, Edmondo Adorisio, Morena D'Avenia, Michela Iacobellis, Rosanna Piluscio, Giorgio Dirani, Vittorio Sambri, Simona Semprini, Silvia Zanoli, Francesco Curcio, Stefania Marzinotto, Andreina Baj, Fausto Sessa. |
| EPI_ISL_1014237 | University of Bari Biomedical Sciences and Human Oncology | University of Bari Biomedical Sciences and Human Oncology | Chironna M., Sallustio A., Loconsole D., Accogli M. |
| EPI_ISL_1014545 | Department of Infectious Diseases, Istituto Superiore di Sanità, Rome, Italy; Università degli Studi di Perugia, Perugia, Italy | Istituto Superiore di Sanità (ISS) | Paola Stefanelli, Alessandra Lo Presti, Angela Di Martino, Stefano Fiore, Antonella Mencacci, Barbara Camilloni, Manuela Marra, Maria Carollo, Marco Crescenzi |
| EPI_ISL_1014675 | Department of Infectious Diseases, Istituto Superiore di Sanità, | Istituto Superiore di Sanità (ISS) | Paola Stefanelli, Alessandra Lo Presti, Angela Di Martino, Stefano Fiore, Antonella Mencacci, Barbara Camilloni, Manuela Marra, Maria Carollo, Marco |

| | | | |
|---|---|---|---|
| | Rome, Italy; Università degli Studi di Perugia, Perugia, Italy | | Crescenzi, Luca De Sabato |
| EPI_ISL_1014690 | University of Bari Biomedical Sciences and Human Oncology | University of Bari Biomedical Sciences and Human Oncology | Chironna M., Sallustio A., Loconsole D., Accogli M. |
| EPI_ISL_1015515, EPI_ISL_1015516, EPI_ISL_1015517, EPI_ISL_1015518, EPI_ISL_1015519, EPI_ISL_1015520, EPI_ISL_1015521, EPI_ISL_1015532, EPI_ISL_1015533, EPI_ISL_1015534, EPI_ISL_1015535, EPI_ISL_1015536, EPI_ISL_1015537, EPI_ISL_1015538, EPI_ISL_1015539, EPI_ISL_1015540, EPI_ISL_1015541, EPI_ISL_1015542, EPI_ISL_1015543, EPI_ISL_1015544, EPI_ISL_1015545, EPI_ISL_1015546, EPI_ISL_1015547, EPI_ISL_1015548, EPI_ISL_1015549, EPI_ISL_1015550, EPI_ISL_1015551, EPI_ISL_1015552, EPI_ISL_1015553, EPI_ISL_1015554, EPI_ISL_1015555, EPI_ISL_1015556, EPI_ISL_1015557, EPI_ISL_1015558, EPI_ISL_1015559, EPI_ISL_1015560 | | | |
| see above | Azienda Sanitaria dell'Alto Adige Laboratorio Aziendale di Microbiologia e Virologia | Istituto di Genomica Applicata | Elisabetta Pagani, Irene Bianconi, Elisabetta Giacobazzi, Elisa Masi, Stefanie Wieser, Irena Jurman, Vera Vendramin, Gabriele Magris, Eleonora Paparelli, Davide Scaglione, Michele Morgante |
| EPI_ISL_1020310, EPI_ISL_1020314, EPI_ISL_1020329, EPI_ISL_1020331, EPI_ISL_1020333, EPI_ISL_1020334, EPI_ISL_1020335, EPI_ISL_1020336, EPI_ISL_1020337 | Laboratorio di Riferimento Regionale della Sicilia Occidentale per l'Emergenza COVID-19 | Laboratorio di Riferimento Regionale della Sicilia Occidentale per l'Emergenza COVID-19 | Fabio Tramuto, Carmelo Massimo Maida, Daniela Di Naro, Giulia Randazzo, Walter Mazzucco, Giorgio Graziano, Vincenzo Restivo, Claudio Costantino, Francesco Vitale |
| EPI_ISL_1023524 | Ospedale San Camillo De Lellis di Rieti | INMI Lazzaro Spallanzani IRCCS | Cesare E.M. Gruber, Barbara Bartolini, Emanuela Giombini, Francesco Messina, Martina Rueca, Ornella Butera, Stefano Venarubea, Luca Casertano, Luisa Marchioni, Antonino Di Caro, Maria R. Capobianchi |
| EPI_ISL_1034144 | MEDICO COMPETENTE P.O. L'AQUILA | Istituto Zooprofilattico Sperimentale dell'Abruzzo e Molise "G. Caporale" | Lorusso A, Marcacci M, Di Domenico M, Ancora M, Curini V, Mangone I, Rinaldi A, Scialabba S, Di Pasquale A, Cammà C, Puglia I, Calistri P, Savini G |
| EPI_ISL_1034280 | INMI Lazzaro Spallanzani IRCCS | INMI Lazzaro Spallanzani IRCCS | Cesare E.M. Gruber, Barbara Bartolini, Emanuela Giombini, Francesco Messina, Martina Rueca, Ornella Butera, Emanuele Nicastri, Alessandra D'Abramo, Antonino Di Caro, Maria R. Capobianchi |
| EPI_ISL_1034748, EPI_ISL_1034749, EPI_ISL_1034750, EPI_ISL_1034751, EPI_ISL_1034752, EPI_ISL_1034753 | Center of Advanced Studies and Technology, Molecular Genetics Laboratory | Center of Advanced Studies and Technology, Molecular Genetics Laboratory | Ferrante Rossella, Mandatori Domitilla, De Fabritiis Simone, Damiani Verena, Anaclerio Federico |
| EPI_ISL_1034793 | Virologia Dipartimento di Scienze Biomediche Università di Sassari Viale San Pietro, 43/B - Sassari | Laboratorio Specialistico di Ematologia, Ospedale "San Francesco", via Manniron i 1, 08100 Nuoro | Giovanna Piras, Tatiana Fancello, Maria Monne, Rosanna Asproni, Caterina Serra, Elena Rimini, Salvatore Rubino |
| EPI_ISL_1034916 | Laboratorio Biologia Molecolare Sars Cov2 - UOC Laboratorio Analisi - Servizio Medicina di Laboratorio, Ospedale "San Francesco" - ATS-ASSL Nuoro | Laboratorio specialistico UOC Ematologia - Ospedale "San Francesco" - ATS-ASSL Nuoro | Piras Giovanna, Asproni Rosanna, Malune Paolo, Fiamma Maura, Monne Maria Itria, Palmas Angelo Domenico, Lo Maglio Iana, Mameli Giuseppe |
| EPI_ISL_1035871 | Department of Infectious Diseases, Istituto Superiore di Sanità, Rome, Italy; Università degli Studi di Siena, Siena, Italy | Istituto Superiore di Sanità (ISS) | Paola Stefanelli, Angela Di Martino, Alessandra Lo Presti, Stefano Fiore, Maria Grazia Cusi, Gabriele Anichini, Claudia Gandolfo, Gianni Gori Savellini, Manuela Marra, Maria Carollo, Marco Crescenzi |
| EPI_ISL_1035873, EPI_ISL_1035874, EPI_ISL_1035875, EPI_ISL_1035876, EPI_ISL_1035877, EPI_ISL_1035878, EPI_ISL_1035879, EPI_ISL_1035880, EPI_ISL_1035881, EPI_ISL_1035882, EPI_ISL_1035883, EPI_ISL_1035884, EPI_ISL_1035885, EPI_ISL_1035886, EPI_ISL_1035887, EPI_ISL_1035888 | | | |
| see above | Istituto Zooprofilattico Sperimentale del Mezzogiorno | IZSM-U.O.C. Virologia | Maurizio Viscardi, Lorena Cardillo, Pellegrino Cerino, Massimo Zollo,Giovanna Fusco, Esterina De Carlo, Antonio Limone |
| EPI_ISL_1035893, EPI_ISL_1035894, EPI_ISL_1035895, EPI_ISL_1035896, EPI_ISL_1035897, EPI_ISL_1035898, EPI_ISL_1035899 | Istituto Zooprofilattico Sperimentale del Mezzogiorno | IZSM-U.O.C. Virologia | Maurizio Viscardi, Lorena Cardillo, Sergio Brandi, Pellegrino Cerino, Massimo Zollo,Giovanna Fusco, Esterina De Carlo, Antonio Limone |
| EPI_ISL_1035900, EPI_ISL_1035901, EPI_ISL_1035966 | Department of Infectious Diseases, Istituto Superiore di Sanità, Rome, Italy; Università degli Studi di Siena, Siena, Italy | Istituto Superiore di Sanità (ISS) | Paola Stefanelli, Angela Di Martino, Alessandra Lo Presti, Stefano Fiore, Maria Grazia Cusi, Gabriele Anichini, Claudia Gandolfo, Gianni Gori Savellini, Manuela Marra, Maria Carollo, Marco Crescenzi |
| EPI_ISL_1036140 | SIESP TERAMO | Istituto Zooprofilattico Sperimentale dell'Abruzzo e Molise "G. Caporale" | Lorusso A, Marcacci M, Di Domenico M, Ancora M, Curini V, Mangone I, Rinaldi A, Scialabba S, Di Pasquale A, Cammà C, Puglia I, Calistri P, Savini G |
| EPI_ISL_1036141 | SIESP SULM | Istituto Zooprofilattico Sperimentale dell'Abruzzo e Molise "G. Caporale" | Lorusso A, Marcacci M, Di Domenico M, Ancora M, Curini V, Mangone I, Rinaldi A, Scialabba S, Di Pasquale A, Cammà C, Puglia I, Calistri P, Savini G |
| EPI_ISL_1036142 | OSP CIV ATRI MEDICINA INT | Istituto Zooprofilattico Sperimentale dell'Abruzzo e Molise "G. Caporale" | Lorusso A, Marcacci M, Di Domenico M, Ancora M, Curini V, Mangone I, Rinaldi A, Scialabba S, Di Pasquale A, Cammà C, Puglia I, Calistri P, Savini G |
| EPI_ISL_1036143 | OSP SAN SALVATORE MED INT | Istituto Zooprofilattico Sperimentale dell'Abruzzo e Molise "G. Caporale" | Lorusso A, Marcacci M, Di Domenico M, Ancora M, Curini V, Mangone I, Rinaldi A, Scialabba S, Di Pasquale A, Cammà C, Puglia I, Calistri P, Savini G |
| EPI_ISL_1036144, EPI_ISL_1036145 | SIESP SULM | Istituto Zooprofilattico Sperimentale dell'Abruzzo e Molise "G. Caporale" | Lorusso A, Marcacci M, Di Domenico M, Ancora M, Curini V, Mangone I, Rinaldi A, Scialabba S, Di Pasquale A, Cammà C, Puglia I, Calistri P, Savini G |
| EPI_ISL_1036146 | SIESP CH | Istituto Zooprofilattico Sperimentale dell'Abruzzo e Molise "G. Caporale" | Lorusso A, Marcacci M, Di Domenico M, Ancora M, Curini V, Mangone I, Rinaldi A, Scialabba S, Di Pasquale A, Cammà C, Puglia I, Calistri P, Savini G |
| EPI_ISL_1036147 | SIESP DIP PREV CHIETI | Istituto Zooprofilattico Sperimentale dell'Abruzzo e Molise "G. Caporale" | Lorusso A, Marcacci M, Di Domenico M, Ancora M, Curini V, Mangone I, Rinaldi A, Scialabba S, Di Pasquale A, Cammà C, Puglia I, Calistri P, Savini G |
| EPI_ISL_1036148 | USCA AVEZZANO | Istituto Zooprofilattico Sperimentale dell'Abruzzo e Molise "G. Caporale" | Lorusso A, Marcacci M, Di Domenico M, Ancora M, Curini V, Mangone I, Rinaldi A, Scialabba S, Di Pasquale A, Cammà C, Puglia I, Calistri P, Savini G |
| EPI_ISL_1036149 | OSP SAN SALVATORE MED INT | Istituto Zooprofilattico Sperimentale dell'Abruzzo e Molise "G. Caporale" | Lorusso A, Marcacci M, Di Domenico M, Ancora M, Curini V, Mangone I, Rinaldi A, Scialabba S, Di Pasquale A, Cammà C, Puglia I, Calistri P, Savini G |
| EPI_ISL_1036150 | SIESP CH | Istituto Zooprofilattico Sperimentale dell'Abruzzo e Molise "G. Caporale" | Lorusso A, Marcacci M, Di Domenico M, Ancora M, Curini V, Mangone I, Rinaldi A, Scialabba S, Di Pasquale A, Cammà C, Puglia I, Calistri P, Savini G |
| EPI_ISL_1036151 | PRES OSP TAGLIACOZZO | Istituto Zooprofilattico Sperimentale dell'Abruzzo e Molise "G. Caporale" | Lorusso A, Marcacci M, Di Domenico M, Ancora M, Curini V, Mangone I, Rinaldi A, Scialabba S, Di Pasquale A, Cammà C, Puglia I, Calistri P, Savini G |
| EPI_ISL_1036153 | SIESP AQ | Istituto Zooprofilattico Sperimentale dell'Abruzzo e Molise "G. Caporale" | Lorusso A, Marcacci M, Di Domenico M, Ancora M, Curini V, Mangone I, Rinaldi A, Scialabba S, Di Pasquale A, Cammà C, Puglia I, Calistri P, Savini G |
| EPI_ISL_1036154, EPI_ISL_1036155 | SIESP TERAMO | Istituto Zooprofilattico Sperimentale dell'Abruzzo e Molise "G. Caporale" | Lorusso A, Marcacci M, Di Domenico M, Ancora M, Curini V, Mangone I, Rinaldi A, Scialabba S, Di Pasquale A, Cammà C, Puglia I, Calistri P, Savini G |
| EPI_ISL_1036156 | SIESP DIP PREV CHIETI | Istituto Zooprofilattico Sperimentale dell'Abruzzo e Molise "G. Caporale" | Lorusso A, Marcacci M, Di Domenico M, Ancora M, Curini V, Mangone I, Rinaldi A, Scialabba S, Di Pasquale A, Cammà C, Puglia I, Calistri P, Savini G |
| EPI_ISL_1036157 | SIESP TERAMO | Istituto Zooprofilattico Sperimentale dell'Abruzzo e Molise "G. Caporale" | Lorusso A, Marcacci M, Di Domenico M, Ancora M, Curini V, Mangone I, Rinaldi A, Scialabba S, Di Pasquale A, Cammà C, Puglia I, Calistri P, Savini G |
| EPI_ISL_1036158 | OSP SAN SALVATORE MED INT | Istituto Zooprofilattico Sperimentale dell'Abruzzo e Molise "G. Caporale" | Lorusso A, Marcacci M, Di Domenico M, Ancora M, Curini V, Mangone I, Rinaldi A, Scialabba S, Di Pasquale A, Cammà C, Puglia I, Calistri P, Savini G |
| EPI_ISL_1036159 | OSP CIV GIULIANOVA PRONTO SOCC | Istituto Zooprofilattico Sperimentale dell'Abruzzo e Molise "G. Caporale" | Lorusso A, Marcacci M, Di Domenico M, Ancora M, Curini V, Mangone I, Rinaldi A, Scialabba S, Di Pasquale A, Cammà C, Puglia I, Calistri P, Savini G |
| EPI_ISL_1036160 | SIESP SULM | Istituto Zooprofilattico Sperimentale dell'Abruzzo e Molise "G. | Lorusso A, Marcacci M, Di Domenico M, Ancora M, Curini V, Mangone I, Rinaldi A, Scialabba S, Di Pasquale A, Cammà C, Puglia I, Calistri P, Savini G |

| | | | |
|---|---|---|---|
| | | Caporale" | |
| EPI_ISL_1036161 | OSP CIV TERAMO RIANIMAZIONE CARDIOCHIR | Istituto Zooprofilattico Sperimentale dell'Abruzzo e Molise "G. Caporale" | Lorusso A, Marcacci M, Di Domenico M, Ancora M, Curini V, Mangone I, Rinaldi A, Scialabba S, Di Pasquale A, Cammà C, Puglia I, Calistri P, Savini G |
| EPI_ISL_1036162 | SIESP TERAMO | Istituto Zooprofilattico Sperimentale dell'Abruzzo e Molise "G. Caporale" | Lorusso A, Marcacci M, Di Domenico M, Ancora M, Curini V, Mangone I, Rinaldi A, Scialabba S, Di Pasquale A, Cammà C, Puglia I, Calistri P, Savini G |
| EPI_ISL_1036163, EPI_ISL_1036164 | SIESP CH | Istituto Zooprofilattico Sperimentale dell'Abruzzo e Molise "G. Caporale" | Lorusso A, Marcacci M, Di Domenico M, Ancora M, Curini V, Mangone I, Rinaldi A, Scialabba S, Di Pasquale A, Cammà C, Puglia I, Calistri P, Savini G |
| EPI_ISL_1036165 | SIESP TERAMO | Istituto Zooprofilattico Sperimentale dell'Abruzzo e Molise "G. Caporale" | Lorusso A, Marcacci M, Di Domenico M, Ancora M, Curini V, Mangone I, Rinaldi A, Scialabba S, Di Pasquale A, Cammà C, Puglia I, Calistri P, Savini G |
| EPI_ISL_1036166, EPI_ISL_1036167 | SIESP CH | Istituto Zooprofilattico Sperimentale dell'Abruzzo e Molise "G. Caporale" | Lorusso A, Marcacci M, Di Domenico M, Ancora M, Curini V, Mangone I, Rinaldi A, Scialabba S, Di Pasquale A, Cammà C, Puglia I, Calistri P, Savini G |
| EPI_ISL_1036168, EPI_ISL_1036169 | SIESP TERAMO | Istituto Zooprofilattico Sperimentale dell'Abruzzo e Molise "G. Caporale" | Lorusso A, Marcacci M, Di Domenico M, Ancora M, Curini V, Mangone I, Rinaldi A, Scialabba S, Di Pasquale A, Cammà C, Puglia I, Calistri P, Savini G |
| EPI_ISL_1036170, EPI_ISL_1036171 | SIESP CH | Istituto Zooprofilattico Sperimentale dell'Abruzzo e Molise "G. Caporale" | Lorusso A, Marcacci M, Di Domenico M, Ancora M, Curini V, Mangone I, Rinaldi A, Scialabba S, Di Pasquale A, Cammà C, Puglia I, Calistri P, Savini G |
| EPI_ISL_1036172 | SIESP TERAMO | Istituto Zooprofilattico Sperimentale dell'Abruzzo e Molise "G. Caporale" | Lorusso A, Marcacci M, Di Domenico M, Ancora M, Curini V, Mangone I, Rinaldi A, Scialabba S, Di Pasquale A, Cammà C, Puglia I, Calistri P, Savini G |
| EPI_ISL_1036173, EPI_ISL_1036174, EPI_ISL_1036175, EPI_ISL_1036176 | SIESP CH | Istituto Zooprofilattico Sperimentale dell'Abruzzo e Molise "G. Caporale" | Lorusso A, Marcacci M, Di Domenico M, Ancora M, Curini V, Mangone I, Rinaldi A, Scialabba S, Di Pasquale A, Cammà C, Puglia I, Calistri P, Savini G |
| EPI_ISL_1036177 | SIESP DIP PREV CHIETI | Istituto Zooprofilattico Sperimentale dell'Abruzzo e Molise "G. Caporale" | Lorusso A, Marcacci M, Di Domenico M, Ancora M, Curini V, Mangone I, Rinaldi A, Scialabba S, Di Pasquale A, Cammà C, Puglia I, Calistri P, Savini G |
| EPI_ISL_1036178 | SIESP SULM | Istituto Zooprofilattico Sperimentale dell'Abruzzo e Molise "G. Caporale" | Lorusso A, Marcacci M, Di Domenico M, Ancora M, Curini V, Mangone I, Rinaldi A, Scialabba S, Di Pasquale A, Cammà C, Puglia I, Calistri P, Savini G |
| EPI_ISL_1036179 | SIESP CH DISTRETTO SANITARIO CHIETI | Istituto Zooprofilattico Sperimentale dell'Abruzzo e Molise "G. Caporale" | Lorusso A, Marcacci M, Di Domenico M, Ancora M, Curini V, Mangone I, Rinaldi A, Scialabba S, Di Pasquale A, Cammà C, Puglia I, Calistri P, Savini G |
| EPI_ISL_1036180, EPI_ISL_1036181 | SIESP TERAMO | Istituto Zooprofilattico Sperimentale dell'Abruzzo e Molise "G. Caporale" | Lorusso A, Marcacci M, Di Domenico M, Ancora M, Curini V, Mangone I, Rinaldi A, Scialabba S, Di Pasquale A, Cammà C, Puglia I, Calistri P, Savini G |
| EPI_ISL_1036182 | SIESP CH | Istituto Zooprofilattico Sperimentale dell'Abruzzo e Molise "G. Caporale" | Lorusso A, Marcacci M, Di Domenico M, Ancora M, Curini V, Mangone I, Rinaldi A, Scialabba S, Di Pasquale A, Cammà C, Puglia I, Calistri P, Savini G |
| EPI_ISL_1036183, EPI_ISL_1036184 | SIESP SULM | Istituto Zooprofilattico Sperimentale dell'Abruzzo e Molise "G. Caporale" | Lorusso A, Marcacci M, Di Domenico M, Ancora M, Curini V, Mangone I, Rinaldi A, Scialabba S, Di Pasquale A, Cammà C, Puglia I, Calistri P, Savini G |
| EPI_ISL_1036185 | SIESP DIP PREV CHIETI | Istituto Zooprofilattico Sperimentale dell'Abruzzo e Molise "G. Caporale" | Lorusso A, Marcacci M, Di Domenico M, Ancora M, Curini V, Mangone I, Rinaldi A, Scialabba S, Di Pasquale A, Cammà C, Puglia I, Calistri P, Savini G |
| EPI_ISL_1036186 | OSP SAN SALVATORE MED INT | Istituto Zooprofilattico Sperimentale dell'Abruzzo e Molise "G. Caporale" | Lorusso A, Marcacci M, Di Domenico M, Ancora M, Curini V, Mangone I, Rinaldi A, Scialabba S, Di Pasquale A, Cammà C, Puglia I, Calistri P, Savini G |
| EPI_ISL_1036187 | SIESP TERAMO | Istituto Zooprofilattico Sperimentale dell'Abruzzo e Molise "G. Caporale" | Lorusso A, Marcacci M, Di Domenico M, Ancora M, Curini V, Mangone I, Rinaldi A, Scialabba S, Di Pasquale A, Cammà C, Puglia I, Calistri P, Savini G |
| EPI_ISL_1036188 | OSP SAN SALVATORE UOC MAL INF | Istituto Zooprofilattico Sperimentale dell'Abruzzo e Molise "G. Caporale" | Lorusso A, Marcacci M, Di Domenico M, Ancora M, Curini V, Mangone I, Rinaldi A, Scialabba S, Di Pasquale A, Cammà C, Puglia I, Calistri P, Savini G |
| EPI_ISL_1036189 | OSP CIV TERAMO PNEUMOLOGIA | Istituto Zooprofilattico Sperimentale dell'Abruzzo e Molise "G. Caporale" | Lorusso A, Marcacci M, Di Domenico M, Ancora M, Curini V, Mangone I, Rinaldi A, Scialabba S, Di Pasquale A, Cammà C, Puglia I, Calistri P, Savini G |
| EPI_ISL_1036190 | SIESP CH | Istituto Zooprofilattico Sperimentale dell'Abruzzo e Molise "G. Caporale" | Lorusso A, Marcacci M, Di Domenico M, Ancora M, Curini V, Mangone I, Rinaldi A, Scialabba S, Di Pasquale A, Cammà C, Puglia I, Calistri P, Savini G |
| EPI_ISL_1036191 | SIESP SULM | Istituto Zooprofilattico Sperimentale dell'Abruzzo e Molise "G. Caporale" | Lorusso A, Marcacci M, Di Domenico M, Ancora M, Curini V, Mangone I, Rinaldi A, Scialabba S, Di Pasquale A, Cammà C, Puglia I, Calistri P, Savini G |
| EPI_ISL_1036192 | SIESP TERAMO | Istituto Zooprofilattico Sperimentale dell'Abruzzo e Molise "G. Caporale" | Lorusso A, Marcacci M, Di Domenico M, Ancora M, Curini V, Mangone I, Rinaldi A, Scialabba S, Di Pasquale A, Cammà C, Puglia I, Calistri P, Savini G |
| EPI_ISL_1036193, EPI_ISL_1036194, EPI_ISL_1036195, EPI_ISL_1036196, EPI_ISL_1036197 | SIESP CH | Istituto Zooprofilattico Sperimentale dell'Abruzzo e Molise "G. Caporale" | Lorusso A, Marcacci M, Di Domenico M, Ancora M, Curini V, Mangone I, Rinaldi A, Scialabba S, Di Pasquale A, Cammà C, Puglia I, Calistri P, Savini G |
| EPI_ISL_1036198 | ASL AVEZZANO-SULMONA-L'AQUILA DIP PREV SERV IGIENE E SAN PUBBLICA | Istituto Zooprofilattico Sperimentale dell'Abruzzo e Molise "G. Caporale" | Lorusso A, Marcacci M, Di Domenico M, Ancora M, Curini V, Mangone I, Rinaldi A, Scialabba S, Di Pasquale A, Cammà C, Puglia I, Calistri P, Savini G |
| EPI_ISL_1036199, EPI_ISL_1036200, EPI_ISL_1036201 | SIESP CH | Istituto Zooprofilattico Sperimentale dell'Abruzzo e Molise "G. Caporale" | Lorusso A, Marcacci M, Di Domenico M, Ancora M, Curini V, Mangone I, Rinaldi A, Scialabba S, Di Pasquale A, Cammà C, Puglia I, Calistri P, Savini G |
| EPI_ISL_1036202 | SIESP TERAMO | Istituto Zooprofilattico Sperimentale dell'Abruzzo e Molise "G. Caporale" | Lorusso A, Marcacci M, Di Domenico M, Ancora M, Curini V, Mangone I, Rinaldi A, Scialabba S, Di Pasquale A, Cammà C, Puglia I, Calistri P, Savini G |
| EPI_ISL_1036203, EPI_ISL_1036204 | SIESP CH | Istituto Zooprofilattico Sperimentale dell'Abruzzo e Molise "G. Caporale" | Lorusso A, Marcacci M, Di Domenico M, Ancora M, Curini V, Mangone I, Rinaldi A, Scialabba S, Di Pasquale A, Cammà C, Puglia I, Calistri P, Savini G |
| EPI_ISL_1036205 | PRES TERR CASOLI | Istituto Zooprofilattico Sperimentale dell'Abruzzo e Molise "G. Caporale" | Lorusso A, Marcacci M, Di Domenico M, Ancora M, Curini V, Mangone I, Rinaldi A, Scialabba S, Di Pasquale A, Cammà C, Puglia I, Calistri P, Savini G |
| EPI_ISL_1036206 | SIESP AQ | Istituto Zooprofilattico Sperimentale dell'Abruzzo e Molise "G. Caporale" | Lorusso A, Marcacci M, Di Domenico M, Ancora M, Curini V, Mangone I, Rinaldi A, Scialabba S, Di Pasquale A, Cammà C, Puglia I, Calistri P, Savini G |
| EPI_ISL_1036207, EPI_ISL_1036208 | SIESP TERAMO | Istituto Zooprofilattico Sperimentale dell'Abruzzo e Molise "G. Caporale" | Lorusso A, Marcacci M, Di Domenico M, Ancora M, Curini V, Mangone I, Rinaldi A, Scialabba S, Di Pasquale A, Cammà C, Puglia I, Calistri P, Savini G |
| EPI_ISL_1036209 | SIESP DIP PREV CHIETI | Istituto Zooprofilattico Sperimentale dell'Abruzzo e Molise "G. Caporale" | Lorusso A, Marcacci M, Di Domenico M, Ancora M, Curini V, Mangone I, Rinaldi A, Scialabba S, Di Pasquale A, Cammà C, Puglia I, Calistri P, Savini G |
| EPI_ISL_1036210 | SIESP TERAMO | Istituto Zooprofilattico Sperimentale dell'Abruzzo e Molise "G. Caporale" | Lorusso A, Marcacci M, Di Domenico M, Ancora M, Curini V, Mangone I, Rinaldi A, Scialabba S, Di Pasquale A, Cammà C, Puglia I, Calistri P, Savini G |
| EPI_ISL_1036211 | ASL AVEZZANO-SULMONA-L'AQUILA | Istituto Zooprofilattico Sperimentale dell'Abruzzo e Molise "G. Caporale" | Lorusso A, Marcacci M, Di Domenico M, Ancora M, Curini V, Mangone I, Rinaldi A, Scialabba S, Di Pasquale A, Cammà C, Puglia I, Calistri P, Savini G |

| | | | |
|---|---|---|---|
| EPI_ISL_1036212, EPI_ISL_1036215, EPI_ISL_1036216 | SIESP TERAMO | Istituto Zooprofilattico Sperimentale dell'Abruzzo e Molise "G. Caporale" | Lorusso A, Marcacci M, Di Domenico M, Ancora M, Curini V, Mangone I, Rinaldi A, Scialabba S, Di Pasquale A, Cammà C, Puglia I, Calistri P, Savini G |
| EPI_ISL_1036217 | FONDAZIONE PICCOLA OPERA CARITAS | Istituto Zooprofilattico Sperimentale dell'Abruzzo e Molise "G. Caporale" | Lorusso A, Marcacci M, Di Domenico M, Ancora M, Curini V, Mangone I, Rinaldi A, Scialabba S, Di Pasquale A, Cammà C, Puglia I, Calistri P, Savini G |
| EPI_ISL_1036221 | OSP SAN SALVATORE UOC MAL INF | Istituto Zooprofilattico Sperimentale dell'Abruzzo e Molise "G. Caporale" | Lorusso A, Marcacci M, Di Domenico M, Ancora M, Curini V, Mangone I, Rinaldi A, Scialabba S, Di Pasquale A, Cammà C, Puglia I, Calistri P, Savini G |
| EPI_ISL_1036222 | SIESP TERAMO | Istituto Zooprofilattico Sperimentale dell'Abruzzo e Molise "G. Caporale" | Lorusso A, Marcacci M, Di Domenico M, Ancora M, Curini V, Mangone I, Rinaldi A, Scialabba S, Di Pasquale A, Cammà C, Puglia I, Calistri P, Savini G |
| EPI_ISL_1036223 | OSP SAN SALVATORE MED INT | Istituto Zooprofilattico Sperimentale dell'Abruzzo e Molise "G. Caporale" | Lorusso A, Marcacci M, Di Domenico M, Ancora M, Curini V, Mangone I, Rinaldi A, Scialabba S, Di Pasquale A, Cammà C, Puglia I, Calistri P, Savini G |
| EPI_ISL_1036224, EPI_ISL_1036225, EPI_ISL_1036226, EPI_ISL_1036227, EPI_ISL_1036228, EPI_ISL_1036229 | P.O.CARDARELLI | Istituto Zooprofilattico Sperimentale dell'Abruzzo e Molise "G. Caporale" | Scutellà M, Niro G, Lorusso A, Marcacci M, Di Domenico M, Ancora M, Curini V, Mangone I, Rinaldi A, Scialabba S, Di Pasquale A, Cammà C, Puglia I, Calistri P, Savini G |
| EPI_ISL_1036238, EPI_ISL_1036239 | Center of Advanced Studies and Technology, Molecular Genetics Laboratory | Center of Advanced Studies and Technology, Molecular Genetics Laboratory | Ferrante Rossella, Mandatori Domitilla, De Fabritiis Simone, Damiani Verena, Anaclerio Federico |
| EPI_ISL_1036755 | Unità Operativa di Microbiologia, IRCCS Policlinico di Sant'Orsola, Azienda Ospedaliero-Universitaria di Bologna | Unità di Analisi del Rischio ed Epidemiologia Genomica, Istituto Zooprofilattico Sperimentale dell'Emilia Romagna e della Lombardia (IZSLER) | Giada Rossini, Giuliano Furlini, Tiziana Lazzarotto, Marina Morganti, Ilaria Menozzi, Erika Scaltriti, Stefano Pongolini |
| EPI_ISL_1039782 | INMI Lazzaro Spallanzani IRCCS | INMI Lazzaro Spallanzani IRCCS | CEM Gruber, B Bartolini, E Giombini, F Messina, M Rueca, O Butera, A Di Caro, MR Capobianchi |
| EPI_ISL_1039783 | INMI Lazzaro Spallanzani IRCCS | INMI Lazzaro Spallanzani IRCCS | B Bartolini, M. Rueca, E Giombini, O Butera, C.E.M Gruber, F Messina, MR Capobianchi, A Di Caro |
| EPI_ISL_1039784 | Azienda Ospedaliera San Camillo Forlanini | INMI Lazzaro Spallanzani IRCCS | CEM Gruber, B Bartolini, E Giombini, M Rueca, O Butera, F Messina, D.Gallone, G Parisi, ML Guarino, A Di Caro, MR Capobianchi |
| EPI_ISL_1039785 | Ospedale "F. Spaziani" Frosinone | INMI Lazzaro Spallanzani IRCCS | B Bartolini, E Giombini, C.E.M Gruber, F Messina, M Rueca, O Butera, C. Gargiulo, C Sias, R Pulselli, A Di Caro, MR Capobianchi |
| EPI_ISL_1040918, EPI_ISL_1040919, EPI_ISL_1040920 | Medicine and Surgery, University of Insubria | Medicine and Surgery, University of Insubria | Novazzi,F., Genoni,A., Baj,A., Spezia,P.G., Focosi,D., Zago,C., Colombo,A., Cassani,G., Pasciuta,R., Tamborini,A., Rossi,A., Prestia,M., Capuano,R., Maggi,F. |
| EPI_ISL_1048822, EPI_ISL_1048823, EPI_ISL_1048824, EPI_ISL_1048825, EPI_ISL_1048826, EPI_ISL_1048827, EPI_ISL_1048828 | Azienda Sanitaria dell'Alto Adige Laboratorio Aziendale di Microbiologia e Virologia | Istituto di Genomica Applicata | Elisabetta Pagani, Irene Bianconi, Elisabetta Giacobazzi, Elisa Masi, Stefanie Wieser, Irena Jurman, Vera Vendramin, Gabriele Magris, Eleonora Paparelli, Davide Scaglione, Michele Morgante |
| EPI_ISL_1049260 | Struttura Semplice Dipartimentale di Virologia e Microbiologia molecolare, Azienda Ospedaliero-Universitaria, Policlinico di Modena | U.O. Microbiologia, Laboratorio Unico Centro Servizi - AUSL della Romagna | Pecorari Monica, Gennari William, Fregni Serpini Giulia, Giorgio Dirani, Silvia Zannoli, Vittorio Sambri |
| EPI_ISL_1049261 | U.O. Microbiologia Laboratorio Unico Centro Servizi AUSL della Romagna | U.O. Microbiologia Laboratorio Unico Centro Servizi AUSL della Romagna | Silvia Zannoli, Giorgio Dirani, Vittorio Sambri |
| EPI_ISL_1055765 | Virology Unit, Pisa University Hospital | Virology Unit, AOUP | Marialinda Vatteroni, Susi Frateschi |
| EPI_ISL_1055792 | Virology Unit, Pisa University Hospital | Virology Unit, AOUP, Pisa | Marialinda Vatteroni, Susi Frateschi |
| EPI_ISL_1055800 | Virology Unit, AOUP | Virology Unit, AOUP, Pisa | Marialinda Vatteroni, Susi Frateschi |
| EPI_ISL_1055811 | Virology Unit, AOUP | Virolgy Unit, AOUP | Marialinda Vatteroni, Susi Frateschi |
| EPI_ISL_1055815 | Virology Unit, AOUP | Virology Unit, AOUP, Pisa | Marialinda Vatteroni, Susi Frateschi, Mauro Pistello |
| EPI_ISL_1055820 | Virology Unit, AOUP | Virology Unit, AOUP | Marialinda Vatteroni, Susi Frateschi |
| EPI_ISL_1055824 | Virology Unit, AOUP, Pisa, Italy | Virology Unit, AOUP, Pisa | Marialinda Vatteroni, Susi Frateschi, Mauro Pistello |
| EPI_ISL_1055825 | Virology Unit, AOUP | Virology Unit, AOUP | Marialinda Vatteroni, Susi Frateschi |
| EPI_ISL_1058040 | SIESP CHIETI DRIVE IN LANCIANO | Istituto Zooprofilattico Sperimentale dell'Abruzzo e Molise "G. Caporale" | Lorusso A, Marcacci M, Di Domenico M, Ancora M, Curini V, Mangone I, Rinaldi A, Scialabba S, Di Pasquale A, Cammà C, Puglia I, Calistri P, Savini G |
| EPI_ISL_1058041 | SIESP CHIETI | Istituto Zooprofilattico Sperimentale dell'Abruzzo e Molise "G. Caporale" | Lorusso A, Marcacci M, Di Domenico M, Ancora M, Curini V, Mangone I, Rinaldi A, Scialabba S, Di Pasquale A, Cammà C, Puglia I, Calistri P, Savini G |
| EPI_ISL_1058042 | SIESP CHIETI - DRIVE IN ORTONA | Istituto Zooprofilattico Sperimentale dell'Abruzzo e Molise "G. Caporale" | Lorusso A, Marcacci M, Di Domenico M, Ancora M, Curini V, Mangone I, Rinaldi A, Scialabba S, Di Pasquale A, Cammà C, Puglia I, Calistri P, Savini G |
| EPI_ISL_1058043 | SIESP CHIETI - DRIVE IN CHIETI | Istituto Zooprofilattico Sperimentale dell'Abruzzo e Molise "G. Caporale" | Lorusso A, Marcacci M, Di Domenico M, Ancora M, Curini V, Mangone I, Rinaldi A, Scialabba S, Di Pasquale A, Cammà C, Puglia I, Calistri P, Savini G |
| EPI_ISL_1058044, EPI_ISL_1058045 | SIESP CHIETI - DRIVE IN ORTONA | Istituto Zooprofilattico Sperimentale dell'Abruzzo e Molise "G. Caporale" | Lorusso A, Marcacci M, Di Domenico M, Ancora M, Curini V, Mangone I, Rinaldi A, Scialabba S, Di Pasquale A, Cammà C, Puglia I, Calistri P, Savini G |
| EPI_ISL_1061067, EPI_ISL_1061091, EPI_ISL_1061092, EPI_ISL_1061093, EPI_ISL_1061094, EPI_ISL_1061095, EPI_ISL_1061096, EPI_ISL_1061097, EPI_ISL_1061098, EPI_ISL_1061100, EPI_ISL_1061101, EPI_ISL_1061102, EPI_ISL_1061103, EPI_ISL_1061104, EPI_ISL_1061105, EPI_ISL_1061106, EPI_ISL_1061107, EPI_ISL_1061108, EPI_ISL_1061109, EPI_ISL_1061110, EPI_ISL_1061111, EPI_ISL_1061112, EPI_ISL_1061113, EPI_ISL_1061114, EPI_ISL_1061115, EPI_ISL_1061116, EPI_ISL_1061117, EPI_ISL_1061118, EPI_ISL_1061119, EPI_ISL_1061120, EPI_ISL_1061121, EPI_ISL_1061122, EPI_ISL_1061123, EPI_ISL_1061124, EPI_ISL_1061125, EPI_ISL_1061126, EPI_ISL_1061127, EPI_ISL_1061128, EPI_ISL_1061129, EPI_ISL_1061130, EPI_ISL_1061141, EPI_ISL_1061150, EPI_ISL_1061151, EPI_ISL_1061152, EPI_ISL_1061153, EPI_ISL_1061154, EPI_ISL_1061157, EPI_ISL_1061192, EPI_ISL_1061206, EPI_ISL_1061207, EPI_ISL_1061208, EPI_ISL_1061209, EPI_ISL_1061210, EPI_ISL_1061225, EPI_ISL_1061252, EPI_ISL_1061253, EPI_ISL_1061254, EPI_ISL_1061255, EPI_ISL_1061256, EPI_ISL_1061257, EPI_ISL_1061258, EPI_ISL_1061259 | | | |
| see above | Istituto Zooprofilattico Sperimentale del Mezzogiorno (IZSM) | Telethon Institute of Genetics and Medicine (TIGEM) | Antonio Grimaldi, Patrizia Annunziata, Francesco Panariello, Biancamaria Pierri, Valentina Bouche, Chiara Colantuono, Maria Concetta Cuomo, Denise Di Concilio, Lucio Di Filippo, Anna Manfredi, Marcello Salvi, Antonio Limone, Pellegrino Cerino, Andrea Ballabio, Davide Cacchiarelli. |
| EPI_ISL_1063404, EPI_ISL_1063405, EPI_ISL_1063406, EPI_ISL_1063407, EPI_ISL_1063408, EPI_ISL_1063409, EPI_ISL_1063410, EPI_ISL_1063411, EPI_ISL_1063412, EPI_ISL_1063413, EPI_ISL_1063414, EPI_ISL_1063415, EPI_ISL_1063416, EPI_ISL_1063417, EPI_ISL_1063418, EPI_ISL_1063419, EPI_ISL_1063420, EPI_ISL_1063421, EPI_ISL_1063422, EPI_ISL_1063423, EPI_ISL_1063424, EPI_ISL_1063425, EPI_ISL_1063426, EPI_ISL_1063427, EPI_ISL_1063428, EPI_ISL_1063429, EPI_ISL_1063430, EPI_ISL_1063431, EPI_ISL_1063432, EPI_ISL_1063433, EPI_ISL_1063434, EPI_ISL_1063435, EPI_ISL_1063436, EPI_ISL_1063437, EPI_ISL_1063438, EPI_ISL_1063439, EPI_ISL_1063440, EPI_ISL_1063441, EPI_ISL_1063442, EPI_ISL_1063443, EPI_ISL_1063444, EPI_ISL_1063445, EPI_ISL_1063446, EPI_ISL_1063447, EPI_ISL_1063448, EPI_ISL_1063449, EPI_ISL_1063450, EPI_ISL_1063451, EPI_ISL_1063452, EPI_ISL_1063453, EPI_ISL_1063454, EPI_ISL_1063455, EPI_ISL_1063456, EPI_ISL_1063457, EPI_ISL_1063458, EPI_ISL_1063459, EPI_ISL_1063460, EPI_ISL_1063461, EPI_ISL_1063462, EPI_ISL_1063463, EPI_ISL_1063464, EPI_ISL_1063465, EPI_ISL_1063466, EPI_ISL_1063467, EPI_ISL_1063468, EPI_ISL_1063469, EPI_ISL_1063470, EPI_ISL_1063471, EPI_ISL_1063472, EPI_ISL_1063473, EPI_ISL_1063474, EPI_ISL_1063475, EPI_ISL_1063476, EPI_ISL_1063477, EPI_ISL_1063478, EPI_ISL_1063479, EPI_ISL_1063480, EPI_ISL_1063481, EPI_ISL_1063482, EPI_ISL_1063483, EPI_ISL_1063484, EPI_ISL_1063485, EPI_ISL_1063486, EPI_ISL_1063487, EPI_ISL_1063488, EPI_ISL_1063489, EPI_ISL_1063490, EPI_ISL_1063491, EPI_ISL_1063492, EPI_ISL_1063493 | | | |
| see above | Azienda Sanitaria dell'Alto Adige Laboratorio Aziendale di Microbiologia e Virologia | Istituto di Genomica Applicata | Elisabetta Pagani, Irene Bianconi, Elisabetta Giacobazzi, Elisa Masi, Stefanie Wieser, Irena Jurman, Vera Vendramin, Gabriele Magris, Eleonora Paparelli, Davide Scaglione, Michele Morgante |
| EPI_ISL_1063785, EPI_ISL_1063786, EPI_ISL_1063787 | SIESP CHIETI - DRIVE IN ORTONA | Istituto Zooprofilattico Sperimentale dell'Abruzzo e Molise "G. Caporale" | Lorusso A, Marcacci M, Di Domenico M, Ancora M, Curini V, Mangone I, Rinaldi A, Scialabba S, Di Pasquale A, Cammà C, Puglia I, Calistri P, Savini G |
| EPI_ISL_1063788 | P.O.CARDARELLI | Istituto Zooprofilattico Sperimentale dell'Abruzzo e Molise "G. Caporale" | Scutellà M, Niro G, Lorusso A, Marcacci M, Di Domenico M, Ancora M, Curini V, Mangone I, Rinaldi A, Scialabba S, Di Pasquale A, Cammà C, Puglia I, Calistri P, Savini G |
| EPI_ISL_1063909 | Laboratorio Genzano - ASL RM 6 | INMI Lazzaro Spallanzani IRCCS | B Bartolini, O Butera, C.E.M Gruber, M Rueca, F Messina, E Giombini, G Tramini, E Conti, MR Capobianchi, A Di Caro |

| | | | |
|---|---|---|---|
| EPI_ISL_1063910 | Ospedale San Filippo Neri | INMI Lazzaro Spallanzani IRCCS | M Rueca, O Butera, F Messina, CEM Gruber, B Bartolini, E Giombini, M Melandri, ML Schiavone, A Di Caro, MR Capobianchi |
| EPI_ISL_1063911 | INMI Lazzaro Spallanzani IRCCS | INMI Lazzaro Spallanzani IRCCS | E Giombini, M. Rueca, B Bartolini, O Butera, CEM Gruber, F Messina, A Di Caro, MR Capobianchi |
| EPI_ISL_1063912 | Laboratorio Biologia Molecolare Sars Cov2 - UOC Laboratorio Analisi - Servizio Medicina di Laboratorio, Ospedale "San Francesco" - ATS-ASSL Nuoro | Laboratorio specialistico UOC Ematologia - Ospedale "San Francesco" - ATS-ASSL Nuoro | Piras Giovanna, Asproni Rosanna, Malune Paolo, Fiamma Maura, Monne Maria Itria, Palmas Angelo Domenico, Lo Maglio Iana, Mameli Giuseppe |
| EPI_ISL_1069204 | Azienda Sanitaria Locale Napoli 1 | Centro Polidiagnostico strumentale AMES | Sirica Roberto |
| EPI_ISL_1073034 | ASL Napoli 1 Centro | AMES Centro Polidiagnostico Strumentale S.r.l. | Giovanni Savarese, Raffaella Ruggiero, Eloisa Evangelista, Antonella Di Carlo, Luisa Circelli, Luigi D'Amore, Roberto Sirica, Antonio Fico |
| EPI_ISL_1074622, EPI_ISL_1074623 | ASL Napoli 1 Centro | AMES Centro Polidiagnostico Strumentale S.r.l. | "Giovanni Savarese, Raffaella Ruggiero, Eloisa Evangelista, Antonella Di Carlo, Luisa Circelli, Luigi D'Amore, Roberto Sirica, Antonio Fico" |
| EPI_ISL_1079352 | Genomic Medicine Laboratory, IRCCS Santa Lucia Foundation | INMI Lazzaro Spallanzani IRCCS | CEM Gruber, B Bartolini, E Giombini, F Messina, M Rueca, O Butera, E Giardina, A Di Caro, MR Capobianchi |
| EPI_ISL_1079353 | IRCCS San Raffaele | INMI Lazzaro Spallanzani IRCCS | B Bartolini, E Giombini, F Messina, M Rueca, O Butera, CEM Gruber, D Russo, D Limongi, MR Capobianchi, A Di Caro |
| EPI_ISL_1079355 | Genomic Medicine Laboratory, IRCCS Santa Lucia Foundation | INMI Lazzaro Spallanzani IRCCS | B Bartolini, E Giombini, F Messina, M Rueca, O Butera, CEM Gruber, E Giardina, MR Capobianchi, A Di Caro |
| EPI_ISL_1079356 | Ospedale S.M. Goretti di Latina - ASL Latina presidio ospedaliero Nord | INMI Lazzaro Spallanzani IRCCS | F Messina, M Rueca, O Butera, CEM Gruber, B Bartolini, E Giombini, L Corso, L Di Biase, MR Capobianchi, A Di Caro |
| EPI_ISL_1079358 | Ospedale S.M. Goretti di Latina - ASL Latina presidio ospedaliero Nord | INMI Lazzaro Spallanzani IRCCS | CEM Gruber, B Bartolini, E Giombini, F Messina, O Butera, M Rueca, L Corso, A Lucci, MR Capobianchi, A Di Caro |
| EPI_ISL_1079359 | Fondazione Policlinico Tor Vergata | INMI Lazzaro Spallanzani IRCCS | CEM Gruber, B Bartolini, E Giombini, F Messina, M Rueca, O Butera, S Grelli, M Ciotti, C D'Agostini, A Di Caro, MR Capobianchi |
| EPI_ISL_1079360 | Ospedale "F. Spaziani" Frosinone | INMI Lazzaro Spallanzani IRCCS | F Messina, M Rueca, O Butera, CEM Gruber, B Bartolini, E Giombini, C. Sias, R Pulselli, MR Capobianchi, A Di Caro |
| EPI_ISL_1079362 | Azienda Ospedaliera San Camillo Forlanini | INMI Lazzaro Spallanzani IRCCS | M Rueca, O Butera, CEM Gruber, B Bartolini, E Giombini, F Messina, D Gallone, A D'Agostino, A Di Caro, MR Capobianchi |
| EPI_ISL_1079363 | Azienda Ospedaliera San Camillo Forlanini | INMI Lazzaro Spallanzani IRCCS | B Bartolini, E Giombini, M Rueca, O Butera, CEM Gruber,F Messina, D Gallone, G Meoni, A Di Caro, MR Capobianchi |
| EPI_ISL_1079364 | Ospedale San Camillo De Lellis di Rieti | INMI Lazzaro Spallanzani IRCCS | B Bartolini, E Giombini, F Messina, M Rueca, O Butera, CEM Gruber, S Venarubea, A De Luca, MR Capobianchi, A Di Caro |
| EPI_ISL_1079366 | Ospedale San Camillo De Lellis di Rieti | INMI Lazzaro Spallanzani IRCCS | E Giombini, F Messina, M Rueca, O Butera, CEM Gruber, B Bartolini, A De Luca, L Casertano, A Di Caro, MR Capobianchi |
| EPI_ISL_1079368 | Ospedale Sandro Pertini ASL Roma2 | INMI Lazzaro Spallanzani IRCCS | F Messina, M Rueca, O Butera, CEM Gruber, B Bartolini, E Giombini, MC Cava, G Cappiello, MR Capobianchi, A Di Caro |
| EPI_ISL_1079370 | Ospedale Sandro Pertini ASL Roma2 | INMI Lazzaro Spallanzani IRCCS | M Rueca, O Butera, CEM Gruber, B Bartolini, E Giombini, F Messina, MC Cava, S Romano, A Di Caro, MR Capobianchi |
| EPI_ISL_1079371 | Fondazione Policlinico Tor Vergata | INMI Lazzaro Spallanzani IRCCS | CEM Gruber, B Bartolini, E Giombini, F Messina, O Butera, M Rueca, S Grelli, P Paba, D Ombres, MR Capobianchi, A Di Caro |
| EPI_ISL_1079373 | Fondazione Policlinico Tor Vergata | INMI Lazzaro Spallanzani IRCCS | E Giombini, F Messina, M Rueca, O Butera, CEM Gruber, B Bartolini, S Grelli, C Fontana, D Di Cave, A Di Caro, MR Capobianchi |
| EPI_ISL_1079376 | San Gallicano Dermatological Institute I.F.O. | INMI Lazzaro Spallanzani IRCCS | B Bartolini, E Giombini, F Messina, M Rueca, O Butera, CEM Gruber, F Pimpinelli, F Ensoli, G D'Agosto, MR Capobianchi, A Di Caro |
| EPI_ISL_1079378 | San Gallicano Dermatological Institute I.F.O. | INMI Lazzaro Spallanzani IRCCS | E Giombini, F Messina, M Rueca, O Butera, CEM Gruber, B Bartolini, F Ensoli, F Pimpinelli, E Trento, A Di Caro, MR Capobianchi |
| EPI_ISL_1079380 | Sant'Eugenio/CTO ASL Roma 2 | INMI Lazzaro Spallanzani IRCCS | F Messina, M Rueca, O Butera, CEM Gruber, B Bartolini, E Giombini, F Bondanini, GC Cocciolillo, MR Capobianchi, A Di Caro |
| EPI_ISL_1079382 | Sant'Eugenio/CTO ASL Roma 2 | INMI Lazzaro Spallanzani IRCCS | M Rueca, O Butera, CEM Gruber, B Bartolini, E Giombini, F Messina, F Bondanini, C Disegni, A Di Caro, MR Capobianchi |
| EPI_ISL_1079383 | Azienda Ospedaliera San Camillo Forlanini | INMI Lazzaro Spallanzani IRCCS | CEM Gruber, B Bartolini, O Butera, E Giombini, F Messina, M Rueca, R.Sperning, D Gallone, MR Capobianchi, A Di Caro |
| EPI_ISL_1079386 | Azienda Ospedaliera San Camillo Forlanini | INMI Lazzaro Spallanzani IRCCS | E Giombini, M Rueca, O Butera, CEM Gruber, B Bartolini,F Messina, D Gallone, M Irno Consalvo, A Di Caro, MR Capobianchi |
| EPI_ISL_1079388 | University Hospital Sant'Andrea-Sapienza | INMI Lazzaro Spallanzani IRCCS | E Giombini, F Messina, M Rueca, O Butera, CEM Gruber, B Bartolini, I Santino, M Simmaco, A Di Caro, MR Capobianchi |
| EPI_ISL_1079390 | Laboratorio Genzano - ASL RM 6 | INMI Lazzaro Spallanzani IRCCS | F Messina, M Rueca, O Butera, CEM Gruber, B Bartolini, E Giombini, G Tramini, E Conti, MR Capobianchi, A Di Caro |
| EPI_ISL_1079391 | Laboratorio Genzano - ASL RM 6 | INMI Lazzaro Spallanzani IRCCS | M Rueca, O Butera, CEM Gruber, B Bartolini, E Giombini, F Messina, E Conti, G Tramini, A Di Caro, MR Capobianchi |
| EPI_ISL_1079392 | Policlinico Universitario Campus Bio-Medico | INMI Lazzaro Spallanzani IRCCS | O Butera, CEM Gruber, B Bartolini, E Giombini, F Messina, M Rueca, E Riva, S Angeletti, MR Capobianchi, A Di Caro |
| EPI_ISL_1079394 | Policlinico Universitario Campus Bio-Medico | INMI Lazzaro Spallanzani IRCCS | CEM Gruber, B Bartolini, E Giombini, F Messina, M Rueca, O Butera, M Fogolari, L De Florio, E Riva, A Di Caro, MR Capobianchi |
| EPI_ISL_1079395 | IRCCS San Raffaele | INMI Lazzaro Spallanzani IRCCS | E Giombini, F Messina, M Rueca, O Butera, CEM Gruber, B Bartolini, D Limongi, D Russo, A Di Caro, MR Capobianchi |
| EPI_ISL_1079398 | Ospedale "F. Spaziani" Frosinone | INMI Lazzaro Spallanzani IRCCS | F Messina, M Rueca, O Butera, CEM Gruber, B Bartolini, E Giombini, R Pulselli, G Brocco, MR Capobianchi, A Di Caro |
| EPI_ISL_1079400 | Ospedale San Filippo Neri | INMI Lazzaro Spallanzani IRCCS | M Rueca, O Butera, F Messina, CEM Gruber, B Bartolini, E Giombini, M Melandri, ML Schiavone, A Tamburro, A Di Caro, MR Capobianchi |
| EPI_ISL_1079992 | Ospedale S.M. Goretti di Latina - ASL Latina presidio ospedaliero Nord | INMI Lazzaro Spallanzani IRCCS | CEM Gruber, B Bartolini, E Giombini, F Messina, O Butera, M Rueca, S Pignalosa, L Corso, MR Capobianchi, A Di Caro |
| EPI_ISL_1079993 | Ospedale S.M. Goretti di Latina - ASL Latina presidio ospedaliero Nord | INMI Lazzaro Spallanzani IRCCS | B Bartolini, E Giombini, F Messina, O Butera, M Rueca, CEM Gruber, L Di Biase, A Lucci, A Di Caro, MR Capobianchi |
| EPI_ISL_1079994 | Azienda Ospedaliera San Camillo Forlanini | INMI Lazzaro Spallanzani IRCCS | O Butera, E Giombini, F Messina, M Rueca, CEM Gruber, B Bartolini, G Parisi, MI Consalvo, MR Capobianchi, A Di Caro |
| EPI_ISL_1079995 | INMI Lazzaro Spallanzani IRCCS | INMI Lazzaro Spallanzani IRCCS | E Giombini, F Messina, M Rueca, O Butera, CEM Gruber, B Bartolini, A Di Caro, MR Capobianchi |
| EPI_ISL_1079996 | Ospedale Sandro Pertini ASL Roma2 | INMI Lazzaro Spallanzani IRCCS | F Messina, M Rueca, O Butera, CEM Gruber, B Bartolini, E Giombini, MC Cava, G Cappiello, MR Capobianchi, A Di Caro |
| EPI_ISL_1079998 | Ospedale San Filippo Neri | INMI Lazzaro Spallanzani IRCCS | M Rueca, O Butera, F Messina, CEM Gruber, B Bartolini, E Giombini, M Meledandri, ML Schiavone, MR Capobianchi |
| EPI_ISL_1079999 | Ospedale San Filippo Neri | INMI Lazzaro Spallanzani IRCCS | CEM Gruber, B Bartolini, E Giombini, F Messina, O Butera, M Rueca, ML Schiavone, A Tamburro, A Di Caro, MR Capobianchi |
| EPI_ISL_1080001 | Ospedale Sandro Pertini ASL Roma2 | INMI Lazzaro Spallanzani IRCCS | B Bartolini, E Giombini, F Messina, O Butera, M Rueca, CEM Gruber, R Longo, V Michela, MR Capobianchi, A Di Caro |
| EPI_ISL_1080464, EPI_ISL_1080465, EPI_ISL_1080466, EPI_ISL_1080467, EPI_ISL_1080468, EPI_ISL_1080469, EPI_ISL_1080470, EPI_ISL_1080471, EPI_ISL_1080472, EPI_ISL_1080473, EPI_ISL_1080474, EPI_ISL_1080475, EPI_ISL_1080476, EPI_ISL_1080477, EPI_ISL_1080478, EPI_ISL_1080479, EPI_ISL_1080481, EPI_ISL_1080482, EPI_ISL_1080483, EPI_ISL_1080484, EPI_ISL_1080485, EPI_ISL_1080486, EPI_ISL_1080487, EPI_ISL_1080488, EPI_ISL_1080489, EPI_ISL_1080490, EPI_ISL_1080491, EPI_ISL_1080492, EPI_ISL_1080493, EPI_ISL_1080494, EPI_ISL_1080495 | | | |
| see above | Istituto Zooprofilattico Sperimentale del Mezzogiorno | Istituto Zooprofilattico Sperimentale del Mezzogiorno-U.O.C. Virologia | Maurizio Viscardi, Lorena Cardillo, Loredana Cozzolino, Pellegrino Cerino, Massimo Zollo,Giovanna Fusco, Esterina De Carlo, Antonio Limone |
| EPI_ISL_1080605, EPI_ISL_1080606, EPI_ISL_1080607, EPI_ISL_1080608, EPI_ISL_1080609, EPI_ISL_1080610, EPI_ISL_1080611, EPI_ISL_1080612, EPI_ISL_1080613, EPI_ISL_1080614, EPI_ISL_1080615, EPI_ISL_1080616, EPI_ISL_1080617, EPI_ISL_1080618, EPI_ISL_1080619, EPI_ISL_1080620, EPI_ISL_1080621, EPI_ISL_1080622, EPI_ISL_1080623, EPI_ISL_1080624, EPI_ISL_1080625, EPI_ISL_1080626, EPI_ISL_1080627, EPI_ISL_1080628, EPI_ISL_1080629, EPI_ISL_1080630, EPI_ISL_1080631, EPI_ISL_1080632, EPI_ISL_1080633, EPI_ISL_1080634, EPI_ISL_1080635, EPI_ISL_1080636, EPI_ISL_1080637, EPI_ISL_1080638, EPI_ISL_1080639, EPI_ISL_1080640, EPI_ISL_1080641, EPI_ISL_1080642, EPI_ISL_1080643, EPI_ISL_1080644, EPI_ISL_1080645, EPI_ISL_1080646, EPI_ISL_1080647, EPI_ISL_1080648, EPI_ISL_1080649, EPI_ISL_1080650, EPI_ISL_1080651, EPI_ISL_1080652, EPI_ISL_1080653, EPI_ISL_1080654, EPI_ISL_1080655, EPI_ISL_1080656, EPI_ISL_1080657, EPI_ISL_1080658, EPI_ISL_1080659, EPI_ISL_1080660, EPI_ISL_1080661, EPI_ISL_1080662, EPI_ISL_1080663, EPI_ISL_1080664, EPI_ISL_1080665, EPI_ISL_1080666, EPI_ISL_1080667, EPI_ISL_1080668, EPI_ISL_1080669, EPI_ISL_1080670, EPI_ISL_1080671, EPI_ISL_1080672, EPI_ISL_1080673, EPI_ISL_1080674, EPI_ISL_1080675, EPI_ISL_1080676, EPI_ISL_1080677, EPI_ISL_1080678, EPI_ISL_1080679, EPI_ISL_1080681, EPI_ISL_1080682, EPI_ISL_1080683, EPI_ISL_1080684, EPI_ISL_1080685, EPI_ISL_1080686, EPI_ISL_1080687, EPI_ISL_1080688, EPI_ISL_1080689, EPI_ISL_1080690, EPI_ISL_1080691, EPI_ISL_1080692, EPI_ISL_1080693, EPI_ISL_1080694, EPI_ISL_1080695, EPI_ISL_1080696, EPI_ISL_1080697, EPI_ISL_1080698, EPI_ISL_1080699, EPI_ISL_1080700, EPI_ISL_1080701, EPI_ISL_1080702, EPI_ISL_1080703, EPI_ISL_1080704, EPI_ISL_1080705, EPI_ISL_1080706, EPI_ISL_1080707, EPI_ISL_1080708, EPI_ISL_1080709, EPI_ISL_1080710, EPI_ISL_1080711, EPI_ISL_1080712, EPI_ISL_1080713, EPI_ISL_1080714, EPI_ISL_1080715, EPI_ISL_1080716, EPI_ISL_1080717, EPI_ISL_1080718, EPI_ISL_1080719, EPI_ISL_1080720, EPI_ISL_1080721, EPI_ISL_1080722, EPI_ISL_1080723, EPI_ISL_1080724, EPI_ISL_1080725, EPI_ISL_1080726, EPI_ISL_1080727, EPI_ISL_1080728, EPI_ISL_1080729, EPI_ISL_1080730, EPI_ISL_1080731, EPI_ISL_1080732, EPI_ISL_1080733, EPI_ISL_1080734, EPI_ISL_1080735, EPI_ISL_1080736, EPI_ISL_1080737, EPI_ISL_1080738, EPI_ISL_1080739, EPI_ISL_1080740, EPI_ISL_1080741, EPI_ISL_1080742, EPI_ISL_1080743, EPI_ISL_1080744, EPI_ISL_1080745, EPI_ISL_1080746, EPI_ISL_1080747, EPI_ISL_1080748 | | | |
| see above | ASL Napoli 1 Centro | AMES Centro Polidiagnostico Strumentale S.r.l. | "Giovanni Savarese, Raffaella Ruggiero, Eloisa Evangelista, Antonella Di Carlo, Luisa Circelli, Luigi D'Amore, Roberto Sirica, Antonio Fico" |
| EPI_ISL_1081953 | Laboratorio di Riferimento Regionale della Sicilia Occidentale | Laboratorio di Riferimento Regionale della Sicilia Occidentale | Fabio Tramuto, Carmelo Massimo Maida, Daniela Di Naro, Giulia Randazzo, Walter Mazzucco, Giorgio Graziano, Vincenzo Restivo, Claudio Costantino, |

| | | | |
|---|---|---|---|
| | per l'Emergenza COVID-19 | per l'Emergenza COVID-19 | Stefano Giaramida, Francesco Vitale |
| EPI_ISL_1082253 | Laboratorio Biologia Molecolare Sars Cov2 - UOC Laboratorio Analisi - Servizio Medicina di Laboratorio, Ospedale "San Francesco" - ATS-ASSL Nuoro | Laboratorio specialistico UOC Ematologia - Ospedale "San Francesco" - ATS-ASSL Nuoro | Giovanna Piras, Rosanna Asproni, Paolo Malune, Maura Fiamma, Maria Itria Monne, Angelo Domenico Palmas, Iana Lo Maglio, Giuseppe Mameli |
| EPI_ISL_1082254 | Virologia, Dipartimento di Scienze Biomediche, Università di Sassari, Viale San Pietro, 43/B - Sassari | Laboratorio specialistico UOC Ematologia - Ospedale "San Francesco" - ATS-ASSL Nuoro | Giovanna Piras, Asproni Rosanna, Paolo Malune, Maria Itria Monne, Angelo Domenico Palmas, Caterina Serra, Elena Rimini, Salvatore Rubino |
| EPI_ISL_1082292, EPI_ISL_1082293, EPI_ISL_1082295 | INT Fondazione Pascale | INT Fondazione Pascale | INT Fondazione Pascale |
| EPI_ISL_1082454, EPI_ISL_1082455, EPI_ISL_1082456, EPI_ISL_1082457, EPI_ISL_1082458, EPI_ISL_1082459, EPI_ISL_1082460, EPI_ISL_1082461, EPI_ISL_1082462, EPI_ISL_1082463, EPI_ISL_1082464, EPI_ISL_1082465, EPI_ISL_1082466, EPI_ISL_1082467, EPI_ISL_1082468, EPI_ISL_1082469, EPI_ISL_1082470, EPI_ISL_1082471, EPI_ISL_1082472, EPI_ISL_1082473, EPI_ISL_1082474, EPI_ISL_1082475, EPI_ISL_1082476, EPI_ISL_1082477, EPI_ISL_1082478, EPI_ISL_1082479, EPI_ISL_1082480, EPI_ISL_1082481, EPI_ISL_1082482, EPI_ISL_1082483, EPI_ISL_1082484, EPI_ISL_1082485, EPI_ISL_1082486, EPI_ISL_1082487, EPI_ISL_1082488, EPI_ISL_1082489, EPI_ISL_1082490, EPI_ISL_1082491, EPI_ISL_1082492, EPI_ISL_1082493, EPI_ISL_1082494, EPI_ISL_1082495, EPI_ISL_1082496, EPI_ISL_1082497, EPI_ISL_1082498, EPI_ISL_1082499, EPI_ISL_1082500, EPI_ISL_1082501, EPI_ISL_1082502, EPI_ISL_1082503, EPI_ISL_1082504, EPI_ISL_1082505, EPI_ISL_1082506, EPI_ISL_1082507, EPI_ISL_1082508, EPI_ISL_1082509, EPI_ISL_1082510, EPI_ISL_1082511, EPI_ISL_1082512, EPI_ISL_1082513, EPI_ISL_1082514, EPI_ISL_1082515, EPI_ISL_1082516, EPI_ISL_1082517, EPI_ISL_1082518, EPI_ISL_1082519, EPI_ISL_1082520, EPI_ISL_1082521, EPI_ISL_1082522, EPI_ISL_1082523, EPI_ISL_1082524, EPI_ISL_1082525, EPI_ISL_1085025, EPI_ISL_1085026, EPI_ISL_1085027, EPI_ISL_1085028, EPI_ISL_1085029, EPI_ISL_1085030, EPI_ISL_1085032, EPI_ISL_1085033, EPI_ISL_1085034, EPI_ISL_1085035, EPI_ISL_1085036, EPI_ISL_1085037, EPI_ISL_1085038, EPI_ISL_1085039, EPI_ISL_1085040, EPI_ISL_1085041, EPI_ISL_1085042, EPI_ISL_1085043, EPI_ISL_1085044, EPI_ISL_1085045, EPI_ISL_1085046, EPI_ISL_1085047, EPI_ISL_1085048, EPI_ISL_1085049, EPI_ISL_1085050, EPI_ISL_1085051, EPI_ISL_1085052, EPI_ISL_1085053, EPI_ISL_1085054, EPI_ISL_1085055, EPI_ISL_1085056, EPI_ISL_1085057, EPI_ISL_1085058, EPI_ISL_1085059, EPI_ISL_1085060, EPI_ISL_1085061, EPI_ISL_1085062, EPI_ISL_1085063, EPI_ISL_1085064, EPI_ISL_1085065, EPI_ISL_1085066, EPI_ISL_1085067, EPI_ISL_1085068, EPI_ISL_1085069, EPI_ISL_1085070, EPI_ISL_1085071, EPI_ISL_1085072, EPI_ISL_1085073, EPI_ISL_1085074, EPI_ISL_1085075, EPI_ISL_1085076, EPI_ISL_1085077, EPI_ISL_1085078, EPI_ISL_1085079, EPI_ISL_1085080, EPI_ISL_1085081, EPI_ISL_1085082, EPI_ISL_1085083, EPI_ISL_1085084, EPI_ISL_1085085, EPI_ISL_1085086, EPI_ISL_1085087, EPI_ISL_1085088, EPI_ISL_1085089, EPI_ISL_1085090 | | | |
| see above | ASL Napoli 1 Centro | AMES Centro Polidiagnostico Strumentale S.r.l. | "Giovanni Savarese, Raffaella Ruggiero, Eloisa Evangelista, Antonella Di Carlo, Luisa Circelli, Luigi D'Amore, Roberto Sirica, Nadia Petrillo, Monica Ianniello, Antonio Fico" |
| EPI_ISL_1085094 | Laboratory of Infectious Diseases, Department of Biomedical and Clinical Sciences L. Sacco, University of Milan | Laboratory of Infectious Diseases, Department of Biomedical and Clinical Sciences L. Sacco, University of Milan | Alessia Lai, Annalisa Bergna, Carla Della Ventura, Claudia Balotta, Massimo Galli, Gianguglielmo Zehender on behalf of SARS-CoV-2 ITALIAN RESEARCH ENTERPRISE-(SCIRE) Collaborative Group |
| EPI_ISL_1085096, EPI_ISL_1085097, EPI_ISL_1085098, EPI_ISL_1085099, EPI_ISL_1085100, EPI_ISL_1085101, EPI_ISL_1085102, EPI_ISL_1085103, EPI_ISL_1085104, EPI_ISL_1085105, EPI_ISL_1085106, EPI_ISL_1085107, EPI_ISL_1085108, EPI_ISL_1085109, EPI_ISL_1085110, EPI_ISL_1085111, EPI_ISL_1085112, EPI_ISL_1085113, EPI_ISL_1085114, EPI_ISL_1085115, EPI_ISL_1085116, EPI_ISL_1085117, EPI_ISL_1085118, EPI_ISL_1085119, EPI_ISL_1085120, EPI_ISL_1085121, EPI_ISL_1085122, EPI_ISL_1085123, EPI_ISL_1085124, EPI_ISL_1085125, EPI_ISL_1085126, EPI_ISL_1085127, EPI_ISL_1085128, EPI_ISL_1085129, EPI_ISL_1085130, EPI_ISL_1085131, EPI_ISL_1085132, EPI_ISL_1085133, EPI_ISL_1085134, EPI_ISL_1085135, EPI_ISL_1085136, EPI_ISL_1085137, EPI_ISL_1085138, EPI_ISL_1085139, EPI_ISL_1085140, EPI_ISL_1085141, EPI_ISL_1085142, EPI_ISL_1085143, EPI_ISL_1085144, EPI_ISL_1085145, EPI_ISL_1085146, EPI_ISL_1085147, EPI_ISL_1085148, EPI_ISL_1085149, EPI_ISL_1085150, EPI_ISL_1085151, EPI_ISL_1085152, EPI_ISL_1085153, EPI_ISL_1085154, EPI_ISL_1085155, EPI_ISL_1085156, EPI_ISL_1085157, EPI_ISL_1085158, EPI_ISL_1085159, EPI_ISL_1085160 | | | |
| see above | ASL Napoli 1 Centro | AMES Centro Polidiagnostico Strumentale S.r.l. | "Giovanni Savarese, Raffaella Ruggiero, Eloisa Evangelista, Antonella Di Carlo, Luisa Circelli, Luigi D'Amore, Roberto Sirica, Nadia Petrillo, Monica Ianniello, Antonio Fico" |
| EPI_ISL_1085162, EPI_ISL_1085164, EPI_ISL_1085165, EPI_ISL_1085167, EPI_ISL_1085169, EPI_ISL_1085170, EPI_ISL_1085173, EPI_ISL_1085174 | Laboratory of Infectious Diseases, Department of Biomedical and Clinical Sciences L. Sacco, University of Milan | Laboratory of Infectious Diseases, Department of Biomedical and Clinical Sciences L. Sacco, University of Milan | Alessia Lai, Annalisa Bergna, Carla Della Ventura, Claudia Balotta, Massimo Galli, Gianguglielmo Zehender on behalf of SARS-CoV-2 ITALIAN RESEARCH ENTERPRISE-(SCIRE) Collaborative Group |
| EPI_ISL_1085176, EPI_ISL_1085177, EPI_ISL_1085178, EPI_ISL_1085179, EPI_ISL_1085180, EPI_ISL_1085181, EPI_ISL_1085182, EPI_ISL_1085183, EPI_ISL_1085184, EPI_ISL_1085185, EPI_ISL_1085186, EPI_ISL_1085187, EPI_ISL_1085188, EPI_ISL_1085189, EPI_ISL_1085190, EPI_ISL_1085191, EPI_ISL_1085192, EPI_ISL_1085193, EPI_ISL_1085194, EPI_ISL_1085195, EPI_ISL_1085196, EPI_ISL_1085197, EPI_ISL_1085198, EPI_ISL_1085199, EPI_ISL_1085200, EPI_ISL_1085201, EPI_ISL_1085202, EPI_ISL_1085203, EPI_ISL_1085204, EPI_ISL_1085205, EPI_ISL_1085206, EPI_ISL_1085207, EPI_ISL_1085208, EPI_ISL_1085209, EPI_ISL_1085210, EPI_ISL_1085211, EPI_ISL_1085212, EPI_ISL_1085213, EPI_ISL_1085214, EPI_ISL_1085215, EPI_ISL_1085216, EPI_ISL_1085217, EPI_ISL_1085218, EPI_ISL_1085219, EPI_ISL_1085220, EPI_ISL_1085221, EPI_ISL_1085222, EPI_ISL_1085223, EPI_ISL_1085224, EPI_ISL_1085225, EPI_ISL_1085226, EPI_ISL_1085227, EPI_ISL_1085228, EPI_ISL_1085229 | | | |
| see above | ASL Napoli 1 Centro | AMES Centro Polidiagnostico Strumentale S.r.l. | "Giovanni Savarese, Raffaella Ruggiero, Eloisa Evangelista, Antonella Di Carlo, Luisa Circelli, Luigi D'Amore, Roberto Sirica, Nadia Petrillo, Monica Ianniello, Antonio Fico" |
| EPI_ISL_1085230 | Laboratory of Infectious Diseases, Department of Biomedical and Clinical Sciences L. Sacco, University of Milan | Laboratory of Infectious Diseases, Department of Biomedical and Clinical Sciences L. Sacco, University of Milan | Alessia Lai, Annalisa Bergna, Carla Della Ventura, Claudia Balotta, Massimo Galli, Gianguglielmo Zehender on behalf of SARS-CoV-2 ITALIAN RESEARCH ENTERPRISE-(SCIRE) Collaborative Group |
| EPI_ISL_1087353, EPI_ISL_1087354, EPI_ISL_1087355, EPI_ISL_1087356, EPI_ISL_1087357, EPI_ISL_1087358, EPI_ISL_1087359, EPI_ISL_1087360, EPI_ISL_1087361, EPI_ISL_1087362, EPI_ISL_1087363, EPI_ISL_1087364, EPI_ISL_1087365, EPI_ISL_1087366, EPI_ISL_1087367, EPI_ISL_1087368, EPI_ISL_1087369, EPI_ISL_1087370, EPI_ISL_1087371, EPI_ISL_1087372, EPI_ISL_1087373, EPI_ISL_1087374, EPI_ISL_1087375, EPI_ISL_1087376, EPI_ISL_1087377, EPI_ISL_1087378, EPI_ISL_1087379, EPI_ISL_1087380, EPI_ISL_1087381, EPI_ISL_1087382, EPI_ISL_1087383, EPI_ISL_1087384, EPI_ISL_1087385, EPI_ISL_1087386, EPI_ISL_1087387, EPI_ISL_1087388, EPI_ISL_1087389, EPI_ISL_1087390, EPI_ISL_1087391, EPI_ISL_1087392, EPI_ISL_1087393, EPI_ISL_1087394, EPI_ISL_1087395, EPI_ISL_1087396, EPI_ISL_1087397, EPI_ISL_1087398, EPI_ISL_1087399, EPI_ISL_1087400, EPI_ISL_1087401, EPI_ISL_1087402, EPI_ISL_1087403, EPI_ISL_1087404, EPI_ISL_1087405, EPI_ISL_1087406, EPI_ISL_1087407, EPI_ISL_1087408, EPI_ISL_1087409, EPI_ISL_1087410, EPI_ISL_1087411, EPI_ISL_1087412, EPI_ISL_1087413, EPI_ISL_1088008, EPI_ISL_1088009, EPI_ISL_1088010, EPI_ISL_1088011, EPI_ISL_1088012, EPI_ISL_1088013, EPI_ISL_1088014, EPI_ISL_1088015, EPI_ISL_1088016, EPI_ISL_1088017, EPI_ISL_1088018, EPI_ISL_1088019, EPI_ISL_1088020, EPI_ISL_1088021, EPI_ISL_1088022, EPI_ISL_1088023, EPI_ISL_1088024, EPI_ISL_1088025, EPI_ISL_1088026, EPI_ISL_1088027, EPI_ISL_1088028, EPI_ISL_1088029, EPI_ISL_1088030, EPI_ISL_1088031, EPI_ISL_1088032, EPI_ISL_1088033, EPI_ISL_1088034, EPI_ISL_1088035, EPI_ISL_1088036, EPI_ISL_1088037, EPI_ISL_1088038, EPI_ISL_1088039, EPI_ISL_1088040, EPI_ISL_1088041, EPI_ISL_1088042, EPI_ISL_1088043, EPI_ISL_1088044, EPI_ISL_1088045, EPI_ISL_1088046, EPI_ISL_1088047, EPI_ISL_1088048, EPI_ISL_1088049, EPI_ISL_1088050, EPI_ISL_1088051, EPI_ISL_1088052, EPI_ISL_1088053, EPI_ISL_1088054, EPI_ISL_1088055, EPI_ISL_1088056, EPI_ISL_1088057, EPI_ISL_1088058, EPI_ISL_1088059, EPI_ISL_1088060, EPI_ISL_1088061, EPI_ISL_1088062, EPI_ISL_1088063, EPI_ISL_1088064, EPI_ISL_1088065, EPI_ISL_1088066, EPI_ISL_1088067, EPI_ISL_1088068, EPI_ISL_1088069, EPI_ISL_1088070, EPI_ISL_1088071, EPI_ISL_1088072, EPI_ISL_1088073, EPI_ISL_1088074, EPI_ISL_1088075, EPI_ISL_1088076 | | | |
| see above | ASL Napoli 1 Centro | AMES Centro Polidiagnostico Strumentale S.r.l. | "Giovanni Savarese, Raffaella Ruggiero, Eloisa Evangelista, Antonella Di Carlo, Luisa Circelli, Luigi D'Amore, Roberto Sirica, Nadia Petrillo, Monica Ianniello, Antonio Fico" |
| EPI_ISL_1091275 | Ospedale S.M. Goretti di Latina - ASL Latina presidio ospedaliero Nord | INMI Lazzaro Spallanzani IRCCS | CEM Gruber, B Bartolini, E Giombini, F Messina, O Butera, M Rueca, S Pignalosa, L Corso, MR Capobianchi, A Di Caro |
| EPI_ISL_1091276 | INMI Lazzaro Spallanzani IRCCS | INMI Lazzaro Spallanzani IRCCS | O Butera, E Giombini, F Messina, M Rueca, CEM Gruber, B Bartolini, MR Capobianchi, A Di Caro |
| EPI_ISL_1091277 | INMI Lazzaro Spallanzani IRCCS | INMI Lazzaro Spallanzani IRCCS | CEM Gruber, B Bartolini, O Butera, E Giombini, F Messina, M Rueca, A Di Caro, MR Capobianchi |
| EPI_ISL_1091278 | INMI Lazzaro Spallanzani IRCCS | INMI Lazzaro Spallanzani IRCCS | B Bartolini, E Giombini, F Messina, O Butera, M Rueca, CEM Gruber, A Di Caro, MR Capobianchi |
| EPI_ISL_1091279 | INMI Lazzaro Spallanzani IRCCS | INMI Lazzaro Spallanzani IRCCS | M Rueca, CEM Gruber, B Bartolini, O Butera, E Giombini, F Messina, A Di Caro, MR Capobianchi |
| EPI_ISL_1091280 | Azienda Ospedaliera San Camillo Forlanini | INMI Lazzaro Spallanzani IRCCS | F Messina, M Rueca, O Butera, CEM Gruber, B Bartolini, E Giombini, G Parisi, D Gallone, MR Capobianchi, A Di Caro |
| EPI_ISL_1091281 | Azienda Ospedaliera San Camillo Forlanini | INMI Lazzaro Spallanzani IRCCS | M Rueca, CEM Gruber, B Bartolini, O Butera, E Giombini, F Messina, G Parisi, A D'Agostino, A Di Caro, MR Capobianchi |
| EPI_ISL_1091282 | Azienda Ospedaliera San Camillo Forlanini | INMI Lazzaro Spallanzani IRCCS | O Butera, E Giombini, F Messina, M Rueca, CEM Gruber, B Bartolini, G Parisi, ML Guarino, MR Capobianchi, A Di Caro |
| EPI_ISL_1091283 | Azienda Ospedaliera San Camillo Forlanini | INMI Lazzaro Spallanzani IRCCS | E Giombini, F Messina, M Rueca, O Butera, CEM Gruber, B Bartolini, G Parisi, MI Consalvo, A Di Caro, MR Capobianchi |
| EPI_ISL_1091284 | Azienda Ospedaliera San Camillo Forlanini | INMI Lazzaro Spallanzani IRCCS | B Bartolini, E Giombini, F Messina, O Butera, M Rueca, CEM Gruber, G Parisi, F Basile, MR Capobianchi, A Di Caro |
| EPI_ISL_1091285 | Azienda Ospedaliera San Camillo Forlanini | INMI Lazzaro Spallanzani IRCCS | F Messina, M Rueca, O Butera, CEM Gruber, B Bartolini, E Giombini, G Parisi, G Meoni, A Di Caro, MR Capobianchi |
| EPI_ISL_1091286 | Azienda Ospedaliera San Camillo Forlanini | INMI Lazzaro Spallanzani IRCCS | CEM Gruber, B Bartolini, E Giombini, F Messina, O Butera, M Rueca, G Parisi, ML Guarino, A Di Caro, MR Capobianchi |
| EPI_ISL_1091287 | Ospedale Sandro Pertini ASL Roma2 | INMI Lazzaro Spallanzani IRCCS | CEM Gruber, B Bartolini, O Butera, E Giombini, F Messina, M Rueca, MC Cava, R Longo, A Di Caro, MR Capobianchi |
| EPI_ISL_1091288 | Ospedale Sandro Pertini ASL Roma3 | INMI Lazzaro Spallanzani IRCCS | B Bartolini, E Giombini, F Messina, O Butera, M Rueca, CEM Gruber, G Cappiello, V Michela, MR Capobianchi, A Di Caro |
| EPI_ISL_1091289 | Ospedale Sandro Pertini ASL Roma4 | INMI Lazzaro Spallanzani IRCCS | O Butera, E Giombini, F Messina, M Rueca, CEM Gruber, B Bartolini, MC Cava, S Romano, MR Capobianchi, A Di Caro |
| EPI_ISL_1091290 | Ospedale San Filippo Neri | INMI Lazzaro Spallanzani IRCCS | M Rueca, CEM Gruber, B Bartolini, O Butera, E Giombini, F Messina, ML Schiavone, A Tamburro, MR Capobianchi, A Di Caro |
| EPI_ISL_1091291 | Ospedale San Filippo Neri | INMI Lazzaro Spallanzani IRCCS | F Messina, M Rueca, O Butera, CEM Gruber, B Bartolini, E Giombini, A Tamburro, M Meledandri, MR Capobianchi, A Di Caro |
| EPI_ISL_1091787 | San Gallicano Dermatological Institute I.F.O. | INMI Lazzaro Spallanzani IRCCS | B Bartolini, E Giombini, F Messina, M Rueca, O Butera, CEM Gruber, F Pimpinelli, F Ensoli, G Prignano, A Di Caro, MR Capobianchi |
| EPI_ISL_1091788 | INMI Lazzaro Spallanzani IRCCS | INMI Lazzaro Spallanzani IRCCS | E Giombini, F Messina, M Rueca, O Butera, CEM Gruber, B Bartolini, A Di Caro, MR Capobianchi |
| EPI_ISL_1091789 | INMI Lazzaro Spallanzani IRCCS | INMI Lazzaro Spallanzani IRCCS | O Butera, CEM Gruber, B Bartolini, E Giombini, F Messina, M Rueca, MR Capobianchi, A Di Caro |

| | | | |
|---|---|---|---|
| EPI_ISL_1091790 | San Gallicano Dermatological Institute I.F.O. | INMI Lazzaro Spallanzani IRCCS | M Rueca, O Butera, CEM Gruber, B Bartolini, E Giombini, F Messina, F Pimpinelli, F Ensoli, G D'Agosto, A Di Caro, MR Capobianchi |
| EPI_ISL_1091791 | Laboratorio Genzano - ASL RM 6 | INMI Lazzaro Spallanzani IRCCS | O Butera, CEM Gruber, B Bartolini, E Giombini, F Messina, M Rueca, G Tramini, E Conti, MR Capobianchi, A Di Caro |
| EPI_ISL_1091792 | Laboratorio Genzano - ASL RM 6 | INMI Lazzaro Spallanzani IRCCS | B Bartolini, E Giombini, F Messina, M Rueca, O Butera, CEM Gruber, G Tramini, E Conti, A Di Caro, MR Capobianchi |
| EPI_ISL_1091793 | Laboratorio Genzano - ASL RM 6 | INMI Lazzaro Spallanzani IRCCS | CEM Gruber, B Bartolini, E Giombini, F Messina, M Rueca, O Butera, G Tramini, E Conti, MR Capobianchi, A Di Caro |
| EPI_ISL_1091794 | Ospedale "F. Spaziani" Frosinone | INMI Lazzaro Spallanzani IRCCS | F Messina, M Rueca, O Butera, CEM Gruber, B Bartolini, E Giombini, R Pulselli, C Sias, G Brocco, MR Capobianchi, A Di Caro |
| EPI_ISL_1091795 | Laboratorio Genzano - ASL RM 6 | INMI Lazzaro Spallanzani IRCCS | M Rueca, O Butera, CEM Gruber, B Bartolini, E Giombini, F Messina, G Tramini, E Conti, A Di Caro, MR Capobianchi |
| EPI_ISL_1091796 | Ospedale S.M. Goretti di Latina - ASL Latina presidio ospedaliero Nord | INMI Lazzaro Spallanzani IRCCS | B Bartolini, E Giombini, F Messina, M Rueca, O Butera, CEM Gruber, A Lucci, S Pignalosa, L Corso, A Di Caro, MR Capobianchi |
| EPI_ISL_1091797 | Azienda Ospedaliera San Camillo Forlanini | INMI Lazzaro Spallanzani IRCCS | M Rueca, O Butera, CEM Gruber, B Bartolini, E Giombini, F Messina, G Parisi, D Gallone, F Basile, A Di Caro, MR Capobianchi |
| EPI_ISL_1091798 | Synlab Lazio S.r.l. | INMI Lazzaro Spallanzani IRCCS | E Giombini, F Messina, M Rueca, O Butera, CEM Gruber, B Bartolini, E Trappolini, SA Santini, A Di Caro, MR Capobianchi |
| EPI_ISL_1093477, EPI_ISL_1093478, EPI_ISL_1093479, EPI_ISL_1093480, EPI_ISL_1093481, EPI_ISL_1093482, EPI_ISL_1093483, EPI_ISL_1093484, EPI_ISL_1093485, EPI_ISL_1093486, EPI_ISL_1093487, EPI_ISL_1093488, EPI_ISL_1093489, EPI_ISL_1093490, EPI_ISL_1093491 | | | |
| see above | Center of Advanced Studies and Technology, Molecular Genetics Laboratory | Center of Advanced Studies and Technology, Molecular Genetics Laboratory | Ferrante Rossella, Mandatori Domitilla, De Fabritiis Simone, Damiani Verena, Anaclerio Federico |
| EPI_ISL_1095366, EPI_ISL_1095367, EPI_ISL_1095368, EPI_ISL_1095369, EPI_ISL_1095370, EPI_ISL_1095371, EPI_ISL_1095372, EPI_ISL_1095373, EPI_ISL_1095374, EPI_ISL_1095375, EPI_ISL_1095376, EPI_ISL_1095377, EPI_ISL_1095378, EPI_ISL_1095379, EPI_ISL_1095380, EPI_ISL_1095381, EPI_ISL_1095382, EPI_ISL_1095383 | | | |
| see above | Istituto Zooprofilattico Sperimentale del Mezzogiorno | Lab. Microbiologia e Virologia, Cotugno, A.O. dei Colli | Luigi Atripaldi, Claudia Tiberio, Anna Perfetti, Pellegrino Cerino, Biancamaria Pierri, Maria Concetta Cuomo |
| EPI_ISL_1095384 | Laboratorio Microbiologia e Virologia P.O. Cotugno A.O. dei Colli | Laboratorio Microbiologia e Virologia P.O. Cotugno A.O. dei Colli | Luigi Atripaldi, Claudia Tiberio, Anna Perfetti, Pellegrino Cerino, Biancamaria Pierri, Maria Concetta Cuomo |
| EPI_ISL_1095385, EPI_ISL_1095386, EPI_ISL_1095387, EPI_ISL_1095388, EPI_ISL_1095389, EPI_ISL_1095390, EPI_ISL_1095391, EPI_ISL_1095392, EPI_ISL_1095393, EPI_ISL_1095394, EPI_ISL_1095395, EPI_ISL_1095396, EPI_ISL_1095397, EPI_ISL_1095398, EPI_ISL_1095399, EPI_ISL_1095400, EPI_ISL_1095401, EPI_ISL_1095402, EPI_ISL_1095403, EPI_ISL_1095404, EPI_ISL_1095405, EPI_ISL_1095406, EPI_ISL_1095407 | | | |
| see above | Istituto Zooprofilattico Sperimentale del Mezzogiorno | Lab. Microbiologia e Virologia, Cotugno, A.O. dei Colli | Luigi Atripaldi, Claudia Tiberio, Anna Perfetti, Pellegrino Cerino, Biancamaria Pierri, Maria Concetta Cuomo |
| EPI_ISL_1095449, EPI_ISL_1096228, EPI_ISL_1096229, EPI_ISL_1096230, EPI_ISL_1096231, EPI_ISL_1096232, EPI_ISL_1096233, EPI_ISL_1096342 | Laboratorio Microbiologia e Virologia P.O. Cotugno A.O. dei Colli | Laboratorio Microbiologia e Virologia P.O. Cotugno A.O. dei Colli | Luigi Atripaldi, Claudia Tiberio, Anna Perfetti, Pellegrino Cerino, Biancamaria Pierri, Maria Concetta Cuomo |
| EPI_ISL_1096349, EPI_ISL_1096350, EPI_ISL_1096351, EPI_ISL_1096352, EPI_ISL_1096353, EPI_ISL_1096354, EPI_ISL_1096355, EPI_ISL_1096356, EPI_ISL_1096357, EPI_ISL_1096358, EPI_ISL_1096359, EPI_ISL_1096360, EPI_ISL_1096361, EPI_ISL_1096362, EPI_ISL_1096363, EPI_ISL_1096364, EPI_ISL_1096365, EPI_ISL_1096366, EPI_ISL_1096367, EPI_ISL_1096368, EPI_ISL_1096369, EPI_ISL_1096370, EPI_ISL_1096371, EPI_ISL_1096372, EPI_ISL_1096373, EPI_ISL_1096374, EPI_ISL_1096375, EPI_ISL_1096376, EPI_ISL_1096377, EPI_ISL_1096378, EPI_ISL_1096379, EPI_ISL_1096380, EPI_ISL_1096381, EPI_ISL_1096382, EPI_ISL_1096383, EPI_ISL_1096384, EPI_ISL_1096385, EPI_ISL_1096386, EPI_ISL_1096387, EPI_ISL_1096388, EPI_ISL_1096389, EPI_ISL_1096390, EPI_ISL_1096391, EPI_ISL_1096392, EPI_ISL_1096393, EPI_ISL_1096394, EPI_ISL_1096395, EPI_ISL_1096396, EPI_ISL_1096397, EPI_ISL_1096398, EPI_ISL_1096399, EPI_ISL_1096400, EPI_ISL_1096401, EPI_ISL_1096402, EPI_ISL_1096403, EPI_ISL_1096404, EPI_ISL_1096405, EPI_ISL_1096406, EPI_ISL_1096407, EPI_ISL_1096408, EPI_ISL_1096409, EPI_ISL_1096410, EPI_ISL_1096411, EPI_ISL_1096412, EPI_ISL_1096413, EPI_ISL_1096414, EPI_ISL_1096415, EPI_ISL_1096416, EPI_ISL_1096417, EPI_ISL_1096418, EPI_ISL_1096419, EPI_ISL_1096420, EPI_ISL_1096421, EPI_ISL_1096422, EPI_ISL_1096423, EPI_ISL_1096424, EPI_ISL_1096425, EPI_ISL_1096426, EPI_ISL_1096427, EPI_ISL_1096428, EPI_ISL_1096429, EPI_ISL_1096430, EPI_ISL_1096431, EPI_ISL_1096432, EPI_ISL_1096433, EPI_ISL_1096434, EPI_ISL_1096435, EPI_ISL_1096436, EPI_ISL_1096437, EPI_ISL_1096438, EPI_ISL_1096439, EPI_ISL_1096440, EPI_ISL_1096441, EPI_ISL_1096442, EPI_ISL_1096443, EPI_ISL_1096444, EPI_ISL_1096445, EPI_ISL_1096446, EPI_ISL_1096447, EPI_ISL_1096448, EPI_ISL_1096449, EPI_ISL_1096450, EPI_ISL_1096451, EPI_ISL_1096452, EPI_ISL_1096453, EPI_ISL_1096454, EPI_ISL_1096455, EPI_ISL_1096456, EPI_ISL_1096457, EPI_ISL_1096458, EPI_ISL_1096459, EPI_ISL_1096460, EPI_ISL_1096461, EPI_ISL_1096462, EPI_ISL_1096463, EPI_ISL_1096464, EPI_ISL_1096465, EPI_ISL_1096466, EPI_ISL_1096467, EPI_ISL_1096468, EPI_ISL_1096469, EPI_ISL_1096470, EPI_ISL_1096471, EPI_ISL_1096472, EPI_ISL_1096473, EPI_ISL_1096474, EPI_ISL_1096475, EPI_ISL_1096476, EPI_ISL_1096477, EPI_ISL_1096478, EPI_ISL_1096479, EPI_ISL_1096480, EPI_ISL_1096481, EPI_ISL_1096482, EPI_ISL_1096483, EPI_ISL_1096484, EPI_ISL_1096485, EPI_ISL_1096486, EPI_ISL_1096487, EPI_ISL_1096488, EPI_ISL_1096489, EPI_ISL_1096490, EPI_ISL_1096491, EPI_ISL_1096492, EPI_ISL_1096493, EPI_ISL_1096494, EPI_ISL_1096495, EPI_ISL_1096496, EPI_ISL_1096497, EPI_ISL_1096498, EPI_ISL_1096499, EPI_ISL_1096500, EPI_ISL_1096501, EPI_ISL_1096502, EPI_ISL_1096503, EPI_ISL_1096504, EPI_ISL_1096505, EPI_ISL_1096506, EPI_ISL_1096507, EPI_ISL_1096508, EPI_ISL_1096509, EPI_ISL_1096510, EPI_ISL_1096511, EPI_ISL_1096512, EPI_ISL_1096513, EPI_ISL_1096514, EPI_ISL_1096515, EPI_ISL_1096516, EPI_ISL_1096517, EPI_ISL_1096518, EPI_ISL_1096519, EPI_ISL_1096520, EPI_ISL_1096521, EPI_ISL_1096522, EPI_ISL_1096523, EPI_ISL_1096524, EPI_ISL_1096525, EPI_ISL_1096526, EPI_ISL_1096527, EPI_ISL_1096528, EPI_ISL_1096529, EPI_ISL_1096530, EPI_ISL_1096531, EPI_ISL_1096532, EPI_ISL_1096533, EPI_ISL_1096534, EPI_ISL_1096535, EPI_ISL_1096536, EPI_ISL_1096537, EPI_ISL_1096538, EPI_ISL_1096539, EPI_ISL_1096540, EPI_ISL_1096541, EPI_ISL_1096542, EPI_ISL_1096543, EPI_ISL_1096544, EPI_ISL_1096545, EPI_ISL_1096546, EPI_ISL_1096547, EPI_ISL_1096548, EPI_ISL_1096549, EPI_ISL_1096550, EPI_ISL_1096551, EPI_ISL_1096552, EPI_ISL_1096553, EPI_ISL_1096554, EPI_ISL_1096555, EPI_ISL_1096556, EPI_ISL_1096557, EPI_ISL_1096558, EPI_ISL_1096559, EPI_ISL_1096560, EPI_ISL_1096561, EPI_ISL_1096562, EPI_ISL_1096563, EPI_ISL_1096564, EPI_ISL_1096565, EPI_ISL_1096566, EPI_ISL_1096567, EPI_ISL_1096568, EPI_ISL_1096569, EPI_ISL_1096570, EPI_ISL_1096571, EPI_ISL_1096572, EPI_ISL_1096573, EPI_ISL_1096574, EPI_ISL_1096575, EPI_ISL_1096576, EPI_ISL_1096577, EPI_ISL_1096578, EPI_ISL_1096579, EPI_ISL_1096580, EPI_ISL_1096581, EPI_ISL_1096582, EPI_ISL_1096583, EPI_ISL_1096584, EPI_ISL_1096585, EPI_ISL_1096586, EPI_ISL_1096587, EPI_ISL_1096588, EPI_ISL_1096589, EPI_ISL_1096590, EPI_ISL_1096591, EPI_ISL_1096592, EPI_ISL_1096593, EPI_ISL_1096594, EPI_ISL_1096595, EPI_ISL_1096596, EPI_ISL_1096597, EPI_ISL_1096598, EPI_ISL_1096599, EPI_ISL_1096600, EPI_ISL_1096601, EPI_ISL_1096602, EPI_ISL_1096603, EPI_ISL_1096604, EPI_ISL_1096605, EPI_ISL_1096606, EPI_ISL_1096607, EPI_ISL_1096608, EPI_ISL_1096609, EPI_ISL_1096610, EPI_ISL_1096611, EPI_ISL_1096612, EPI_ISL_1096613, EPI_ISL_1096614, EPI_ISL_1096615, EPI_ISL_1096616, EPI_ISL_1096617, EPI_ISL_1096618, EPI_ISL_1096619, EPI_ISL_1096620, EPI_ISL_1096621, EPI_ISL_1096622, EPI_ISL_1096623, EPI_ISL_1096624, EPI_ISL_1096625, EPI_ISL_1096626, EPI_ISL_1096627, EPI_ISL_1096628, EPI_ISL_1096629, EPI_ISL_1096630, EPI_ISL_1096631, EPI_ISL_1096632, EPI_ISL_1096633, EPI_ISL_1096634, EPI_ISL_1096635, EPI_ISL_1096636, EPI_ISL_1096637, EPI_ISL_1096638, EPI_ISL_1096639, EPI_ISL_1096640, EPI_ISL_1096641, EPI_ISL_1096642, EPI_ISL_1096643, EPI_ISL_1096644, EPI_ISL_1096645, EPI_ISL_1096646, EPI_ISL_1096647, EPI_ISL_1096648, EPI_ISL_1096649, EPI_ISL_1096650, EPI_ISL_1096651, EPI_ISL_1096652, EPI_ISL_1096653, EPI_ISL_1096654, EPI_ISL_1096655, EPI_ISL_1096656, EPI_ISL_1096657, EPI_ISL_1096658, EPI_ISL_1096659, EPI_ISL_1096660, EPI_ISL_1096661, EPI_ISL_1096662, EPI_ISL_1096663, EPI_ISL_1096664, EPI_ISL_1096665, EPI_ISL_1096666, EPI_ISL_1096667, EPI_ISL_1096668, EPI_ISL_1096669, EPI_ISL_1096670, EPI_ISL_1096671, EPI_ISL_1096672, EPI_ISL_1096673, EPI_ISL_1096674, EPI_ISL_1096675, EPI_ISL_1096676, EPI_ISL_1096677, EPI_ISL_1096678, EPI_ISL_1096679, EPI_ISL_1096680, EPI_ISL_1096681, EPI_ISL_1096682, EPI_ISL_1096683, EPI_ISL_1096684, EPI_ISL_1096685, EPI_ISL_1096686, EPI_ISL_1096687, EPI_ISL_1096688, EPI_ISL_1096689, EPI_ISL_1096690, EPI_ISL_1096691, EPI_ISL_1096692, EPI_ISL_1096693, EPI_ISL_1096694, EPI_ISL_1096695, EPI_ISL_1096696, EPI_ISL_1096697, EPI_ISL_1096698, EPI_ISL_1096699, EPI_ISL_1096700, EPI_ISL_1096701, EPI_ISL_1096702, EPI_ISL_1096703, EPI_ISL_1096704, EPI_ISL_1096705, EPI_ISL_1096706, EPI_ISL_1096707, EPI_ISL_1096708, EPI_ISL_1096709, EPI_ISL_1096710, EPI_ISL_1096711, EPI_ISL_1096712, EPI_ISL_1096713, EPI_ISL_1096714, EPI_ISL_1096715, EPI_ISL_1096716, EPI_ISL_1096717, EPI_ISL_1096718, EPI_ISL_1096719, EPI_ISL_1096720, EPI_ISL_1096721, EPI_ISL_1096722, EPI_ISL_1096723, EPI_ISL_1096724, EPI_ISL_1096725, EPI_ISL_1096726, EPI_ISL_1096727, EPI_ISL_1096728, EPI_ISL_1096729, EPI_ISL_1096730, EPI_ISL_1096731, EPI_ISL_1096732, EPI_ISL_1096733, EPI_ISL_1096734, EPI_ISL_1096735, EPI_ISL_1096736, EPI_ISL_1096737, EPI_ISL_1096738, EPI_ISL_1096739, EPI_ISL_1096740, EPI_ISL_1096741, EPI_ISL_1096742, EPI_ISL_1096743, EPI_ISL_1096744, EPI_ISL_1096745, EPI_ISL_1096746, EPI_ISL_1096747, EPI_ISL_1096748, EPI_ISL_1096749, EPI_ISL_1096750, EPI_ISL_1096751, EPI_ISL_1096752, EPI_ISL_1096753, EPI_ISL_1096754, EPI_ISL_1096755, EPI_ISL_1096756, EPI_ISL_1096757, EPI_ISL_1096758, EPI_ISL_1096759, EPI_ISL_1096760, EPI_ISL_1096761, EPI_ISL_1096762, EPI_ISL_1096763, EPI_ISL_1096764, EPI_ISL_1096765, EPI_ISL_1096766, EPI_ISL_1096767, EPI_ISL_1096768, EPI_ISL_1096769, EPI_ISL_1096770, EPI_ISL_1096771, EPI_ISL_1096772, EPI_ISL_1096773, EPI_ISL_1096774, EPI_ISL_1096775, EPI_ISL_1096776, EPI_ISL_1096777, EPI_ISL_1096778, EPI_ISL_1096779, EPI_ISL_1096780, EPI_ISL_1096781, EPI_ISL_1096782, EPI_ISL_1096783, EPI_ISL_1096784, EPI_ISL_1096785, EPI_ISL_1096786, EPI_ISL_1096787, EPI_ISL_1096788, EPI_ISL_1096789, EPI_ISL_1096790, EPI_ISL_1096791, EPI_ISL_1096792, EPI_ISL_1096793, EPI_ISL_1096794, EPI_ISL_1096795, EPI_ISL_1096796, EPI_ISL_1096797, EPI_ISL_1096798, EPI_ISL_1096799, EPI_ISL_1096800, EPI_ISL_1096801, EPI_ISL_1096802, EPI_ISL_1096803, EPI_ISL_1096804, EPI_ISL_1096805, EPI_ISL_1096806, EPI_ISL_1096807, EPI_ISL_1096808, EPI_ISL_1096809, EPI_ISL_1096810, EPI_ISL_1096811, EPI_ISL_1096812, EPI_ISL_1096813, EPI_ISL_1096814, EPI_ISL_1096815, EPI_ISL_1096816, EPI_ISL_1096817, EPI_ISL_1096818, EPI_ISL_1096819, EPI_ISL_1096820, EPI_ISL_1096821, EPI_ISL_1096822, EPI_ISL_1096823, EPI_ISL_1096824, EPI_ISL_1096825, EPI_ISL_1096826, EPI_ISL_1096827, EPI_ISL_1096828, EPI_ISL_1096829, EPI_ISL_1096830, EPI_ISL_1096831, EPI_ISL_1096832, EPI_ISL_1096833, EPI_ISL_1096834, EPI_ISL_1096835, EPI_ISL_1096836, EPI_ISL_1096837, EPI_ISL_1096838, EPI_ISL_1096839, EPI_ISL_1096840, EPI_ISL_1096841, EPI_ISL_1096842, EPI_ISL_1096843, EPI_ISL_1096844, EPI_ISL_1096845, EPI_ISL_1096846, EPI_ISL_1096847, EPI_ISL_1096848, EPI_ISL_1096849, EPI_ISL_1096850, EPI_ISL_1096851, EPI_ISL_1096852, EPI_ISL_1096853, EPI_ISL_1096854, EPI_ISL_1096855, EPI_ISL_1096856, EPI_ISL_1096857, EPI_ISL_1096858, EPI_ISL_1096859, EPI_ISL_1096860, EPI_ISL_1096861, EPI_ISL_1096862, EPI_ISL_1096863, EPI_ISL_1096864, EPI_ISL_1096865, EPI_ISL_1096866, EPI_ISL_1096867, EPI_ISL_1096868, EPI_ISL_1096869, EPI_ISL_1096870, EPI_ISL_1096871, EPI_ISL_1096872, EPI_ISL_1096873, EPI_ISL_1096874, EPI_ISL_1096875, EPI_ISL_1096876, EPI_ISL_1096877, EPI_ISL_1096878, EPI_ISL_1096879, EPI_ISL_1096880, EPI_ISL_1096881, EPI_ISL_1096882, EPI_ISL_1096883, EPI_ISL_1096884, EPI_ISL_1096885, EPI_ISL_1096886, EPI_ISL_1096887, EPI_ISL_1096888, EPI_ISL_1096889, EPI_ISL_1096890, EPI_ISL_1096891, EPI_ISL_1096892, EPI_ISL_1096893, EPI_ISL_1096894, EPI_ISL_1096895, EPI_ISL_1096896, EPI_ISL_1096897, EPI_ISL_1096898, EPI_ISL_1096899, EPI_ISL_1096900, EPI_ISL_1096901, EPI_ISL_1096902, EPI_ISL_1096903, EPI_ISL_1096904, EPI_ISL_1096905, EPI_ISL_1096906, EPI_ISL_1096907, EPI_ISL_1096908, EPI_ISL_1096909, EPI_ISL_1096910, EPI_ISL_1096911, EPI_ISL_1096912, EPI_ISL_1096913, EPI_ISL_1096914, EPI_ISL_1096915, EPI_ISL_1096916, EPI_ISL_1096917, EPI_ISL_1096918, EPI_ISL_1096919, EPI_ISL_1096920, EPI_ISL_1096921, EPI_ISL_1096922, EPI_ISL_1096923, EPI_ISL_1096924, EPI_ISL_1096925, EPI_ISL_1096926, EPI_ISL_1096927, EPI_ISL_1096928, EPI_ISL_1096929, EPI_ISL_1096930, EPI_ISL_1096931, EPI_ISL_1096932, EPI_ISL_1096933, EPI_ISL_1096934, EPI_ISL_1096935, EPI_ISL_1096936, EPI_ISL_1096937, EPI_ISL_1096938, EPI_ISL_1096939, EPI_ISL_1096940, EPI_ISL_1096941, EPI_ISL_1096942, EPI_ISL_1096943, EPI_ISL_1096944, EPI_ISL_1096945, EPI_ISL_1096946, EPI_ISL_1096947, EPI_ISL_1096948, EPI_ISL_1096949, EPI_ISL_1096950, EPI_ISL_1096951, EPI_ISL_1096952, EPI_ISL_1096953, EPI_ISL_1096954, EPI_ISL_1096955, EPI_ISL_1096956, EPI_ISL_1096957, EPI_ISL_1096958, EPI_ISL_1096959, EPI_ISL_1096960, EPI_ISL_1096961, EPI_ISL_1096962, EPI_ISL_1096963, EPI_ISL_1096964, EPI_ISL_1096965, EPI_ISL_1096966, EPI_ISL_1096967, EPI_ISL_1096968, EPI_ISL_1096969, EPI_ISL_1096970, EPI_ISL_1096971, EPI_ISL_1096972, EPI_ISL_1096973, EPI_ISL_1096974, EPI_ISL_1096975, EPI_ISL_1096976, EPI_ISL_1096977, EPI_ISL_1096978, EPI_ISL_1096979, EPI_ISL_1096980, EPI_ISL_1096981 | | | |
| see above | ASL Napoli 1 Centro | AMES Centro Polidiagnostico Strumentale S.r.l. | "Giovanni Savarese, Raffaella Ruggiero, Eloisa Evangelista, Antonella Di Carlo, Luisa Circelli, Luigi D'Amore, Roberto Sirica, Antonio Fico" |
| EPI_ISL_1096982, EPI_ISL_1096984, EPI_ISL_1096986, EPI_ISL_1096989, EPI_ISL_1096991, EPI_ISL_1096993, EPI_ISL_1096994, EPI_ISL_1096996, EPI_ISL_1096998, EPI_ISL_1097000 | Laboratorio Microbiologia e Virologia P.O. Cotugno A.O. dei Colli | Laboratorio Microbiologia e Virologia P.O. Cotugno A.O. dei Colli | Luigi Atripaldi, Claudia Tiberio, Anna Perfetti, Pellegrino Cerino, Biancamaria Pierri, Maria Concetta Cuomo |
| EPI_ISL_1097002 | Center of Advanced Studies and Technology, Molecular Genetics Laboratory | Center of Advanced Studies and Technology, Molecular Genetics Laboratory | Ferrante Rossella, Mandatori Domitilla, De Fabritiis Simone, Damiani Verena, Anaclerio Federico |
| EPI_ISL_1097004, EPI_ISL_1097005, EPI_ISL_1097007, EPI_ISL_1097008, EPI_ISL_1097009, EPI_ISL_1097010, EPI_ISL_1097011, EPI_ISL_1097012, EPI_ISL_1097013, EPI_ISL_1097014, EPI_ISL_1097015 | | | |
| see above | Laboratorio Microbiologia e Virologia P.O. Cotugno A.O. dei Colli | Laboratorio Microbiologia e Virologia P.O. Cotugno A.O. dei Colli | Luigi Atripaldi, Claudia Tiberio, Anna Perfetti, Pellegrino Cerino, Biancamaria Pierri, Maria Concetta Cuomo |
| EPI_ISL_1097016 | Laboratorio Microbiologia e Virologia P.O. Cotugno A.O. dei Col | Laboratorio Microbiologia e Virologia P.O. Cotugno A.O. dei Colli | Luigi Atripaldi, Claudia Tiberio, Anna Perfetti, Pellegrino Cerino, Biancamaria Pierri, Maria Concetta Cuomo |

| | | | |
|---|---|---|---|
| EPI_ISL_1097017, EPI_ISL_1097019, EPI_ISL_1097020, EPI_ISL_1097021, EPI_ISL_1097022 | Laboratorio Microbiologia e Virologia P.O. Cotugno A.O. dei Colli | Laboratorio Microbiologia e Virologia P.O. Cotugno A.O. dei Colli | Luigi Atripaldi, Claudia Tiberio, Anna Perfetti, Pellegrino Cerino, Biancamaria Pierri, Maria Concetta Cuomo |
| EPI_ISL_1104645, EPI_ISL_1104647, EPI_ISL_1104649 | Virologia, Dipartimento di Scienze Biomediche, Università di Sassari, Viale San Pietro, 43/B - Sassari | Laboratorio specialistico UOC Ematologia - Ospedale "San Francesco" - ATS-ASSL Nuoro | Giovanna Piras, Asproni Rosanna, Paolo Malune, Maria Itria Monne, Angelo Domenico Palmas, Caterina Serra, Elena Rimini, Salvatore Rubino |
| EPI_ISL_1104650 | Laboratorio Biologia Molecolare Sars Cov2 - UOC Laboratorio Analisi - Servizio Medicina di Laboratorio, Ospedale "San Francesco" - ATS-ASSL Nuoro | Laboratorio specialistico UOC Ematologia - Ospedale "San Francesco" - ATS-ASSL Nuoro | Giovanna Piras, Rosanna Asproni, Paolo Malune, Maura Fiamma, Maria Itria Monne, Angelo Domenico Palmas, Iana Lo Maglio, Giuseppe Mameli |
| EPI_ISL_1104652, EPI_ISL_1104654, EPI_ISL_1104656 | Virologia, Dipartimento di Scienze Biomediche, Università di Sassari, Viale San Pietro, 43/B - Sassari | Laboratorio specialistico UOC Ematologia - Ospedale "San Francesco" - ATS-ASSL Nuoro | Giovanna Piras, Asproni Rosanna, Paolo Malune, Maria Itria Monne, Angelo Domenico Palmas, Caterina Serra, Elena Rimini, Salvatore Rubino |
| EPI_ISL_1109574 | SC (UCO) Igiene e Sanità Pubblica (funzione integrata con SC Microbiologia e Virologia) | ARGO Laboratorio Genomica ed Epigenomica | Licastro D, Dal Monego S, Degasperi M, Marcello A, D'Agaro P, Lombardo F |
| EPI_ISL_1109575 | SC (UCO) Igiene e Sanità Pubblica (funzione integrata con SC Microbiologia e Virologia) e Azienda Ospedaliera Pordenone | ARGO Laboratorio Genomica ed Epigenomica | Licastro D, Dal Monego S, Degasperi M, Marcello A, D'Agaro P, De Rosa R |
| EPI_ISL_1109576 | SC (UCO) Igiene e Sanità Pubblica (funzione integrata con SC Microbiologia e Virologia) | ARGO Laboratorio Genomica ed Epigenomica | Licastro D, Dal Monego S, Degasperi M, Marcello A, Segat L, Piscianz E, D'Agaro P |
| EPI_ISL_1109577 | SC (UCO) Igiene e Sanità Pubblica (funzione integrata con SC Microbiologia e Virologia) | ARGO Laboratorio Genomica ed Epigenomica | Licastro D, Dal Monego S, Degasperi M, Marcello A, D'Agaro P, Lombardo F |
| EPI_ISL_1109578, EPI_ISL_1109579, EPI_ISL_1109580, EPI_ISL_1109581 | SC (UCO) Igiene e Sanità Pubblica (funzione integrata con SC Microbiologia e Virologia) e Az. Ospedaliero-Universitaria Udine | ARGO Laboratorio Genomica ed Epigenomica | Licastro D, Dal Monego S, Degasperi M, Marcello A, D'Agaro P, Pipan C |
| EPI_ISL_1109582 | SC (UCO) Igiene e Sanità Pubblica (funzione integrata con SC Microbiologia e Virologia) | ARGO Laboratorio Genomica ed Epigenomica | Licastro D, Dal Monego S, Degasperi M, Marcello A, D'Agaro P, Lombardo F |
| EPI_ISL_1109583, EPI_ISL_1109584, EPI_ISL_1109585, EPI_ISL_1109586 | SC (UCO) Igiene e Sanità Pubblica (funzione integrata con SC Microbiologia e Virologia) e Az. Ospedaliero-Universitaria Udine | ARGO Laboratorio Genomica ed Epigenomica | Licastro D, Dal Monego S, Degasperi M, Marcello A, D'Agaro P, Pipan C |
| EPI_ISL_1109587 | SC (UCO) Igiene e Sanità Pubblica (funzione integrata con SC Microbiologia e Virologia) e Azienda Ospedaliera Pordenone | ARGO Laboratorio Genomica ed Epigenomica | Licastro D, Dal Monego S, Degasperi M, Marcello A, D'Agaro P, De Rosa R |
| EPI_ISL_1109588 | SC (UCO) Igiene e Sanità Pubblica (funzione integrata con SC Microbiologia e Virologia) | ARGO Laboratorio Genomica ed Epigenomica | Licastro D, Dal Monego S, Degasperi M, Marcello A, D'Agaro P, Lombardo F |
| EPI_ISL_1109589, EPI_ISL_1109590 | SC (UCO) Igiene e Sanità Pubblica (funzione integrata con SC Microbiologia e Virologia) e Az. Ospedaliero-Universitaria Udine | ARGO Laboratorio Genomica ed Epigenomica | Licastro D, Dal Monego S, Degasperi M, Marcello A, D'Agaro P, Pipan C |
| EPI_ISL_1109591 | SC (UCO) Igiene e Sanità Pubblica (funzione integrata con SC Microbiologia e Virologia) | ARGO Laboratorio Genomica ed Epigenomica | Licastro D, Dal Monego S, Degasperi M, Marcello A, D'Agaro P, Lombardo F |
| EPI_ISL_1109592, EPI_ISL_1109593 | SC (UCO) Igiene e Sanità Pubblica (funzione integrata con SC Microbiologia e Virologia) | ARGO Laboratorio Genomica ed Epigenomica | Licastro D, Dal Monego S, Degasperi M, Marcello A, Segat L, Piscianz E, D'Agaro P |
| EPI_ISL_1109594 | SC (UCO) Igiene e Sanità Pubblica (funzione integrata con SC Microbiologia e Virologia) e Azienda Ospedaliera Pordenone | ARGO Laboratorio Genomica ed Epigenomica | Licastro D, Dal Monego S, Degasperi M, Marcello A, D'Agaro P, De Rosa R |
| EPI_ISL_1109595, EPI_ISL_1109596, EPI_ISL_1109597, EPI_ISL_1109598, EPI_ISL_1109599 | SC (UCO) Igiene e Sanità Pubblica (funzione integrata con SC Microbiologia e Virologia) e Az. Ospedaliero-Universitaria Udine | ARGO Laboratorio Genomica ed Epigenomica | Licastro D, Dal Monego S, Degasperi M, Marcello A, D'Agaro P, Pipan C |
| EPI_ISL_1109600 | SC (UCO) Igiene e Sanità Pubblica (funzione integrata con SC Microbiologia e Virologia) | ARGO Laboratorio Genomica ed Epigenomica | Licastro D, Dal Monego S, Degasperi M, Marcello A, D'Agaro P, Lombardo F |
| EPI_ISL_1109601, EPI_ISL_1109602, EPI_ISL_1109603, EPI_ISL_1109604, EPI_ISL_1109605, EPI_ISL_1109606, EPI_ISL_1109607 | SC (UCO) Igiene e Sanità Pubblica (funzione integrata con SC Microbiologia e Virologia) e Az. Ospedaliero-Universitaria Udine | ARGO Laboratorio Genomica ed Epigenomica | Licastro D, Dal Monego S, Degasperi M, Marcello A, D'Agaro P, Pipan C |
| EPI_ISL_1109608, EPI_ISL_1109609 | SC (UCO) Igiene e Sanità Pubblica (funzione integrata con SC Microbiologia e Virologia) | ARGO Laboratorio Genomica ed Epigenomica | Licastro D, Dal Monego S, Degasperi M, Marcello A, Segat L, Piscianz E, D'Agaro P |
| EPI_ISL_1109610, EPI_ISL_1109611, EPI_ISL_1109612 | SC (UCO) Igiene e Sanità Pubblica (funzione integrata con SC Microbiologia e Virologia) e Azienda Ospedaliera Pordenone | ARGO Laboratorio Genomica ed Epigenomica | Licastro D, Dal Monego S, Degasperi M, Marcello A, D'Agaro P, De Rosa R |
| EPI_ISL_1109613 | SC (UCO) Igiene e Sanità Pubblica (funzione integrata con SC Microbiologia e Virologia) e Az. Ospedaliero-Universitaria Udine | ARGO Laboratorio Genomica ed Epigenomica | Licastro D, Dal Monego S, Degasperi M, Marcello A, D'Agaro P, Pipan C |
| EPI_ISL_1109614, EPI_ISL_1109615 | SC (UCO) Igiene e Sanità Pubblica (funzione integrata con SC Microbiologia e Virologia) e Azienda Ospedaliera Pordenone | ARGO Laboratorio Genomica ed Epigenomica | Licastro D, Dal Monego S, Degasperi M, Marcello A, D'Agaro P, De Rosa R |
| EPI_ISL_1109616, EPI_ISL_1109617, EPI_ISL_1109618, EPI_ISL_1109619 | SC (UCO) Igiene e Sanità Pubblica (funzione integrata con SC Microbiologia e Virologia) e Az. Ospedaliero-Universitaria Udine | ARGO Laboratorio Genomica ed Epigenomica | Licastro D, Dal Monego S, Degasperi M, Marcello A, D'Agaro P, Pipan C |
| EPI_ISL_1109620 | SC (UCO) Igiene e Sanità Pubblica (funzione integrata con SC Microbiologia e Virologia) | ARGO Laboratorio Genomica ed Epigenomica | Licastro D, Dal Monego S, Degasperi M, Marcello A, Segat L, Piscianz E, D'Agaro P |
| EPI_ISL_1109621 | SC (UCO) Igiene e Sanità Pubblica (funzione integrata con SC Microbiologia e Virologia) e Az. Ospedaliero-Universitaria Udine | ARGO Laboratorio Genomica ed Epigenomica | Licastro D, Dal Monego S, Degasperi M, Marcello A, D'Agaro P, Pipan C |
| EPI_ISL_1110295, EPI_ISL_1110296, EPI_ISL_1110297, EPI_ISL_1110298, EPI_ISL_1110299, EPI_ISL_1110300, EPI_ISL_1110301, EPI_ISL_1110302, EPI_ISL_1110303, EPI_ISL_1110304, EPI_ISL_1110305, EPI_ISL_1110306, EPI_ISL_1110307, EPI_ISL_1110308, EPI_ISL_1110309, EPI_ISL_1110310, EPI_ISL_1110311, EPI_ISL_1110312, EPI_ISL_1110313, EPI_ISL_1110314, EPI_ISL_1110315, EPI_ISL_1110316, EPI_ISL_1110317, EPI_ISL_1110318, EPI_ISL_1110319, EPI_ISL_1110320, EPI_ISL_1110321, EPI_ISL_1110322, EPI_ISL_1110323, EPI_ISL_1110324, EPI_ISL_1110325, EPI_ISL_1110326, EPI_ISL_1110327, EPI_ISL_1110328, EPI_ISL_1110329, EPI_ISL_1110330, EPI_ISL_1110331, EPI_ISL_1110332, EPI_ISL_1110333, EPI_ISL_1110334, EPI_ISL_1110335, EPI_ISL_1110336, EPI_ISL_1110337, EPI_ISL_1110338, EPI_ISL_1110339, EPI_ISL_1110340, EPI_ISL_1110341, EPI_ISL_1110342, EPI_ISL_1110343, EPI_ISL_1110344, EPI_ISL_1110345, EPI_ISL_1110346, EPI_ISL_1110347, EPI_ISL_1110348, EPI_ISL_1110349, EPI_ISL_1110350, EPI_ISL_1110351, EPI_ISL_1110352, EPI_ISL_1110353, EPI_ISL_1110354, EPI_ISL_1110355, EPI_ISL_1110356, EPI_ISL_1110357, EPI_ISL_1110358, EPI_ISL_1110359, EPI_ISL_1110360, EPI_ISL_1110361, EPI_ISL_1110362, EPI_ISL_1110363, EPI_ISL_1110364, EPI_ISL_1110365, EPI_ISL_1110366, EPI_ISL_1110367, EPI_ISL_1110368, EPI_ISL_1110369, EPI_ISL_1110370, EPI_ISL_1110371, EPI_ISL_1110372, EPI_ISL_1110373, EPI_ISL_1110374, EPI_ISL_1110375, EPI_ISL_1110376, EPI_ISL_1110377, EPI_ISL_1110378, EPI_ISL_1110379, EPI_ISL_1110380, EPI_ISL_1110381, EPI_ISL_1110382, EPI_ISL_1110383, EPI_ISL_1110384, EPI_ISL_1110385, EPI_ISL_1110386, EPI_ISL_1110387, EPI_ISL_1110388 | | | |
| see above | Azienda Sanitaria dell'Alto Adige Laboratorio Aziendale di Microbiologia e Virologia | Istituto di Genomica Applicata | Elisabetta Pagani, Irene Bianconi, Elisabetta Giacobazzi, Elisa Masi, Stefanie Wieser, Irena Jurman, Vera Vendramin, Gabriele Magris, Eleonora Paparelli, Davide Scaglione, Michele Morgante |

| | | | |
|---|---|---|---|
| EPI_ISL_1110581, EPI_ISL_1110582, EPI_ISL_1110583, EPI_ISL_1110584, EPI_ISL_1110585, EPI_ISL_1110586, EPI_ISL_1110587, EPI_ISL_1110588, EPI_ISL_1110589, EPI_ISL_1110590, EPI_ISL_1110591, EPI_ISL_1110592, EPI_ISL_1110593, EPI_ISL_1110594, EPI_ISL_1110595 | | | |
| see above | LAB ANALISI PO CARDARELLI CB | Istituto Zooprofilattico Sperimentale dell'Abruzzo e Molise "G. Caporale" | Scutellà M, Niro G, Lorusso A, Marcacci M, Di Domenico M, Ancora M, Curini V, Mangone I, Rinaldi A, Scialabba S, Di Pasquale A, Cammà C, Puglia I, Calistri P, Savini G |
| EPI_ISL_1110596 | SIESP L'Aquila | Istituto Zooprofilattico Sperimentale dell'Abruzzo e Molise "G. Caporale" | Lorusso A, Marcacci M, Di Domenico M, Ancora M, Curini V, Mangone I, Rinaldi A, Scialabba S, Di Pasquale A, Cammà C, Puglia I, Calistri P, Savini G |
| EPI_ISL_1110597 | SIESP CHIETI-Drive in Chieti | Istituto Zooprofilattico Sperimentale dell'Abruzzo e Molise "G. Caporale" | Lorusso A, Marcacci M, Di Domenico M, Ancora M, Curini V, Mangone I, Rinaldi A, Scialabba S, Di Pasquale A, Cammà C, Puglia I, Calistri P, Savini G |
| EPI_ISL_1110598, EPI_ISL_1110599, EPI_ISL_1110600 | SIESP CH-Drive in VASTO | Istituto Zooprofilattico Sperimentale dell'Abruzzo e Molise "G. Caporale" | Lorusso A, Marcacci M, Di Domenico M, Ancora M, Curini V, Mangone I, Rinaldi A, Scialabba S, Di Pasquale A, Cammà C, Puglia I, Calistri P, Savini G |
| EPI_ISL_1110601 | Ospedale Civile Teramo | Istituto Zooprofilattico Sperimentale dell'Abruzzo e Molise "G. Caporale" | Lorusso A, Marcacci M, Di Domenico M, Ancora M, Curini V, Mangone I, Rinaldi A, Scialabba S, Di Pasquale A, Cammà C, Puglia I, Calistri P, Savini G |
| EPI_ISL_1110602 | SIESP Dip. Prev Teramo | Istituto Zooprofilattico Sperimentale dell'Abruzzo e Molise "G. Caporale" | Lorusso A, Marcacci M, Di Domenico M, Ancora M, Curini V, Mangone I, Rinaldi A, Scialabba S, Di Pasquale A, Cammà C, Puglia I, Calistri P, Savini G |
| EPI_ISL_1112221, EPI_ISL_1112222 | Ospedale Santissima Annunziata | Istituto Zooprofilattico Sperimentale della Puglia e della Basilicata | Parisi A., Bianco A., Capozzi L., Del Sambro L., Simone D., Giannico A., Ridolfi D. |
| EPI_ISL_1112223, EPI_ISL_1112224, EPI_ISL_1112225, EPI_ISL_1112226 | Ospedale Di Venere - Carbonara | Istituto Zooprofilattico Sperimentale della Puglia e della Basilicata | Parisi A., Bianco A., Capozzi L., Del Sambro L., Simone D., Giannico A., Ridolfi D. |
| EPI_ISL_1112227, EPI_ISL_1112228, EPI_ISL_1112229, EPI_ISL_1112230, EPI_ISL_1112231, EPI_ISL_1112232, EPI_ISL_1112233, EPI_ISL_1112234 | Istituto Zooprofilattico Sperimentale della Puglia e della Basilicata | Istituto Zooprofilattico Sperimentale della Puglia e della Basilicata | Parisi A., Bianco A., Capozzi L., Del Sambro L., Simone D., Giannico A., Ridolfi D. |
| EPI_ISL_1112235 | Ospedali Riuniti Azienda Ospedaliera Universitaria - Foggia | Istituto Zooprofilattico Sperimentale della Puglia e della Basilicata | Parisi A., Bianco A., Capozzi L., Del Sambro L., Simone D., Giannico A., Ridolfi D. |
| EPI_ISL_1112236 | Dipartimento di Scienze Biomediche e Oncologia Umana - Azienda Ospedaliero Universitaria Consorziale Policlinico | Istituto Zooprofilattico Sperimentale della Puglia e della Basilicata | Parisi A., Bianco A., Capozzi L., Del Sambro L., Simone D., Giannico A., Ridolfi D. |
| EPI_ISL_1112237, EPI_ISL_1112238, EPI_ISL_1112239, EPI_ISL_1112240, EPI_ISL_1112241, EPI_ISL_1112242, EPI_ISL_1112243, EPI_ISL_1112244, EPI_ISL_1112245, EPI_ISL_1112246, EPI_ISL_1112247, EPI_ISL_1112248, EPI_ISL_1112249, EPI_ISL_1112250, EPI_ISL_1112251, EPI_ISL_1112252, EPI_ISL_1112253, EPI_ISL_1112254, EPI_ISL_1112255, EPI_ISL_1112256, EPI_ISL_1112257, EPI_ISL_1112258, EPI_ISL_1112259, EPI_ISL_1112260, EPI_ISL_1112261, EPI_ISL_1112262, EPI_ISL_1112263, EPI_ISL_1112264, EPI_ISL_1112265, EPI_ISL_1112266, EPI_ISL_1112267, EPI_ISL_1112268, EPI_ISL_1112269, EPI_ISL_1112270, EPI_ISL_1112271, EPI_ISL_1112272, EPI_ISL_1112273, EPI_ISL_1112274, EPI_ISL_1112275, EPI_ISL_1112276, EPI_ISL_1112277, EPI_ISL_1112278, EPI_ISL_1112279, EPI_ISL_1112280, EPI_ISL_1112281, EPI_ISL_1112282, EPI_ISL_1112283, EPI_ISL_1112284, EPI_ISL_1112285, EPI_ISL_1112286, EPI_ISL_1112287, EPI_ISL_1112288, EPI_ISL_1112289, EPI_ISL_1112290, EPI_ISL_1112291 | | | |
| see above | Dipartimento di Scienze Biomediche e Oncologia Umana - Azienda Ospedaliero Universitaria Consorziale Policlinico | Istituto Zooprofilattico Sperimentale della Puglia e della Basilicata | Parisi A., Bianco A., Capozzi L., Del Sambro L., Simone D., Chironna M., Loconsole D., Sallustio A. |
| EPI_ISL_1114766 | SIESP L'Aquila | Istituto Zooprofilattico Sperimentale dell'Abruzzo e Molise "G. Caporale" | Lorusso A, Marcacci M, Di Domenico M, Ancora M, Curini V, Mangone I, Rinaldi A, Scialabba S, Di Pasquale A, Cammà C, Puglia I, Calistri P, Savini G |
| EPI_ISL_1114769 | Virologia, Dipartimento di Scienze Biomediche, Università di Sassari, Viale San Pietro, 43/B - Sassari | Laboratorio specialistico UOC Ematologia - Ospedale "San Francesco" - ATS-ASSL Nuoro | Giovanna Piras, Rosanna Asproni, Paolo Malune, Maria Itria Monne, Angelo Domenico Palmas, Caterina Serra, Elena Rimini, Salvatore Rubino |
| EPI_ISL_1116471, EPI_ISL_1116472 | Dipartimento di Scienze Biomediche e Oncologia Umana - Azienda Ospedaliero Universitaria Consorziale Policlinico | Istituto Zooprofilattico Sperimentale della Puglia e della Basilicata | Parisi A., Bianco A., Capozzi L., Del Sambro L., Simone D., Chironna M., Loconsole D., Sallustio A. |
| EPI_ISL_1116473 | Istituto Zooprofilattico Sperimentale della Puglia e della Basilicata | Istituto Zooprofilattico Sperimentale della Puglia e della Basilicata | Parisi A., Bianco A., Capozzi L., Del Sambro L., Simone D., Chironna M., Loconsole D., Sallustio A., Ridolfi D. |
| EPI_ISL_1116474 | Istituto Zooprofilattico Sperimentale della Puglia e della Basilicata | Istituto Zooprofilattico Sperimentale della Puglia e della Basilicata | Parisi A., Bianco A., Capozzi L., Del Sambro L., Simone D., Chironna M., Loconsole D., Sallustio A., Giannico A. |
| EPI_ISL_1116475 | Dipartimento di Scienze Biomediche e Oncologia Umana - Azienda Ospedaliero Universitaria Consorziale Policlinico | Istituto Zooprofilattico Sperimentale della Puglia e della Basilicata | Parisi A., Bianco A., Capozzi L., Del Sambro L., Simone D., Chironna M., Loconsole D., Sallustio A. |
| EPI_ISL_1117458 | OSP SAN SALVATORE, MEDICINA INTERNA | Istituto Zooprofilattico Sperimentale dell'Abruzzo e Molise "G. Caporale" | Lorusso A, Marcacci M, Di Domenico M, Ancora M, Curini V, Mangone I, Rinaldi A, Scialabba S, Di Pasquale A, Cammà C, Scialabba S, Puglia I, Calistri P, Savini G |
| EPI_ISL_1117459 | SIESP L'AQUILA, | Istituto Zooprofilattico Sperimentale dell'Abruzzo e Molise "G. Caporale" | Lorusso A, Marcacci M, Di Domenico M, Ancora M, Curini V, Mangone I, Rinaldi A, Scialabba S, Di Pasquale A, Cammà C, Scialabba S, Puglia I, Calistri P, Savini G |
| EPI_ISL_1117460 | SIESP DIP PREV CHIETI | Istituto Zooprofilattico Sperimentale dell'Abruzzo e Molise "G. Caporale" | Lorusso A, Marcacci M, Di Domenico M, Ancora M, Curini V, Mangone I, Rinaldi A, Scialabba S, Di Pasquale A, Cammà C, Scialabba S, Puglia I, Calistri P, Savini G |
| EPI_ISL_1117461 | SIESP CHIETI,DRIVE IN CHIETI | Istituto Zooprofilattico Sperimentale dell'Abruzzo e Molise "G. Caporale" | Lorusso A, Marcacci M, Di Domenico M, Ancora M, Curini V, Mangone I, Rinaldi A, Scialabba S, Di Pasquale A, Cammà C, Scialabba S, Puglia I, Calistri P, Savini G |
| EPI_ISL_1117462 | SIESP CHIETI, DRIVE IN CHIETI | Istituto Zooprofilattico Sperimentale dell'Abruzzo e Molise "G. Caporale" | Lorusso A, Marcacci M, Di Domenico M, Ancora M, Curini V, Mangone I, Rinaldi A, Scialabba S, Di Pasquale A, Cammà C, Scialabba S, Puglia I, Calistri P, Savini G |
| EPI_ISL_1117463 | SIESP CHIETI, DRIVE IN LANCIANO | Istituto Zooprofilattico Sperimentale dell'Abruzzo e Molise "G. Caporale" | Lorusso A, Marcacci M, Di Domenico M, Ancora M, Curini V, Mangone I, Rinaldi A, Scialabba S, Di Pasquale A, Cammà C, Scialabba S, Puglia I, Calistri P, Savini G |
| EPI_ISL_1117464 | SIESP CHIETI, DRIVE IN CHIETI | Istituto Zooprofilattico Sperimentale dell'Abruzzo e Molise "G. Caporale" | Lorusso A, Marcacci M, Di Domenico M, Ancora M, Curini V, Mangone I, Rinaldi A, Scialabba S, Di Pasquale A, Cammà C, Scialabba S, Puglia I, Calistri P, Savini G |
| EPI_ISL_1117465 | SIESP DIP PREV CHIETI | Istituto Zooprofilattico Sperimentale dell'Abruzzo e Molise "G. Caporale" | Lorusso A, Marcacci M, Di Domenico M, Ancora M, Curini V, Mangone I, Rinaldi A, Scialabba S, Di Pasquale A, Cammà C, Scialabba S, Puglia I, Calistri P, Savini G |
| EPI_ISL_1117466, EPI_ISL_1117467 | SIESP DIP PREV TERAMO | Istituto Zooprofilattico Sperimentale dell'Abruzzo e Molise "G. Caporale" | Lorusso A, Marcacci M, Di Domenico M, Ancora M, Curini V, Mangone I, Rinaldi A, Scialabba S, Di Pasquale A, Cammà C, Scialabba S, Puglia I, Calistri P, Savini G |
| EPI_ISL_1117468 | OSP SAN SALVATORE, MEDICINA INTERNA | Istituto Zooprofilattico Sperimentale dell'Abruzzo e Molise "G. Caporale" | Lorusso A, Marcacci M, Di Domenico M, Ancora M, Curini V, Mangone I, Rinaldi A, Scialabba S, Di Pasquale A, Cammà C, Scialabba S, Puglia I, Calistri P, Savini G |
| EPI_ISL_1117469, EPI_ISL_1117470, EPI_ISL_1117471, EPI_ISL_1117472 | SIESP DIP PREV TERAMO | Istituto Zooprofilattico Sperimentale dell'Abruzzo e Molise "G. Caporale" | Lorusso A, Marcacci M, Di Domenico M, Ancora M, Curini V, Mangone I, Rinaldi A, Scialabba S, Di Pasquale A, Cammà C, Scialabba S, Puglia I, Calistri P, Savini G |
| EPI_ISL_1117473 | OSP SAN SALVATORE, MEDICINA INTERNA | Istituto Zooprofilattico Sperimentale dell'Abruzzo e Molise "G. Caporale" | Lorusso A, Marcacci M, Di Domenico M, Ancora M, Curini V, Mangone I, Rinaldi A, Scialabba S, Di Pasquale A, Cammà C, Scialabba S, Puglia I, Calistri P, Savini G |
| EPI_ISL_1117474 | SIESP DIP PREV CHIETI | Istituto Zooprofilattico Sperimentale dell'Abruzzo e Molise "G. Caporale" | Lorusso A, Marcacci M, Di Domenico M, Ancora M, Curini V, Mangone I, Rinaldi A, Scialabba S, Di Pasquale A, Cammà C, Scialabba S, Puglia I, Calistri P, Savini G |
| EPI_ISL_1117475 | SIESP SULMONA | Istituto Zooprofilattico Sperimentale dell'Abruzzo e Molise "G. Caporale" | Lorusso A, Marcacci M, Di Domenico M, Ancora M, Curini V, Mangone I, Rinaldi A, Scialabba S, Di Pasquale A, Cammà C, Scialabba S, Puglia I, Calistri P, Savini G |

| | | | |
|---|---|---|---|
| EPI_ISL_1117476, EPI_ISL_1117477 | SIESP CHIETI, DRIVE IN CHIETI | Istituto Zooprofilattico Sperimentale dell'Abruzzo e Molise "G. Caporale" | Lorusso A, Marcacci M, Di Domenico M, Ancora M, Curini V, Mangone I, Rinaldi A, Scialabba S, Di Pasquale A, Cammà C, Scialabba S, Puglia I, Calistri P, Savini G |
| EPI_ISL_1117478 | SIESP CHIETI, DRIVE IN ORTONA | Istituto Zooprofilattico Sperimentale dell'Abruzzo e Molise "G. Caporale" | Lorusso A, Marcacci M, Di Domenico M, Ancora M, Curini V, Mangone I, Rinaldi A, Scialabba S, Di Pasquale A, Cammà C, Scialabba S, Puglia I, Calistri P, Savini G |
| EPI_ISL_1117479 | SIESP CHIETI, DRIVE IN LANCIANO | Istituto Zooprofilattico Sperimentale dell'Abruzzo e Molise "G. Caporale" | Lorusso A, Marcacci M, Di Domenico M, Ancora M, Curini V, Mangone I, Rinaldi A, Scialabba S, Di Pasquale A, Cammà C, Scialabba S, Puglia I, Calistri P, Savini G |
| EPI_ISL_1117480, EPI_ISL_1117481, EPI_ISL_1117482 | SIESP DIP PREV TERAMO | Istituto Zooprofilattico Sperimentale dell'Abruzzo e Molise "G. Caporale" | Lorusso A, Marcacci M, Di Domenico M, Ancora M, Curini V, Mangone I, Rinaldi A, Scialabba S, Di Pasquale A, Cammà C, Scialabba S, Puglia I, Calistri P, Savini G |
| EPI_ISL_1117483, EPI_ISL_1117484 | OSP SAN SALVATORE, MEDICINA INTERNA | Istituto Zooprofilattico Sperimentale dell'Abruzzo e Molise "G. Caporale" | Lorusso A, Marcacci M, Di Domenico M, Ancora M, Curini V, Mangone I, Rinaldi A, Scialabba S, Di Pasquale A, Cammà C, Scialabba S, Puglia I, Calistri P, Savini G |
| EPI_ISL_1117485 | SIESP DIP PREV TERAMO | Istituto Zooprofilattico Sperimentale dell'Abruzzo e Molise "G. Caporale" | Lorusso A, Marcacci M, Di Domenico M, Ancora M, Curini V, Mangone I, Rinaldi A, Scialabba S, Di Pasquale A, Cammà C, Scialabba S, Puglia I, Calistri P, Savini G |
| EPI_ISL_1117486, EPI_ISL_1117487 | SIESP DIP PREV CHIETI | Istituto Zooprofilattico Sperimentale dell'Abruzzo e Molise "G. Caporale" | Lorusso A, Marcacci M, Di Domenico M, Ancora M, Curini V, Mangone I, Rinaldi A, Scialabba S, Di Pasquale A, Cammà C, Scialabba S, Puglia I, Calistri P, Savini G |
| EPI_ISL_1117488 | DIP PREV AVEZZANO, SERVIZIO DI IGIENE EPIDEMIOLOGIA E SANITÀ PUBBLICA | Istituto Zooprofilattico Sperimentale dell'Abruzzo e Molise "G. Caporale" | Lorusso A, Marcacci M, Di Domenico M, Ancora M, Curini V, Mangone I, Rinaldi A, Scialabba S, Di Pasquale A, Cammà C, Scialabba S, Puglia I, Calistri P, Savini G |
| EPI_ISL_1117489 | SIESP DIP PREV TERAMO | Istituto Zooprofilattico Sperimentale dell'Abruzzo e Molise "G. Caporale" | Lorusso A, Marcacci M, Di Domenico M, Ancora M, Curini V, Mangone I, Rinaldi A, Scialabba S, Di Pasquale A, Cammà C, Scialabba S, Puglia I, Calistri P, Savini G |
| EPI_ISL_1117490 | SIESP CHIETI, DRIVE IN VASTO | Istituto Zooprofilattico Sperimentale dell'Abruzzo e Molise "G. Caporale" | Lorusso A, Marcacci M, Di Domenico M, Ancora M, Curini V, Mangone I, Rinaldi A, Scialabba S, Di Pasquale A, Cammà C, Scialabba S, Puglia I, Calistri P, Savini G |
| EPI_ISL_1117491, EPI_ISL_1117492 | SIESP DIP PREV CHIETI | Istituto Zooprofilattico Sperimentale dell'Abruzzo e Molise "G. Caporale" | Lorusso A, Marcacci M, Di Domenico M, Ancora M, Curini V, Mangone I, Rinaldi A, Scialabba S, Di Pasquale A, Cammà C, Scialabba S, Puglia I, Calistri P, Savini G |
| EPI_ISL_1117493 | OSP SAN SALVATORE, MEDICINA INTERNA | Istituto Zooprofilattico Sperimentale dell'Abruzzo e Molise "G. Caporale" | Lorusso A, Marcacci M, Di Domenico M, Ancora M, Curini V, Mangone I, Rinaldi A, Scialabba S, Di Pasquale A, Cammà C, Scialabba S, Puglia I, Calistri P, Savini G |
| EPI_ISL_1117494 | SIESP DIP PREV CHIETI | Istituto Zooprofilattico Sperimentale dell'Abruzzo e Molise "G. Caporale" | Lorusso A, Marcacci M, Di Domenico M, Ancora M, Curini V, Mangone I, Rinaldi A, Scialabba S, Di Pasquale A, Cammà C, Scialabba S, Puglia I, Calistri P, Savini G |
| EPI_ISL_1117495 | SIESP DIP PREV TERAMO | Istituto Zooprofilattico Sperimentale dell'Abruzzo e Molise "G. Caporale" | Lorusso A, Marcacci M, Di Domenico M, Ancora M, Curini V, Mangone I, Rinaldi A, Scialabba S, Di Pasquale A, Cammà C, Scialabba S, Puglia I, Calistri P, Savini G |
| EPI_ISL_1117496 | USCA PESCINA | Istituto Zooprofilattico Sperimentale dell'Abruzzo e Molise "G. Caporale" | Lorusso A, Marcacci M, Di Domenico M, Ancora M, Curini V, Mangone I, Rinaldi A, Scialabba S, Di Pasquale A, Cammà C, Scialabba S, Puglia I, Calistri P, Savini G |
| EPI_ISL_1117497 | SIESP CHIETI, DRIVE IN ORTONA | Istituto Zooprofilattico Sperimentale dell'Abruzzo e Molise "G. Caporale" | Lorusso A, Marcacci M, Di Domenico M, Ancora M, Curini V, Mangone I, Rinaldi A, Scialabba S, Di Pasquale A, Cammà C, Scialabba S, Puglia I, Calistri P, Savini G |
| EPI_ISL_1117498 | PRESIDIO TERRITORIALE CASOLI | Istituto Zooprofilattico Sperimentale dell'Abruzzo e Molise "G. Caporale" | Lorusso A, Marcacci M, Di Domenico M, Ancora M, Curini V, Mangone I, Rinaldi A, Scialabba S, Di Pasquale A, Cammà C, Scialabba S, Puglia I, Calistri P, Savini G |
| EPI_ISL_1117499 | SIESP CHIETI, DRIVE IN VASTO | Istituto Zooprofilattico Sperimentale dell'Abruzzo e Molise "G. Caporale" | Lorusso A, Marcacci M, Di Domenico M, Ancora M, Curini V, Mangone I, Rinaldi A, Scialabba S, Di Pasquale A, Cammà C, Scialabba S, Puglia I, Calistri P, Savini G |
| EPI_ISL_1117500 | SIESP CHIETI, DRIVE IN ORTONA | Istituto Zooprofilattico Sperimentale dell'Abruzzo e Molise "G. Caporale" | Lorusso A, Marcacci M, Di Domenico M, Ancora M, Curini V, Mangone I, Rinaldi A, Scialabba S, Di Pasquale A, Cammà C, Scialabba S, Puglia I, Calistri P, Savini G |
| EPI_ISL_1117501 | OSP SAN SALVATORE, MEDICINA INTERNA | Istituto Zooprofilattico Sperimentale dell'Abruzzo e Molise "G. Caporale" | Lorusso A, Marcacci M, Di Domenico M, Ancora M, Curini V, Mangone I, Rinaldi A, Scialabba S, Di Pasquale A, Cammà C, Scialabba S, Puglia I, Calistri P, Savini G |
| EPI_ISL_1117502 | SIESP DIP PREV CHIETI | Istituto Zooprofilattico Sperimentale dell'Abruzzo e Molise "G. Caporale" | Lorusso A, Marcacci M, Di Domenico M, Ancora M, Curini V, Mangone I, Rinaldi A, Scialabba S, Di Pasquale A, Cammà C, Scialabba S, Puglia I, Calistri P, Savini G |
| EPI_ISL_1117503 | SIESP DIP PREV TERAMO | Istituto Zooprofilattico Sperimentale dell'Abruzzo e Molise "G. Caporale" | Lorusso A, Marcacci M, Di Domenico M, Ancora M, Curini V, Mangone I, Rinaldi A, Scialabba S, Di Pasquale A, Cammà C, Scialabba S, Puglia I, Calistri P, Savini G |
| EPI_ISL_1117504, EPI_ISL_1117505 | OSP SAN SALVATORE, MEDICINA INTERNA | Istituto Zooprofilattico Sperimentale dell'Abruzzo e Molise "G. Caporale" | Lorusso A, Marcacci M, Di Domenico M, Ancora M, Curini V, Mangone I, Rinaldi A, Scialabba S, Di Pasquale A, Cammà C, Scialabba S, Puglia I, Calistri P, Savini G |
| EPI_ISL_1117506 | SIESP DIP PREV CHIETI | Istituto Zooprofilattico Sperimentale dell'Abruzzo e Molise "G. Caporale" | Lorusso A, Marcacci M, Di Domenico M, Ancora M, Curini V, Mangone I, Rinaldi A, Scialabba S, Di Pasquale A, Cammà C, Scialabba S, Puglia I, Calistri P, Savini G |
| EPI_ISL_1117507, EPI_ISL_1117508 | SIESP CHIETI, DRIVE IN LANCIANO | Istituto Zooprofilattico Sperimentale dell'Abruzzo e Molise "G. Caporale" | Lorusso A, Marcacci M, Di Domenico M, Ancora M, Curini V, Mangone I, Rinaldi A, Scialabba S, Di Pasquale A, Cammà C, Scialabba S, Puglia I, Calistri P, Savini G |
| EPI_ISL_1117509, EPI_ISL_1117510, EPI_ISL_1117511, EPI_ISL_1117512, EPI_ISL_1117513 | SIESP DIP PREV CHIETI | Istituto Zooprofilattico Sperimentale dell'Abruzzo e Molise "G. Caporale" | Lorusso A, Marcacci M, Di Domenico M, Ancora M, Curini V, Mangone I, Rinaldi A, Scialabba S, Di Pasquale A, Cammà C, Scialabba S, Puglia I, Calistri P, Savini G |
| EPI_ISL_1117514 | PRES OSP SAN LIBERATORE | Istituto Zooprofilattico Sperimentale dell'Abruzzo e Molise "G. Caporale" | Lorusso A, Marcacci M, Di Domenico M, Ancora M, Curini V, Mangone I, Rinaldi A, Scialabba S, Di Pasquale A, Cammà C, Scialabba S, Puglia I, Calistri P, Savini G |
| EPI_ISL_1117515 | SIESP DIP PREV TERAMO | Istituto Zooprofilattico Sperimentale dell'Abruzzo e Molise "G. Caporale" | Lorusso A, Marcacci M, Di Domenico M, Ancora M, Curini V, Mangone I, Rinaldi A, Scialabba S, Di Pasquale A, Cammà C, Scialabba S, Puglia I, Calistri P, Savini G |
| EPI_ISL_1117516 | SIESP CHIETI, DRIVE IN LANCIANO | Istituto Zooprofilattico Sperimentale dell'Abruzzo e Molise "G. Caporale" | Lorusso A, Marcacci M, Di Domenico M, Ancora M, Curini V, Mangone I, Rinaldi A, Scialabba S, Di Pasquale A, Cammà C, Scialabba S, Puglia I, Calistri P, Savini G |
| EPI_ISL_1117517 | SIESP CHIETI, DRIVE IN ORTONA | Istituto Zooprofilattico Sperimentale dell'Abruzzo e Molise "G. Caporale" | Lorusso A, Marcacci M, Di Domenico M, Ancora M, Curini V, Mangone I, Rinaldi A, Scialabba S, Di Pasquale A, Cammà C, Scialabba S, Puglia I, Calistri P, Savini G |
| EPI_ISL_1117518 | SIESP DIP PREV TERAMO | Istituto Zooprofilattico Sperimentale dell'Abruzzo e Molise "G. Caporale" | Lorusso A, Marcacci M, Di Domenico M, Ancora M, Curini V, Mangone I, Rinaldi A, Scialabba S, Di Pasquale A, Cammà C, Scialabba S, Puglia I, Calistri P, Savini G |
| EPI_ISL_1117519 | SIESP CHIETI, DRIVE IN CHIETI | Istituto Zooprofilattico Sperimentale dell'Abruzzo e Molise "G. Caporale" | Lorusso A, Marcacci M, Di Domenico M, Ancora M, Curini V, Mangone I, Rinaldi A, Scialabba S, Di Pasquale A, Cammà C, Scialabba S, Puglia I, Calistri P, Savini G |
| EPI_ISL_1117520 | USCA AVEZZANO | Istituto Zooprofilattico Sperimentale dell'Abruzzo e Molise "G. Caporale" | Lorusso A, Marcacci M, Di Domenico M, Ancora M, Curini V, Mangone I, Rinaldi A, Scialabba S, Di Pasquale A, Cammà C, Scialabba S, Puglia I, Calistri P, Savini G |
| EPI_ISL_1117521 | DIP PREV AVEZZANO, SERVIZIO DI IGIENE EPIDEMIOLOGIA E SANITÀ PUBBLICA | Istituto Zooprofilattico Sperimentale dell'Abruzzo e Molise "G. Caporale" | Lorusso A, Marcacci M, Di Domenico M, Ancora M, Curini V, Mangone I, Rinaldi A, Scialabba S, Di Pasquale A, Cammà C, Scialabba S, Puglia I, Calistri P, Savini G |
| EPI_ISL_1117522, EPI_ISL_1117523, EPI_ISL_1117524 | SIESP DIP PREV TERAMO | Istituto Zooprofilattico Sperimentale dell'Abruzzo e Molise "G. Caporale" | Lorusso A, Marcacci M, Di Domenico M, Ancora M, Curini V, Mangone I, Rinaldi A, Scialabba S, Di Pasquale A, Cammà C, Scialabba S, Puglia I, Calistri P, Savini G |

| | | | |
|---|---|---|---|
| EPI_ISL_1117525 | OSP SAN SALVATORE, MEDICINA INTERNA | Istituto Zooprofilattico Sperimentale dell'Abruzzo e Molise "G. Caporale" | Lorusso A, Marcacci M, Di Domenico M, Ancora M, Curini V, Mangone I, Rinaldi A, Scialabba S, Di Pasquale A, Cammà C, Scialabba S, Puglia I, Calistri P, Savini G |
| EPI_ISL_1117526 | IZS TERAMO | Istituto Zooprofilattico Sperimentale dell'Abruzzo e Molise "G. Caporale" | Lorusso A, Marcacci M, Di Domenico M, Ancora M, Curini V, Mangone I, Rinaldi A, Scialabba S, Di Pasquale A, Cammà C, Scialabba S, Puglia I, Calistri P, Savini G |
| EPI_ISL_1117661, EPI_ISL_1117662 | SIESP DIP PREV TERAMO | Istituto Zooprofilattico Sperimentale dell'Abruzzo e Molise "G. Caporale" | Lorusso A, Marcacci M, Di Domenico M, Ancora M, Curini V, Mangone I, Rinaldi A, Scialabba S, Di Pasquale A, Cammà C, Puglia I, Calistri P, Savini G |
| EPI_ISL_1117663 | SIESP CHIETI, DRIVE IN ORTONA | Istituto Zooprofilattico Sperimentale dell'Abruzzo e Molise "G. Caporale" | Lorusso A, Marcacci M, Di Domenico M, Ancora M, Curini V, Mangone I, Rinaldi A, Scialabba S, Di Pasquale A, Cammà C, Puglia I, Calistri P, Savini G |
| EPI_ISL_1117664 | SIESP DIP PREV CHIETI | Istituto Zooprofilattico Sperimentale dell'Abruzzo e Molise "G. Caporale" | Lorusso A, Marcacci M, Di Domenico M, Ancora M, Curini V, Mangone I, Rinaldi A, Scialabba S, Di Pasquale A, Cammà C, Puglia I, Calistri P, Savini G |
| EPI_ISL_1117665, EPI_ISL_1117666 | SIESP DIP PREV TERAMO | Istituto Zooprofilattico Sperimentale dell'Abruzzo e Molise "G. Caporale" | Lorusso A, Marcacci M, Di Domenico M, Ancora M, Curini V, Mangone I, Rinaldi A, Scialabba S, Di Pasquale A, Cammà C, Puglia I, Calistri P, Savini G |
| EPI_ISL_1117667 | SIESP CHIETI, DRIVE IN ORTONA | Istituto Zooprofilattico Sperimentale dell'Abruzzo e Molise "G. Caporale" | Lorusso A, Marcacci M, Di Domenico M, Ancora M, Curini V, Mangone I, Rinaldi A, Scialabba S, Di Pasquale A, Cammà C, Puglia I, Calistri P, Savini G |
| EPI_ISL_1117668 | SIESP DIP PREV TERAMO | Istituto Zooprofilattico Sperimentale dell'Abruzzo e Molise "G. Caporale" | Lorusso A, Marcacci M, Di Domenico M, Ancora M, Curini V, Mangone I, Rinaldi A, Scialabba S, Di Pasquale A, Cammà C, Puglia I, Calistri P, Savini G |
| EPI_ISL_1117669 | SIESP CHIETI, DRIVE IN CHIETI | Istituto Zooprofilattico Sperimentale dell'Abruzzo e Molise "G. Caporale" | Lorusso A, Marcacci M, Di Domenico M, Ancora M, Curini V, Mangone I, Rinaldi A, Scialabba S, Di Pasquale A, Cammà C, Puglia I, Calistri P, Savini G |
| EPI_ISL_1117670, EPI_ISL_1117671 | SIESP DIP PREV TERAMO | Istituto Zooprofilattico Sperimentale dell'Abruzzo e Molise "G. Caporale" | Lorusso A, Marcacci M, Di Domenico M, Ancora M, Curini V, Mangone I, Rinaldi A, Scialabba S, Di Pasquale A, Cammà C, Puglia I, Calistri P, Savini G |
| EPI_ISL_1117672 | SIESP DIP PREV CHIETI | Istituto Zooprofilattico Sperimentale dell'Abruzzo e Molise "G. Caporale" | Lorusso A, Marcacci M, Di Domenico M, Ancora M, Curini V, Mangone I, Rinaldi A, Scialabba S, Di Pasquale A, Cammà C, Puglia I, Calistri P, Savini G |
| EPI_ISL_1117673 | SIESP DIP PREV TERAMO | Istituto Zooprofilattico Sperimentale dell'Abruzzo e Molise "G. Caporale" | Lorusso A, Marcacci M, Di Domenico M, Ancora M, Curini V, Mangone I, Rinaldi A, Scialabba S, Di Pasquale A, Cammà C, Puglia I, Calistri P, Savini G |
| EPI_ISL_1117674 | SIESP CHIETI, DRIVE IN CHIETI | Istituto Zooprofilattico Sperimentale dell'Abruzzo e Molise "G. Caporale" | Lorusso A, Marcacci M, Di Domenico M, Ancora M, Curini V, Mangone I, Rinaldi A, Scialabba S, Di Pasquale A, Cammà C, Puglia I, Calistri P, Savini G |
| EPI_ISL_1117675 | DIP PREV AVEZZANO-SERVIZIO DI IGIENE EPIDEMIOLOGIA E SANITÀ PUBBLICA | Istituto Zooprofilattico Sperimentale dell'Abruzzo e Molise "G. Caporale" | Lorusso A, Marcacci M, Di Domenico M, Ancora M, Curini V, Mangone I, Rinaldi A, Scialabba S, Di Pasquale A, Cammà C, Puglia I, Calistri P, Savini G |
| EPI_ISL_1117676 | SIESP CHIETI, DRIVE IN LANCIANO | Istituto Zooprofilattico Sperimentale dell'Abruzzo e Molise "G. Caporale" | Lorusso A, Marcacci M, Di Domenico M, Ancora M, Curini V, Mangone I, Rinaldi A, Scialabba S, Di Pasquale A, Cammà C, Puglia I, Calistri P, Savini G |
| EPI_ISL_1117677 | USCA CASTEL DI SANGRO | Istituto Zooprofilattico Sperimentale dell'Abruzzo e Molise "G. Caporale" | Lorusso A, Marcacci M, Di Domenico M, Ancora M, Curini V, Mangone I, Rinaldi A, Scialabba S, Di Pasquale A, Cammà C, Puglia I, Calistri P, Savini G |
| EPI_ISL_1117678, EPI_ISL_1117679 | SIESP CHIETI, DRIVE IN VASTO | Istituto Zooprofilattico Sperimentale dell'Abruzzo e Molise "G. Caporale" | Lorusso A, Marcacci M, Di Domenico M, Ancora M, Curini V, Mangone I, Rinaldi A, Scialabba S, Di Pasquale A, Cammà C, Puglia I, Calistri P, Savini G |
| EPI_ISL_1117680 | SIESP DIP PREV TERAMO | Istituto Zooprofilattico Sperimentale dell'Abruzzo e Molise "G. Caporale" | Lorusso A, Marcacci M, Di Domenico M, Ancora M, Curini V, Mangone I, Rinaldi A, Scialabba S, Di Pasquale A, Cammà C, Puglia I, Calistri P, Savini G |
| EPI_ISL_1117681 | SIESP CHIETI, DRIVE IN CHIETI | Istituto Zooprofilattico Sperimentale dell'Abruzzo e Molise "G. Caporale" | Lorusso A, Marcacci M, Di Domenico M, Ancora M, Curini V, Mangone I, Rinaldi A, Scialabba S, Di Pasquale A, Cammà C, Puglia I, Calistri P, Savini G |
| EPI_ISL_1117682 | SIESP CHIETI, DRIVE IN LANCIANO | Istituto Zooprofilattico Sperimentale dell'Abruzzo e Molise "G. Caporale" | Lorusso A, Marcacci M, Di Domenico M, Ancora M, Curini V, Mangone I, Rinaldi A, Scialabba S, Di Pasquale A, Cammà C, Puglia I, Calistri P, Savini G |
| EPI_ISL_1117683, EPI_ISL_1117684 | USCA CASTEL DI SANGRO | Istituto Zooprofilattico Sperimentale dell'Abruzzo e Molise "G. Caporale" | Lorusso A, Marcacci M, Di Domenico M, Ancora M, Curini V, Mangone I, Rinaldi A, Scialabba S, Di Pasquale A, Cammà C, Puglia I, Calistri P, Savini G |
| EPI_ISL_1117685 | SIESP DIP PREV TERAMO | Istituto Zooprofilattico Sperimentale dell'Abruzzo e Molise "G. Caporale" | Lorusso A, Marcacci M, Di Domenico M, Ancora M, Curini V, Mangone I, Rinaldi A, Scialabba S, Di Pasquale A, Cammà C, Puglia I, Calistri P, Savini G |
| EPI_ISL_1117686 | DIP PREV AVEZZANO-SERVIZIO DI IGIENE EPIDEMIOLOGIA E SANITÀ PUBBLICA | Istituto Zooprofilattico Sperimentale dell'Abruzzo e Molise "G. Caporale" | Lorusso A, Marcacci M, Di Domenico M, Ancora M, Curini V, Mangone I, Rinaldi A, Scialabba S, Di Pasquale A, Cammà C, Puglia I, Calistri P, Savini G |
| EPI_ISL_1117687 | SIESP CHIETI, DRIVE IN ORTONA | Istituto Zooprofilattico Sperimentale dell'Abruzzo e Molise "G. Caporale" | Lorusso A, Marcacci M, Di Domenico M, Ancora M, Curini V, Mangone I, Rinaldi A, Scialabba S, Di Pasquale A, Cammà C, Puglia I, Calistri P, Savini G |
| EPI_ISL_1117688 | SIESP DIP PREV CHIETI | Istituto Zooprofilattico Sperimentale dell'Abruzzo e Molise "G. Caporale" | Lorusso A, Marcacci M, Di Domenico M, Ancora M, Curini V, Mangone I, Rinaldi A, Scialabba S, Di Pasquale A, Cammà C, Puglia I, Calistri P, Savini G |
| EPI_ISL_1117689 | SIESP CHIETI, DRIVE IN LANCIANO | Istituto Zooprofilattico Sperimentale dell'Abruzzo e Molise "G. Caporale" | Lorusso A, Marcacci M, Di Domenico M, Ancora M, Curini V, Mangone I, Rinaldi A, Scialabba S, Di Pasquale A, Cammà C, Puglia I, Calistri P, Savini G |
| EPI_ISL_1117690 | DIP PREV AVEZZANO-SERVIZIO DI IGIENE EPIDEMIOLOGIA E SANITÀ PUBBLICA | Istituto Zooprofilattico Sperimentale dell'Abruzzo e Molise "G. Caporale" | Lorusso A, Marcacci M, Di Domenico M, Ancora M, Curini V, Mangone I, Rinaldi A, Scialabba S, Di Pasquale A, Cammà C, Puglia I, Calistri P, Savini G |
| EPI_ISL_1117691 | SIESP DIP PREV TERAMO | Istituto Zooprofilattico Sperimentale dell'Abruzzo e Molise "G. Caporale" | Lorusso A, Marcacci M, Di Domenico M, Ancora M, Curini V, Mangone I, Rinaldi A, Scialabba S, Di Pasquale A, Cammà C, Puglia I, Calistri P, Savini G |
| EPI_ISL_1117692 | PRESIDIO OSP SAN LIBERATORE | Istituto Zooprofilattico Sperimentale dell'Abruzzo e Molise "G. Caporale" | Lorusso A, Marcacci M, Di Domenico M, Ancora M, Curini V, Mangone I, Rinaldi A, Scialabba S, Di Pasquale A, Cammà C, Puglia I, Calistri P, Savini G |
| EPI_ISL_1117693 | SIESP DIP PREV TERAMO | Istituto Zooprofilattico Sperimentale dell'Abruzzo e Molise "G. Caporale" | Lorusso A, Marcacci M, Di Domenico M, Ancora M, Curini V, Mangone I, Rinaldi A, Scialabba S, Di Pasquale A, Cammà C, Puglia I, Calistri P, Savini G |
| EPI_ISL_1117694, EPI_ISL_1117695 | SIESP CHIETI, DRIVE IN LANCIANO | Istituto Zooprofilattico Sperimentale dell'Abruzzo e Molise "G. Caporale" | Lorusso A, Marcacci M, Di Domenico M, Ancora M, Curini V, Mangone I, Rinaldi A, Scialabba S, Di Pasquale A, Cammà C, Puglia I, Calistri P, Savini G |
| EPI_ISL_1117696 | SIESP CHIETI, DRIVE IN VASTO | Istituto Zooprofilattico Sperimentale dell'Abruzzo e Molise "G. Caporale" | Lorusso A, Marcacci M, Di Domenico M, Ancora M, Curini V, Mangone I, Rinaldi A, Scialabba S, Di Pasquale A, Cammà C, Puglia I, Calistri P, Savini G |
| EPI_ISL_1117697 | DIP PREV AVEZZANO-SERVIZIO DI IGIENE EPIDEMIOLOGIA E SANITÀ PUBBLICA | Istituto Zooprofilattico Sperimentale dell'Abruzzo e Molise "G. Caporale" | Lorusso A, Marcacci M, Di Domenico M, Ancora M, Curini V, Mangone I, Rinaldi A, Scialabba S, Di Pasquale A, Cammà C, Puglia I, Calistri P, Savini G |
| EPI_ISL_1117698 | SIESP CHIETI, DRIVE IN LANCIANO | Istituto Zooprofilattico Sperimentale dell'Abruzzo e Molise "G. Caporale" | Lorusso A, Marcacci M, Di Domenico M, Ancora M, Curini V, Mangone I, Rinaldi A, Scialabba S, Di Pasquale A, Cammà C, Puglia I, Calistri P, Savini G |
| EPI_ISL_1117699 | SIESP CHIETI, DRIVE IN VASTO | Istituto Zooprofilattico Sperimentale dell'Abruzzo e Molise "G. Caporale" | Lorusso A, Marcacci M, Di Domenico M, Ancora M, Curini V, Mangone I, Rinaldi A, Scialabba S, Di Pasquale A, Cammà C, Puglia I, Calistri P, Savini G |

| | | | |
|---|---|---|---|
| EPI_ISL_1118159, EPI_ISL_1118160, EPI_ISL_1118161, EPI_ISL_1118162, EPI_ISL_1118163, EPI_ISL_1118164, EPI_ISL_1118165, EPI_ISL_1118166, EPI_ISL_1118167, EPI_ISL_1118168, EPI_ISL_1118169, EPI_ISL_1118170, EPI_ISL_1118171, EPI_ISL_1118172, EPI_ISL_1118173, EPI_ISL_1118174, EPI_ISL_1118175, EPI_ISL_1118176, EPI_ISL_1118177, EPI_ISL_1118178, EPI_ISL_1118179, EPI_ISL_1118180, EPI_ISL_1118181, EPI_ISL_1118182, EPI_ISL_1118183, EPI_ISL_1118184, EPI_ISL_1118185, EPI_ISL_1118186, EPI_ISL_1118187, EPI_ISL_1118188, EPI_ISL_1118189, EPI_ISL_1118190, EPI_ISL_1118191, EPI_ISL_1118192, EPI_ISL_1118193, EPI_ISL_1118194, EPI_ISL_1118195, EPI_ISL_1118196, EPI_ISL_1118197, EPI_ISL_1118198, EPI_ISL_1118199, EPI_ISL_1118200, EPI_ISL_1118201, EPI_ISL_1118202, EPI_ISL_1118203, EPI_ISL_1118204 | | | |
| see above | LAB ANALISI PO CARDARELLI CAMPOBASSO | Istituto Zooprofilattico Sperimentale dell'Abruzzo e Molise "G. Caporale" | Scutellà M, Niro G, Felice V. Lorusso A, Marcacci M, Di Domenico M, Ancora M, Curini V, Mangone I, Rinaldi A, Scialabba S, Di Pasquale A, Cammà C, Puglia I, Calistri P, Savini G |
| EPI_ISL_1118217, EPI_ISL_1118223, EPI_ISL_1118224, EPI_ISL_1118231, EPI_ISL_1118232, EPI_ISL_1118233, EPI_ISL_1118234, EPI_ISL_1118235, EPI_ISL_1118236, EPI_ISL_1118237, EPI_ISL_1118238, EPI_ISL_1118239, EPI_ISL_1118240, EPI_ISL_1118241, EPI_ISL_1118242, EPI_ISL_1118243, EPI_ISL_1118244, EPI_ISL_1118245, EPI_ISL_1118246, EPI_ISL_1118247, EPI_ISL_1118248, EPI_ISL_1118249, EPI_ISL_1118250, EPI_ISL_1118251, EPI_ISL_1118252, EPI_ISL_1118253, EPI_ISL_1118254, EPI_ISL_1118255, EPI_ISL_1118256, EPI_ISL_1118257, EPI_ISL_1118258, EPI_ISL_1118259, EPI_ISL_1118260, EPI_ISL_1118261, EPI_ISL_1118264 | | | |
| see above | Virology Lab, Ospedali Riuniti, Ancona | Dipartimento di Scienze Biomediche e Sanità Pubblica, Università Politecnica delle Marche | Sara Caucci, Roberta Longo, Sofia Maria Luigia Tiano, Patrizia Bagnarelli, Stefano Menzo |
| EPI_ISL_1118930, EPI_ISL_1118932, EPI_ISL_1118934, EPI_ISL_1119567 | Laboratorio Microbiologia e Virologia P.O. Cotugno A.O. dei Colli | Laboratorio Microbiologia e Virologia P.O. Cotugno A.O. dei Colli | Luigi Atripaldi, Claudia Tiberio, Anna Perfetti, Pellegrino Cerino, Biancamaria Pierri, Maria Concetta Cuomo |
| EPI_ISL_1120120 | Area Biologia Molecolare - Istituto Zooprofilattico Sperimentale della Sicilia | Area Biologia Molecolare - Istituto Zooprofilattico Sperimentale della Sicilia | REALE Stefano, SCIBETTA Silvia, PIAZZA Angela, BRUNO Gabriella, CASTELLI Germano, BRUNO Federica, VITALE Francesco, TRAMUTO Fabio, MAIDA Carmelo Massimo, MAZZUCCO Walter, DI NARO Daniela, RANDAZZO Giulia, VITALE Fabrizio |
| EPI_ISL_1121032 | SIESP CHIETI, DRIVE IN LANCIANO | Istituto Zooprofilattico Sperimentale dell'Abruzzo e Molise "G. Caporale" | Lorusso A, Marcacci M, Di Domenico M, Ancora M, Curini V, Mangone I, Rinaldi A, Scialabba S, Di Pasquale A, Cammà C, Puglia I, Calistri P, Savini G |
| EPI_ISL_1121033 | USCA CASTEL DI SANGRO | Istituto Zooprofilattico Sperimentale dell'Abruzzo e Molise "G. Caporale" | Lorusso A, Marcacci M, Di Domenico M, Ancora M, Curini V, Mangone I, Rinaldi A, Scialabba S, Di Pasquale A, Cammà C, Puglia I, Calistri P, Savini G |
| EPI_ISL_1121034, EPI_ISL_1121035, EPI_ISL_1121036 | LAB ANALISI PO CARDARELLI CAMPOBASSO | Istituto Zooprofilattico Sperimentale dell'Abruzzo e Molise "G. Caporale" | Scutellà M, Niro G, Felice V. Lorusso A, Marcacci M, Di Domenico M, Ancora M, Curini V, Mangone I, Rinaldi A, Scialabba S, Di Pasquale A, Cammà C, Puglia I, Calistri P, Savini G |
| EPI_ISL_1121037 | SIESP DIP PREV TERAMO | Istituto Zooprofilattico Sperimentale dell'Abruzzo e Molise "G. Caporale" | Lorusso A, Marcacci M, Di Domenico M, Ancora M, Curini V, Mangone I, Rinaldi A, Scialabba S, Di Pasquale A, Cammà C, Scialabba S, Puglia I, Calistri P, Savini G |
| EPI_ISL_1121038 | OSP SAN SALVATORE, MEDICINA INTERNA | Istituto Zooprofilattico Sperimentale dell'Abruzzo e Molise "G. Caporale" | Lorusso A, Marcacci M, Di Domenico M, Ancora M, Curini V, Mangone I, Rinaldi A, Scialabba S, Di Pasquale A, Cammà C, Scialabba S, Puglia I, Calistri P, Savini G |
| EPI_ISL_1123258 | Virologia, Dipartimento di Scienze Biomediche, Università di Sassari, Viale San Pietro, 43/B - Sassari | Laboratorio specialistico UOC Ematologia - Ospedale "San Francesco" - ATS-ASSL Nuoro | Giovanna Piras, Rosanna Asproni, Paolo Malune, Maria Itria Monne, Angelo Domenico Palmas, Caterina Serra, Elena Rimini, Salvatore Rubino |
| EPI_ISL_1123297 | Virology Unit, AOUP | Virology Unit, AOUP | Mamrialinda Vatteroni, Susi Frateschi, Mauro Pistello |
| EPI_ISL_1123298, EPI_ISL_1123299 | Virology Unit, AOUP | Virology Unit | Marialinda Vatteroni, Susi Frateschi, Mauro Pistello |
| EPI_ISL_1123325, EPI_ISL_1123332 | Virology Unit, AOUP | Virology Unit, AOUP | Marialinda Vatteroni, Susi Frateschi, Mauro Pistello |
| EPI_ISL_1123344 | Virology Unit, AOUP | Virology Unit | Marialinda Vatteroni, Susi Frateschi, Mauro Pistello |
| EPI_ISL_1123345 | Virology Unit, AOUP | Virology Unit, AOUP | Marialinda Vatteroni, Susi Frateschi, Mauro Pistello |
| EPI_ISL_1123346, EPI_ISL_1123347 | Virology Unit, AOUP | Virology Unit | Marialinda Vatteroni, Susi Frateschi, Mauro Pistello |
| EPI_ISL_1123349 | Virology Unit, AOUP | Virology Unit, AOUP | Marialinda Vatteroni, Susi Frateschi, Mauro Pistello |
| EPI_ISL_1129215 | Istituto Zooprofilattico Sperimentale del Mezzogiorno | Lab. Microbiologia e Virologia, Cotugno, A.O. dei Colli | Luigi Atripaldi, Claudia Tiberio, Anna Perfetti, Pellegrino Cerino, Biancamaria Pierri, Maria Concetta Cuomo |
| EPI_ISL_1132668 | Azienda Ospedaliero - Universitaria di Modena Policlinico - Virologia e Microbiologia Molecolare | Zooprofilattico Sperimentale dell'Emilia Romagna e della Lombardia (IZSLER), Risk Analysis and Genomic Epidemiology Unit | Monica Pecorari, William Gennari, Giulia Fregni Serpini, Marina Morganti, Ilaria Menozzi, Erika Scaltriti, Stefano Pongolini |
| EPI_ISL_1140102, EPI_ISL_1140103, EPI_ISL_1140104, EPI_ISL_1140105, EPI_ISL_1140106, EPI_ISL_1140107, EPI_ISL_1140108, EPI_ISL_1140109, EPI_ISL_1140110, EPI_ISL_1140111, EPI_ISL_1140112, EPI_ISL_1140113, EPI_ISL_1140114, EPI_ISL_1140115, EPI_ISL_1140116 | | | |
| see above | Università di Parma, Laboratorio di Igiene e Sanità Pubblica | Istituto Zooprofilattico Sperimentale della Lombardia e dell'Emilia Romagna (IZSLER), Risk Analysis and Genomic Epidemiology Unit | Maria Eugenia Colucci, Licia Veronesi, Paola Affanni, Marina Morganti, Ilaria Menozzi, Erika Scaltriti, Stefano Pongolini |
| EPI_ISL_1154361, EPI_ISL_1154890 | Area Biologia Molecolare - Istituto Zooprofilattico Sperimentale della Sicilia | Area Biologia Molecolare - Istituto Zooprofilattico Sperimentale della Sicilia | REALE Stefano, SCIBETTA Silvia, PIAZZA Angela, BRUNO Gabriella, CASTELLI Germano, BRUNO Federica, VITALE Francesco, TRAMUTO Fabio, MAIDA Carmelo Massimo, MAZZUCCO Walter, DI NARO Daniela, RANDAZZO Giulia, VITALE Fabrizio |
| EPI_ISL_1157894, EPI_ISL_1157895, EPI_ISL_1157896, EPI_ISL_1157897, EPI_ISL_1157898, EPI_ISL_1157899, EPI_ISL_1157900, EPI_ISL_1157901, EPI_ISL_1157902, EPI_ISL_1157903, EPI_ISL_1157904, EPI_ISL_1157905, EPI_ISL_1157906, EPI_ISL_1157907, EPI_ISL_1157908, EPI_ISL_1157909, EPI_ISL_1157910, EPI_ISL_1157911, EPI_ISL_1157912, EPI_ISL_1157913, EPI_ISL_1157914, EPI_ISL_1157915, EPI_ISL_1157916, EPI_ISL_1157917, EPI_ISL_1157918, EPI_ISL_1157919, EPI_ISL_1157920, EPI_ISL_1157921, EPI_ISL_1157922, EPI_ISL_1157923, EPI_ISL_1157924, EPI_ISL_1157925, EPI_ISL_1157926, EPI_ISL_1157927, EPI_ISL_1157928, EPI_ISL_1157929, EPI_ISL_1157930, EPI_ISL_1157931, EPI_ISL_1157932, EPI_ISL_1157933, EPI_ISL_1157934, EPI_ISL_1157935, EPI_ISL_1157936, EPI_ISL_1157937, EPI_ISL_1157938, EPI_ISL_1157939, EPI_ISL_1157940, EPI_ISL_1157941, EPI_ISL_1157942, EPI_ISL_1157943, EPI_ISL_1157944, EPI_ISL_1157945, EPI_ISL_1157946, EPI_ISL_1157947, EPI_ISL_1157948, EPI_ISL_1157949, EPI_ISL_1157950, EPI_ISL_1157951, EPI_ISL_1157952, EPI_ISL_1157953, EPI_ISL_1157954, EPI_ISL_1157955, EPI_ISL_1157956, EPI_ISL_1157957, EPI_ISL_1157958, EPI_ISL_1157959, EPI_ISL_1157960, EPI_ISL_1157961, EPI_ISL_1157962, EPI_ISL_1157963, EPI_ISL_1157964, EPI_ISL_1157965, EPI_ISL_1157966, EPI_ISL_1157967, EPI_ISL_1157968, EPI_ISL_1157969, EPI_ISL_1157970, EPI_ISL_1157971, EPI_ISL_1157972, EPI_ISL_1157973, EPI_ISL_1157974, EPI_ISL_1157975, EPI_ISL_1157976, EPI_ISL_1157977, EPI_ISL_1157978, EPI_ISL_1157979, EPI_ISL_1157980, EPI_ISL_1157981, EPI_ISL_1157982, EPI_ISL_1157983, EPI_ISL_1157984, EPI_ISL_1157985, EPI_ISL_1157986, EPI_ISL_1157987, EPI_ISL_1157988, EPI_ISL_1157989, EPI_ISL_1157990, EPI_ISL_1157991, EPI_ISL_1157992, EPI_ISL_1157993, EPI_ISL_1157994, EPI_ISL_1157995, EPI_ISL_1157996, EPI_ISL_1157997, EPI_ISL_1157998, EPI_ISL_1157999, EPI_ISL_1158000, EPI_ISL_1158001, EPI_ISL_1158002, EPI_ISL_1158003, EPI_ISL_1158004, EPI_ISL_1158005, EPI_ISL_1158006, EPI_ISL_1158007, EPI_ISL_1158008, EPI_ISL_1158009, EPI_ISL_1158010, EPI_ISL_1158011, EPI_ISL_1158012, EPI_ISL_1158013, EPI_ISL_1158014, EPI_ISL_1158015, EPI_ISL_1158016, EPI_ISL_1158017, EPI_ISL_1158018, EPI_ISL_1158019, EPI_ISL_1158020, EPI_ISL_1158021, EPI_ISL_1158022, EPI_ISL_1158023, EPI_ISL_1158024, EPI_ISL_1158025, EPI_ISL_1158026, EPI_ISL_1158027, EPI_ISL_1158028, EPI_ISL_1158029, EPI_ISL_1158030 | | | |
| see above | ASL Napoli 1 Centro | AMES Centro Polidiagnostico Strumentale S.r.l. | "Giovanni Savarese, Raffaella Ruggiero, Eloisa Evangelista, Antonella Di Carlo, Luisa Circelli, Luigi D'Amore, Roberto Sirica, Nadia Petrillo, Monica Ianniello, Antonio Fico" |
| EPI_ISL_1158846 | Unità Operativa di Microbiologia, IRCCS Policlinico di Sant'Orsola, Azienda Ospedaliero-Universitaria di Bologna | U.O. Microbiologia Laboratorio Unico Centro Servizi AUSL della Romagna | Giada Rossini, Giuliano Furlini, Tiziana Lazzarotto, Giorgio Dirani, Silvia Zannoli, Vittorio Sambri |
| EPI_ISL_1163688, EPI_ISL_1163689, EPI_ISL_1163690, EPI_ISL_1163691, EPI_ISL_1163692 | UOC Microbiologia e Virologia, Azienda Ospedaliera Universitaria Senese, Siena, Italy | Dipartimento di Biotecnologie Mediche | Maria Grazia Cusi, David Pinzauti, Claudia Gandolfo, Gabriele Anichini, Gianni Pozzi, Gianni Gori Savellini, Francesco Santoro |
| EPI_ISL_1163795, EPI_ISL_1163796, EPI_ISL_1163797 | Azienda USL Umbria 2 | Istituto Zooprofilattico Sperimentale dell'Abruzzo e Molise "G. Caporale" | Proietti A., Pistoni E., Lorusso A, Marcacci M, Di Domenico M, Ancora M, Curini V, Mangone I, Rinaldi A, Scialabba S, Di Pasquale A, Cammà C, Puglia I, Calistri P, Savini G |
| EPI_ISL_1163798, EPI_ISL_1163799, EPI_ISL_1163800, EPI_ISL_1163801, EPI_ISL_1163802, EPI_ISL_1163803, EPI_ISL_1163804, EPI_ISL_1163805, EPI_ISL_1163806, EPI_ISL_1163807, EPI_ISL_1163808, EPI_ISL_1163809, EPI_ISL_1163810, EPI_ISL_1163811, EPI_ISL_1163812, EPI_ISL_1163813, EPI_ISL_1163814, EPI_ISL_1163815, EPI_ISL_1163816, EPI_ISL_1163817, EPI_ISL_1163818, EPI_ISL_1163819, EPI_ISL_1163820, EPI_ISL_1163821, EPI_ISL_1163822, EPI_ISL_1163823, EPI_ISL_1163824, EPI_ISL_1163825, EPI_ISL_1163826, EPI_ISL_1163827, EPI_ISL_1163828 | | | |
| see above | Università degli Studi di Perugia | Istituto Zooprofilattico Sperimentale dell'Abruzzo e Molise "G. Caporale" | Mencacci A., Camilloni B., Lorusso A, Marcacci M, Di Domenico M, Ancora M, Curini V, Mangone I, Rinaldi A, Scialabba S, Di Pasquale A, Cammà C, Puglia I, Calistri P, Savini G |
| EPI_ISL_1163829, EPI_ISL_1163830 | Istituto Zooprofilattico Sperimentale Umbria e Marche "Togo Rosati" | Istituto Zooprofilattico Sperimentale dell'Abruzzo e Molise "G. Caporale" | Biagetti M., Giammarioli M., Lorusso A, Marcacci M, Di Domenico M, Ancora M, Curini V, Mangone I, Rinaldi A, Scialabba S, Di Pasquale A, Cammà C, Puglia I, Calistri P, Savini G |
| EPI_ISL_1163889, EPI_ISL_1163890, EPI_ISL_1163891, EPI_ISL_1163892, EPI_ISL_1163893, EPI_ISL_1163895 | IZSM | IZSM-U.O.C. Virologia | Maurizio Viscardi, Lorena Cardillo, Luigi Atripaldi,Giovanna Fusco, Esterina De Carlo, Antonio Limone |
| EPI_ISL_1163910 | IZSM | IZSM-U.O.C. Virologia | Maurizio Viscardi, Lorena Cardillo, Giuseppe Prota, Vincenzo Petrella, Luigi Atripaldi, Giovanna Fusco, Esterina De Carlo, Antonio Limone |

| | | | |
|---|---|---|---|
| EPI_ISL_1163911, EPI_ISL_1163912, EPI_ISL_1163913 | IZSM | IZSM-U.O.C. Virologia | Maurizio Viscardi, Lorena Cardillo, Luigi Atripaldi,Giovanna Fusco, Esterina De Carlo, Antonio Limone |
| EPI_ISL_1164754, EPI_ISL_1164755, EPI_ISL_1164756 | Azienda USL Umbria 2 | Istituto Zooprofilattico Sperimentale dell'Abruzzo e Molise "G. Caporale" | Proietti A, Pistoni E, Lorusso A, Marcacci M, Di Domenico M, Ancora M, Curini V, Mangone I, Rinaldi A, Scialabba S, Di Pasquale A, Cammà C, Puglia I, Calistri P, Savini G |
| EPI_ISL_1164816, EPI_ISL_1164853, EPI_ISL_1164854, EPI_ISL_1164930, EPI_ISL_1164969, EPI_ISL_1165000, EPI_ISL_1165066, EPI_ISL_1165077, EPI_ISL_1165138, EPI_ISL_1165143, EPI_ISL_1165899, EPI_ISL_1166083 | | | |
| see above | Struttura Semplice Dipartimentale di Virologia e Microbiologia molecolare, Azienda Ospedaliero-Universitaria, Policlinico di Modena | U.O. Microbiologia, Laboratorio Unico Centro Servizi - AUSL della Romagna | Pecorari Monica, Gennari William, Fregni Serpini Giulia, Giorgio Dirani, Silvia Zannoli, Vittorio Sambri |
| EPI_ISL_1166147, EPI_ISL_1166148 | Laboratorio Analisi Osp. Città di Castello - Azienda USL Umbria1 | Istituto Zooprofilattico Sperimentale dell'Abruzzo e Molise "G. Caporale" | Malagigi V, Tacconi P, Lorusso A, Marcacci M, Di Domenico M, Ancora M, Curini V, Mangone I, Rinaldi A, Scialabba S, Di Pasquale A, Cammà C, Puglia I, Calistri P, Savini G |
| EPI_ISL_1166149, EPI_ISL_1166150, EPI_ISL_1166151, EPI_ISL_1166152, EPI_ISL_1166153, EPI_ISL_1166154, EPI_ISL_1166155, EPI_ISL_1166156, EPI_ISL_1166157, EPI_ISL_1166158, EPI_ISL_1166159, EPI_ISL_1166160, EPI_ISL_1166161, EPI_ISL_1166162, EPI_ISL_1166163, EPI_ISL_1166164, EPI_ISL_1166165, EPI_ISL_1166166, EPI_ISL_1166167, EPI_ISL_1166168, EPI_ISL_1166169, EPI_ISL_1166170, EPI_ISL_1166171 | | | |
| see above | Università degli Studi di Perugia | Istituto Zooprofilattico Sperimentale dell'Abruzzo e Molise "G. Caporale" | Mencacci A, Camilloni B, Lorusso A, Marcacci M, Di Domenico M, Ancora M, Curini V, Mangone I, Rinaldi A, Scialabba S, Di Pasquale A, Cammà C, Puglia I, Calistri P, Savini G |
| EPI_ISL_1166172, EPI_ISL_1166173, EPI_ISL_1166174, EPI_ISL_1166175, EPI_ISL_1166176, EPI_ISL_1166177, EPI_ISL_1166178, EPI_ISL_1166179, EPI_ISL_1166180, EPI_ISL_1166181, EPI_ISL_1166182, EPI_ISL_1166183, EPI_ISL_1166184, EPI_ISL_1166185, EPI_ISL_1166186, EPI_ISL_1166187, EPI_ISL_1166188 | | | |
| see above | Istituto Zooprofilattico Sperimentale Umbria e Marche "Togo Rosati" | Istituto Zooprofilattico Sperimentale dell'Abruzzo e Molise "G. Caporale" | Biagetti M, Giammarioli M, Lorusso A, Marcacci M, Di Domenico M, Ancora M, Curini V, Mangone I, Rinaldi A, Scialabba S, Di Pasquale A, Cammà C, Puglia I, Calistri P, Savini G |
| EPI_ISL_1166221, EPI_ISL_1166222, EPI_ISL_1166223, EPI_ISL_1166224, EPI_ISL_1166225, EPI_ISL_1166226, EPI_ISL_1166227, EPI_ISL_1166228, EPI_ISL_1166229, EPI_ISL_1166230, EPI_ISL_1166231, EPI_ISL_1166232, EPI_ISL_1166233, EPI_ISL_1166234, EPI_ISL_1166235, EPI_ISL_1166236, EPI_ISL_1166237, EPI_ISL_1166238, EPI_ISL_1166239, EPI_ISL_1166240, EPI_ISL_1166241, EPI_ISL_1166242, EPI_ISL_1166243, EPI_ISL_1166244, EPI_ISL_1166245, EPI_ISL_1166246, EPI_ISL_1166247, EPI_ISL_1166248, EPI_ISL_1166249, EPI_ISL_1166250, EPI_ISL_1166251, EPI_ISL_1166252, EPI_ISL_1166253, EPI_ISL_1166254, EPI_ISL_1166255, EPI_ISL_1166256, EPI_ISL_1166257, EPI_ISL_1166258, EPI_ISL_1166259, EPI_ISL_1166260, EPI_ISL_1166261, EPI_ISL_1166262, EPI_ISL_1166263, EPI_ISL_1166264, EPI_ISL_1166265, EPI_ISL_1166266, EPI_ISL_1166267, EPI_ISL_1166268, EPI_ISL_1166269, EPI_ISL_1166270, EPI_ISL_1166271, EPI_ISL_1166272, EPI_ISL_1166273, EPI_ISL_1166274, EPI_ISL_1166275, EPI_ISL_1166276, EPI_ISL_1166277, EPI_ISL_1166278, EPI_ISL_1166279, EPI_ISL_1166280, EPI_ISL_1166281, EPI_ISL_1166282, EPI_ISL_1166283, EPI_ISL_1166284, EPI_ISL_1166285, EPI_ISL_1166286, EPI_ISL_1166287, EPI_ISL_1166288, EPI_ISL_1166289, EPI_ISL_1166290, EPI_ISL_1166291, EPI_ISL_1166292, EPI_ISL_1166293, EPI_ISL_1166294, EPI_ISL_1166295, EPI_ISL_1166296, EPI_ISL_1166297, EPI_ISL_1166298, EPI_ISL_1166299, EPI_ISL_1166300, EPI_ISL_1166301, EPI_ISL_1166302, EPI_ISL_1166303, EPI_ISL_1166304, EPI_ISL_1166305, EPI_ISL_1166306, EPI_ISL_1166307, EPI_ISL_1166308, EPI_ISL_1166309, EPI_ISL_1166310, EPI_ISL_1166311, EPI_ISL_1166312, EPI_ISL_1166313, EPI_ISL_1166314, EPI_ISL_1166315, EPI_ISL_1166316, EPI_ISL_1166317, EPI_ISL_1166318, EPI_ISL_1166319, EPI_ISL_1166320, EPI_ISL_1166321, EPI_ISL_1166322, EPI_ISL_1166323, EPI_ISL_1166324, EPI_ISL_1166325, EPI_ISL_1166326, EPI_ISL_1166327, EPI_ISL_1166328, EPI_ISL_1166329, EPI_ISL_1166330, EPI_ISL_1166331, EPI_ISL_1166332, EPI_ISL_1166333, EPI_ISL_1166334, EPI_ISL_1166335, EPI_ISL_1166336, EPI_ISL_1166337, EPI_ISL_1166338, EPI_ISL_1166339, EPI_ISL_1166340, EPI_ISL_1166341, EPI_ISL_1166342, EPI_ISL_1166343, EPI_ISL_1166344, EPI_ISL_1166345, EPI_ISL_1166346, EPI_ISL_1166347, EPI_ISL_1166348, EPI_ISL_1166349, EPI_ISL_1166350, EPI_ISL_1166351, EPI_ISL_1166352, EPI_ISL_1166353, EPI_ISL_1166354, EPI_ISL_1166355, EPI_ISL_1166356, EPI_ISL_1166357, EPI_ISL_1166358, EPI_ISL_1166359, EPI_ISL_1166360, EPI_ISL_1166361, EPI_ISL_1166362, EPI_ISL_1166363, EPI_ISL_1166364, EPI_ISL_1166365, EPI_ISL_1166366, EPI_ISL_1166367, EPI_ISL_1166368, EPI_ISL_1166369, EPI_ISL_1166370, EPI_ISL_1166371, EPI_ISL_1166372, EPI_ISL_1166373, EPI_ISL_1166374, EPI_ISL_1166375, EPI_ISL_1166376, EPI_ISL_1166377, EPI_ISL_1166378, EPI_ISL_1166379, EPI_ISL_1166380, EPI_ISL_1166381, EPI_ISL_1166382, EPI_ISL_1166383, EPI_ISL_1166384, EPI_ISL_1166385, EPI_ISL_1166386, EPI_ISL_1166387, EPI_ISL_1166388, EPI_ISL_1166389, EPI_ISL_1166390, EPI_ISL_1166391, EPI_ISL_1166392, EPI_ISL_1166393, EPI_ISL_1166394, EPI_ISL_1166395, EPI_ISL_1166396, EPI_ISL_1166397, EPI_ISL_1166398, EPI_ISL_1166399, EPI_ISL_1166400, EPI_ISL_1166401, EPI_ISL_1166402, EPI_ISL_1166403, EPI_ISL_1166404, EPI_ISL_1166405, EPI_ISL_1166406, EPI_ISL_1166407, EPI_ISL_1166408, EPI_ISL_1166409, EPI_ISL_1166410, EPI_ISL_1166411, EPI_ISL_1166412, EPI_ISL_1166413, EPI_ISL_1166414, EPI_ISL_1166415, EPI_ISL_1166416, EPI_ISL_1166417, EPI_ISL_1166418, EPI_ISL_1166419, EPI_ISL_1166420, EPI_ISL_1166421, EPI_ISL_1166422, EPI_ISL_1166423, EPI_ISL_1166424, EPI_ISL_1166425, EPI_ISL_1166426, EPI_ISL_1166427, EPI_ISL_1166428, EPI_ISL_1166429, EPI_ISL_1166430, EPI_ISL_1166431, EPI_ISL_1166432, EPI_ISL_1166433, EPI_ISL_1166434, EPI_ISL_1166435, EPI_ISL_1166436, EPI_ISL_1166437, EPI_ISL_1166438, EPI_ISL_1166439, EPI_ISL_1166440, EPI_ISL_1166441, EPI_ISL_1166442, EPI_ISL_1166443, EPI_ISL_1166444, EPI_ISL_1166445, EPI_ISL_1166446, EPI_ISL_1166447, EPI_ISL_1166448, EPI_ISL_1166449, EPI_ISL_1166450, EPI_ISL_1166451, EPI_ISL_1166452, EPI_ISL_1166453, EPI_ISL_1166454, EPI_ISL_1166455, EPI_ISL_1166456, EPI_ISL_1166457, EPI_ISL_1166458, EPI_ISL_1166459, EPI_ISL_1166460, EPI_ISL_1166461, EPI_ISL_1166462, EPI_ISL_1166463, EPI_ISL_1166464, EPI_ISL_1166465, EPI_ISL_1166466, EPI_ISL_1166467, EPI_ISL_1166468, EPI_ISL_1166469, EPI_ISL_1166470, EPI_ISL_1166471, EPI_ISL_1166472, EPI_ISL_1166473, EPI_ISL_1166474, EPI_ISL_1166475, EPI_ISL_1166476, EPI_ISL_1166477, EPI_ISL_1166478, EPI_ISL_1166479, EPI_ISL_1166480, EPI_ISL_1166481, EPI_ISL_1166482, EPI_ISL_1166483, EPI_ISL_1166484, EPI_ISL_1166485, EPI_ISL_1166486, EPI_ISL_1166487, EPI_ISL_1166488, EPI_ISL_1166489, EPI_ISL_1166490, EPI_ISL_1166491, EPI_ISL_1166492, EPI_ISL_1166493, EPI_ISL_1166494, EPI_ISL_1166495, EPI_ISL_1166496, EPI_ISL_1166497, EPI_ISL_1166498, EPI_ISL_1166499, EPI_ISL_1166500, EPI_ISL_1166501, EPI_ISL_1166502, EPI_ISL_1166503, EPI_ISL_1166504, EPI_ISL_1166505, EPI_ISL_1166506, EPI_ISL_1166507, EPI_ISL_1166508, EPI_ISL_1166509, EPI_ISL_1166510, EPI_ISL_1166511, EPI_ISL_1166512, EPI_ISL_1166513, EPI_ISL_1166514, EPI_ISL_1166515, EPI_ISL_1166516, EPI_ISL_1166517, EPI_ISL_1166518, EPI_ISL_1166519, EPI_ISL_1166520, EPI_ISL_1166521, EPI_ISL_1166522, EPI_ISL_1166523, EPI_ISL_1166524, EPI_ISL_1166525, EPI_ISL_1166526, EPI_ISL_1166527, EPI_ISL_1166528, EPI_ISL_1166529, EPI_ISL_1166530, EPI_ISL_1166531, EPI_ISL_1166532, EPI_ISL_1166533 | | | |
| see above | ASL Napoli 1 Centro | AMES Centro Polidiagnostico Strumentale S.r.l. | "Giovanni Savarese, Raffaella Ruggiero, Eloisa Evangelista, Antonella Di Carlo, Luisa Circelli, Luigi D'Amore, Nadia Petrillo, Monica Ianniello, Roberto Sirica,Maurizio D'Amora, Antonio Fico" |
| EPI_ISL_1166877, EPI_ISL_1166878, EPI_ISL_1166879, EPI_ISL_1166880, EPI_ISL_1166881, EPI_ISL_1166882 | IZSM | IZSM-U.O.C. Virologia | Maurizio Viscardi, Lorena Cardillo, Luigi Atripaldi,Giovanna Fusco, Esterina De Carlo, Antonio Limone |
| EPI_ISL_1167308, EPI_ISL_1167309, EPI_ISL_1167310, EPI_ISL_1167311, EPI_ISL_1167312, EPI_ISL_1167313, EPI_ISL_1167314, EPI_ISL_1167315, EPI_ISL_1167316, EPI_ISL_1167317, EPI_ISL_1167318, EPI_ISL_1167319, EPI_ISL_1167320, EPI_ISL_1167321, EPI_ISL_1167322, EPI_ISL_1167323, EPI_ISL_1167324, EPI_ISL_1167325, EPI_ISL_1167326, EPI_ISL_1167327, EPI_ISL_1167328, EPI_ISL_1167329, EPI_ISL_1167330, EPI_ISL_1167331, EPI_ISL_1167332, EPI_ISL_1167333, EPI_ISL_1167334, EPI_ISL_1167335, EPI_ISL_1167336, EPI_ISL_1167337, EPI_ISL_1167338, EPI_ISL_1167339, EPI_ISL_1167340, EPI_ISL_1167341, EPI_ISL_1167342, EPI_ISL_1167343, EPI_ISL_1167344, EPI_ISL_1167345, EPI_ISL_1167346, EPI_ISL_1167347, EPI_ISL_1167348, EPI_ISL_1167349, EPI_ISL_1167350, EPI_ISL_1167351, EPI_ISL_1167352, EPI_ISL_1167353, EPI_ISL_1167354, EPI_ISL_1167355, EPI_ISL_1167356, EPI_ISL_1167357, EPI_ISL_1167358, EPI_ISL_1167359, EPI_ISL_1167360, EPI_ISL_1167361, EPI_ISL_1167362, EPI_ISL_1167363, EPI_ISL_1167364, EPI_ISL_1167365, EPI_ISL_1167366, EPI_ISL_1167367, EPI_ISL_1167368, EPI_ISL_1167369, EPI_ISL_1167370, EPI_ISL_1167371, EPI_ISL_1167372, EPI_ISL_1167373, EPI_ISL_1167374, EPI_ISL_1167375, EPI_ISL_1167376, EPI_ISL_1167377, EPI_ISL_1167378, EPI_ISL_1167379, EPI_ISL_1167380, EPI_ISL_1167381, EPI_ISL_1167382, EPI_ISL_1167383, EPI_ISL_1167384, EPI_ISL_1167385, EPI_ISL_1167386, EPI_ISL_1167387, EPI_ISL_1167388, EPI_ISL_1167389, EPI_ISL_1167390, EPI_ISL_1167391, EPI_ISL_1167392, EPI_ISL_1167393, EPI_ISL_1167394, EPI_ISL_1167395, EPI_ISL_1167396, EPI_ISL_1167397, EPI_ISL_1167398, EPI_ISL_1167399, EPI_ISL_1167400, EPI_ISL_1167401, EPI_ISL_1167402, EPI_ISL_1167403, EPI_ISL_1167404, EPI_ISL_1167405, EPI_ISL_1167406, EPI_ISL_1167407, EPI_ISL_1167408, EPI_ISL_1167409, EPI_ISL_1167410, EPI_ISL_1167411, EPI_ISL_1167412, EPI_ISL_1167413, EPI_ISL_1167414, EPI_ISL_1167415, EPI_ISL_1167416, EPI_ISL_1167417, EPI_ISL_1167418, EPI_ISL_1167419, EPI_ISL_1167420, EPI_ISL_1167421, EPI_ISL_1167422, EPI_ISL_1167423, EPI_ISL_1167424, EPI_ISL_1167425, EPI_ISL_1167426, EPI_ISL_1167427, EPI_ISL_1167428, EPI_ISL_1167429, EPI_ISL_1167430, EPI_ISL_1167431, EPI_ISL_1167432, EPI_ISL_1167433, EPI_ISL_1167434, EPI_ISL_1167435, EPI_ISL_1167436, EPI_ISL_1167437, EPI_ISL_1167438, EPI_ISL_1167439, EPI_ISL_1167440, EPI_ISL_1167441, EPI_ISL_1167442, EPI_ISL_1167443, EPI_ISL_1167444, EPI_ISL_1167445, EPI_ISL_1167446, EPI_ISL_1167447, EPI_ISL_1167448, EPI_ISL_1167449, EPI_ISL_1167450, EPI_ISL_1167451, EPI_ISL_1167452, EPI_ISL_1167453, EPI_ISL_1167454, EPI_ISL_1167455, EPI_ISL_1167456, EPI_ISL_1167457, EPI_ISL_1167458, EPI_ISL_1167459, EPI_ISL_1167460, EPI_ISL_1167461, EPI_ISL_1167462, EPI_ISL_1167463, EPI_ISL_1167464, EPI_ISL_1167465, EPI_ISL_1167466, EPI_ISL_1167467, EPI_ISL_1167468, EPI_ISL_1167469, EPI_ISL_1167470, EPI_ISL_1167471, EPI_ISL_1167472, EPI_ISL_1167473, EPI_ISL_1167474, EPI_ISL_1167475, EPI_ISL_1167476, EPI_ISL_1167477, EPI_ISL_1167478, EPI_ISL_1167479, EPI_ISL_1167480, EPI_ISL_1167481, EPI_ISL_1167482, EPI_ISL_1167483, EPI_ISL_1167484, EPI_ISL_1167485, EPI_ISL_1167486, EPI_ISL_1167487, EPI_ISL_1167488, EPI_ISL_1167489, EPI_ISL_1167490, EPI_ISL_1167491, EPI_ISL_1167492, EPI_ISL_1167493, EPI_ISL_1167494, EPI_ISL_1167495, EPI_ISL_1167496, EPI_ISL_1167497, EPI_ISL_1167498, EPI_ISL_1167499, EPI_ISL_1167500, EPI_ISL_1167501, EPI_ISL_1167502, EPI_ISL_1167503, EPI_ISL_1167504, EPI_ISL_1167505, EPI_ISL_1167506, EPI_ISL_1167507, EPI_ISL_1167508, EPI_ISL_1167509, EPI_ISL_1167510, EPI_ISL_1167511, EPI_ISL_1167512, EPI_ISL_1167513, EPI_ISL_1167514, EPI_ISL_1167515, EPI_ISL_1167516, EPI_ISL_1167517, EPI_ISL_1167518, EPI_ISL_1167519, EPI_ISL_1167520, EPI_ISL_1167521, EPI_ISL_1167522, EPI_ISL_1167523, EPI_ISL_1167524, EPI_ISL_1167525, EPI_ISL_1167526, EPI_ISL_1167527, EPI_ISL_1167528, EPI_ISL_1167529, EPI_ISL_1167530, EPI_ISL_1167531, EPI_ISL_1167532, EPI_ISL_1167533, EPI_ISL_1167534, EPI_ISL_1167535, EPI_ISL_1167536, EPI_ISL_1167537, EPI_ISL_1167538, EPI_ISL_1167539, EPI_ISL_1167540, EPI_ISL_1167541, EPI_ISL_1167542, EPI_ISL_1167543, EPI_ISL_1167544, EPI_ISL_1167545, EPI_ISL_1167546, EPI_ISL_1169061, EPI_ISL_1169062, EPI_ISL_1169063, EPI_ISL_1169064, EPI_ISL_1169065, EPI_ISL_1169066, EPI_ISL_1169067, EPI_ISL_1169068, EPI_ISL_1169069, EPI_ISL_1169070, EPI_ISL_1169071, EPI_ISL_1169072, EPI_ISL_1169073, EPI_ISL_1169074, EPI_ISL_1169075, EPI_ISL_1169076, EPI_ISL_1169077, EPI_ISL_1169078, EPI_ISL_1169079, EPI_ISL_1169080, EPI_ISL_1169081, EPI_ISL_1169082, EPI_ISL_1169083, EPI_ISL_1169084, EPI_ISL_1169085, EPI_ISL_1169086, EPI_ISL_1169087, EPI_ISL_1169088, EPI_ISL_1169089, EPI_ISL_1169090, EPI_ISL_1169091, EPI_ISL_1169092, EPI_ISL_1169093, EPI_ISL_1169094, EPI_ISL_1169095, EPI_ISL_1169096, EPI_ISL_1169097, EPI_ISL_1169098, EPI_ISL_1169099, EPI_ISL_1169100, EPI_ISL_1169101, EPI_ISL_1169102, EPI_ISL_1169103, EPI_ISL_1169104, EPI_ISL_1169105, EPI_ISL_1169106, EPI_ISL_1169107, EPI_ISL_1169108, EPI_ISL_1169109, EPI_ISL_1169110, EPI_ISL_1169111, EPI_ISL_1169112, EPI_ISL_1169113, EPI_ISL_1169114, EPI_ISL_1169115, EPI_ISL_1169116, EPI_ISL_1169117, EPI_ISL_1169118, EPI_ISL_1169119, EPI_ISL_1169120, EPI_ISL_1169121, EPI_ISL_1169122, EPI_ISL_1169123, EPI_ISL_1169124, EPI_ISL_1169125, EPI_ISL_1169126, EPI_ISL_1169127, EPI_ISL_1169128, EPI_ISL_1169129, EPI_ISL_1169130, EPI_ISL_1169131, EPI_ISL_1169132, EPI_ISL_1169133, EPI_ISL_1169134, EPI_ISL_1169135, EPI_ISL_1169136, EPI_ISL_1169137, EPI_ISL_1169138, EPI_ISL_1169139, EPI_ISL_1169140, EPI_ISL_1169141, EPI_ISL_1169142, EPI_ISL_1169143, EPI_ISL_1169144, EPI_ISL_1169145, EPI_ISL_1169146, EPI_ISL_1169147, EPI_ISL_1169148, EPI_ISL_1169149, EPI_ISL_1169150, EPI_ISL_1169151, EPI_ISL_1169152, EPI_ISL_1169153, EPI_ISL_1169154, EPI_ISL_1169155, EPI_ISL_1169156, EPI_ISL_1169157, EPI_ISL_1169158, EPI_ISL_1169159, EPI_ISL_1169160, EPI_ISL_1169161, EPI_ISL_1169162, EPI_ISL_1169163, EPI_ISL_1169164, EPI_ISL_1169165, EPI_ISL_1169166, EPI_ISL_1169167, EPI_ISL_1169168, EPI_ISL_1169169, EPI_ISL_1169170, EPI_ISL_1169171, EPI_ISL_1169172, EPI_ISL_1169173, EPI_ISL_1169174, EPI_ISL_1169175, EPI_ISL_1169176, EPI_ISL_1169177, EPI_ISL_1169178, EPI_ISL_1169179, EPI_ISL_1169180, EPI_ISL_1169181, EPI_ISL_1169182, EPI_ISL_1169183, EPI_ISL_1169184, EPI_ISL_1169185, EPI_ISL_1169186, EPI_ISL_1169187, EPI_ISL_1169188, EPI_ISL_1169189, EPI_ISL_1169190, EPI_ISL_1169191, EPI_ISL_1169192, EPI_ISL_1169193, EPI_ISL_1169194, EPI_ISL_1169195, EPI_ISL_1169196, EPI_ISL_1169197, EPI_ISL_1169198, EPI_ISL_1169199, EPI_ISL_1169200, EPI_ISL_1169201, EPI_ISL_1169202, EPI_ISL_1169203, EPI_ISL_1169204, EPI_ISL_1169205, EPI_ISL_1169206, EPI_ISL_1169207, EPI_ISL_1169208, EPI_ISL_1169209, EPI_ISL_1169210, EPI_ISL_1169211, EPI_ISL_1169212, EPI_ISL_1169213, EPI_ISL_1169214, EPI_ISL_1169215, EPI_ISL_1169216, EPI_ISL_1169217, EPI_ISL_1169218, EPI_ISL_1169219, EPI_ISL_1169220, EPI_ISL_1169221, EPI_ISL_1169222, EPI_ISL_1169223, EPI_ISL_1169224, EPI_ISL_1169225, EPI_ISL_1169226, EPI_ISL_1169227, EPI_ISL_1169228, EPI_ISL_1169229, EPI_ISL_1169230, EPI_ISL_1169231, EPI_ISL_1169232, EPI_ISL_1169233, EPI_ISL_1169234, EPI_ISL_1169235, EPI_ISL_1169236, EPI_ISL_1169237, EPI_ISL_1169238, EPI_ISL_1169239, EPI_ISL_1169240, EPI_ISL_1169241, EPI_ISL_1169242, EPI_ISL_1169243, EPI_ISL_1169244, EPI_ISL_1169245, EPI_ISL_1169246, EPI_ISL_1169247, EPI_ISL_1169248, EPI_ISL_1169249, EPI_ISL_1169250, EPI_ISL_1169251, EPI_ISL_1169252, EPI_ISL_1169253, EPI_ISL_1169254, EPI_ISL_1169255, EPI_ISL_1169256, EPI_ISL_1169257, EPI_ISL_1169258, EPI_ISL_1169259, EPI_ISL_1169260, EPI_ISL_1169261, EPI_ISL_1169262, EPI_ISL_1169263, EPI_ISL_1169264, EPI_ISL_1169265, EPI_ISL_1169266, EPI_ISL_1169267, EPI_ISL_1169268, EPI_ISL_1169269, EPI_ISL_1169270, EPI_ISL_1169271, EPI_ISL_1169272, EPI_ISL_1169273, EPI_ISL_1169274, EPI_ISL_1169275, EPI_ISL_1169276, EPI_ISL_1169277, EPI_ISL_1169278, EPI_ISL_1169279, EPI_ISL_1169280, EPI_ISL_1169281, EPI_ISL_1169282, EPI_ISL_1169283, EPI_ISL_1169284, EPI_ISL_1169285, EPI_ISL_1169286, EPI_ISL_1169287, EPI_ISL_1169288, EPI_ISL_1169289, EPI_ISL_1169290, EPI_ISL_1169291, EPI_ISL_1169292, EPI_ISL_1169293, EPI_ISL_1169294, EPI_ISL_1169295, EPI_ISL_1169296, EPI_ISL_1169297, EPI_ISL_1169298, EPI_ISL_1169299, EPI_ISL_1169300, EPI_ISL_1169301, EPI_ISL_1169302, EPI_ISL_1169303, EPI_ISL_1169304, EPI_ISL_1169305, EPI_ISL_1169306, EPI_ISL_1169307, EPI_ISL_1169308, EPI_ISL_1169309, EPI_ISL_1169310, EPI_ISL_1169311, EPI_ISL_1169312, EPI_ISL_1169313, EPI_ISL_1169314, EPI_ISL_1169315, EPI_ISL_1169316, EPI_ISL_1169317, EPI_ISL_1169318, EPI_ISL_1169319, EPI_ISL_1169320, EPI_ISL_1169321, EPI_ISL_1169322, EPI_ISL_1169323, EPI_ISL_1169324, EPI_ISL_1169325, EPI_ISL_1169326, EPI_ISL_1169327, EPI_ISL_1169328, EPI_ISL_1169329, EPI_ISL_1169330, EPI_ISL_1169331, EPI_ISL_1169332, EPI_ISL_1169333, EPI_ISL_1169334, EPI_ISL_1169335, EPI_ISL_1169336, EPI_ISL_1169337, EPI_ISL_1169338, EPI_ISL_1169339, EPI_ISL_1169340, EPI_ISL_1169341, EPI_ISL_1169342, EPI_ISL_1169343, EPI_ISL_1169344, EPI_ISL_1169345, EPI_ISL_1169346, EPI_ISL_1169347, EPI_ISL_1169348, EPI_ISL_1169349, EPI_ISL_1169350, EPI_ISL_1169351, EPI_ISL_1169352, EPI_ISL_1169353, EPI_ISL_1169354, EPI_ISL_1169355, EPI_ISL_1169356, EPI_ISL_1169357, EPI_ISL_1169358, EPI_ISL_1169359, EPI_ISL_1169360, EPI_ISL_1169361, EPI_ISL_1169362, EPI_ISL_1169363, EPI_ISL_1169364, EPI_ISL_1169365, EPI_ISL_1169366, EPI_ISL_1169367, EPI_ISL_1169368, EPI_ISL_1169369, EPI_ISL_1169370, EPI_ISL_1169371, EPI_ISL_1169372, EPI_ISL_1169373, EPI_ISL_1169374, EPI_ISL_1169375, EPI_ISL_1169376, EPI_ISL_1169377, EPI_ISL_1169378, EPI_ISL_1169379, EPI_ISL_1169380, EPI_ISL_1169381, EPI_ISL_1169382, EPI_ISL_1169383, EPI_ISL_1169384, EPI_ISL_1169385, EPI_ISL_1169386, EPI_ISL_1169387, EPI_ISL_1169388, EPI_ISL_1169389, EPI_ISL_1169390, EPI_ISL_1169391, EPI_ISL_1169392, EPI_ISL_1169393, EPI_ISL_1169394, EPI_ISL_1169395, EPI_ISL_1169396, EPI_ISL_1169397, EPI_ISL_1169398, EPI_ISL_1169399, EPI_ISL_1169400, EPI_ISL_1169401, EPI_ISL_1169402, EPI_ISL_1169403, EPI_ISL_1169404, EPI_ISL_1169405, EPI_ISL_1169406, EPI_ISL_1169407, EPI_ISL_1169408, EPI_ISL_1169409, EPI_ISL_1169410, EPI_ISL_1169411, EPI_ISL_1169412, EPI_ISL_1169413, EPI_ISL_1169414, EPI_ISL_1169415, EPI_ISL_1169416, EPI_ISL_1169417, EPI_ISL_1169418, EPI_ISL_1169419, EPI_ISL_1169420, EPI_ISL_1169421, EPI_ISL_1169422, EPI_ISL_1169423, EPI_ISL_1169424, EPI_ISL_1169425, EPI_ISL_1169426, EPI_ISL_1169427, EPI_ISL_1169428, EPI_ISL_1169429, EPI_ISL_1169430, EPI_ISL_1169431, EPI_ISL_1169432, EPI_ISL_1169433, EPI_ISL_1169434, EPI_ISL_1169435, EPI_ISL_1169436, EPI_ISL_1169437, EPI_ISL_1169438, EPI_ISL_1169439, EPI_ISL_1169440, EPI_ISL_1169441, EPI_ISL_1169442, EPI_ISL_1169443, EPI_ISL_1169444, EPI_ISL_1169445, EPI_ISL_1169446, EPI_ISL_1169447, EPI_ISL_1169448, EPI_ISL_1169449, EPI_ISL_1169450, EPI_ISL_1169451, EPI_ISL_1169452, EPI_ISL_1169453, EPI_ISL_1169454, EPI_ISL_1169455, EPI_ISL_1169456, EPI_ISL_1169457, EPI_ISL_1169458, EPI_ISL_1169459, EPI_ISL_1169460, EPI_ISL_1169461, EPI_ISL_1169462, EPI_ISL_1169463, EPI_ISL_1169464, EPI_ISL_1169465, EPI_ISL_1169466, EPI_ISL_1169467, EPI_ISL_1169468, EPI_ISL_1169469, EPI_ISL_1169470, EPI_ISL_1169471, EPI_ISL_1169472, EPI_ISL_1169473, EPI_ISL_1169474, EPI_ISL_1169475, EPI_ISL_1169476, EPI_ISL_1169477, EPI_ISL_1169478, EPI_ISL_1169479, EPI_ISL_1169480, EPI_ISL_1169481, EPI_ISL_1169482 | | | |
| see above | ASL Napoli 1 Centro | AMES Centro Polidiagnostico Strumentale S.r.l. | "Giovanni Savarese, Raffaella Ruggiero, Eloisa Evangelista, Antonella Di Carlo, Luisa Circelli, Luigi D'Amore, Nadia Petrillo, Monica Ianniello, Roberto |

| | | | |
|---|---|---|---|
| | | | Sirica,Maurizio D'Amora, Antonio Fico" |
| EPI_ISL_1169883 | INMI Lazzaro Spallanzani IRCCS | INMI Lazzaro Spallanzani IRCCS | CEM Gruber, B Bartolini, E Giombini, F Messina, M Rueca, O Butera, MR Capobianchi, A Di Caro |
| EPI_ISL_1169884 | INMI Lazzaro Spallanzani IRCCS | INMI Lazzaro Spallanzani IRCCS | F Messina, M Rueca, O Butera, CEM Gruber, B Bartolini, E Giombini, MR Capobianchi, A Di Caro |
| EPI_ISL_1169885 | INMI Lazzaro Spallanzani IRCCS | INMI Lazzaro Spallanzani IRCCS | M Rueca, O Butera, CEM Gruber, B Bartolini, E Giombini, F Messina, A Di Caro, MR Capobianchi |
| EPI_ISL_1169886 | INMI Lazzaro Spallanzani IRCCS | INMI Lazzaro Spallanzani IRCCS | B Bartolini, E Giombini, F Messina, M Rueca, O Butera, CEM Gruber, A Di Caro, MR Capobianchi |
| EPI_ISL_1169887 | San Gallicano Dermatological Institute I.F.O. | INMI Lazzaro Spallanzani IRCCS | E Giombini, F Messina, M Rueca, O Butera, CEM Gruber, B Bartolini, F Pimpinelli, F Ensoli, EG Di Domenico, A Di Caro, MR Capobianchi |
| EPI_ISL_1169888 | Ospedale "F. Spaziani" Frosinone | INMI Lazzaro Spallanzani IRCCS | E Giombini, F Messina, M Rueca, O Butera, CEM Gruber, B Bartolini, R Pulselli, C Gargiulo, C Sias, A Di Caro, MR Capobianchi |
| EPI_ISL_1169889 | San Gallicano Dermatological Institute I.F.O. | INMI Lazzaro Spallanzani IRCCS | CEM Gruber, B Bartolini, E Giombini, F Messina, M Rueca, O Butera, F Pimpinelli, F Ensoli, G D'Agosto, MR Capobianchi, A Di Caro |
| EPI_ISL_1169890 | Laboratorio Genzano - ASL RM 6 | INMI Lazzaro Spallanzani IRCCS | O Butera, CEM Gruber, B Bartolini, E Giombini, F Messina, M Rueca, G Tramini, E Conti, MR Capobianchi, A Di Caro |
| EPI_ISL_1169891 | Synlab Lazio S.r.l. | INMI Lazzaro Spallanzani IRCCS | F Messina, M Rueca, O Butera, CEM Gruber, B Bartolini, E Giombini, E Trappolini, SA Santini, MR Capobianchi, A Di Caro |
| EPI_ISL_1169892, EPI_ISL_1169893 | Università degli Studi di Perugia | Istituto Zooprofilattico Sperimentale dell'Abruzzo e Molise "G. Caporale" | Mencacci A., Camilloni B., Lorusso A, Marcacci M, Di Domenico M, Ancora M, Curini V, Mangone I, Rinaldi A, Scialabba S, Di Pasquale A, Cammà C, Puglia I, Calistri P, Savini G |
| EPI_ISL_1169894, EPI_ISL_1169895 | Istituto Zooprofilattico Sperimentale Umbria e Marche "Togo Rosati" | Istituto Zooprofilattico Sperimentale dell'Abruzzo e Molise "G. Caporale" | Biagetti M., Giammarioli M., Lorusso A, Marcacci M, Di Domenico M, Ancora M, Curini V, Mangone I, Rinaldi A, Scialabba S, Di Pasquale A, Cammà C, Puglia I, Calistri P, Savini G |
| EPI_ISL_1169905, EPI_ISL_1169906, EPI_ISL_1169907, EPI_ISL_1169908, EPI_ISL_1169909, EPI_ISL_1169910, EPI_ISL_1169911 | UOC Microbiologia e Virologia, Azienda Ospedaliera Universitaria Senese, Siena, Italy | Dipartimento di Biotecnologie Mediche | Maria Grazia Cusi, David Pinzauti, Claudia Gandolfo, Gabriele Anichini, Gianni Pozzi, Gianni Gori Savellini, Francesco Santoro |
| EPI_ISL_1171675, EPI_ISL_1171676, EPI_ISL_1171677, EPI_ISL_1171678, EPI_ISL_1171679, EPI_ISL_1171680, EPI_ISL_1171681, EPI_ISL_1171682, EPI_ISL_1171683, EPI_ISL_1171684, EPI_ISL_1171685, EPI_ISL_1171686, EPI_ISL_1171687, EPI_ISL_1171688, EPI_ISL_1171689, EPI_ISL_1171690, EPI_ISL_1171691, EPI_ISL_1171692, EPI_ISL_1171693, EPI_ISL_1171694, EPI_ISL_1171695, EPI_ISL_1171696, EPI_ISL_1171697, EPI_ISL_1171698, EPI_ISL_1171699, EPI_ISL_1171700, EPI_ISL_1171701, EPI_ISL_1171702, EPI_ISL_1171703, EPI_ISL_1171704, EPI_ISL_1171705 | | | |
| see above | UOC LAB ANALISI PO CARDARELLI CAMPOBASSO | Istituto Zooprofilattico Sperimentale dell'Abruzzo e Molise "G. Caporale" | Scutellà M, Niro G, Felice V. Lorusso A, Marcacci M, Di Domenico M, Ancora M, Curini V, Mangone I, Rinaldi A, Scialabba S, Di Pasquale A, Cammà C, Puglia I, Calistri P, Savini G |
| EPI_ISL_1173204, EPI_ISL_1173205 | Istituto Zooprofilattico Sperimentale del Mezzogiorno | Istituto Zooprofilattico Sperimentale del Mezzogiorno-U.O.C. Virologia | Maurizio Viscardi, Lorena Cardillo, Luigi Atripaldi,Giovanna Fusco, Esterina De Carlo, Antonio Limone |
| EPI_ISL_1173769, EPI_ISL_1173770, EPI_ISL_1173771, EPI_ISL_1173772, EPI_ISL_1173773, EPI_ISL_1173774, EPI_ISL_1173775, EPI_ISL_1173776, EPI_ISL_1173777, EPI_ISL_1173778, EPI_ISL_1173779, EPI_ISL_1173780, EPI_ISL_1173781, EPI_ISL_1173784, EPI_ISL_1173787, EPI_ISL_1173789, EPI_ISL_1173792, EPI_ISL_1173795, EPI_ISL_1173798, EPI_ISL_1173801, EPI_ISL_1173804, EPI_ISL_1173808, EPI_ISL_1173811, EPI_ISL_1173814, EPI_ISL_1173817, EPI_ISL_1173820, EPI_ISL_1173822, EPI_ISL_1173825, EPI_ISL_1173828, EPI_ISL_1173832, EPI_ISL_1173835, EPI_ISL_1173838, EPI_ISL_1173840, EPI_ISL_1173843, EPI_ISL_1173847, EPI_ISL_1173850, EPI_ISL_1173853, EPI_ISL_1173856, EPI_ISL_1173859, EPI_ISL_1173862, EPI_ISL_1173865, EPI_ISL_1173868, EPI_ISL_1173871, EPI_ISL_1173874, EPI_ISL_1173876, EPI_ISL_1173879, EPI_ISL_1173882, EPI_ISL_1173886, EPI_ISL_1173889, EPI_ISL_1173891, EPI_ISL_1173895, EPI_ISL_1173897, EPI_ISL_1173901, EPI_ISL_1173904, EPI_ISL_1173907, EPI_ISL_1173910, EPI_ISL_1173914, EPI_ISL_1173916, EPI_ISL_1173919, EPI_ISL_1173922, EPI_ISL_1173925, EPI_ISL_1173928, EPI_ISL_1173931, EPI_ISL_1173935, EPI_ISL_1173938, EPI_ISL_1173940, EPI_ISL_1173946, EPI_ISL_1173950, EPI_ISL_1173953, EPI_ISL_1173956, EPI_ISL_1173959, EPI_ISL_1173962 | | | |
| see above | Università Federico II - Dipartimento di scienze mediche traslazionali - Napoli | TIGEM | Antonio Grimaldi Patrizia Annunziata Francesco Panariello Michele Cennamo Valentina Bouche Chiara Colantuono Lucio Di Filippo Mariano Fiorenza Anna Manfredi Marcello Salvi Giuseppe Portella Andrea Ballabio Davide Cacchiarelli |
| EPI_ISL_1176355 | Laboratorio di Riferimento Regionale della Sicilia Occidentale per l'Emergenza COVID-19 | Laboratorio di Riferimento Regionale della Sicilia Occidentale per l'Emergenza COVID-19 | Fabio Tramuto, Carmelo Massimo Maida, Daniela Di Naro, Giulia Randazzo, Walter Mazzucco, Giorgio Graziano, Vincenzo Restivo, Claudio Costantino, Francesco Vitale |
| EPI_ISL_1178582 | Department of Infectious Diseases, Istituto Superiore di Sanità, Rome, Italy; Università degli Studi di Perugia, Perugia, Italy | Istituto Superiore di Sanità (ISS) | Paola Stefanelli, Alessandra Lo Presti, Angela Di Martino, Stefano Fiore, Antonella Mencacci, Barbara Camilloni, Manuela Marra, Maria Carollo, Marco Crescenzi |
| EPI_ISL_1180141, EPI_ISL_1180142, EPI_ISL_1180143, EPI_ISL_1180144 | Laboratorio Biologia Molecolare Sars Cov2 - UOC Laboratorio Analisi - Servizio Medicina di Laboratorio, Ospedale "San Francesco" - ATS-ASSL Nuoro | Laboratorio specialistico UOC Ematologia - Ospedale "San Francesco" - ATS-ASSL Nuoro | Giovanna Piras, Rosanna Asproni, Paolo Malune, Maria Itria Monne, Maura Fiamma, Angelo Domenico Palmas, Iana Lo Maglio, Giuseppe Mameli |
| EPI_ISL_1181344 | Department of Infectious Diseases, Istituto Superiore di Sanità, Rome, Italy; AOR San Carlo, Potenza, Italy | Istituto Superiore di Sanità (ISS) | Paola Stefanelli, Angela Di Martino, Alessandra Lo Presti, Stefano Fiore, Antonio Picerno, Lopizzo Teresa, Anna Curci, Angela Menchise, Manuela Marra, Maria Carollo, Marco Crescenzi |
| EPI_ISL_1181347 | Department of Infectious Diseases, Istituto Superiore di Sanità, Rome, Italy; U.O.S.D. Laboratorio Analisi Distretto 4 Ovest AULSS 9 Scaligera, Italy | Istituto Superiore di Sanità (ISS) | Paola Stefanelli, Angela Di Martino, Alessandra Lo Presti, Stefano Fiore, Maria Pompilia Visconti, Rosa Sforza, Manuela Marra, Maria Carollo, Marco Crescenzi |
| EPI_ISL_1181627 | Department of Infectious Diseases, Istituto Superiore di Sanità, Rome, Italy; Azienda Usl Toscana sud est, Arezzo, Italy | Istituto Superiore di Sanità (ISS) | Paola Stefanelli, Angela Di Martino, Alessandra Lo Presti, Stefano Fiore, Agostino Ognibene, Manuela Marra, Maria Carollo, Marco Crescenzi |
| EPI_ISL_1181933, EPI_ISL_1181934, EPI_ISL_1181935, EPI_ISL_1181936, EPI_ISL_1181937, EPI_ISL_1181938, EPI_ISL_1181939, EPI_ISL_1181940, EPI_ISL_1181941, EPI_ISL_1181942, EPI_ISL_1181943, EPI_ISL_1181944, EPI_ISL_1181945 | | | |
| see above | Azienda Ospedaliero - Universitaria di Modena Policlinico - Virologia e Microbiologia Molecolare | Istituto Zooprofilattico Sperimentale della Lombardia e dell'Emilia Romagna (IZSLER), Risk Analysis and Genomic Epidemiology Unit | Monica Pecorari, William Gennari, Giulia Fregni Serpini, Marina Morganti, Ilaria Menozzi, Erika Scaltriti, Stefano Pongolini |
| EPI_ISL_410545 | INMI Lazzaro Spallanzani IRCCS | Laboratory of Virology, INMI Lazzaro Spallanzani IRCCS | Maria R. Capobianchi, Cesare E. M. Gruber, Martina Rueca, Barbara Bartolini, Francesco Messina, Emanuela Giombini, Francesca Colavita, Concetta Castilletti, Eleonora Lalle, Fabrizio Carletti, Emanuele Nicastri, Giuseppe Ippolito. |
| EPI_ISL_410546 | INMI Lazzaro Spallanzani IRCCS | Laboratory of Virology, INMI Lazzaro Spallanzani IRCCS | Maria R. Capobianchi, Cesare E. M. Gruber, Martina Rueca, Fabrizio Carletti, Barbara Bartolini, Francesco Messina, Emanuela Giombini, Francesca Colavita, Concetta Castilletti, Eleonora Lalle, Emanuele Nicastri, Giuseppe Ippolito. |
| EPI_ISL_412973 | Department of Infectious Diseases, Istituto Superiore di Sanità, Roma , Italy | Virology Laboratory, Scientific Department, Army Medical Center | Paola Stefanelli, Stefano Fiore, Antonella Marchi, Eleonora Benedetti, Concetta Fabiani, Giovanni Faggioni, Antonella Fortunato, Riccardo De Santis, Silvia Fillo, Anna Anselmo, Andrea Ciammaruconi, Stefano Palomba, Florigio Lista |
| EPI_ISL_412974 | Department of Infectious Diseases, Istituto Superiore di Sanità, Rome, Italy | Virology Laboratory, Scientific Department, Army Medical Center | Paola Stefanelli, Stefano Fiore, Antonella Marchi, Eleonora Benedetti, Concetta Fabiani, Giovanni Faggioni, Antonella Fortunato, Silvia Fillo, Riccardo De Santis, Andrea Ciammaruconi, Giancarlo Petralito, Filippo Molinari, Florigio Lista |
| EPI_ISL_413489 | Laboratorio di Microbiologia e Virologia, Università Vita-Salute San Raffaele, Milano | Laboratorio di Microbiologia e Virologia, Università Vita-Salute San Raffaele, Milano | R.A Diotti, E. Criscuolo, M. Castelli, V. Caputo, R. Ferrarese, M. Sampaolo, E. Boeri, I. Negri, V. Amato, G. Lo Raso, C. Di Resta, R. Burioni, M. Clementi, N. Mancini & N. Clementi |
| EPI_ISL_417418 | Laboratory of Molecular Virology International Center fro Genetic Engineering and Biotechnology (ICGEB) | ARGO Open Lab Platform for Genome sequencing | Licastro D, Rajasekharan S, Dal Monego S, Segat L, D'Agaro P, Marcello A |
| EPI_ISL_417419, EPI_ISL_417421 | Laboratory of Molecular Virology International Center for Genetic Engineering and Biotechnology (ICGEB) | ARGO Open Lab Platform for Genome sequencing | Licastro D, Rajasekharan S, Dal Monego S, Segat L, D'Agaro P, Marcello A |
| EPI_ISL_417423 | Laboratory of Molecular Virology International Center for Genetic Engineering and Biotechnology (ICGEB) | ARGO Open Lab Platform for Genome sequencing | Licastro D, Rajasekharan, Dal Monego S, Segat L, D'Agaro P, Marcello A |
| EPI_ISL_417445, EPI_ISL_417446, EPI_ISL_417447 | Laboratory of Infectious Diseases, Department of Biomedical and Clinical Sciences L. Sacco, University of Milan | Laboratory of Infectious Diseases, Department of Biomedical and Clinical Sciences L. Sacco, University of Milan | Gianguglielmo Zehender, Alessia Lai, Annalisa Bergna, Luca Meroni, Agostino Riva, Claudia Balotta, Maciej Tarkowski, Arianna Gabrieli, Dario Bernacchia, Stefano Rusconi, Giuliano Rizzardini, Spinello Antinori, Massimo Galli |
| EPI_ISL_417491 | Virology Laboratory, Department of Biomedical Sciences and Public Health, University Politecnica delle Marche | Virology and Legal Medicine Laboratories, Department of Biomedical Sciences and Public Health, University Politecnica delle Marche | Bagnarelli,P., Caucci,S., Di Sante,L., Menzo,S., Alessandrini,F., Onofri,V., Turchi,C., Tagliabracci,A. |

| | | | |
|---|---|---|---|
| EPI_ISL_417921 | INMI Lazzaro Spallanzani IRCCS | Laboratory of Virology, INMI Lazzaro Spallanzani IRCCS | Martina Rueca, Barbara Bartolini, Francesco Messina, Cesare E. M. Gruber, Emanuela Giombini, Maria R. Capobianchi, Fabrizio Carletti, Francesca Colavita, Concetta Castilletti, Eleonora Lalle, Daniele Lapa, Giuseppe Ippolito. |
| EPI_ISL_417922 | INMI Lazzaro Spallanzani IRCCS | Laboratory of Virology, INMI Lazzaro Spallanzani IRCCS | Cesare E. M. Gruber, Martina Rueca, Barbara Bartolini, Francesco Messina, Emanuela Giombini, Maria R. Capobianchi, Fabrizio Carletti, Francesca Colavita, Concetta Castilletti, Eleonora Lalle, Daniele Lapa, Giuseppe Ippolito. |
| EPI_ISL_417923 | INMI Lazzaro Spallanzani IRCCS | Laboratory of Virology, INMI Lazzaro Spallanzani IRCCS | Francesco Messina, Barbara Bartolini, Martina Rueca, Cesare E. M. Gruber, Emanuela Giombini, Maria R. Capobianchi, Fabrizio Carletti, Francesca Colavita, Concetta Castilletti, Eleonora Lalle, Daniele Lapa, Giuseppe Ippolito. |
| EPI_ISL_418255 | Presidio Ospedaliero "S. Spirito" - PESCARA | Istituto Zooprofilattico Sperimentale dell'Abruzzo e Molise "G. Caporale" | Lorusso A, Marcacci M, Cammà C, Monaco F, Puglia I, Di Pasquale A, Rinaldi A, Mangone I, Savini G |
| EPI_ISL_418256 | Ospedale "San Liberatore" di Atri | Istituto Zooprofilattico Sperimentale dell'Abruzzo e Molise "G. Caporale" | Lorusso A, Marcacci M, Di Domenico M, Puglia I, Curini V, Ancora M, Di Pasquale A, Rinaldi A, Mangone I, Cammà C, Savini G. |
| EPI_ISL_418257 | Ospedale Civile Giuseppe Mazzini, Teramo | Istituto Zooprofilattico Sperimentale dell'Abruzzo e Molise "G.Caporale" | Lorusso A, Marcacci M, Di Domenico M, Puglia I, Curini V, Ancora M, Di Pasquale A, Rinaldi A, Mangone I, Cammà C, Savini G. |
| EPI_ISL_418258, EPI_ISL_418259 | Presidio ospedaliero "Santo Spirito" | Istituto Zooprofilattico Sperimentale dell'Abruzzo e Molise "G. Caporale" | Lorusso A, Marcacci M, Di Domenico M, Puglia I, Curini V, Ancora M, Di Pasquale A, Rinaldi A, Mangone I, Cammà C, Savini G. |
| EPI_ISL_418260, EPI_ISL_418261 | Ospedale Civile Giuseppe Mazzini | Istituto Zooprofilattico Sperimentale dell'Abruzzo e Molise "G. Caporale" | Lorusso A, Marcacci M, Di Domenico M, Puglia I, Curini V, Ancora M, Di Pasquale A, Rinaldi A, Mangone I, Cammà C, Savini G. |
| EPI_ISL_419254 | INMI Lazzaro Spallanzani IRCCS | Laboratory of Virology, INMI Lazzaro Spallanzani IRCCS | Barbara Bartolini, Martina Rueca, Francesco Messina, Cesare E. M. Gruber, Emanuela Giombini, Maria R. Capobianchi, Fabrizio Carletti, Francesca Colavita, Concetta Castilletti, Eleonora Lalle, Daniele Lapa, Giuseppe Ippolito. |
| EPI_ISL_419255 | INMI Lazzaro Spallanzani IRCCS | INMI Lazzaro Spallanzani IRCCS | Antonino Di Caro, Cesare E. M. Gruber, Martina Rueca, Barbara Bartolini, Francesco Messina, Emanuela Giombini, Maria R. Capobianchi, Fabrizio Carletti, Francesca Colavita, Concetta Castilletti, Eleonora Lalle, Daniele Lapa, Giuseppe Ippolito. |
| EPI_ISL_420563 | Ospedale Civile Giuseppe Mazzini | Istituto Zooprofilattico Sperimentale dell'Abruzzo e Molise "G. Caporale" | Lorusso A, Marcacci M, Di Domenico M, Ancora M, Curini V, Mangone I, Rinaldi A, Di Pasquale A, Cammà C, Puglia I, Savini G |
| EPI_ISL_420564 | Ospedale Civile Castel Di Sangro | Istituto Zooprofilattico Sperimentale dell'Abruzzo e Molise "G.Caporale" | Lorusso A, Marcacci M, Di Domenico M, Ancora M, Curini V, Mangone I, Rinaldi A, Di Pasquale A, Cammà C, Puglia I, Savini G |
| EPI_ISL_420565 | Ospedale Civile Giuseppe Mazzini | Istituto Zooprofilattico Sperimentale dell'Abruzzo e Molise "G.Caporale" | Lorusso A, Marcacci M, Di Domenico M, Ancora M, Curini V, Mangone I, Rinaldi A, Di Pasquale A, Cammà C, Puglia I, Savini G |
| EPI_ISL_420566, EPI_ISL_420567 | Ospedale Regionale San Salvatore | Istituto Zooprofilattico Sperimentale dell'Abruzzo e Molise "G.Caporale" | Lorusso A, Marcacci M, Di Domenico M, Ancora M, Curini V, Mangone I, Rinaldi A, Di Pasquale A, Cammà C, Puglia I, Savini G |
| EPI_ISL_420568, EPI_ISL_420569, EPI_ISL_420583, EPI_ISL_420592 | Ospedale Civile Giuseppe Mazzini | Istituto Zooprofilattico Sperimentale dell'Abruzzo e Molise "G.Caporale" | Lorusso A, Marcacci M, Di Domenico M, Ancora M, Curini V, Mangone I, Rinaldi A, Di Pasquale A, Cammà C, Puglia I, Savini G |
| EPI_ISL_422437, EPI_ISL_422438 | ULSS9 Distretto di Bussolengo | Istituto Zooprofilattico Sperimentale delle Venezie | Adelaide Milani, Alessia Schivo, Annalisa Salviato, Erika Giorgia Quaranta, Ambra Pastori, Bianca Zecchin, Alice Fusaro, Isabella Monne, Calogero Terregino, Antonia Ricci |
| EPI_ISL_424342 | INMI Lazzaro Spallanzani IRCCS | Laboratory of Virology, INMI Lazzaro Spallanzani IRCCS | Concetta Castilletti, Barbara Bartolini, Martina Rueca, Cesare Ernesto Maria Gruber, Francesco Messina, Fabrizio Carletti, Eleonora Lalle, Licia Bordi, Giulia Matusali, Francesca Colavita, Maria Rosaria Capobianchi, Francesco Vairo, Giuseppe Ippolito, Antonino Di Caro |
| EPI_ISL_424343 | INMI Lazzaro Spallanzani IRCCS | Laboratory of Virology, INMI Lazzaro Spallanzani IRCCS | Fabrizio Carletti, Barbara Bartolini, Martina Rueca, Cesare Ernesto Maria Gruber, Francesco Messina, Eleonora Lalle, Licia Bordi, Giulia Matusali, Francesca Colavita, Maria Rosaria Capobianchi, Concetta Castilletti, Francesco Vairo, Giuseppe Ippolito, Antonino Di Caro |
| EPI_ISL_424344 | INMI Lazzaro Spallanzani IRCCS | Laboratory of Virology, INMI Lazzaro Spallanzani IRCCS | Eleonora Lalle, Barbara Bartolini, Martina Rueca, Cesare Ernesto Maria Gruber, Francesco Messina, Fabrizio Carletti, Licia Bordi, Giulia Matusali, Francesca Colavita, Maria Rosaria Capobianchi, Concetta Castilletti, Francesco Vairo, Giuseppe Ippolito, Antonino Di Caro |
| EPI_ISL_428853 | Laboratory of Molecular Virology International Center for Genetic Engineering and Biotechnology (ICGEB) | ARGO Open Lab Platform for Genome Sequencing | Licastro D, Rajasekharan S, Dal Monego S, Segat L, D'Agaro P, Marcello A |
| EPI_ISL_428854 | Laboratory of Molecular Virology International Center for Genetic Engineering and Biotechnology (ICGEB) | ARGO Open Lab Platform for Genome sequencing | Licastro D, Rajasekharan S, Dal Monego S, Segat L, D'Agaro P, Marcello A |
| EPI_ISL_429226, EPI_ISL_429227 | Presidio Ospedaliero Santo Spirito | Istituto Zooprofilattico Sperimentale dell'Abruzzo e Molise "G. Caporale" | Lorusso A, Marcacci M, Di Domenico M, Ancora M, Curini V, Mangone I, Rinaldi A, Di Pasquale A, Camma C, Puglia I, Savini G |
| EPI_ISL_429228 | Ospedale Civile Giuseppe Mazzini | Istituto Zooprofilattico Sperimentale dell'Abruzzo e Molise "G. Caporale" | Lorusso A, Marcacci M, Di Domenico M, Ancora M, Curini V, Mangone I, Rinaldi A, Di Pasquale A, Camma C, Puglia I, Savini G |
| EPI_ISL_429229 | Ospedale Regionale San Salvatore | Istituto Zooprofilattico Sperimentale dell'Abruzzo e Molise "G. Caporale" | Lorusso A, Marcacci M, Di Domenico M, Ancora M, Curini V, Mangone I, Rinaldi A, Di Pasquale A, Camma C, Puglia I, Savini G |
| EPI_ISL_429230, EPI_ISL_429231, EPI_ISL_429232, EPI_ISL_429233, EPI_ISL_429234, EPI_ISL_429235 | Ospedale Civile Giuseppe Mazzini | Istituto Zooprofilattico Sperimentale dell'Abruzzo e Molise "G. Caporale" | Lorusso A, Marcacci M, Di Domenico M, Ancora M, Curini V, Mangone I, Rinaldi A, Di Pasquale A, Camma C, Puglia I, Savini G |
| EPI_ISL_429236 | Ospedale Civile S. Liberatore di Atri | Istituto Zooprofilattico Sperimentale dell'Abruzzo e Molise "G. Caporale" | Lorusso A, Marcacci M, Di Domenico M, Ancora M, Curini V, Mangone I, Rinaldi A, Di Pasquale A, Camma C, Puglia I, Savini G |
| EPI_ISL_435145 | Ospedale Civile Giuseppe Mazzini | Istituto Zooprofilattico Sperimentale dell'Abruzzo e Molise "G.Caporale" | Lorusso A, Marcacci M, Di Domenico M, Ancora M, Curini V, Mangone I, Rinaldi A, Di Pasquale A, Cammà C, Puglia I, Savini G |
| EPI_ISL_435146, EPI_ISL_435147 | Villa Serena del Dr. Leonardo Petruzzi | Istituto Zooprofilattico Sperimentale dell'Abruzzo e Molise "G.Caporale" | Lorusso A, Marcacci M, Di Domenico M, Ancora M, Curini V, Mangone I, Rinaldi A, Di Pasquale A, Cammà C, Puglia I, Savini G |
| EPI_ISL_435148 | Ospedale SS Annunziata | Istituto Zooprofilattico Sperimentale dell'Abruzzo e Molise "G.Caporale" | Lorusso A, Marcacci M, Di Domenico M, Ancora M, Curini V, Mangone I, Rinaldi A, Di Pasquale A, Cammà C, Puglia I, Savini G |
| EPI_ISL_435149 | SERVIZIO DI IGIENE E SANITÀ PUBBLICA ASL Teramo | Istituto Zooprofilattico Sperimentale dell'Abruzzo e Molise "G.Caporale" | Lorusso A, Marcacci M, Di Domenico M, Ancora M, Curini V, Mangone I, Rinaldi A, Di Pasquale A, Cammà C, Puglia I, Savini G |
| EPI_ISL_435150, EPI_ISL_435151 | Ospedale SS Annunziata | Istituto Zooprofilattico Sperimentale dell'Abruzzo e Molise "G.Caporale" | Lorusso A, Marcacci M, Di Domenico M, Ancora M, Curini V, Mangone I, Rinaldi A, Di Pasquale A, Cammà C, Puglia I, Savini G |
| EPI_ISL_435152 | Servizio di Igiene, Epidemiologia e Sanità Pubblica (SIESP) Avezzano | Istituto Zooprofilattico Sperimentale dell'Abruzzo e Molise "G.Caporale" | Lorusso A, Marcacci M, Di Domenico M, Ancora M, Curini V, Mangone I, Rinaldi A, Di Pasquale A, Cammà C, Puglia I, Savini G |
| EPI_ISL_435153, EPI_ISL_435154, EPI_ISL_435155 | SERVIZIO DI IGIENE E SANITÀ PUBBLICA ASL Teramo | Istituto Zooprofilattico Sperimentale dell'Abruzzo e Molise "G.Caporale" | Lorusso A, Marcacci M, Di Domenico M, Ancora M, Curini V, Mangone I, Rinaldi A, Di Pasquale A, Cammà C, Puglia I, Savini G |
| EPI_ISL_436718 | Ospedale Regionale San Salvatore | Istituto Zooprofilattico Sperimentale dell'Abruzzo e Molise "G.Caporale" | Lorusso A, Marcacci M, Di Domenico M, Ancora M, Curini V, Mangone I, Rinaldi A, Di Pasquale A, Cammà C, Puglia I, Savini G |
| EPI_ISL_436719, EPI_ISL_436720, EPI_ISL_436721, EPI_ISL_436722 | Ospedale Civile S. Liberatore di Atri | Istituto Zooprofilattico Sperimentale dell'Abruzzo e Molise "G.Caporale" | Lorusso A, Marcacci M, Di Domenico M, Ancora M, Curini V, Mangone I, Rinaldi A, Di Pasquale A, Cammà C, Puglia I, Savini G |

| | | | |
|---|---|---|---|
| EPI_ISL_436723 | Ospedale Civile Giuseppe Mazzini | Istituto Zooprofilattico Sperimentale dell'Abruzzo e Molise "G.Caporale" | Lorusso A, Marcacci M, Di Domenico M, Ancora M, Curini V, Mangone I, Rinaldi A, Di Pasquale A, Cammà C, Puglia I, Savini G |
| EPI_ISL_436724 | Ospedale Civile S. Liberatore di Atri | Istituto Zooprofilattico Sperimentale dell'Abruzzo e Molise "G.Caporale" | Lorusso A, Marcacci M, Di Domenico M, Ancora M, Curini V, Mangone I, Rinaldi A, Di Pasquale A, Cammà C, Puglia I, Savini G |
| EPI_ISL_436725 | RSA/RP Villa San Giovanni - Gruppo Edos | Istituto Zooprofilattico Sperimentale dell'Abruzzo e Molise "G.Caporale" | Lorusso A, Marcacci M, Di Domenico M, Ancora M, Curini V, Mangone I, Rinaldi A, Di Pasquale A, Cammà C, Puglia I, Savini G |
| EPI_ISL_436726, EPI_ISL_436727, EPI_ISL_436728, EPI_ISL_436729 | SERVIZIO DI IGIENE E SANITÀ PUBBLICA ASL Teramo | Istituto Zooprofilattico Sperimentale dell'Abruzzo e Molise "G.Caporale" | Lorusso A, Marcacci M, Di Domenico M, Ancora M, Curini V, Mangone I, Rinaldi A, Di Pasquale A, Cammà C, Puglia I, Savini G |
| EPI_ISL_436730 | Servizio di igiene epidemiologia e sanità pubblica (Siesp) Chieti | Istituto Zooprofilattico Sperimentale dell'Abruzzo e Molise "G.Caporale" | Lorusso A, Marcacci M, Di Domenico M, Ancora M, Curini V, Mangone I, Rinaldi A, Di Pasquale A, Cammà C, Puglia I, Savini G |
| EPI_ISL_436731, EPI_ISL_436732 | Ospedale Civile S. Liberatore di Atri | Istituto Zooprofilattico Sperimentale dell'Abruzzo e Molise "G.Caporale" | Lorusso A, Marcacci M, Di Domenico M, Ancora M, Curini V, Mangone I, Rinaldi A, Di Pasquale A, Cammà C, Puglia I, Savini G |
| EPI_ISL_451298 | Laboratory of Virology, INMI Lazzaro Spallanzani IRCCS | Laboratory of Virology, INMI Lazzaro Spallanzani IRCCS | Cesare E.M. Gruber, Martina Rueca, Barbara Bartolini, Francesco Messina, Antonino Di Caro, Maria R. Capobianchi, Giuseppe Ippolito |
| EPI_ISL_451299 | Laboratory of Virology, INMI Lazzaro Spallanzani IRCCS | Laboratory of Virology, INMI Lazzaro Spallanzani IRCCS | Martina Rueca, Cesare E.M. Gruber, Barbara Bartolini, Francesco Messina, Antonino Di Caro, Maria R. Capobianchi, Giuseppe Ippolito |
| EPI_ISL_451300 | Laboratory of Virology, INMI Lazzaro Spallanzani IRCCS | Laboratory of Virology, INMI Lazzaro Spallanzani IRCCS | Cesare E.M. Gruber, Martina Rueca, Barbara Bartolini, Francesco Messina, Antonino Di Caro, Maria R. Capobianchi, Giuseppe Ippolito |
| EPI_ISL_451301 | Laboratory of Virology, INMI Lazzaro Spallanzani IRCCS | Laboratory of Virology, INMI Lazzaro Spallanzani IRCCS | Martina Rueca, Cesare E.M. Gruber, Barbara Bartolini, Francesco Messina, Antonino Di Caro, Maria R. Capobianchi, Giuseppe Ippolito |
| EPI_ISL_451302 | Laboratory of Virology, INMI Lazzaro Spallanzani IRCCS | Laboratory of Virology, INMI Lazzaro Spallanzani IRCCS | Cesare E.M. Gruber, Martina Rueca, Barbara Bartolini, Francesco Messina, Antonino Di Caro, Maria R. Capobianchi, Giuseppe Ippolito |
| EPI_ISL_451303 | Laboratory of Virology, INMI Lazzaro Spallanzani IRCCS | Laboratory of Virology, INMI Lazzaro Spallanzani IRCCS | Martina Rueca, Cesare E.M. Gruber, Barbara Bartolini, Francesco Messina, Antonino Di Caro, Maria R. Capobianchi, Giuseppe Ippolito |
| EPI_ISL_451304 | Laboratory of Virology, INMI Lazzaro Spallanzani IRCCS | Laboratory of Virology, INMI Lazzaro Spallanzani IRCCS | Cesare E.M. Gruber, Martina Rueca, Barbara Bartolini, Francesco Messina, Antonino Di Caro, Maria R. Capobianchi, Giuseppe Ippolito |
| EPI_ISL_451305 | Laboratory of Virology, INMI Lazzaro Spallanzani IRCCS | Laboratory of Virology, INMI Lazzaro Spallanzani IRCCS | Martina Rueca, Cesare E.M. Gruber, Barbara Bartolini, Francesco Messina, Antonino Di Caro, Maria R. Capobianchi, Giuseppe Ippolito |
| EPI_ISL_451306 | Molecular Virology Unit, Fondazione IRCCS Policlinico San Matteo , Pavia | Laboratory of Virology, INMI Lazzaro Spallanzani IRCCS | Antonio Piralla, Fausto Baldanti, Martina Rueca, Antonino Di Caro, Maria R. Capobianchi, Cesare E.M. Gruber, Barbara Bartolini |
| EPI_ISL_451307 | Molecular Virology Unit, Fondazione IRCCS Policlinico San Matteo , Pavia | Laboratory of Virology, INMI Lazzaro Spallanzani IRCCS | Fausto Baldanti, Antonio Piralla, Antonino Di Caro, Cesare E.M. Gruber, Martina Rueca, Barbara Bartolini, Maria R. Capobianchi |
| EPI_ISL_451308 | Molecular Virology Unit, Fondazione IRCCS Policlinico San Matteo , Pavia | Laboratory of Virology, INMI Lazzaro Spallanzani IRCCS | Antonio Piralla, Fausto Baldanti, Maria R. Capobianchi, Cesare E.M. Gruber, Martina Rueca, Barbara Bartolini, Antonino Di Caro |
| EPI_ISL_451309 | Molecular Virology Unit, Fondazione IRCCS Policlinico San Matteo , Pavia | Laboratory of Virology, INMI Lazzaro Spallanzani IRCCS | Fausto Baldanti, Antonio Piralla, Cesare E.M. Gruber, Maria R. Capobianchi, Antonino Di Caro, Martina Rueca, Barbara Bartolini |
| EPI_ISL_451961 | Istituto Zooprofilattico Sperimentale Puglia e Basilicata; Dipartimento di Bioscienze, Biotecnologie e Biofarmaceutica dell'Università degli Studi di Bari "A.Moro"; Istituto di Biomembrane. Bioenergetica e Biotecnologie Molecolari del Consiglio Nazionale delle Ricerche di Bari | Beaconlab (Bioinformatics Evolution and Comparative Genomics lab), Dept of Biosciences, University of Milan | Parisi A.,Pesole G., Manzari C., Chiara M. |
| EPI_ISL_451962 | Istituto Zooprofilattico Sperimentale Puglia e Basilicata; Dipartimento di Bioscienze, Biotecnologie e Biofarmaceutica dell'Università degli Studi di Bari "A.Moro"; Istituto di Biomembrane. Bioenergetica e Biotecnologie Molecolari del Consiglio Nazionale delle Ricerche di Bari | Beaconlab (Bioinformatics, Evolution and Comparative Genomics lab), Dept of Biosciences, University on Milan | Parisi A.,Pesole G., Manzari C., Chiara M. |
| EPI_ISL_452181, EPI_ISL_452182, EPI_ISL_452183, EPI_ISL_452184, EPI_ISL_452185, EPI_ISL_452186, EPI_ISL_452187, EPI_ISL_452188, EPI_ISL_452189 | ULSS9 Distretto di Bussolengo | Istituto Zooprofilattico Sperimentale delle Venezie | Adelaide Milani, Alessia Schivo, Annalisa Salviato, Erika Giorgia Quaranta, Ambra Pastori, Bianca Zecchin, Alice Fusaro, Isabella Monne, Calogero Terregino, Antonia Ricci |
| EPI_ISL_452190, EPI_ISL_452191 | ULSS9 Distretto di San Bonifacio | Istituto Zooprofilattico Sperimentale delle Venezie | Adelaide Milani, Alessia Schivo, Annalisa Salviato, Erika Giorgia Quaranta, Ambra Pastori, Bianca Zecchin, Alice Fusaro, Isabella Monne, Calogero Terregino, Antonia Ricci |
| EPI_ISL_454733 | Department of Medical, Biotechnologies University of Siena | Department of Medical, Biotechnologies University of Siena | Cusi,M.G., Pinzauti,D., Gandolfo,C., Anichini,G., Pozzi,G. and Santoro,F. |
| EPI_ISL_457699, EPI_ISL_457700 | Department of Infectious Diseases, Istituto Superiore di Sanità, Roma , Italy | Army Medical and Veterinary Research Center | Paola Stefanelli, Alessandra Lo Presti, Stefano Fiore, Antonella Marchi, Eleonora Benedetti, Concetta Fabiani Silvia Fillo, Giovanni Faggioni, Riccardo De Sanctis, Antonella Fortunato, Anna Anselmo, Francesco Giordani, Vanessa Vera Fain, Nino D'Amore, Florigio Lista |
| EPI_ISL_457721, EPI_ISL_457724, EPI_ISL_457728, EPI_ISL_457732, EPI_ISL_457736, EPI_ISL_457749 | Department of Infectious Diseases, Istituto Superiore di Sanità, Roma , Italy | Army Medical and Veterinary Research Center | Paola Stefanelli, Alessandra Lo Presti, Stefano Fiore, Antonella Marchi, Eleonora Benedetti, Concetta Fabiani Silvia Fillo, Giovanni Faggioni, Riccardo De Sanctis, Antonella Fortunato, Anna Anselmo, Francesco Giordani, Vanessa Vera Fain, Nino D'Amore, Florigio Lista |
| EPI_ISL_457825 | Army Medical Research Center - Scientific Department | Army Medical and Veterinary Research Center | Silvia Fillo, Giovanni Faggioni, Riccardo De Sanctis, Antonella Fortunato, Anna Anselmo, Francesco Giordani, Vanessa Vera Fain, Nino D'Amore, Florigio Lista |
| EPI_ISL_457826 | Army Medical Center - Scientific Department | Army Medical and Veterinary Research Center | Silvia Fillo, Giovanni Faggioni, Riccardo De Sanctis, Antonella Fortunato, Anna Anselmo, Francesco Giordani, Vanessa Vera Fain, Nino D'Amore, Florigio Lista |
| EPI_ISL_458084 | Laboratorio Biologia Molecolare Sars Cov2 - UOC Laboratorio Analisi - Servizio Medicina di Laboratorio, Ospedale "San Francesco" - ATS-ASSL Nuoro | Laboratorio specialistico UOC Ematologia - Ospedale "San Francesco" - ATS-ASSL Nuoro | Piras Giovanna, Fancello Tatiana, Asproni Rosanna, Fiamma Maura, Monne Maria Itria, Toja Alessandro, Sanna Filomena, Floris Anna Rita, Sulis Vincenzo, Palmas Angelo Domenico, Casu Gavino, Lo Maglio Iana, Mameli Giuseppe. |
| EPI_ISL_458085 | Laboratorio Biologia Molecolare Sars Cov2 - UOC Laboratorio Analisi - Servizio Medicina di Laboratorio , Ospedale "San Francesco" - ATS- ASSL Nuoro | Laboratorio specialistico UOC Ematologia - Ospedale "San Francesco" - ATS-ASSL Nuoro | Piras Giovanna, Fancello Tatiana, Asproni Rosanna, Fiamma Maura, Monne Maria Itria, Toja Alessandro, Sanna Filomena, Floris Anna Rita, Sulis Vincenzo, Palmas Angelo Domenico, Casu Gavino, Lo Maglio Iana, Mameli Giuseppe. |
| EPI_ISL_460079 | Molecular Virology Unit, Fondazione IRCCS Policlinico San Matteo , Pavia | Laboratory of Virology, INMI Lazzaro Spallanzani IRCCS | Barbara Bartolini, Cesare E.M. Gruber, Maria R. Capobianchi, Martina Rueca, Antonio Piralla, Fausto Baldanti, Antonino Di Caro |
| EPI_ISL_460080 | Molecular Virology Unit, Fondazione IRCCS Policlinico San Matteo , Pavia | Laboratory of Virology, INMI Lazzaro Spallanzani IRCCS | Antonio Piralla, Barbara Bartolini, Fausto Baldanti, Martina Rueca, Antonino Di Caro, Cesare E.M. Gruber, Maria R. Capobianchi |
| EPI_ISL_460081 | Molecular Virology Unit, Fondazione IRCCS Policlinico San Matteo , Pavia | Laboratory of Virology, INMI Lazzaro Spallanzani IRCCS | Fausto Baldanti, Martina Rueca, Antonio Piralla, Antonino Di Caro, Maria R. Capobianchi, Cesare E.M. Gruber, Barbara Bartolini |
| EPI_ISL_460082 | Molecular Virology Unit, Fondazione IRCCS Policlinico San Matteo , Pavia | Laboratory of Virology, INMI Lazzaro Spallanzani IRCCS | Martina Rueca, Cesare E.M. Gruber, Antonio Piralla, Antonino Di Caro, Barbara Bartolini, Maria R. Capobianchi, Fausto Baldanti |
| EPI_ISL_460083 | Molecular Virology Unit, Fondazione IRCCS Policlinico San Matteo , Pavia | Laboratory of Virology, INMI Lazzaro Spallanzani IRCCS | Martina Rueca, Antonino Di Caro, Cesare E.M. Gruber, Barbara Bartolini, Fausto Baldanti, Antonio Piralla, Maria R. Capobianchi |

| | | | |
|---|---|---|---|
| EPI_ISL_460084 | Molecular Virology Unit, Fondazione IRCCS Policlinico San Matteo , Pavia | Laboratory of Virology, INMI Lazzaro Spallanzani IRCCS | Fausto Baldanti, Antonio Piralla, Martina Rueca, Barbara Bartolini, Maria R. Capobianchi, Cesare E.M. Gruber, Antonino Di Caro |
| EPI_ISL_460085 | Molecular Virology Unit, Fondazione IRCCS Policlinico San Matteo , Pavia | Laboratory of Virology, INMI Lazzaro Spallanzani IRCCS | Cesare E.M. Gruber, Maria R. Capobianchi, Barbara Bartolini, Fausto Baldanti, Martina Rueca, Antonio Piralla, Antonino Di Caro |
| EPI_ISL_460086 | Molecular Virology Unit, Fondazione IRCCS Policlinico San Matteo , Pavia | Laboratory of Virology, INMI Lazzaro Spallanzani IRCCS | Maria R. Capobianchi, Fausto Baldanti, Antonio Piralla, Antonino Di Caro, Barbara Bartolini, Cesare E.M. Gruber, Martina Rueca |
| EPI_ISL_460087 | Molecular Virology Unit, Fondazione IRCCS Policlinico San Matteo , Pavia | Laboratory of Virology, INMI Lazzaro Spallanzani IRCCS | Cesare E.M. Gruber, Maria R. Capobianchi, Martina Rueca, Barbara Bartolini, Antonino Di Caro, Antonio Piralla, Fausto Baldanti |
| EPI_ISL_460088 | Molecular Virology Unit, Fondazione IRCCS Policlinico San Matteo , Pavia | Laboratory of Virology, INMI Lazzaro Spallanzani IRCCS | Martina Rueca, Barbara Bartolini, Fausto Baldanti, Maria R. Capobianchi, Cesare E.M. Gruber, Antonino Di Caro, Antonio Piralla |
| EPI_ISL_460089 | Molecular Virology Unit, Fondazione IRCCS Policlinico San Matteo , Pavia | Laboratory of Virology, INMI Lazzaro Spallanzani IRCCS | Antonino Di Caro, Barbara Bartolini, Martina Rueca, Cesare E.M. Gruber, Antonio Piralla, Fausto Baldanti, Maria R. Capobianchi |
| EPI_ISL_460090 | Molecular Virology Unit, Fondazione IRCCS Policlinico San Matteo , Pavia | Laboratory of Virology, INMI Lazzaro Spallanzani IRCCS | Antonio Piralla, Cesare E.M. Gruber, Antonino Di Caro, Maria R. Capobianchi, Martina Rueca, Barbara Bartolini, Fausto Baldanti |
| EPI_ISL_460091 | Molecular Virology Unit, Fondazione IRCCS Policlinico San Matteo , Pavia | Laboratory of Virology, INMI Lazzaro Spallanzani IRCCS | Antonino Di Caro, Antonio Piralla, Martina Rueca, Fausto Baldanti, Barbara Bartolini, Maria R. Capobianchi, Cesare E.M. Gruber |
| EPI_ISL_460092 | Molecular Virology Unit, Fondazione IRCCS Policlinico San Matteo , Pavia | Laboratory of Virology, INMI Lazzaro Spallanzani IRCCS | Cesare E.M. Gruber, Martina Rueca, Maria R. Capobianchi, Antonino Di Caro, Antonio Piralla, Barbara Bartolini, Fausto Baldanti |
| EPI_ISL_460093 | Molecular Virology Unit, Fondazione IRCCS Policlinico San Matteo , Pavia | Laboratory of Virology, INMI Lazzaro Spallanzani IRCCS | Maria R. Capobianchi, Antonio Piralla, Antonino Di Caro, Fausto Baldanti, Martina Rueca, Cesare E.M. Gruber, Barbara Bartolini |
| EPI_ISL_460094 | Molecular Virology Unit, Fondazione IRCCS Policlinico San Matteo , Pavia | Laboratory of Virology, INMI Lazzaro Spallanzani IRCCS | Barbara Bartolini, Maria R. Capobianchi, Antonino Di Caro, Antonio Piralla, Cesare E.M. Gruber, Martina Rueca, Fausto Baldanti |
| EPI_ISL_460095 | Molecular Virology Unit, Fondazione IRCCS Policlinico San Matteo , Pavia | Laboratory of Virology, INMI Lazzaro Spallanzani IRCCS | Barbara Bartolini, Antonino Di Caro, Fausto Baldanti, Cesare E.M. Gruber, Maria R. Capobianchi, Martina Rueca, Antonio Piralla |
| EPI_ISL_468914, EPI_ISL_469016, EPI_ISL_469018, EPI_ISL_469019, EPI_ISL_469020, EPI_ISL_469021, EPI_ISL_469022 | Istituto Zooprofilattico Sperimentale Puglia e Basilicata; Dipartimento di Bioscienze, Biotecnologie e Biofarmaceutica dell'Università degli Studi di Bari "A.Moro"; Istituto di Biomembrane. Bioenergetica e Biotecnologie Molecolari del Consiglio Nazionale delle Ricerche di Bari | Beaconlab (Bioinformatics, Evolution and Comparative Genomics lab), Dept of Biosciences, University on Milan | Parisi A.,Pesole G., Manzari C., Chiara M. |
| EPI_ISL_469023 | Istituto Zooprofilattico Sperimentale Puglia e Basilicata; Dipartimento di Bioscienze, Biotecnologie e Biofarmaceutica dell'Università degli Studi di Bari "A.Moro"; Istituto di Biomembrane. Bioenergetica e Biotecnologie Molecolari del Consiglio Nazionale delle Ricerche di Bari | Beaconlab (Bioinformatics, Evolution and Comparative Genomics lab), Dept of Biosciences, University on Milan | Parisi A.,Pesole G., Manzari C., Chiara M |
| EPI_ISL_477193, EPI_ISL_477194 | Istituto Zooprofilattico Sperimentale Puglia e Basilicata; | Beaconlab (Bioinformatics, Evolution and Comparative Genomics lab), Dept of Biosciences, University on Mila | Parisi A.,Pesole G., Manzari C., Chiara M. |
| EPI_ISL_477195, EPI_ISL_477196, EPI_ISL_477197, EPI_ISL_477198, EPI_ISL_477199, EPI_ISL_477200, EPI_ISL_477201 | Istituto Zooprofilattico Sperimentale Puglia e Basilicata; | Beaconlab (Bioinformatics, Evolution and Comparative Genomics lab), Dept of Biosciences, University on Milan | Parisi A.,Pesole G., Manzari C., Chiara M. |
| EPI_ISL_477202, EPI_ISL_477203 | Istituto Zooprofilattico Sperimentale Puglia e Basilicata; | Beaconlab (Bioinformatics, Evolution and Comparative Genomics lab), Dept of Biosciences, University on Mila | Parisi A.,Pesole G., Manzari C., Chiara M. |
| EPI_ISL_477204 | Prof. Massimo Zollo CEINGE TASK-FORCE COVID19 - Regione Campania | Prof. Massimo Zollo CEINGE TASK-FORCE COVID19 - Regione Campania | Veronica Ferrucci1,2, Dae young Kong8, Fatemeh asadzadeh1,2, Laura Marrone1,2, Roberto Siciliano1,2, Rino Cerino3, Giovanna Fusco3, Marika Comegna1,2, Angelo Boccia2, Maurizio Viscardi3, Giorgia Borriello3, Sergio Brandi3, Claudia Tiberio4, Luigi Atripaldi4, Giovanni Paolella1,2, Giuseppe Castaldo1,2, Stefano Pascarella4, Martina Bianchi4, Lorenzo Chiariotti1,2, Jae Myun Lee5, Jae Ho Jung6, Kyong Seop Yun7, Hong Yeoul Kim 7,8* and Massimo Zollo1,2* 1 CEINGE Biotecnologie Avanzate, Naples, Italia 2 Dipartimento di Medicina Molecolare e Biotecnologie Mediche DMMBM University of Naples Federico II, Italia 3 Istituto Zooprofilattico Sperimentale del Mezzogiorno, Naples, Italia 4 -U.O.C. di Patologia Clinica Ospedale D. Cotugno, Azienda Sanitaria Ospedali dei Colli, Naples, Italy. 5 Università La Sapienza di Roma, Italia 6 Department of Microbiology, Yonsei University College of Medicine, Seoul, Korea 7 Department of Surgery, Yonsei University College of Medicine, Seoul, Korea 8 Haim bio co., Ltd, , Indust |
| EPI_ISL_479616, EPI_ISL_479617 | Laboratory of Molecular Virology of the International Centre for Genetic Engineering and Biotechnology (ICGEB) | ARGO Open Lab Platform for Genome Sequencing | Licastro, D, Rajasekharan S, Dal Monego S, Segat L, D'Agaro P, Salton F, Confalonieri P, Confalonieri M Marcello A |
| EPI_ISL_479618, EPI_ISL_479619, EPI_ISL_479790, EPI_ISL_479791 | Laboratory of Molecular Virology of the International Centre for Genetic Engineering and Biotechnology (ICGEB) | ARGO Open Lab Platform for Genome Sequencing | Licastro, D, Rajasekharan S, Dal Monego S, Segat L, D'Agaro P, Salton F, Confalonieri P, Confalonieri M, Marcello A |
| EPI_ISL_486646, EPI_ISL_486647 | Microbiology, Virology and Biemergency Laboratory-ASST FBF Sacco | Microbiology, Virology and Biemergency Laboratory-ASST FBF Sacco | Mancon A, Comandatore F, Romeri F, Micheli V, Rimoldi SG |
| EPI_ISL_486648 | Microbiology, Virology and Biemergency Laboratory-ASST FBF Sacco | Microbiology, Virology and Biemergency Laboratory-ASST FBF Sacco | Micheli V, Comandatore F, Romeri F, Mancon A, Rimoldi SG |
| EPI_ISL_486649 | Microbiology, Virology and Biemergency Laboratory-ASST FBF Sacco | Microbiology, Virology and Biemergency Laboratory-ASST FBF Sacco | Rimoldi SG, Comandatore F, Romeri F, Mancon A, Micheli V |
| EPI_ISL_486650 | Microbiology, Virology and Biemergency Laboratory-ASST FBF Sacco | Microbiology, Virology and Biemergency Laboratory-ASST FBF Sacco | Romeri F, Comandatore F, Mancon A, Micheli V, Rimoldi SG |
| EPI_ISL_486651 | Microbiology, Virology and Biemergency Laboratory-ASST FBF Sacco | Microbiology, Virology and Biemergency Laboratory-ASST FBF Sacco | Mancon A, Comandatore F, Romeri F, Micheli V, Rimoldi SG |
| EPI_ISL_486652 | Microbiology, Virology and Biemergency Laboratory-ASST FBF Sacco | Microbiology, Virology and Biemergency Laboratory-ASST FBF Sacco | Micheli V, Comandatore F, Romeri F, Mancon A, Rimoldi SG |
| EPI_ISL_486653 | Microbiology, Virology and Biemergency Laboratory-ASST FBF Sacco | Microbiology, Virology and Biemergency Laboratory-ASST FBF Sacco | Rimoldi SG, Comandatore F, Romeri F, Mancon A, Micheli V |
| EPI_ISL_486654 | Microbiology, Virology and Biemergency Laboratory-ASST FBF Sacco | Microbiology, Virology and Biemergency Laboratory-ASST FBF Sacco | Romeri F, Comandatore F, Mancon A, Micheli V, Rimoldi SG |
| EPI_ISL_486655 | Microbiology, Virology and Biemergency Laboratory-ASST FBF Sacco | Microbiology, Virology and Biemergency Laboratory-ASST FBF Sacco | Mancon A, Comandatore F, Romeri F, Micheli V, Rimoldi SG |
| EPI_ISL_486656 | Microbiology, Virology and Biemergency Laboratory-ASST FBF Sacco | Microbiology, Virology and Biemergency Laboratory-ASST FBF Sacco | Micheli V, Comandatore F, Romeri F, Mancon A, Rimoldi SG |

| | | | |
|---|---|---|---|
| EPI_ISL_486657 | Microbiology, Virology and Biemergency Laboratory-ASST FBF Sacco | Microbiology, Virology and Biemergency Laboratory-ASST FBF Sacco | Rimoldi SG, Comandatore F, Romeri F, Mancon A, Micheli V |
| EPI_ISL_486658 | Microbiology, Virology and Biemergency Laboratory-ASST FBF Sacco | Microbiology, Virology and Biemergency Laboratory-ASST FBF Sacco | Romeri F, Comandatore F, Mancon A, Micheli V, Rimoldi SG |
| EPI_ISL_486659 | Microbiology, Virology and Biemergency Laboratory-ASST FBF Sacco | Microbiology, Virology and Biemergency Laboratory-ASST FBF Sacco | Micheli V, Comandatore F, Romeri F, Mancon A, Rimoldi SG |
| EPI_ISL_486660 | Microbiology, Virology and Biemergency Laboratory-ASST FBF Sacco | Microbiology, Virology and Biemergency Laboratory-ASST FBF Sacco | Rimoldi SG, Comandatore F, Romeri F, Mancon A, Micheli V |
| EPI_ISL_486661 | Microbiology, Virology and Biemergency Laboratory-ASST FBF Sacco | Microbiology, Virology and Biemergency Laboratory-ASST FBF Sacco | Romeri F, Comandatore F, Mancon A, Micheli V, Rimoldi SG |
| EPI_ISL_486662 | Microbiology, Virology and Biemergency Laboratory-ASST FBF Sacco | Microbiology, Virology and Biemergency Laboratory-ASST FBF Sacco | Mancon A, Comandatore F, Romeri F, Micheli V, Rimoldi SG |
| EPI_ISL_486663 | Microbiology, Virology and Biemergency Laboratory-ASST FBF Sacco | Microbiology, Virology and Biemergency Laboratory-ASST FBF Sacco | Micheli V, Comandatore F, Romeri F, Mancon A, Rimoldi SG |
| EPI_ISL_486664 | Microbiology, Virology and Biemergency Laboratory-ASST FBF Sacco | Microbiology, Virology and Biemergency Laboratory-ASST FBF Sacco | Rimoldi SG, Comandatore F, Romeri F, Mancon A, Micheli V |
| EPI_ISL_486665 | Microbiology, Virology and Biemergency Laboratory-ASST FBF Sacco | Microbiology, Virology and Biemergency Laboratory-ASST FBF Sacco | Micheli V, Rimoldi SG, Comandatore F, Mancon A, Romeri F |
| EPI_ISL_487276 | Department of Food Safety, Nutrition and Veterinary public health, Istituto Superiore di Sanita' | Department of Biomedical, Surgical and Dental Sciences and Department of Biomedical Sciences for Health | Delbue,S., Ferrante,P., Basilico,N., Parapini,S., Binda,S., D'Alessandro,S., Galli,C., Signorini,L., Primache,V., Anselmi,G., Pariani,E. |
| EPI_ISL_492980, EPI_ISL_492981, EPI_ISL_492982, EPI_ISL_492983, EPI_ISL_492984, EPI_ISL_492985, EPI_ISL_492986, EPI_ISL_492987 | IRCCS Sacro Cuore Don Calabria Hospital, Department of Infectious, Tropical Diseases & Microbiology | University of Verona, Department of Biotechnology | Antonio Mori, Michela Deiana, Elena Pomari, Chiara Piubelli; Giulia Lopatriello, Luca Marcolungo, Cristina Beltrami, Chiara Degli Esposti, Emanuela Cosentino, Massimo Delledonne |
| EPI_ISL_493328 | INMI Lazzaro Spallanzani IRCCS | INMI Lazzaro Spallanzani IRCCS | Martina Rueca, Cesare E.M. Gruber, Barbara Bartolini, Francesco Messina, Maria R. Capobianchi, Antonino Di Caro |
| EPI_ISL_493329 | INMI Lazzaro Spallanzani IRCCS | INMI Lazzaro Spallanzani IRCCS | Barbara Bartolini, Martina Rueca, Cesare E.M. Gruber, Francesco Messina, Antonino Di Caro, Maria R. Capobianchi |
| EPI_ISL_493330 | INMI Lazzaro Spallanzani IRCCS | INMI Lazzaro Spallanzani IRCCS | Cesare E.M. Gruber, Martina Rueca, Barbara Bartolini, Francesco Messina, Maria R. Capobianchi, Antonino Di Caro |
| EPI_ISL_493331 | INMI Lazzaro Spallanzani IRCCS | INMI Lazzaro Spallanzani IRCCS | Martina Rueca, Cesare E.M. Gruber, Barbara Bartolini, Francesco Messina, Maria R. Capobianchi, Antonino Di Caro |
| EPI_ISL_493332 | Istituto Zooprofilattico Sperimentale del Mezzogiorno | INMI Lazzaro Spallanzani IRCCS | Cesare E.M. Gruber, Martina Rueca, Barbara Bartolini, Francesco Messina, Antonino Di Caro, Giovanna Fusco, Maurizio Viscardi, Giorgia Borriello, Maria R. Capobianchi |
| EPI_ISL_493333 | Istituto Zooprofilattico Sperimentale del Mezzogiorno | INMI Lazzaro Spallanzani IRCCS | Barbara Bartolini, Martina Rueca, Cesare E.M. Gruber, Francesco Messina, Antonino Di Caro, Giovanna Fusco, Maurizio Viscardi, Giorgia Borriello, Maria R. Capobianchi |
| EPI_ISL_496482 | Dept. Infectious, Tropical Diseases & Microbiology, IRCCS Sacro Cuore Don Calabria Hospital | 1) Dept. Infectious, Tropical Diseases & Microbiology, IRCCS Sacro Cuore Don Calabria Hospital; 2) Centro Piattaforme Tecnologiche, University of Verona; 3) Dept. Neurosciences, Biomedicine and Movement Sciences, University of Verona. | 1) Antonio Mori, Michela Deiana, Elena Pomari, Chiara Piubelli; 2) Monica Castellucci and Francesca Griggio; 3) Giovanni Malerba |
| EPI_ISL_498559, EPI_ISL_498560, EPI_ISL_498561, EPI_ISL_498562, EPI_ISL_498563 | Laboratory of Molecular Virology International Center for Genetic Engineering and Biotechnology (ICGEB) | ARGO Open Lab Platform for Genome Sequencing | Licastro D, Rajasekharan S, Dal Monego S, Segat L, D'Agaro P, Marcello A |
| EPI_ISL_514432 | Prof. Massimo Zollo CEINGE TASK-FORCE COVID19 - Regione Campania | Prof. Massimo Zollo CEINGE TASK-FORCE COVID19 - Regione Campania | Veronica Ferrucci, Dae young Kong, Fatemeh asadzadeh, Laura Marrone, Roberto Siciliano, Rino Cerino, Giovanna Fusco, Marika Comegna, Angelo Boccia, Maurizio Viscardi, Giorgia Borriello, Sergio Brandi, Claudia Tiberio, Luigi Atripaldi, Giovanni Paolella, Giuseppe Castaldo, Stefano Pascarella, Martina Bianchi, Lorenzo Chiariotti, Jae Myun Lee, Jae Ho Jung, Kyong Seop Yun, Hong Yeoul Kim and Massimo Zollo |
| EPI_ISL_514751 | CoronaNet Lab- TaskForce Regione Campania, CEINGE Biotecnologie Avanzate, Via G. Salvatore | CoronaNet Lab- TaskForce Regione Campania, CEINGE Biotecnologie Avanzate, Via G. Salvatore | Zollo,M., Ferrucci,V., Kong,Dy., Asadzadeh,F., Marrone,L.,Siciliano,R., Cerino,R., Fusco,G., Comegna,M., Boccia,A.,Viscardi,M., Borriello,G., Brandi,S., Tiberio,C., Atripaldi,L.,Paolella,G., Castaldo,G., Pascarella,S., Bianchi,M., Chiariotti,L.,Lee,J.M., Jung,J.H., Yun,K.S. and Kim,H.Y. |
| EPI_ISL_516079, EPI_ISL_516080, EPI_ISL_516081, EPI_ISL_516082, EPI_ISL_516083, EPI_ISL_516084, EPI_ISL_516085, EPI_ISL_516086, EPI_ISL_516087, EPI_ISL_516088 | Biomedical Sciences and Public Health, Polytechnic University of Marche | Biomedical Sciences and Public Health, Polytechnic University of Marche | Bagnarelli,P., Caucci,S., Di Sante,L., Menzo,S., Alessandrini,F., Onofri,V., Turchi,C., Melchionda,F., Tagliabracci,A. |
| EPI_ISL_522855 | ULSS9 Distretto di Bussolengo | Istituto Zooprofilattico Sperimentale delle Venezie | Adelaide Milani, Alessia Schivo, Annalisa Salviato, Erika Giorgia Quaranta, Ambra Pastori, Bianca Zecchin, Alice Fusaro, Isabella Monne, Calogero Terregino, Antonia Ricci |
| EPI_ISL_522856 | ULSS9 Distretto di San Bonifacio | Istituto Zooprofilattico Sperimentale delle Venezie | Adelaide Milani, Alessia Schivo, Annalisa Salviato, Erika Giorgia Quaranta, Ambra Pastori, Bianca Zecchin, Alice Fusaro, Isabella Monne, Calogero Terregino, Antonia Ricci |
| EPI_ISL_522857 | ULSS9 Scaligera | Istituto Zooprofilattico Sperimentale delle Venezie | Adelaide Milani, Alessia Schivo, Annalisa Salviato, Erika Giorgia Quaranta, Ambra Pastori, Bianca Zecchin, Alice Fusaro, Isabella Monne, Calogero Terregino, Antonia Ricci |
| EPI_ISL_522858 | ULSS9 Distretto di San Bonifacio | Istituto Zooprofilattico Sperimentale delle Venezie | Adelaide Milani, Alessia Schivo, Annalisa Salviato, Erika Giorgia Quaranta, Ambra Pastori, Bianca Zecchin, Alice Fusaro, Isabella Monne, Calogero Terregino, Antonia Ricci |
| EPI_ISL_522859 | ULSS9 Scaligera | Istituto Zooprofilattico Sperimentale delle Venezie | Adelaide Milani, Alessia Schivo, Annalisa Salviato, Erika Giorgia Quaranta, Ambra Pastori, Bianca Zecchin, Alice Fusaro, Isabella Monne, Calogero Terregino, Antonia Ricci |
| EPI_ISL_522860, EPI_ISL_522861, EPI_ISL_522862, EPI_ISL_522863, EPI_ISL_522864, EPI_ISL_522865, EPI_ISL_522866, EPI_ISL_522867, EPI_ISL_522868 | ULSS9 Distretto di Bussolengo | Istituto Zooprofilattico Sperimentale delle Venezie | Adelaide Milani, Alessia Schivo, Annalisa Salviato, Erika Giorgia Quaranta, Ambra Pastori, Bianca Zecchin, Alice Fusaro, Isabella Monne, Calogero Terregino, Antonia Ricci |
| EPI_ISL_525495, EPI_ISL_525496 | Laboratory of Molecular Virology of the International Centre for Genetic Engineering and Biotechnology (ICGEB) | ARGO Open Lab Platform for Genome Sequencing | Licastro D, Rajasekharan S, Dal Monego S, Segat L, D'Agaro P, Marcello A |
| EPI_ISL_525553, EPI_ISL_525554, EPI_ISL_525555, EPI_ISL_525556, EPI_ISL_525557, EPI_ISL_525558, EPI_ISL_525559, EPI_ISL_525560, EPI_ISL_525561, EPI_ISL_525562, EPI_ISL_525563, EPI_ISL_525564, EPI_ISL_525565, EPI_ISL_525566, EPI_ISL_525567, EPI_ISL_525568, EPI_ISL_525569, EPI_ISL_525570, EPI_ISL_525571, EPI_ISL_525572, EPI_ISL_525573, EPI_ISL_525574, EPI_ISL_527380 | | | |
| see above | Istituto Zooprofilattico Sperimentale Puglia e Basilicata; Dipartimento di Bioscienze, Biotecnologie e Biofarmaceutica | Beaconlab (Bioinformatics, Evolution and Comparative Genomics lab), Dept of Biosciences, University on Milan | Parisi A.,Pesole G., Manzari C., Chiara M |

| | | | |
|---|---|---|---|
| | dell'Università degli Studi di Bari "A.Moro"; Istituto di Biomembrane. Bioenergetica e Biotecnologie Molecolari del Consiglio Nazionale delle Ricerche di Bari | | |
| EPI_ISL_528919 | Ospedale Civile S. Liberatore-Atri | Istitituto Zooprofilattico Sperimentale dell'Abruzzo e Molise "G.Caporale" | Lorusso A, Marcacci M, Di Domenico M, Curini V, Ancora M, Cammà C, Rinaldi A, Mangone I, Di Pasquale A, Puglia I, Savini G. |
| EPI_ISL_528920, EPI_ISL_528921 | Presidio Ospedaliero "Santo Spirito"-Pescara | Istitituto Zooprofilattico Sperimentale dell'Abruzzo e Molise "G.Caporale" | Lorusso A, Marcacci M, Di Domenico M, Curini V, Ancora M, Cammà C, Rinaldi A, Mangone I, Di Pasquale A, Puglia I, Savini G. |
| EPI_ISL_528922 | Ospedale "Giuseppe Mazzini"-Teramo | Istitituto Zooprofilattico Sperimentale dell'Abruzzo e Molise "G.Caporale" | Lorusso A, Marcacci M, Di Domenico M, Curini V, Ancora M, Cammà C, Rinaldi A, Mangone I, Di Pasquale A, Puglia I, Savini G. |
| EPI_ISL_528923 | Ospedale Civile S. Liberatore-Atri | Istitituto Zooprofilattico Sperimentale dell'Abruzzo e Molise "G.Caporale" | Lorusso A, Marcacci M, Di Domenico M, Curini V, Ancora M, Cammà C, Rinaldi A, Mangone I, Di Pasquale A, Puglia I, Savini G. |
| EPI_ISL_528924 | Ospedale "Giuseppe Mazzini"-Teramo | Istitituto Zooprofilattico Sperimentale dell'Abruzzo e Molise "G.Caporale" | Lorusso A, Marcacci M, Di Domenico M, Curini V, Ancora M, Cammà C, Rinaldi A, Mangone I, Di Pasquale A, Puglia I, Savini G. |
| EPI_ISL_528925 | Ospedale Regionale San Salvatore-L'Aquila | Istitituto Zooprofilattico Sperimentale dell'Abruzzo e Molise "G.Caporale" | Lorusso A, Marcacci M, Di Domenico M, Curini V, Ancora M, Cammà C, Rinaldi A, Mangone I, Di Pasquale A, Puglia I, Savini G. |
| EPI_ISL_528926, EPI_ISL_528927, EPI_ISL_528928 | Ospedale "Giuseppe Mazzini"-Teramo | Istitituto Zooprofilattico Sperimentale dell'Abruzzo e Molise "G.Caporale" | Lorusso A, Marcacci M, Di Domenico M, Curini V, Ancora M, Cammà C, Rinaldi A, Mangone I, Di Pasquale A, Puglia I, Savini G. |
| EPI_ISL_528929 | Ospedale Civile S. Liberatore-Atri | Istitituto Zooprofilattico Sperimentale dell'Abruzzo e Molise "G.Caporale" | Lorusso A, Marcacci M, Di Domenico M, Curini V, Ancora M, Cammà C, Rinaldi A, Mangone I, Di Pasquale A, Puglia I, Savini G. |
| EPI_ISL_528934, EPI_ISL_528935, EPI_ISL_528936, EPI_ISL_528937, EPI_ISL_528938, EPI_ISL_528939, EPI_ISL_528940, EPI_ISL_528941, EPI_ISL_528942, EPI_ISL_528943, EPI_ISL_528944, EPI_ISL_528945, EPI_ISL_528946, EPI_ISL_528947, EPI_ISL_528948, EPI_ISL_528949 | | | |
| see above | Agenzia di Tutela della Salute di Bergamo | Istitituto Zooprofilattico Sperimentale dell'Abruzzo e Molise "G.Caporale" | Lorusso A, Marcacci M, Di Domenico M, Curini V, Ancora M, Cammà C, Rinaldi A, Mangone I, Di Pasquale A, Puglia I, Savini G. |
| EPI_ISL_528990 | Ospedale Civile Maria SS. dello Splendore | Istitituto Zooprofilattico Sperimentale dell'Abruzzo e Molise "G.Caporale" | Lorusso A, Marcacci M, Di Domenico M, Curini V, Ancora M, Cammà C, Rinaldi A, Mangone I, Di Pasquale A, Puglia I, Savini G. |
| EPI_ISL_528991, EPI_ISL_528992 | Ospedale SS Annunziata-Sulmona | Istitituto Zooprofilattico Sperimentale dell'Abruzzo e Molise "G.Caporale" | Lorusso A, Marcacci M, Di Domenico M, Curini V, Ancora M, Cammà C, Rinaldi A, Mangone I, Di Pasquale A, Puglia I, Savini G. |
| EPI_ISL_528993 | Ospedale Civile S. Liberatore-Atri | Istitituto Zooprofilattico Sperimentale dell'Abruzzo e Molise "G.Caporale" | Lorusso A, Marcacci M, Di Domenico M, Curini V, Ancora M, Cammà C, Rinaldi A, Mangone I, Di Pasquale A, Puglia I, Savini G. |
| EPI_ISL_528994, EPI_ISL_528995, EPI_ISL_528996, EPI_ISL_528997, EPI_ISL_528998, EPI_ISL_528999, EPI_ISL_529000, EPI_ISL_529001, EPI_ISL_529002, EPI_ISL_529003, EPI_ISL_529004, EPI_ISL_529005 | | | |
| see above | Servizio di igiene epidemiologia e sanità pubblica (SIESP)-Chieti | Istitituto Zooprofilattico Sperimentale dell'Abruzzo e Molise "G.Caporale" | Lorusso A, Marcacci M, Di Domenico M, Curini V, Ancora M, Cammà C, Rinaldi A, Mangone I, Di Pasquale A, Puglia I, Savini G. |
| EPI_ISL_529006 | Servizio di igiene e sanità pubblica (SIESP)-Teramo | Istitituto Zooprofilattico Sperimentale dell'Abruzzo e Molise "G.Caporale" | Lorusso A, Marcacci M, Di Domenico M, Curini V, Ancora M, Cammà C, Rinaldi A, Mangone I, Di Pasquale A, Puglia I, Savini G. |
| EPI_ISL_529007 | Ospedale Civile S. Liberatore-Atri | Istitituto Zooprofilattico Sperimentale dell'Abruzzo e Molise "G.Caporale" | Lorusso A, Marcacci M, Di Domenico M, Curini V, Ancora M, Cammà C, Rinaldi A, Mangone I, Di Pasquale A, Puglia I, Savini G. |
| EPI_ISL_529008 | Servizio di igiene e sanità pubblica (SIESP)-Teramo | Istitituto Zooprofilattico Sperimentale dell'Abruzzo e Molise "G.Caporale" | Lorusso A, Marcacci M, Di Domenico M, Curini V, Ancora M, Cammà C, Rinaldi A, Mangone I, Di Pasquale A, Puglia I, Savini G. |
| EPI_ISL_529009 | Ospedale Civile S. Liberatore-Atri | Istitituto Zooprofilattico Sperimentale dell'Abruzzo e Molise "G.Caporale" | Lorusso A, Marcacci M, Di Domenico M, Curini V, Ancora M, Cammà C, Rinaldi A, Mangone I, Di Pasquale A, Puglia I, Savini G. |
| EPI_ISL_529010, EPI_ISL_529011, EPI_ISL_529012 | Servizio Igiene Epidemiologia e Sanità Pubblica (SIESP)-L'Aquila | Istitituto Zooprofilattico Sperimentale dell'Abruzzo e Molise "G.Caporale" | Lorusso A, Marcacci M, Di Domenico M, Curini V, Ancora M, Cammà C, Rinaldi A, Mangone I, Di Pasquale A, Puglia I, Savini G. |
| EPI_ISL_529013 | Presidio Ospedaliero "S.Filippo e Nicola"-Avezzano | Istitituto Zooprofilattico Sperimentale dell'Abruzzo e Molise "G.Caporale" | Lorusso A, Marcacci M, Di Domenico M, Curini V, Ancora M, Cammà C, Rinaldi A, Mangone I, Di Pasquale A, Puglia I, Savini G. |
| EPI_ISL_529014, EPI_ISL_529015 | Ospedale "Ss. Annunziata" | Istitituto Zooprofilattico Sperimentale dell'Abruzzo e Molise "G.Caporale" | Lorusso A, Marcacci M, Di Domenico M, Curini V, Ancora M, Cammà C, Rinaldi A, Mangone I, Di Pasquale A, Puglia I, Savini G. |
| EPI_ISL_529016 | Ospedale SS Annunziata-Sulmona | Istitituto Zooprofilattico Sperimentale dell'Abruzzo e Molise "G.Caporale" | Lorusso A, Marcacci M, Di Domenico M, Curini V, Ancora M, Cammà C, Rinaldi A, Mangone I, Di Pasquale A, Puglia I, Savini G. |
| EPI_ISL_529017 | RSA San Raffaele Sulmona | Istitituto Zooprofilattico Sperimentale dell'Abruzzo e Molise "G.Caporale" | Lorusso A, Marcacci M, Di Domenico M, Curini V, Ancora M, Cammà C, Rinaldi A, Mangone I, Di Pasquale A, Puglia I, Savini G. |
| EPI_ISL_529018 | Ospedale "Giuseppe Mazzini"-Teramo | Istitituto Zooprofilattico Sperimentale dell'Abruzzo e Molise "G.Caporale" | Lorusso A, Marcacci M, Di Domenico M, Curini V, Ancora M, Cammà C, Rinaldi A, Mangone I, Di Pasquale A, Puglia I, Savini G. |
| EPI_ISL_529019 | RSA/RP Villa San Giovanni - Gruppo Edos | Istitituto Zooprofilattico Sperimentale dell'Abruzzo e Molise "G.Caporale" | Lorusso A, Marcacci M, Di Domenico M, Curini V, Ancora M, Cammà C, Rinaldi A, Mangone I, Di Pasquale A, Puglia I, Savini G. |
| EPI_ISL_529020, EPI_ISL_529021 | Ospedale Civile S. Liberatore-Atri | Istitituto Zooprofilattico Sperimentale dell'Abruzzo e Molise "G.Caporale" | Lorusso A, Marcacci M, Di Domenico M, Curini V, Ancora M, Cammà C, Rinaldi A, Mangone I, Di Pasquale A, Puglia I, Savini G. |
| EPI_ISL_529022 | Ospedale "Ss. Annunziata" | Istitituto Zooprofilattico Sperimentale dell'Abruzzo e Molise "G.Caporale" | Lorusso A, Marcacci M, Di Domenico M, Curini V, Ancora M, Cammà C, Rinaldi A, Mangone I, Di Pasquale A, Puglia I, Savini G. |
| EPI_ISL_529023, EPI_ISL_529024, EPI_ISL_529025 | Servizio di igiene epidemiologia e sanità pubblica (SIESP)-Chieti | Istitituto Zooprofilattico Sperimentale dell'Abruzzo e Molise "G.Caporale" | Lorusso A, Marcacci M, Di Domenico M, Curini V, Ancora M, Cammà C, Rinaldi A, Mangone I, Di Pasquale A, Puglia I, Savini G. |
| EPI_ISL_529026 | Ospedale "Giuseppe Mazzini"-Teramo | Istitituto Zooprofilattico Sperimentale dell'Abruzzo e Molise "G.Caporale" | Lorusso A, Marcacci M, Di Domenico M, Curini V, Ancora M, Cammà C, Rinaldi A, Mangone I, Di Pasquale A, Puglia I, Savini G. |
| EPI_ISL_542098, EPI_ISL_542099, EPI_ISL_542100, EPI_ISL_542101, EPI_ISL_542102, EPI_ISL_542103, EPI_ISL_542104, EPI_ISL_542105, EPI_ISL_542106, EPI_ISL_542107, EPI_ISL_542108, EPI_ISL_542109, EPI_ISL_542110, EPI_ISL_542111, EPI_ISL_542112, EPI_ISL_542113, EPI_ISL_542114, EPI_ISL_542115, EPI_ISL_542116, EPI_ISL_542117, EPI_ISL_542118, EPI_ISL_542119, EPI_ISL_542120, EPI_ISL_542121, EPI_ISL_542122, EPI_ISL_542123, EPI_ISL_542124, EPI_ISL_542125, EPI_ISL_542126, EPI_ISL_542127, EPI_ISL_542128, EPI_ISL_542129, EPI_ISL_542130, EPI_ISL_542131, EPI_ISL_542132, EPI_ISL_542133, EPI_ISL_542134, EPI_ISL_542135, EPI_ISL_542136, EPI_ISL_542137, EPI_ISL_542138, EPI_ISL_542139, EPI_ISL_542140, EPI_ISL_542141, EPI_ISL_542142, EPI_ISL_542143, EPI_ISL_542144, EPI_ISL_542145, EPI_ISL_542146, EPI_ISL_542147, EPI_ISL_542148, EPI_ISL_542149, EPI_ISL_542150, EPI_ISL_542151, EPI_ISL_542152, EPI_ISL_542153, EPI_ISL_542154, EPI_ISL_542155, EPI_ISL_542156, EPI_ISL_542157, EPI_ISL_542158, EPI_ISL_542159, EPI_ISL_542160, EPI_ISL_542161, EPI_ISL_542162, EPI_ISL_542163, EPI_ISL_542164, EPI_ISL_542165, EPI_ISL_542166, EPI_ISL_542167, EPI_ISL_542168, EPI_ISL_542169, EPI_ISL_542170, EPI_ISL_542171, EPI_ISL_542172, EPI_ISL_542173, EPI_ISL_542174, EPI_ISL_542175, EPI_ISL_542176, EPI_ISL_542177, EPI_ISL_542178, EPI_ISL_542179, EPI_ISL_542180, EPI_ISL_542181, EPI_ISL_542182, EPI_ISL_542183, EPI_ISL_542184, EPI_ISL_542185, EPI_ISL_542186, EPI_ISL_542187, EPI_ISL_542188, EPI_ISL_542189, EPI_ISL_542190, EPI_ISL_542191, EPI_ISL_542192, EPI_ISL_542193, EPI_ISL_542194, EPI_ISL_542195, EPI_ISL_542196, EPI_ISL_542197, EPI_ISL_542198, EPI_ISL_542199, EPI_ISL_542200, EPI_ISL_542201, EPI_ISL_542202, EPI_ISL_542203, EPI_ISL_542204, EPI_ISL_542205, EPI_ISL_542206, EPI_ISL_542207, EPI_ISL_542208, EPI_ISL_542209, EPI_ISL_542210, EPI_ISL_542211, EPI_ISL_542212, EPI_ISL_542213, EPI_ISL_542214, EPI_ISL_542215, EPI_ISL_542216, EPI_ISL_542217, EPI_ISL_542218, EPI_ISL_542219, EPI_ISL_542220, EPI_ISL_542221, EPI_ISL_542222, EPI_ISL_542223, EPI_ISL_542224, EPI_ISL_542225, EPI_ISL_542226, EPI_ISL_542227, EPI_ISL_542228, EPI_ISL_542229, EPI_ISL_542230, EPI_ISL_542231, EPI_ISL_542232, EPI_ISL_542233, EPI_ISL_542234, EPI_ISL_542235, EPI_ISL_542236, EPI_ISL_542237, EPI_ISL_542238, EPI_ISL_542239, EPI_ISL_542240, EPI_ISL_542241, EPI_ISL_542242, EPI_ISL_542243, EPI_ISL_542244, EPI_ISL_542245, EPI_ISL_542246, EPI_ISL_542247, EPI_ISL_542248, EPI_ISL_542249, EPI_ISL_542250, EPI_ISL_542251, EPI_ISL_542252, EPI_ISL_542253, EPI_ISL_542254, EPI_ISL_542255, EPI_ISL_542256, EPI_ISL_542257, EPI_ISL_542258, EPI_ISL_542259, EPI_ISL_542260, EPI_ISL_542261, EPI_ISL_542262, EPI_ISL_542263, EPI_ISL_542264, EPI_ISL_542265, EPI_ISL_542266, EPI_ISL_542267, EPI_ISL_542268, EPI_ISL_542269, EPI_ISL_542270, EPI_ISL_542271, EPI_ISL_542272, EPI_ISL_542273, EPI_ISL_542274, EPI_ISL_542275, EPI_ISL_542276, EPI_ISL_542277 | | | |
| see above | ASST GOM Niguarda | Dep. Of Oncology and Hemato-Oncology University of Milan | Claudia Alteri, Valeria Cento, Antonio Piralla, Valentino Costabile, Monica Tallarita, Luna Colagrossi, Silvia Renica, Federica Giardina, Federica Novazzi, |

| | | | |
|---|---|---|---|
| | | | Stefano Gaiarsa, Elisa Matarazzo, Maria Antonello, Chiara Vismara, Roberto Fumagalli, Oscar Massimiliano Epis, Massimo Puoti, Carlo Federico Perno, Fausto Baldanti |
| EPI_ISL_542278, EPI_ISL_542279, EPI_ISL_542280, EPI_ISL_542281, EPI_ISL_542282, EPI_ISL_542283, EPI_ISL_542284, EPI_ISL_542285, EPI_ISL_542286, EPI_ISL_542287, EPI_ISL_542288, EPI_ISL_542289, EPI_ISL_542290, EPI_ISL_542291, EPI_ISL_542292, EPI_ISL_542293, EPI_ISL_542294, EPI_ISL_542295, EPI_ISL_542296, EPI_ISL_542297, EPI_ISL_542298, EPI_ISL_542299, EPI_ISL_542300, EPI_ISL_542301, EPI_ISL_542302, EPI_ISL_542303, EPI_ISL_542304, EPI_ISL_542305, EPI_ISL_542306, EPI_ISL_542307, EPI_ISL_542308, EPI_ISL_542309, EPI_ISL_542310, EPI_ISL_542311, EPI_ISL_542312, EPI_ISL_542313, EPI_ISL_542314, EPI_ISL_542315, EPI_ISL_542316, EPI_ISL_542317, EPI_ISL_542318, EPI_ISL_542319, EPI_ISL_542320, EPI_ISL_542321, EPI_ISL_542322, EPI_ISL_542323, EPI_ISL_542324, EPI_ISL_542325, EPI_ISL_542326, EPI_ISL_542327, EPI_ISL_542328, EPI_ISL_542329, EPI_ISL_542330, EPI_ISL_542331, EPI_ISL_542332, EPI_ISL_542333, EPI_ISL_542334, EPI_ISL_542335, EPI_ISL_542336, EPI_ISL_542337, EPI_ISL_542338, EPI_ISL_542339, EPI_ISL_542340, EPI_ISL_542341, EPI_ISL_542342, EPI_ISL_542343, EPI_ISL_542344, EPI_ISL_542345, EPI_ISL_542346, EPI_ISL_542347, EPI_ISL_542348, EPI_ISL_542349, EPI_ISL_542350, EPI_ISL_542351, EPI_ISL_542352, EPI_ISL_542353, EPI_ISL_542354, EPI_ISL_542355, EPI_ISL_542356, EPI_ISL_542357, EPI_ISL_542358, EPI_ISL_542359, EPI_ISL_542360, EPI_ISL_542361, EPI_ISL_542362, EPI_ISL_542363, EPI_ISL_542364, EPI_ISL_542365, EPI_ISL_542366, EPI_ISL_542367, EPI_ISL_542368, EPI_ISL_542369, EPI_ISL_542370, EPI_ISL_542371, EPI_ISL_542372, EPI_ISL_542373, EPI_ISL_542374, EPI_ISL_542375, EPI_ISL_542376, EPI_ISL_542377, EPI_ISL_542378, EPI_ISL_542379, EPI_ISL_542380, EPI_ISL_542381, EPI_ISL_542382, EPI_ISL_542383, EPI_ISL_542384, EPI_ISL_542385, EPI_ISL_542386, EPI_ISL_542387, EPI_ISL_542388, EPI_ISL_542389, EPI_ISL_542390, EPI_ISL_542391, EPI_ISL_542392, EPI_ISL_542393, EPI_ISL_542394, EPI_ISL_542395, EPI_ISL_542396, EPI_ISL_542397, EPI_ISL_542398, EPI_ISL_542399 | | | |
| see above | San Matteo Hospital Pavia | Dep. Of Oncology and Hemato-Oncology University of Milan | Claudia Alteri, Valeria Cento, Antonio Piralla, Valentino Costabile, Monica Tallarita, Luna Colagrossi, Silvia Renica, Federica Giardina, Federica Novazzi, Stefano Gaiarsa, Elisa Matarazzo, Maria Antonello, Chiara Vismara, Roberto Fumagalli, Oscar Massimiliano Epis, Massimo Puoti, Carlo Federico Perno, Fausto Baldanti |
| EPI_ISL_542400, EPI_ISL_542401, EPI_ISL_542402, EPI_ISL_542403, EPI_ISL_542404, EPI_ISL_542405, EPI_ISL_542406, EPI_ISL_542407, EPI_ISL_542408, EPI_ISL_542409, EPI_ISL_542410, EPI_ISL_542411, EPI_ISL_542412, EPI_ISL_542413, EPI_ISL_542414, EPI_ISL_542415, EPI_ISL_542416, EPI_ISL_542417, EPI_ISL_542418, EPI_ISL_542419, EPI_ISL_542420, EPI_ISL_542421, EPI_ISL_542422, EPI_ISL_542423, EPI_ISL_542424, EPI_ISL_542425, EPI_ISL_542426, EPI_ISL_542427, EPI_ISL_542428, EPI_ISL_542429, EPI_ISL_542430, EPI_ISL_542431, EPI_ISL_542432, EPI_ISL_542433, EPI_ISL_542434, EPI_ISL_542435, EPI_ISL_542436, EPI_ISL_542437, EPI_ISL_542438, EPI_ISL_542439, EPI_ISL_542440, EPI_ISL_542441, EPI_ISL_542442, EPI_ISL_542443 | | | |
| see above | ASST GOM Niguarda | Dep. Of Oncology and Hemato-Oncology University of Milan | Claudia Alteri, Valeria Cento, Antonio Piralla, Valentino Costabile, Monica Tallarita, Luna Colagrossi, Silvia Renica, Federica Giardina, Federica Novazzi, Stefano Gaiarsa, Elisa Matarazzo, Maria Antonello, Chiara Vismara, Roberto Fumagalli, Oscar Massimiliano Epis, Massimo Puoti, Carlo Federico Perno, Fausto Baldanti |
| EPI_ISL_547965 | Laboratorio Biologia Molecolare SarsCov2 UOC LAboratorio Analisi SErvizio Medicina di Laboratorio Ospedale San Francesco ATS-ASSL Nuoro | Laboratorio Specialistico UOC Ematologia Ospedale San Francesco - ATS ASSL NUORO | Piras Giovanna, Aspronі Rosanna, Monne Maria Itria, Fancello Tatiana,Fiamma Maura,Toja Alessandro, Sanna Filomena, Floris Anna Rita, Sulis Vincenzo, Palmas Angelo Domenico, Casu Gavino, Lo Maglio Iana, Mameli Giuseppe. |
| EPI_ISL_560407 | Istituto Zooprofilattico Sperimentale del Mezzogiorno | INMI Lazzaro Spallanzani IRCCS | Barbara Bartolini, Cesare E.M. Gruber, Martina Rueca, Francesco Messina, Antonino Di Caro, Giovanna Fusco, Maurizio Viscardi, Giorgia Borriello, Sergio Brandi, Maria R. Capobianchi |
| EPI_ISL_568579 | Virus Molecular Laboratory of the Microbiology and Virology Department | INMI Lazzaro Spallanzani IRCCS | Cesare E.M. Gruber, Martina Rueca, Barbara Bartolini, Francesco Messina, Silvia Meschi, Francesca Colavita, Concetta Castilletti, Elena Percivalle, Irene Cassaniti, Edoardo Vecchio Nepita, Fausto Baldanti, Maria R. Capobianchi, Antonino Di Caro |
| EPI_ISL_569865, EPI_ISL_569866, EPI_ISL_569867, EPI_ISL_569868, EPI_ISL_569869, EPI_ISL_569870, EPI_ISL_569871, EPI_ISL_569872, EPI_ISL_569873, EPI_ISL_569874, EPI_ISL_569875, EPI_ISL_569876, EPI_ISL_569877, EPI_ISL_569878, EPI_ISL_569879, EPI_ISL_569880, EPI_ISL_569881, EPI_ISL_569882, EPI_ISL_569883, EPI_ISL_569884, EPI_ISL_569885, EPI_ISL_569886 | | | |
| see above | Amedeo di savoia | Crosetto lab, Karolinska Institutet, SciLifeLab | Michele Simonetti, Maria Grazia Milia, Luuk Harbers, Ning Zhang, Anna Sapino, Valeria Ghisetti, Nicola Crosetto |
| EPI_ISL_572320, EPI_ISL_572321, EPI_ISL_572322, EPI_ISL_572323, EPI_ISL_572324 | IZSM | IZSM | Maurizio Viscardi, Lorena Cardillo, Giovanna Fusco |
| EPI_ISL_582123, EPI_ISL_583954, EPI_ISL_583955, EPI_ISL_583956, EPI_ISL_583957, EPI_ISL_583958, EPI_ISL_583959, EPI_ISL_583960, EPI_ISL_583961, EPI_ISL_583962, EPI_ISL_583963, EPI_ISL_583964, EPI_ISL_583965, EPI_ISL_583966, EPI_ISL_583967 | | | |
| see above | UOC Microbiologia e Virologia, Azienda Ospedaliera Universitaria Senese, Siena, Italy | Dipartimento di Biotecnologie Mediche | Maria Grazia Cusi, David Pinzauti, Claudia Gandolfo, Gabriele Anichini, Gianni Pozzi, Francesco Santoro |
| EPI_ISL_584048 | Laboratory of Molecular Virology, Department of Biomedical, Surgical and Dental Sciences University of Milano | Laboratory of Molecular Virology, Department of Biomedical, Surgical and Dental Sciences University of Milano | Delbue,S., Modenese,A., Bianchi,M., Fattori,M., D'Alessandro,S., Pariani,E., Basilico,N., Galli,C. and Ferrante,P. |
| EPI_ISL_584049 | Laboratory of Molecular Virology, Department of Biomedical, Surgical and Dental Sciences University of Milano | Laboratory of Molecular Virology, Department of Biomedical, Surgical and Dental Sciences University of Milano | Delbue,S., Modenese,A., Bianchi,M., Fattori,M., D'Alessandro,S.,Pariani,E., Basilico,N., Galli,C. and Ferrante,P. |
| EPI_ISL_584051 | Laboratory of Molecular Virology, Department of Biomedical, Surgical and Dental Sciences University of Milano | Laboratory of Molecular Virology, Department of Biomedical, Surgical and Dental Sciences University of Milano | Delbue,S., Ferrante,P., Basilico,N., Parapini,S., Binda,S., D'Alessandro,S., Galli,C., Signorini,L., Primache,V., Anselmi,G. and Pariani,E |
| EPI_ISL_584052 | Laboratory of Molecular Virology, Department of Biomedical, Surgical and Dental Sciences University of Milano | Laboratory of Molecular Virology, Department of Biomedical, Surgical and Dental Sciences University of Milano | Delbue,S., D'Alessandro,S., Modenese,A., Signorini,L., Parapini,S.,Dolci,M., Binda,S., Primache,V., Taramelli,D., Incorvaia,B. and Ferrante,P. |
| EPI_ISL_584069, EPI_ISL_584071, EPI_ISL_584072 | IZSM | IZSM | Maurizio Viscardi, Lorena Cardillo, Giovanna Fusco |
| EPI_ISL_590693 | INMI Lazzaro Spallanzani IRCCS | INMI Lazzaro Spallanzani IRCCS | Martina Rueca, Barbara Bartolini, Cesare E.M. Gruber, Francesco Messina, Emanuela Giombini, Beatrice Valli, Eleonora Lalle, Simone Lanini, Francesco Vairo, Maria R. Capobianchi, Antonino Di Caro |
| EPI_ISL_590694 | INMI Lazzaro Spallanzani IRCCS | INMI Lazzaro Spallanzani IRCCS | Barbara Bartolini, Martina Rueca, Francesco Messina, Cesare E.M. Gruber, Emanuela Giombini, Beatrice Valli, Eleonora Lalle, Simone Lanini, Francesco Vairo, Maria R. Capobianchi, Antonino Di Caro |
| EPI_ISL_590695 | INMI Lazzaro Spallanzani IRCCS | INMI Lazzaro Spallanzani IRCCS | Cesare E.M. Gruber, Francesco Messina, Barbara Bartolini, Martina Rueca, Emanuela Giombini, Beatrice Valli, Eleonora Lalle, Simone Lanini, Francesco Vairo, Antonino Di Caro, Maria R. Capobianchi |
| EPI_ISL_590696 | INMI Lazzaro Spallanzani IRCCS | INMI Lazzaro Spallanzani IRCCS | Cesare E.M. Gruber, Barbara Bartolini, Francesco Messina, Martina Rueca, Emanuela Giombini, Beatrice Valli, Eleonora Lalle, Simone Lanini, Francesco Vairo, Antonino Di Caro, Maria R. Capobianchi |
| EPI_ISL_590697 | INMI Lazzaro Spallanzani IRCCS | INMI Lazzaro Spallanzani IRCCS | Martina Rueca, Cesare E.M. Gruber, Barbara Bartolini, Francesco Messina, Martina Rueca, Emanuela Giombini, Beatrice Valli, Eleonora Lalle, Simone Lanini, Francesco Vairo, Antonino Di Caro, Maria R. Capobianchi |
| EPI_ISL_590698 | INMI Lazzaro Spallanzani IRCCS | INMI Lazzaro Spallanzani IRCCS | Barbara Bartolini, Francesco Messina, Cesare E.M. Gruber, Martina Rueca, Emanuela Giombini, Beatrice Valli, Eleonora Lalle, Simone Lanini, Francesco Vairo, Maria R. Capobianchi, Antonino Di Caro |
| EPI_ISL_591327, EPI_ISL_591328, EPI_ISL_591329, EPI_ISL_591330, EPI_ISL_591331, EPI_ISL_591332, EPI_ISL_591333, EPI_ISL_591334, EPI_ISL_591335 | Dipartimento di Biotecnologie Mediche, University of Siena | Dipartimento di Biotecnologie Mediche, University of Siena | Cusi,M.G., Pinzauti,D., Gandolfo,C., Anichini,G., Pozzi,G., Santoro,F. |
| EPI_ISL_591336 | Dipartimento di Biotecnologie Mediche, University of Siena | Dipartimento di Biotecnologie Mediche, University of Siena | COVID |
| EPI_ISL_591337, EPI_ISL_591338, EPI_ISL_591339, EPI_ISL_591340 | Dipartimento di Biotecnologie Mediche, University of Siena | Dipartimento di Biotecnologie Mediche, University of Siena | Cusi,M.G., Pinzauti,D., Gandolfo,C., Anichini,G., Pozzi,G., Santoro,F. |
| EPI_ISL_602304 | Istituto Zooprofilattico Sperimentale del Mezzogiorno | U.O. Diagnostica Virologica Dip. Sanità Animale IZSM | Maurizio Viscardi, Lorena Cardillo , e Giovanna Fusco |
| EPI_ISL_603137 | INMI Lazzaro Spallanzani IRCCS | INMI Lazzaro Spallanzani IRCCS | Cesare E.M. Gruber, Martina Rueca, Barbara Bartolini, Francesco Messina, Emanuela Giombini, Simone Lanini, Antonino Di Caro, Maria R. Capobianchi |
| EPI_ISL_603138 | INMI Lazzaro Spallanzani IRCCS | INMI Lazzaro Spallanzani IRCCS | Martina Rueca, Francesco Messina, Barbara Bartolini, Cesare E.M. Gruber, Emanuela Giombini, Simone Lanini, Antonino Di Caro, Maria R. Capobianchi |
| EPI_ISL_603139 | INMI Lazzaro Spallanzani IRCCS | INMI Lazzaro Spallanzani IRCCS | Martina Rueca, Cesare E.M. Gruber, Barbara Bartolini, Francesco Messina, Emanuela Giombini, Simone Lanini, Antonino Di Caro, Maria R. Capobianchi |
| EPI_ISL_603140 | INMI Lazzaro Spallanzani IRCCS | INMI Lazzaro Spallanzani IRCCS | Martina Rueca, Cesare E.M. Gruber, Francesco Messina, Barbara Bartolini, Emanuela Giombini, Simone Lanini, Antonino Di Caro, Maria R. Capobianchi |
| EPI_ISL_603141 | INMI Lazzaro Spallanzani IRCCS | INMI Lazzaro Spallanzani IRCCS | Francesco Messina, Cesare E.M. Gruber, Martina Rueca, Barbara Bartolini, Emanuela Giombini, Simone Lanini, Maria R. Capobianchi, Antonino Di Caro |
| EPI_ISL_603142 | INMI Lazzaro Spallanzani IRCCS | INMI Lazzaro Spallanzani IRCCS | Barbara Bartolini, Cesare E.M. Gruber, Francesco Messina, Martina Rueca, Simone Lanini, Emanuela Giombini, Maria R. Capobianchi, Antonino Di Caro |

| | | | |
|---|---|---|---|
| EPI_ISL_603143 | INMI Lazzaro Spallanzani IRCCS | INMI Lazzaro Spallanzani IRCCS | Martina Rueca, Francesco Messina, Cesare E.M. Gruber, Barbara Bartolini, Emanuela Giombini, Simone Lanini, Antonino Di Caro, Maria R. Capobianchi |
| EPI_ISL_603144 | INMI Lazzaro Spallanzani IRCCS | INMI Lazzaro Spallanzani IRCCS | Cesare E.M. Gruber, Martina Rueca, Francesco Messina, Barbara Bartolini, Emanuela Giombini, Simone Lanini, Maria R. Capobianchi, Antonino Di Caro |
| EPI_ISL_603145 | INMI Lazzaro Spallanzani IRCCS | INMI Lazzaro Spallanzani IRCCS | Francesco Messina, Cesare E.M. Gruber, Martina Rueca, Barbara Bartolini, Emanuela Giombini, Simone Lanini, Antonino Di Caro, Maria R. Capobianchi |
| EPI_ISL_603146 | INMI Lazzaro Spallanzani IRCCS | INMI Lazzaro Spallanzani IRCCS | Cesare E.M. Gruber, Martina Rueca, Barbara Bartolini, Emanuela Giombini, Francesco Messina, Simone Lanini, Maria R. Capobianchi, Antonino Di Caro |
| EPI_ISL_603147 | INMI Lazzaro Spallanzani IRCCS | INMI Lazzaro Spallanzani IRCCS | Cesare E.M. Gruber, Barbara Bartolini, Francesco Messina, Martina Rueca, Simone Lanini, Emanuela Giombini, Antonino Di Caro, Maria R. Capobianchi |
| EPI_ISL_603148 | INMI Lazzaro Spallanzani IRCCS | INMI Lazzaro Spallanzani IRCCS | Cesare E.M. Gruber, Francesco Messina, Martina Rueca, Simone Lanini, Emanuela Giombini, Barbara Bartolini, Maria R. Capobianchi, Antonino Di Caro |
| EPI_ISL_603149 | INMI Lazzaro Spallanzani IRCCS | INMI Lazzaro Spallanzani IRCCS | Francesco Messina, Barbara Bartolini, Cesare E.M. Gruber, Martina Rueca, Simone Lanini, Emanuela Giombini, Patrizia Massarelli, Antonino Di Caro, Maria R. Capobianchi |
| EPI_ISL_603150 | INMI Lazzaro Spallanzani IRCCS | INMI Lazzaro Spallanzani IRCCS | Barbara Bartolini, Francesco Messina, Martina Rueca, Cesare E.M. Gruber, Simone Lanini, Emanuela Giombini, Patrizia Massarelli, Maria R. Capobianchi, Antonino Di Caro |
| EPI_ISL_603151 | INMI Lazzaro Spallanzani IRCCS | INMI Lazzaro Spallanzani IRCCS | Francesco Messina, Barbara Bartolini, Cesare E.M. Gruber, Martina Rueca, Emanuela Giombini, Simone Lanini, Patrizia Massarelli, Maria R. Capobianchi, Antonino Di Caro |
| EPI_ISL_603152 | INMI Lazzaro Spallanzani IRCCS | INMI Lazzaro Spallanzani IRCCS | Barbara Bartolini, Cesare E.M. Gruber, Francesco Messina, Martina Rueca, Emanuela Giombini, Simone Lanini, Patrizia Massarelli, Antonino Di Caro, Maria R. Capobianchi |
| EPI_ISL_603153 | INMI Lazzaro Spallanzani IRCCS | INMI Lazzaro Spallanzani IRCCS | Francesco Messina, Cesare E.M. Gruber, Barbara Bartolini, Simone Lanini, Emanuela Giombini, Martina Rueca, Marcello Meledandri, Maria Letizia Schiavone, Annunziata Tamburro, Antonino Di Caro, Maria R. Capobianchi |
| EPI_ISL_603154 | INMI Lazzaro Spallanzani IRCCS | INMI Lazzaro Spallanzani IRCCS | Martina Rueca, Francesco Messina, Cesare E.M. Gruber, Barbara Bartolini, Emanuela Giombini, Simone Lanini, Antonino Di Caro, Maria R. Capobianchi |
| EPI_ISL_603155 | INMI Lazzaro Spallanzani IRCCS | INMI Lazzaro Spallanzani IRCCS | Martina Rueca, Francesco Messina, Barbara Bartolini, Cesare E.M. Gruber, Simone Lanini, Emanuela Giombini, Maria R. Capobianchi, Antonino Di Caro |
| EPI_ISL_603156 | INMI Lazzaro Spallanzani IRCCS | INMI Lazzaro Spallanzani IRCCS | Barbara Bartolini, Francesco Messina, Cesare E.M. Gruber, Martina Rueca, Emanuela Giombini, Simone Lanini, Maria R. Capobianchi, Antonino Di Caro |
| EPI_ISL_603157 | INMI Lazzaro Spallanzani IRCCS | INMI Lazzaro Spallanzani IRCCS | Francesco Messina, Martina Rueca, Barbara Bartolini, Cesare E.M. Gruber, Simone Lanini, Emanuela Giombini, Fulvia Pimpinelli, Antonino Di Caro, Maria R. Capobianchi |
| EPI_ISL_603158 | INMI Lazzaro Spallanzani IRCCS | INMI Lazzaro Spallanzani IRCCS | Barbara Bartolini, Cesare E.M. Gruber, Francesco Messina, Martina Rueca, Emanuela Giombini, Simone Lanini, Fulvia Pimpinelli, Maria R. Capobianchi, Antonino Di Caro |
| EPI_ISL_603159 | INMI Lazzaro Spallanzani IRCCS | INMI Lazzaro Spallanzani IRCCS | Barbara Bartolini, Francesco Messina, Cesare E.M. Gruber, Martina Rueca, Simone Lanini, Emanuela Giombini, Fulvia Pimpinelli, Maria R. Capobianchi, Antonino Di Caro |
| EPI_ISL_603160 | INMI Lazzaro Spallanzani IRCCS | INMI Lazzaro Spallanzani IRCCS | Barbara Bartolini, Simone Lanini, Emanuela Giombini, Francesco Messina, Cesare E.M. Gruber, Martina Rueca, Fulvia Pimpinelli, Maria R. Capobianchi, Antonino Di Caro |
| EPI_ISL_603161 | INMI Lazzaro Spallanzani IRCCS | INMI Lazzaro Spallanzani IRCCS | Cesare E.M. Gruber, Barbara Bartolini, Francesco Messina, Martina Rueca, Emanuela Giombini, Simone Lanini, Fulvia Pimpinelli, Antonino Di Caro, Maria R. Capobianchi |
| EPI_ISL_603162 | INMI Lazzaro Spallanzani IRCCS | INMI Lazzaro Spallanzani IRCCS | Cesare E.M. Gruber, Francesco Messina, Martina Rueca, Barbara Bartolini, Emanuela Giombini, Simone Lanini, Fulvia Pimpinelli, Antonino Di Caro, Maria R. Capobianchi |
| EPI_ISL_603163 | INMI Lazzaro Spallanzani IRCCS | INMI Lazzaro Spallanzani IRCCS | Cesare E.M. Gruber, Martina Rueca, Emanuela Giombini, Simone Lanini, Francesco Messina, Barbara Bartolini, Fulvia Pimpinelli, Maria R. Capobianchi, Antonino Di Caro |
| EPI_ISL_603164 | INMI Lazzaro Spallanzani IRCCS | INMI Lazzaro Spallanzani IRCCS | Cesare E.M. Gruber, Francesco Messina, Martina Rueca, Barbara Bartolini, Emanuela Giombini, Simone Lanini, Fulvia Pimpinelli, Antonino Di Caro, Maria R. Capobianchi |
| EPI_ISL_603165 | INMI Lazzaro Spallanzani IRCCS | INMI Lazzaro Spallanzani IRCCS | Martina Rueca, Francesco Messina, Barbara Bartolini, Cesare E.M. Gruber, Emanuela Giombini, Simone Lanini, Fulvia Pimpinelli, Maria R. Capobianchi, Antonino Di Caro |
| EPI_ISL_603166 | INMI Lazzaro Spallanzani IRCCS | INMI Lazzaro Spallanzani IRCCS | Martina Rueca, Cesare E.M. Gruber, Barbara Bartolini, Francesco Messina, Simone Lanini, Emanuela Giombini, Fulvia Pimpinelli, Maria R. Capobianchi, Antonino Di Caro |
| EPI_ISL_603167 | INMI Lazzaro Spallanzani IRCCS | INMI Lazzaro Spallanzani IRCCS | Martina Rueca, Cesare E.M. Gruber, Francesco Messina, Barbara Bartolini, Simone Lanini, Emanuela Giombini, Fulvia Pimpinelli, Maria R. Capobianchi, Antonino Di Caro |
| EPI_ISL_603168 | INMI Lazzaro Spallanzani IRCCS | INMI Lazzaro Spallanzani IRCCS | Martina Rueca, Barbara Bartolini, Cesare E.M. Gruber, Simone Lanini, Emanuela Giombini, Francesco Messina, Fulvia Pimpinelli, Antonino Di Caro, Maria R. Capobianchi |
| EPI_ISL_603169 | INMI Lazzaro Spallanzani IRCCS | INMI Lazzaro Spallanzani IRCCS | Francesco Messina, Barbara Bartolini, Cesare E.M. Gruber, Martina Rueca, Simone Lanini, Emanuela Giombini, Fulvia Pimpinelli, Maria R. Capobianchi, Antonino Di Caro |
| EPI_ISL_603170 | INMI Lazzaro Spallanzani IRCCS | INMI Lazzaro Spallanzani IRCCS | Martina Rueca, Barbara Bartolini, Cesare E.M. Gruber, Francesco Messina, Simone Lanini, Emanuela Giombini, Maria R. Capobianchi, Antonino Di Caro |
| EPI_ISL_603171 | INMI Lazzaro Spallanzani IRCCS | INMI Lazzaro Spallanzani IRCCS | Barbara Bartolini, Cesare E.M. Gruber, Francesco Messina, Martina Rueca, Emanuela Giombini, Simone Lanini, Mariarosa Gaudio, Antonino Di Caro, Maria R. Capobianchi |
| EPI_ISL_603172 | INMI Lazzaro Spallanzani IRCCS | INMI Lazzaro Spallanzani IRCCS | Cesare E.M. Gruber, Martina Rueca, Barbara Bartolini, Simone Lanini, Francesco Messina, Emanuela Giombini, Mariarosa Gaudio, Antonino Di Caro, Maria R. Capobianchi |
| EPI_ISL_603173 | INMI Lazzaro Spallanzani IRCCS | INMI Lazzaro Spallanzani IRCCS | Barbara Bartolini, Cesare E.M. Gruber, Martina Rueca, Francesco Messina, Simone Lanini, Emanuela Giombini, Giuseppina Cappiello, Maria R. Capobianchi, Antonino Di Caro |
| EPI_ISL_603174 | INMI Lazzaro Spallanzani IRCCS | INMI Lazzaro Spallanzani IRCCS | Francesco Messina, Barbara Bartolini, Martina Rueca, Emanuela Giombini, Simone Lanini, Cesare E.M. Gruber, Mariarosa Gaudio, Maria R. Capobianchi, Antonino Di Caro |
| EPI_ISL_603175 | INMI Lazzaro Spallanzani IRCCS | INMI Lazzaro Spallanzani IRCCS | Barbara Bartolini, Cesare E.M. Gruber, Simone Lanini, Emanuela Giombini, Francesco Messina, Martina Rueca, Mariarosa Gaudio, Antonino Di Caro, Maria R. Capobianchi |
| EPI_ISL_603176 | INMI Lazzaro Spallanzani IRCCS | INMI Lazzaro Spallanzani IRCCS | Cesare E.M. Gruber, Barbara Bartolini, Francesco Messina, Martina Rueca, Simone Lanini, Emanuela Giombini, Antonino Di Caro, Maria R. Capobianchi |
| EPI_ISL_603177 | INMI Lazzaro Spallanzani IRCCS | INMI Lazzaro Spallanzani IRCCS | Barbara Bartolini, Francesco Messina, Emanuela Giombini, Martina Rueca, Cesare E.M. Gruber, Simone Lanini, Maria R. Capobianchi, Antonino Di Caro |
| EPI_ISL_603178 | INMI Lazzaro Spallanzani IRCCS | INMI Lazzaro Spallanzani IRCCS | Martina Rueca, Francesco Messina, Cesare E.M. Gruber, Barbara Bartolini, Simone Lanini, Emanuela Giombini, Antonino Di Caro, Maria R. Capobianchi |
| EPI_ISL_603179 | INMI Lazzaro Spallanzani IRCCS | INMI Lazzaro Spallanzani IRCCS | Cesare E.M. Gruber, Barbara Bartolini, Francesco Messina, Martina Rueca, Emanuela Giombini, Simone Lanini, Antonino Di Caro, Maria R. Capobianchi |
| EPI_ISL_603180 | INMI Lazzaro Spallanzani IRCCS | INMI Lazzaro Spallanzani IRCCS | Barbara Bartolini, Cesare E.M. Gruber, Francesco Messina, Simone Lanini, Martina Rueca, Emanuela Giombini, Maria R. Capobianchi, Antonino Di Caro |
| EPI_ISL_603181 | INMI Lazzaro Spallanzani IRCCS | INMI Lazzaro Spallanzani IRCCS | Francesco Messina, Cesare E.M. Gruber, Barbara Bartolini, Martina Rueca, Simone Lanini, Emanuela Giombini, Maria R. Capobianchi, Antonino Di Caro |
| EPI_ISL_603182 | INMI Lazzaro Spallanzani IRCCS | INMI Lazzaro Spallanzani IRCCS | Martina Rueca, Barbara Bartolini, Cesare E.M. Gruber, Francesco Messina, Emanuela Giombini, Simone Lanini, Maria R. Capobianchi, Antonino Di Caro |
| EPI_ISL_603183 | INMI Lazzaro Spallanzani IRCCS | INMI Lazzaro Spallanzani IRCCS | Barbara Bartolini, Francesco Messina, Martina Rueca, Cesare E.M. Gruber, Emanuela Giombini, Simone Lanini, Maria R. Capobianchi, Antonino Di Caro |
| EPI_ISL_603184 | INMI Lazzaro Spallanzani IRCCS | INMI Lazzaro Spallanzani IRCCS | Martina Rueca, Barbara Bartolini, Francesco Messina, Cesare E.M. Gruber, Emanuela Giombini, Simone Lanini, Maria R. Capobianchi, Antonino Di Caro |
| EPI_ISL_603185 | INMI Lazzaro Spallanzani IRCCS | INMI Lazzaro Spallanzani IRCCS | Cesare E.M. Gruber, Martina Rueca, Barbara Bartolini, Francesco Messina, Simone Lanini, Emanuela Giombini, Maria R. Capobianchi, Antonino Di Caro |

| | | | |
|---|---|---|---|
| EPI_ISL_603186 | INMI Lazzaro Spallanzani IRCCS | INMI Lazzaro Spallanzani IRCCS | Francesco Messina, Martina Rueca, Barbara Bartolini, Cesare E.M. Gruber, Emanuela Giombini, Simone Lanini, Maria R. Capobianchi, Antonino Di Caro |
| EPI_ISL_603187 | INMI Lazzaro Spallanzani IRCCS | INMI Lazzaro Spallanzani IRCCS | Cesare E.M. Gruber, Martina Rueca, Francesco Messina, Barbara Bartolini, Simone Lanini, Emanuela Giombini, Antonino Di Caro, Maria R. Capobianchi |
| EPI_ISL_609989 | INMI Lazzaro Spallanzani IRCCS | INMI Lazzaro Spallanzani IRCCS | C.E.M Gruber, B Bartolini, M Rueca, F Messina, E Giombini, A Di Caro, MR Capobianchi |
| EPI_ISL_609990 | INMI Lazzaro Spallanzani IRCCS | INMI Lazzaro Spallanzani IRCCS | B Bartolini, C.E.M Gruber, M Rueca, F Messina, E Giombini, MR Capobianchi, A Di Caro |
| EPI_ISL_609991 | INMI Lazzaro Spallanzani IRCCS | INMI Lazzaro Spallanzani IRCCS | M Rueca, B Bartolini, C.E.M Gruber, F Messina, E Giombini, A Di Caro, MR Capobianchi |
| EPI_ISL_609992 | INMI Lazzaro Spallanzani IRCCS | INMI Lazzaro Spallanzani IRCCS | F Messina, E Giombini, M Rueca, B Bartolini, C.E.M Gruber, MR Capobianchi, A Di Caro |
| EPI_ISL_609993 | INMI Lazzaro Spallanzani IRCCS | INMI Lazzaro Spallanzani IRCCS | E Giombini, M Rueca, B Bartolini, C.E.M Gruber, F Messina, A Di Caro, MR Capobianchi |
| EPI_ISL_609994 | INMI Lazzaro Spallanzani IRCCS | INMI Lazzaro Spallanzani IRCCS | C.E.M Gruber, F Messina, M Rueca, B Bartolini, E Giombini, MR Capobianchi, A Di Caro |
| EPI_ISL_609995 | INMI Lazzaro Spallanzani IRCCS | INMI Lazzaro Spallanzani IRCCS | E Giombini, C.E.M Gruber, M Rueca, B Bartolini, F Messina, A Di Caro, MR Capobianchi |
| EPI_ISL_609996 | INMI Lazzaro Spallanzani IRCCS | INMI Lazzaro Spallanzani IRCCS | F Messina, M Rueca, B Bartolini, C.E.M Gruber, E Giombini, MR Capobianchi, A Di Caro |
| EPI_ISL_609997 | INMI Lazzaro Spallanzani IRCCS | INMI Lazzaro Spallanzani IRCCS | M Rueca, B Bartolini, C.E.M Gruber, F Messina, E Giombini, A Di Caro, MR Capobianchi |
| EPI_ISL_609998 | INMI Lazzaro Spallanzani IRCCS | INMI Lazzaro Spallanzani IRCCS | F Messina, B Bartolini, M Rueca, C.E.M Gruber, E Giombini, A Di Caro, MR Capobianchi |
| EPI_ISL_609999 | INMI Lazzaro Spallanzani IRCCS | INMI Lazzaro Spallanzani IRCCS | B Bartolini, M Rueca, C.E.M Gruber, F Messina, E Giombini, MR Capobianchi, A Di Caro |
| EPI_ISL_613560 | Laboratorio Biologia Molecolare Sars Cov2 - UOC Laboratorio Analisi - Servizio Medicina di Laboratorio, Ospedale "San Francesco" - ATS-ASSL Nuoro Via Mannironi 1, 08100 Nuoro | Laboratorio specialistico UOC Ematologia - Ospedale "San Francesco" - ATS-ASSL Nuoro Nuoro | Piras Giovanna, Fancello Tatiana, Asproni Rosanna, Fiamma Maura, Monne Maria Itria, Toja Alessandro, Sanna Filomena, Floris Anna Rita, Sulis Vincenzo, Palmas Angelo Domenico, Casu Gavino, Lo Maglio Iana, Mameli Giuseppe |
| EPI_ISL_613706 | Laboratorio Biologia Molecolare Sars Cov2 - UOC Laboratorio Analisi - Servizio Medicina di Laboratorio, Ospedale "San Francesco" - ATS-ASSL Nuoro | Laboratorio specialistico UOC Ematologia - Ospedale "San Francesco" - ATS-ASSL Nuoro | Piras Giovanna, Fancello Tatiana, Asproni Rosanna, Fiamma Maura, Monne Maria Itria, Toja Alessandro, Sanna Filomena, Floris Anna Rita, Sulis Vincenzo, Palmas Angelo Domenico, Casu Gavino, Lo Maglio Iana, Mameli Giuseppe |
| EPI_ISL_613710 | Laboratorio Biologia Molecolare Sars Cov2 - UOC Laboratorio Analisi - Servizio Medicina di Laboratorio, Ospedale "San Francesco" - ATS-ASSL Nuoro | Laboratorio specialistico UOC Ematologia - Ospedale "San Francesco" - ATS-ASSL Nuoro | Piras Giovanna, Fancello Tatiana, Asproni Rosanna, Fiamma Maura, Monne Maria Itria, Toja Alessandro, Sanna Filomena, Floris Anna Rita, Sulis Vincenzo, Palmas Angelo Domenico, Casu Gavino, Lo Maglio Iana, Mameli Giuseppe. |
| EPI_ISL_613953, EPI_ISL_613955, EPI_ISL_614396, EPI_ISL_614397, EPI_ISL_614398, EPI_ISL_614889 | Laboratorio Biologia Molecolare Sars Cov2 - UOC Laboratorio Analisi - Servizio Medicina di Laboratorio, Ospedale "San Francesco" - ATS-ASSL Nuoro | Laboratorio specialistico UOC Ematologia - Ospedale "San Francesco" - ATS-ASSL Nuoro | Piras Giovanna, Fancello Tatiana, Asproni Rosanna, Fiamma Maura, Monne Maria Itria, Toja Alessandro, Sanna Filomena, Floris Anna Rita, Sulis Vincenzo, Palmas Angelo Domenico, Casu Gavino, Lo Maglio Iana, Mameli Giuseppe |
| EPI_ISL_636462, EPI_ISL_636463 | ULSS6 Euganea | Istituto Zooprofilattico Sperimentale delle Venezie | Adelaide Milani, Alessia Schivo, Annalisa Salviato, Erika Giorgia Quaranta, Ambra Pastori, Bianca Zecchin, Alice Fusaro, Isabella Monne, Calogero Terregino, Antonia Ricci |
| EPI_ISL_636464 | ULSS6 Piove di Sacco | Istituto Zooprofilattico Sperimentale delle Venezie | Adelaide Milani, Alessia Schivo, Annalisa Salviato, Erika Giorgia Quaranta, Ambra Pastori, Bianca Zecchin, Alice Fusaro, Isabella Monne, Calogero Terregino, Antonia Ricci |
| EPI_ISL_636465, EPI_ISL_636466 | ULSS6 Euganea | Istituto Zooprofilattico Sperimentale delle Venezie | Adelaide Milani, Alessia Schivo, Annalisa Salviato, Erika Giorgia Quaranta, Ambra Pastori, Bianca Zecchin, Alice Fusaro, Isabella Monne, Calogero Terregino, Antonia Ricci |
| EPI_ISL_636467, EPI_ISL_636468, EPI_ISL_636469, EPI_ISL_636470 | ULSS6 Distretto Padova Terme Colli | Istituto Zooprofilattico Sperimentale delle Venezie | Adelaide Milani, Alessia Schivo, Annalisa Salviato, Erika Giorgia Quaranta, Ambra Pastori, Bianca Zecchin, Alice Fusaro, Isabella Monne, Calogero Terregino, Antonia Ricci |
| EPI_ISL_636471, EPI_ISL_636472, EPI_ISL_636473 | ULSS6 Piove di Sacco | Istituto Zooprofilattico Sperimentale delle Venezie | Adelaide Milani, Alessia Schivo, Annalisa Salviato, Erika Giorgia Quaranta, Ambra Pastori, Bianca Zecchin, Alice Fusaro, Isabella Monne, Calogero Terregino, Antonia Ricci |
| EPI_ISL_636474, EPI_ISL_636475 | ULSS6 Distretto Padova Terme Colli | Istituto Zooprofilattico Sperimentale delle Venezie | Adelaide Milani, Alessia Schivo, Annalisa Salviato, Erika Giorgia Quaranta, Ambra Pastori, Bianca Zecchin, Alice Fusaro, Isabella Monne, Calogero Terregino, Antonia Ricci |
| EPI_ISL_636488 | ULSS9 Scaligera | Istituto Zooprofilattico Sperimentale delle Venezie | Adelaide Milani, Alessia Schivo, Annalisa Salviato, Erika Giorgia Quaranta, Ambra Pastori, Bianca Zecchin, Alice Fusaro, Isabella Monne, Calogero Terregino, Antonia Ricci |
| EPI_ISL_637107, EPI_ISL_637108, EPI_ISL_637109 | Laboratorio Biologia Molecolare Sars Cov2 - UOC Laboratorio Analisi - Servizio Medicina di Laboratorio, Ospedale "San Francesco" - ATS-ASSL Nuoro | Laboratorio specialistico UOC Ematologia - Ospedale "San Francesco" - ATS-ASSL Nuoro | Piras Giovanna, Fancello Tatiana, Asproni Rosanna, Fiamma Maura, Monne Maria Itria, Toja Alessandro, Sanna Filomena, Floris Anna Rita, Sulis Vincenzo, Palmas Angelo Domenico, Casu Gavino, Lo Maglio Iana, Mameli Giuseppe |
| EPI_ISL_649189, EPI_ISL_649190 | Istituto Zooprofilattico Sperimentale della Puglia e della Basilicata | Istituto Zooprofilattico Sperimentale della Puglia e della Basilicata | Parisi A., Bianco A., Capozzi L., Del Sambro L., Manzulli V, Rondinone V., Pace L., Galante D., Cipolletta D. |
| EPI_ISL_649191 | Istituto Zooprofilattico Sperimentale della Puglia e della Basilicata | Istituto Zooprofilattico Sperimentale della Puglia e della Basilicata | Parisi A., Bianco A., Capozzi L., Del Sambro L., Manzulli V, Rondinone V., Pace L., Cipolletta D., Galante D. |
| EPI_ISL_649785 | I.R.C.C.S. "S. De Bellis" - Ente Ospedaliero | Istituto Zooprofilattico Sperimentale della Puglia e della Basilicata | Parisi A., Bianco A., Capozzi L., Del Sambro L., Lippolis A., Notarnicola M., Manzulli V, Rondinone V., Pace L. |
| EPI_ISL_649938 | I.R.C.C.S. "S. De Bellis" - Ente Ospedaliero | Istituto Zooprofilattico Sperimentale della Puglia e della Basilicata | Parisi A., Bianco A., Capozzi L., Del Sambro L., Lippolis A., Notarnicola M., Cipolletta D., Galante D. |
| EPI_ISL_649939 | I.R.C.C.S. "S. De Bellis" - Ente Ospedaliero | Istituto Zooprofilattico Sperimentale della Puglia e della Basilicata | Parisi A., Bianco A., Capozzi L., Del Sambro L., Lippolis A., Notarnicola M., Manzulli V, Rondinone V., Pace L. |
| EPI_ISL_649940 | Istituto Zooprofilattico Sperimentale della Puglia e della Basilicata | Istituto Zooprofilattico Sperimentale della Puglia e della Basilicata | Parisi A., Bianco A., Capozzi L., Del Sambro L., Manzulli V, Rondinone V., Pace L., Cipolletta D., Galante D. |
| EPI_ISL_653763, EPI_ISL_653764, EPI_ISL_653765, EPI_ISL_653766, EPI_ISL_653767, EPI_ISL_653768, EPI_ISL_653769, EPI_ISL_653770, EPI_ISL_653771, EPI_ISL_653772, EPI_ISL_653773, EPI_ISL_653774, EPI_ISL_653775, EPI_ISL_653776, EPI_ISL_653777, EPI_ISL_653778, EPI_ISL_653779, EPI_ISL_653780, EPI_ISL_653781, EPI_ISL_653782, EPI_ISL_653783 | | | |
| see above | I.R.C.C.S. "S. De Bellis" - Ente Ospedaliero | Istituto Zooprofilattico Sperimentale della Puglia e della Basilicata | Parisi A., Bianco A., Capozzi L., Del Sambro L., Lippolis A., Notarnicola M., Manzulli V, Rondinone V., Pace L. |
| EPI_ISL_653784, EPI_ISL_653785, EPI_ISL_653786 | Istituto Zooprofilattico Sperimentale della Puglia e della Basilicata | Istituto Zooprofilattico Sperimentale della Puglia e della Basilicata | Parisi A., Bianco A., Capozzi L., Del Sambro L., Manzulli V, Rondinone V., Pace L., Cipolletta D., Galante D. |
| EPI_ISL_653787, EPI_ISL_653788, EPI_ISL_653789, EPI_ISL_653790, EPI_ISL_653791, EPI_ISL_653792, EPI_ISL_653793 | I.R.C.C.S. "S. De Bellis" - Ente Ospedaliero | Istituto Zooprofilattico Sperimentale della Puglia e della Basilicata | Parisi A., Bianco A., Capozzi L., Del Sambro L., Lippolis A., Notarnicola M., Manzulli V, Rondinone V., Pace L. |
| EPI_ISL_653794, EPI_ISL_653795, EPI_ISL_653796, EPI_ISL_653797, EPI_ISL_653798, EPI_ISL_653799, EPI_ISL_653800, EPI_ISL_653801, EPI_ISL_653802, EPI_ISL_653803, EPI_ISL_653804, EPI_ISL_653805, EPI_ISL_653806, EPI_ISL_653807, EPI_ISL_653808, EPI_ISL_653809, EPI_ISL_653810, EPI_ISL_653811, EPI_ISL_653812 | | | |
| see above | I.R.C.C.S. "S. De Bellis" - Ente Ospedaliero | Istituto Zooprofilattico Sperimentale della Puglia e della Basilicata | Parisi A., Bianco A., Capozzi L., Del Sambro L., Lippolis A., Notarnicola M., Cipolletta D., Galante D. |
| EPI_ISL_653813 | Istituto Zooprofilattico Sperimentale della Puglia e della | Istituto Zooprofilattico Sperimentale della Puglia e della | Parisi A., Bianco A., Capozzi L., Del Sambro L., Manzulli V, Rondinone V., Pace L., Cipolletta D., Galante D. |

| | | | |
|---|---|---|---|
| | Basilicata | Basilicata | |
| EPI_ISL_653814, EPI_ISL_653815, EPI_ISL_653816, EPI_ISL_653817, EPI_ISL_653818, EPI_ISL_653819, EPI_ISL_653820, EPI_ISL_653821, EPI_ISL_653822, EPI_ISL_653823 | I.R.C.C.S. "S. De Bellis" - Ente Ospedaliero | Istituto Zooprofilattico Sperimentale della Puglia e della Basilicata | Parisi A., Bianco A., Capozzi L., Del Sambro L., Lippolis A., Notarnicola M., Cipolletta D., Galante D. |
| EPI_ISL_710503 | Laboratorio specialistico UOC Ematologia - Ospedale "San Francesco" - ATS-ASSL Nuoro | Laboratorio specialistico UOC Ematologia - Ospedale "San Francesco" - ATS-ASSL Nuoro | Giovanna Piras |
| EPI_ISL_710542 | National Institute for Infectious Diseases, INMI, "L. Spallanzani" IRCCS | National Institute for Infectious Diseases, INMI, "L. Spallanzani" IRCCS | C.E.M Gruber, B Bartolini, M Rueca, F Messina, E Giombini, A Di Caro, MR Capobianchi |
| EPI_ISL_710543 | National Institute for Infectious Diseases, INMI, "L. Spallanzani" IRCCS | National Institute for Infectious Diseases, INMI, "L. Spallanzani" IRCCS | B Bartolini, C.E.M Gruber, M Rueca, F Messina, E Giombini, MR Capobianchi, A Di Caro |
| EPI_ISL_710544 | National Institute for Infectious Diseases, INMI, "L. Spallanzani" IRCCS | National Institute for Infectious Diseases, INMI, "L. Spallanzani" IRCCS | M Rueca, B Bartolini, C.E.M Gruber, F Messina, E Giombini, A Di Caro, MR Capobianchi |
| EPI_ISL_710545 | National Institute for Infectious Diseases, INMI, "L. Spallanzani" IRCCS | National Institute for Infectious Diseases, INMI, "L. Spallanzani" IRCCS | F Messina, E Giombini, M Rueca, B Bartolini, C.E.M Gruber, MR Capobianchi, A Di Caro |
| EPI_ISL_710546 | National Institute for Infectious Diseases, INMI, "L. Spallanzani" IRCCS | National Institute for Infectious Diseases, INMI, "L. Spallanzani" IRCCS | E Giombini, M Rueca, B Bartolini, C.E.M Gruber, F Messina, A Di Caro, MR Capobianchi |
| EPI_ISL_717978 | Army Medical Center, Scientific Department, Virology Laboratory | Army Medical Center, Scientific Department, Virology Laboratory | Silvia Fillo, Giovanni Faggioni, Riccardo De Santis, Antonella Fortunato, Anna Anselmo, Vanessa Vera Fain, Francesco Giordani, Nino D'Amore, Anella Monte, Marzia Cavalli, Alessandra Amoroso, Stella Lia, Roberta Sorrentino, Rossella Tirelli, Federica Galeano, Annalisa Pelo, Margherita De Santis, Giulia Campoli, Andrea Ciammaruconi, Florigio Lista. |
| EPI_ISL_718262 | National Institute for Infectious Diseases, INMI, "L. Spallanzani" IRCCS | National Institute for Infectious Diseases, INMI, "L. Spallanzani" IRCCS | C.E.M Gruber, F Messina, M Rueca, B Bartolini, E Giombini, MR Capobianchi, A Di Caro |
| EPI_ISL_718263 | National Institute for Infectious Diseases, INMI, "L. Spallanzani" IRCCS | National Institute for Infectious Diseases, INMI, "L. Spallanzani" IRCCS | E Giombini, C.E.M Gruber, M Rueca, B Bartolini, F Messina, A Di Caro, MR Capobianchi |
| EPI_ISL_718264 | National Institute for Infectious Diseases, INMI, "L. Spallanzani" IRCCS | National Institute for Infectious Diseases, INMI, "L. Spallanzani" IRCCS | F Messina, M Rueca, B Bartolini, C.E.M Gruber, E Giombini, MR Capobianchi, A Di Caro |
| EPI_ISL_718265 | National Institute for Infectious Diseases, INMI, "L. Spallanzani" IRCCS | National Institute for Infectious Diseases, INMI, "L. Spallanzani" IRCCS | B Bartolini, M Rueca, C.E.M Gruber, F Messina, E Giombini, A Di Caro, MR Capobianchi |
| EPI_ISL_721624 | National Institute for Infectious Diseases, INMI, "L. Spallanzani" IRCCS | National Institute for Infectious Diseases, INMI, "L. Spallanzani" IRCCS | C.E.M Gruber, B Bartolini, M Rueca, F Messina, E Giombini, A Di Caro, MR Capobianchi |
| EPI_ISL_721625 | National Institute for Infectious Diseases, INMI, "L. Spallanzani" IRCCS | National Institute for Infectious Diseases, INMI, "L. Spallanzani" IRCCS | B Bartolini, C.E.M Gruber, M Rueca, F Messina, E Giombini, MR Capobianchi, A Di Caro |
| EPI_ISL_721626 | National Institute for Infectious Diseases, INMI, "L. Spallanzani" IRCCS | National Institute for Infectious Diseases, INMI, "L. Spallanzani" IRCCS | M Rueca, B Bartolini, C.E.M Gruber, F Messina, E Giombini, A Di Caro, MR Capobianchi |
| EPI_ISL_721627 | National Institute for Infectious Diseases, INMI, "L. Spallanzani" IRCCS | National Institute for Infectious Diseases, INMI, "L. Spallanzani" IRCCS | F Messina, E Giombini, M Rueca, B Bartolini, C.E.M Gruber, MR Capobianchi, A Di Caro |
| EPI_ISL_721628 | National Institute for Infectious Diseases, INMI, "L. Spallanzani" IRCCS | National Institute for Infectious Diseases, INMI, "L. Spallanzani" IRCCS | E Giombini, M Rueca, B Bartolini, C.E.M Gruber, F Messina, A Di Caro, MR Capobianchi |
| EPI_ISL_722851 | I.R.C.C.S. "S. De Bellis" - Ente Ospedaliero | Istituto Zooprofilattico Sperimentale della Puglia e della Basilicata | Parisi A., Bianco A., Capozzi L., Del Sambro L., Lippolis A., Notarnicola M., Manzulli V, Rondinone V., Pace L. |
| EPI_ISL_722852 | I.R.C.C.S. "S. De Bellis" - Ente Ospedaliero | Istituto Zooprofilattico Sperimentale della Puglia e della Basilicata | Parisi A., Bianco A., Capozzi L., Del Sambro L., Lippolis A., Notarnicola M., Cipolletta D., Galante D. |
| EPI_ISL_722853 | I.R.C.C.S. "S. De Bellis" - Ente Ospedaliero | Istituto Zooprofilattico Sperimentale della Puglia e della Basilicata | Parisi A., Bianco A., Capozzi L., Del Sambro L., Lippolis A., Notarnicola M., Manzulli V, Rondinone V., Pace L. |
| EPI_ISL_722854 | I.R.C.C.S. "S. De Bellis" - Ente Ospedaliero | Istituto Zooprofilattico Sperimentale della Puglia e della Basilicata | Parisi A., Bianco A., Capozzi L., Del Sambro L., Lippolis A., Notarnicola M., Cipolletta D., Galante D. |
| EPI_ISL_722855, EPI_ISL_722856, EPI_ISL_722857, EPI_ISL_722858 | Dipartimento di Scienze Biomediche e Oncologia Umana - Azienda Ospedaliero Universitaria Consorziale Policlinico | Istituto Zooprofilattico Sperimentale della Puglia e della Basilicata | Parisi A., Bianco A., Capozzi L., Del Sambro L., Chironna M., Loconsole D. |
| EPI_ISL_722859, EPI_ISL_722860 | I.R.C.C.S. "S. De Bellis" - Ente Ospedaliero | Istituto Zooprofilattico Sperimentale della Puglia e della Basilicata | Parisi A., Bianco A., Capozzi L., Del Sambro L., Lippolis A., Notarnicola M., Manzulli V, Rondinone V., Pace L. |
| EPI_ISL_722861 | I.R.C.C.S. "S. De Bellis" - Ente Ospedaliero | Istituto Zooprofilattico Sperimentale della Puglia e della Basilicata | Parisi A., Bianco A., Capozzi L., Del Sambro L., Lippolis A., Notarnicola M., Cipolletta D., Galante D. |
| EPI_ISL_722862 | I.R.C.C.S. "S. De Bellis" - Ente Ospedaliero | Istituto Zooprofilattico Sperimentale della Puglia e della Basilicata | Parisi A., Bianco A., Capozzi L., Del Sambro L., Lippolis A., Notarnicola M., Manzulli V, Rondinone V., Pace L. |
| EPI_ISL_722863 | I.R.C.C.S. "S. De Bellis" - Ente Ospedaliero | Istituto Zooprofilattico Sperimentale della Puglia e della Basilicata | Parisi A., Bianco A., Capozzi L., Del Sambro L., Lippolis A., Notarnicola M., Cipolletta D., Galante D. |
| EPI_ISL_722864, EPI_ISL_722865 | I.R.C.C.S. "S. De Bellis" - Ente Ospedaliero | Istituto Zooprofilattico Sperimentale della Puglia e della Basilicata | Parisi A., Bianco A., Capozzi L., Del Sambro L., Lippolis A., Notarnicola M., Manzulli V, Rondinone V., Pace L. |
| EPI_ISL_722866 | I.R.C.C.S. "S. De Bellis" - Ente Ospedaliero | Istituto Zooprofilattico Sperimentale della Puglia e della Basilicata | Parisi A., Bianco A., Capozzi L., Del Sambro L., Lippolis A., Notarnicola M., Cipolletta D., Galante D. |
| EPI_ISL_722867, EPI_ISL_722868 | I.R.C.C.S. "S. De Bellis" - Ente Ospedaliero | Istituto Zooprofilattico Sperimentale della Puglia e della Basilicata | Parisi A., Bianco A., Capozzi L., Del Sambro L., Lippolis A., Notarnicola M., Manzulli V, Rondinone V., Pace L. |
| EPI_ISL_722869 | I.R.C.C.S. "S. De Bellis" - Ente Ospedaliero | Istituto Zooprofilattico Sperimentale della Puglia e della Basilicata | Parisi A., Bianco A., Capozzi L., Del Sambro L., Lippolis A., Notarnicola M., Cipolletta D., Galante D. |
| EPI_ISL_722870 | I.R.C.C.S. "S. De Bellis" - Ente Ospedaliero | Istituto Zooprofilattico Sperimentale della Puglia e della Basilicata | Parisi A., Bianco A., Capozzi L., Del Sambro L., Lippolis A., Notarnicola M., Manzulli V, Rondinone V., Pace L. |
| EPI_ISL_722871 | I.R.C.C.S. "S. De Bellis" - Ente Ospedaliero | Istituto Zooprofilattico Sperimentale della Puglia e della Basilicata | Parisi A., Bianco A., Capozzi L., Del Sambro L., Lippolis A., Notarnicola M., Cipolletta D., Galante D. |
| EPI_ISL_722872 | Dipartimento di Scienze Biomediche e Oncologia Umana - Azienda Ospedaliero Universitaria Consorziale Policlinico | Istituto Zooprofilattico Sperimentale della Puglia e della Basilicata | Parisi A., Bianco A., Capozzi L., Del Sambro L., Chironna M., Loconsole D. |

| | | | |
|---|---|---|---|
| EPI_ISL_722873, EPI_ISL_722874, EPI_ISL_722875, EPI_ISL_722876, EPI_ISL_722877, EPI_ISL_722878, EPI_ISL_722879, EPI_ISL_722880, EPI_ISL_722881, EPI_ISL_722882, EPI_ISL_722883, EPI_ISL_722884, EPI_ISL_722885, EPI_ISL_722886, EPI_ISL_722887, EPI_ISL_722888, EPI_ISL_722889, EPI_ISL_722890, EPI_ISL_722891, EPI_ISL_722892, EPI_ISL_722893, EPI_ISL_722894 | | | |
| see above | Istituto Zooprofilattico Sperimentale della Puglia e della Basilicata | Istituto Zooprofilattico Sperimentale della Puglia e della Basilicata | Parisi A., Bianco A., Capozzi L., Del Sambro L., Manzulli V, Rondinone V., Pace L., Cipolletta D., Galante D. |
| EPI_ISL_722895 | I.R.C.C.S. "S. De Bellis" - Ente Ospedaliero | Istituto Zooprofilattico Sperimentale della Puglia e della Basilicata | Parisi A., Bianco A., Capozzi L., Del Sambro L., Lippolis A., Notarnicola M., Cipolletta D., Galante D. |
| EPI_ISL_722896, EPI_ISL_722897 | Dipartimento di Scienze Biomediche e Oncologia Umana - Azienda Ospedaliero Universitaria Consorziale Policlinico | Istituto Zooprofilattico Sperimentale della Puglia e della Basilicata | Parisi A., Bianco A., Capozzi L., Del Sambro L., Chironna M., Loconsole D. |
| EPI_ISL_722898, EPI_ISL_722900 | I.R.C.C.S. "S. De Bellis" - Ente Ospedaliero | Istituto Zooprofilattico Sperimentale della Puglia e della Basilicata | Parisi A., Bianco A., Capozzi L., Del Sambro L., Lippolis A., Notarnicola M., Cipolletta D., Galante D. |
| EPI_ISL_722901, EPI_ISL_722902, EPI_ISL_722903, EPI_ISL_722904, EPI_ISL_722905, EPI_ISL_722906, EPI_ISL_722907, EPI_ISL_722908, EPI_ISL_722909, EPI_ISL_722910, EPI_ISL_722911, EPI_ISL_722913, EPI_ISL_722914, EPI_ISL_722915, EPI_ISL_722916, EPI_ISL_722917, EPI_ISL_722918, EPI_ISL_722919, EPI_ISL_722920, EPI_ISL_722921, EPI_ISL_722922, EPI_ISL_722923, EPI_ISL_722924, EPI_ISL_722925 | | | |
| see above | Istituto Zooprofilattico Sperimentale della Puglia e della Basilicata | Istituto Zooprofilattico Sperimentale della Puglia e della Basilicata | Parisi A., Bianco A., Capozzi L., Del Sambro L., Manzulli V, Rondinone V., Pace L., Cipolletta D., Galante D. |
| EPI_ISL_728279 | National Institute for Infectious Diseases, INMI, "L. Spallanzani" IRCCS | National Institute for Infectious Diseases, INMI, "L. Spallanzani" IRCCS | E Giombini, C.E.M Gruber, M Rueca, B Bartolini, F Messina, A Di Caro, MR Capobianchi |
| EPI_ISL_728280 | National Institute for Infectious Diseases, INMI, "L. Spallanzani" IRCCS | National Institute for Infectious Diseases, INMI, "L. Spallanzani" IRCCS | F Messina, M Rueca, B Bartolini, C.E.M Gruber, E Giombini, MR Capobianchi, A Di Caro |
| EPI_ISL_728281 | National Institute for Infectious Diseases, INMI, "L. Spallanzani" IRCCS | National Institute for Infectious Diseases, INMI, "L. Spallanzani" IRCCS | B Bartolini, M Rueca, C.E.M Gruber, F Messina, E Giombini, A Di Caro, MR Capobianchi |
| EPI_ISL_728282 | National Institute for Infectious Diseases, INMI, "L. Spallanzani" IRCCS | National Institute for Infectious Diseases, INMI, "L. Spallanzani" IRCCS | M. Rueca, C.E.M Gruber, B Bartolini, F Messina, E Giombini, A Di Caro, MR Capobianchi |
| EPI_ISL_728283 | National Institute for Infectious Diseases, INMI, "L. Spallanzani" IRCCS | National Institute for Infectious Diseases, INMI, "L. Spallanzani" IRCCS | B Bartolini, C.E.M Gruber, M Rueca, F Messina, E Giombini, MR Capobianchi, A Di Caro |
| EPI_ISL_728284 | National Institute for Infectious Diseases, INMI, "L. Spallanzani" IRCCS | National Institute for Infectious Diseases, INMI, "L. Spallanzani" IRCCS | M Rueca, B Bartolini, C.E.M Gruber, F Messina, E Giombini, A Di Caro, MR Capobianchi |
| EPI_ISL_728285 | National Institute for Infectious Diseases, INMI, "L. Spallanzani" IRCCS | National Institute for Infectious Diseases, INMI, "L. Spallanzani" IRCCS | F Messina, E Giombini, M Rueca, B Bartolini, C.E.M Gruber, MR Capobianchi, A Di Caro |
| EPI_ISL_728286 | National Institute for Infectious Diseases, INMI, "L. Spallanzani" IRCCS | National Institute for Infectious Diseases, INMI, "L. Spallanzani" IRCCS | E Giombini, M Rueca, B Bartolini, C.E.M Gruber, F Messina, A Di Caro, MR Capobianchi |
| EPI_ISL_728287 | National Institute for Infectious Diseases, INMI, "L. Spallanzani" IRCCS | National Institute for Infectious Diseases, INMI, "L. Spallanzani" IRCCS | C.E.M Gruber, F Messina, M Rueca, B Bartolini, E Giombini, MR Capobianchi, A Di Caro |
| EPI_ISL_730653 | University of Bari, Valenzano, Italy | Istituto Zooprofilattico Sperimentale dell'Abruzzo e del Molise "G. Caporale". | N. Decaro, E. Lorusso, G. Elia, C. Desario, D. Buonavoglia, V., Martella, C. Buonavoglia, A. Lorusso, C. Cammà, V. Curini |
| EPI_ISL_735504 | University of Bari Biomedical Sciences and Human Oncology | University of Bari Biomedical Sciences and Human Oncology | Maria Chironna, Anna Sallustio, Daniela Loconsole |
| EPI_ISL_735505, EPI_ISL_735506 | University of Bari Biomedical Sciences and Human Oncology | University of Bari Biomedical Sciences and Human Oncology | Maria Chironna, Anna Sallustio, Daniela Loconsole, Marisa Accogli |
| EPI_ISL_735510 | National Institute for Infectious Diseases, INMI, "L. Spallanzani" IRCCS | National Institute for Infectious Diseases, INMI, "L. Spallanzani" IRCCS | M. Rueca, C.E.M Gruber, E. Giombini, F. Messina, B. Bartolini, F. Carletti, A. Di Caro, M.R Capobianchi |
| EPI_ISL_735511 | Clinical Pathology and Microbiology, San Gallicano Dermatologic Institute IRCCS | National Institute for Infectious Diseases, INMI, "L. Spallanzani" IRCCS | B. Bartolini, C.E.M Gruber, F. Messina, E. Giombini, M. Rueca, F. Carletti, F. Pimpinelli, F. Ensoli, M.R. Capobianchi, A. Di Caro |
| EPI_ISL_736777, EPI_ISL_736778, EPI_ISL_736779, EPI_ISL_736780, EPI_ISL_736781, EPI_ISL_736782, EPI_ISL_736783, EPI_ISL_736784, EPI_ISL_736785, EPI_ISL_736786, EPI_ISL_736787, EPI_ISL_736788, EPI_ISL_736789, EPI_ISL_736790, EPI_ISL_736791, EPI_ISL_736792, EPI_ISL_736793, EPI_ISL_736794, EPI_ISL_736795, EPI_ISL_736796, EPI_ISL_736797, EPI_ISL_736798, EPI_ISL_736799, EPI_ISL_736800, EPI_ISL_736801, EPI_ISL_736802, EPI_ISL_736803, EPI_ISL_736804, EPI_ISL_736805, EPI_ISL_736806, EPI_ISL_736807, EPI_ISL_736808, EPI_ISL_736809, EPI_ISL_736810, EPI_ISL_736811, EPI_ISL_736812, EPI_ISL_736813, EPI_ISL_736814, EPI_ISL_736815, EPI_ISL_736816, EPI_ISL_736817, EPI_ISL_736818, EPI_ISL_736819, EPI_ISL_736820, EPI_ISL_736821, EPI_ISL_736822, EPI_ISL_736823, EPI_ISL_736824, EPI_ISL_736825, EPI_ISL_736826, EPI_ISL_736827, EPI_ISL_736828, EPI_ISL_736829, EPI_ISL_736830, EPI_ISL_736831, EPI_ISL_736832, EPI_ISL_736833, EPI_ISL_736834, EPI_ISL_736835, EPI_ISL_736836, EPI_ISL_736837, EPI_ISL_736838, EPI_ISL_736839, EPI_ISL_736840, EPI_ISL_736841, EPI_ISL_736842, EPI_ISL_736843, EPI_ISL_736844, EPI_ISL_736845, EPI_ISL_736846, EPI_ISL_736847, EPI_ISL_736848, EPI_ISL_736849, EPI_ISL_736850, EPI_ISL_736851, EPI_ISL_736852, EPI_ISL_736853, EPI_ISL_736854, EPI_ISL_736855, EPI_ISL_736856, EPI_ISL_736857, EPI_ISL_736858, EPI_ISL_736859, EPI_ISL_736860, EPI_ISL_736861, EPI_ISL_736862, EPI_ISL_736863, EPI_ISL_736864, EPI_ISL_736865, EPI_ISL_736866, EPI_ISL_736867, EPI_ISL_736868, EPI_ISL_736869, EPI_ISL_736870, EPI_ISL_736871, EPI_ISL_736872, EPI_ISL_736873, EPI_ISL_736874, EPI_ISL_736875, EPI_ISL_736876, EPI_ISL_736877, EPI_ISL_736878, EPI_ISL_736879, EPI_ISL_736880, EPI_ISL_736881, EPI_ISL_736882, EPI_ISL_736883, EPI_ISL_736884, EPI_ISL_736885, EPI_ISL_736886, EPI_ISL_736887, EPI_ISL_736888, EPI_ISL_736889, EPI_ISL_736890 | | | |
| see above | Istituto Zooprofilattico Sperimentale del Mezzogiorno | TIGEM | Antonio Grimaldi, Patrizia Annunziata, Francesco Panariello, Biancamaria Pierri, Valentina Bouche, Chiara Colantuono, Maria Concetta Cuomo, Denise Di Concilio, Lucio Di Filippo, Anna Manfredi, Marcello Salvi, Antonio Limone, Pellegrino Cerino, Andrea Ballabio, Davide Cacchiarelli. |
| EPI_ISL_736996 | Istituto Zooprofilattico Sperimentale del Mezzogiorno | National Institute for Infectious Diseases, INMI, "L. Spallanzani" IRCCS | E. Giombini, C.E.M Gruber, F. Messina, M. Rueca, B. Bartolini, F. Carletti, P. Cerino, B. Pierri, C. Buonerba, D. Di Concilio, M.C. Cuomo, A. Di Caro, M.R Capobianchi |
| EPI_ISL_736997 | Istituto Zooprofilattico Sperimentale del Mezzogiorno | National Institute for Infectious Diseases, INMI, "L. Spallanzani" IRCCS | F. Messina, C.E.M Gruber, M. Rueca, B. Bartolini, E. Giombini, F. Carletti, P. Cerino, B. Pierri, C. Buonerba, D. Di Concilio, M.C. Cuomo, M.R. Capobianchi, A. Di Caro |
| EPI_ISL_738044 | SIESP CHIETI - DRIVE IN LANCIANO | Istituto Zooprofilattico Sperimentale dell'Abruzzo e Molise "G. Caporale" | Lorusso A, Marcacci M, Di Domenico M, Ancora M, Curini V, Mangone I, Rinaldi A, Di Pasquale A, Cammà C, Puglia I, Savini G |
| EPI_ISL_738045 | SIESP DIPARTIMENTO DI PREVENZIONE CHIETI | Istituto Zooprofilattico Sperimentale dell'Abruzzo e Molise "G. Caporale" | Lorusso A, Marcacci M, Di Domenico M, Ancora M, Curini V, Mangone I, Rinaldi A, Di Pasquale A, Cammà C, Puglia I, Savini G |
| EPI_ISL_738046, EPI_ISL_738047, EPI_ISL_738048 | SIESP CHIETI - DRIVE IN ORTONA | Istituto Zooprofilattico Sperimentale dell'Abruzzo e Molise "G. Caporale" | Lorusso A, Marcacci M, Di Domenico M, Ancora M, Curini V, Mangone I, Rinaldi A, Di Pasquale A, Cammà C, Puglia I, Savini G |
| EPI_ISL_738121, EPI_ISL_738122, EPI_ISL_738123, EPI_ISL_738124, EPI_ISL_738125, EPI_ISL_738126, EPI_ISL_738127, EPI_ISL_738128, EPI_ISL_738129, EPI_ISL_738130, EPI_ISL_738131, EPI_ISL_738132 | | | |
| see above | IZSM-U.O.C. Virologia | Istituto Zooprofilattico Sperimentale del Mezzogiorno | Maurizio Viscardi, Lorena Cardillo, Giovanna Fusco |
| EPI_ISL_738144 | University of Bari Biomedical Sciences and Human Oncology | University of Bari Biomedical Sciences and Human Oncology | Maria Chironna, Anna Sallustio, Daniela Loconsole, Marisa Accogli |
| EPI_ISL_738147, EPI_ISL_738194, EPI_ISL_738243 | Microbiology and Virology Unit, Florence Careggi University Hospital | Microbiology and Virology Unit, Florence Careggi University Hospital | Vincenzo Di Pilato, Marco Coppi, Alberto Antonelli, Simona Pollini, Gian Maria Rossolini |
| EPI_ISL_745192 | Ospedale Di Venere | Istituto Zooprofilattico Sperimentale della Puglia e della Basilicata | Parisi A., Capozzi L., Del Sambro L., Bianco A., Chiara M., Pesole G., De Sabato L., Iacobellis M. |
| EPI_ISL_745193 | Ospedale Di Venere | Istituto Zooprofilattico Sperimentale della Puglia e della Basilicata | Parisi A., Bianco A., Capozzi L., Del Sambro L., Chiara M., De Sabato L., Pesole G., Iacobellis M |
| EPI_ISL_746826 | National Institute for Infectious Diseases, INMI, "L. Spallanzani" IRCCS | National Institute for Infectious Diseases, INMI, "L. Spallanzani" IRCCS | C.E.M Gruber, B Bartolini, M Rueca, F Messina, E Giombini, A Di Caro, MR Capobianchi |

| | | | |
|---|---|---|---|
| EPI_ISL_746827 | National Institute for Infectious Diseases, INMI, "L. Spallanzani" IRCCS | National Institute for Infectious Diseases, INMI, "L. Spallanzani" IRCCS | B Bartolini, C.E.M Gruber, M Rueca, F Messina, E Giombini, MR Capobianchi, A Di Caro |
| EPI_ISL_746828 | National Institute for Infectious Diseases, INMI, "L. Spallanzani" IRCCS | National Institute for Infectious Diseases, INMI, "L. Spallanzani" IRCCS | M Rueca, B Bartolini, C.E.M Gruber, F Messina, E Giombini, A Di Caro, MR Capobianchi |
| EPI_ISL_746829 | National Institute for Infectious Diseases, INMI, "L. Spallanzani" IRCCS | National Institute for Infectious Diseases, INMI, "L. Spallanzani" IRCCS | F Messina, E Giombini, M Rueca, B Bartolini, C.E.M Gruber, MR Capobianchi, A Di Caro |
| EPI_ISL_746830 | National Institute for Infectious Diseases, INMI, "L. Spallanzani" IRCCS | National Institute for Infectious Diseases, INMI, "L. Spallanzani" IRCCS | E Giombini, M Rueca, B Bartolini, C.E.M Gruber, F Messina, A Di Caro, MR Capobianchi |
| EPI_ISL_746831 | National Institute for Infectious Diseases, INMI, "L. Spallanzani" IRCCS | National Institute for Infectious Diseases, INMI, "L. Spallanzani" IRCCS | C.E.M Gruber, F Messina, M Rueca, B Bartolini, E Giombini, MR Capobianchi, A Di Caro |
| EPI_ISL_746832 | National Institute for Infectious Diseases, INMI, "L. Spallanzani" IRCCS | National Institute for Infectious Diseases, INMI, "L. Spallanzani" IRCCS | E Giombini, C.E.M Gruber, M Rueca, B Bartolini, F Messina, A Di Caro, MR Capobianchi |
| EPI_ISL_746833 | National Institute for Infectious Diseases, INMI, "L. Spallanzani" IRCCS | National Institute for Infectious Diseases, INMI, "L. Spallanzani" IRCCS | F Messina, M Rueca, B Bartolini, C.E.M Gruber, E Giombini, MR Capobianchi, A Di Caro |
| EPI_ISL_746834 | National Institute for Infectious Diseases, INMI, "L. Spallanzani" IRCCS | National Institute for Infectious Diseases, INMI, "L. Spallanzani" IRCCS | B Bartolini, M Rueca, C.E.M Gruber, F Messina, E Giombini, A Di Caro, MR Capobianchi |
| EPI_ISL_746835 | National Institute for Infectious Diseases, INMI, "L. Spallanzani" IRCCS | National Institute for Infectious Diseases, INMI, "L. Spallanzani" IRCCS | M Rueca, E Giombini, B Bartolini, C.E.M Gruber, F Messina, A Di Caro, MR Capobianchi |
| EPI_ISL_747459, EPI_ISL_747460, EPI_ISL_747461, EPI_ISL_747462, EPI_ISL_747463, EPI_ISL_747464 | Ospedale San Bonifacio | Istituto Zooprofilattico Sperimentale delle Venezie | Adelaide Milani, Alessia Schivo, Annalisa Salviato, Erika Giorgia Quaranta, Ambra Pastori, Bianca Zecchin, Alice Fusaro, Isabella Monne, Calogero Terregino, Antonia Ricci |
| EPI_ISL_747465, EPI_ISL_747466, EPI_ISL_747467, EPI_ISL_747468, EPI_ISL_747469, EPI_ISL_747470 | Ospedale Mater Salutis | Istituto Zooprofilattico Sperimentale delle Venezie | Adelaide Milani, Alessia Schivo, Annalisa Salviato, Erika Giorgia Quaranta, Ambra Pastori, Bianca Zecchin, Alice Fusaro, Isabella Monne, Calogero Terregino, Antonia Ricci |
| EPI_ISL_747471, EPI_ISL_747472, EPI_ISL_747473, EPI_ISL_747474, EPI_ISL_747475, EPI_ISL_747476 | ULSS9 Distretto di Bussolengo | Istituto Zooprofilattico Sperimentale delle Venezie | Adelaide Milani, Alessia Schivo, Annalisa Salviato, Erika Giorgia Quaranta, Ambra Pastori, Bianca Zecchin, Alice Fusaro, Isabella Monne, Calogero Terregino, Antonia Ricci |
| EPI_ISL_747477, EPI_ISL_747478, EPI_ISL_747479, EPI_ISL_747480, EPI_ISL_747481 | ULSS 5 Polesana | Istituto Zooprofilattico Sperimentale delle Venezie | Adelaide Milani, Alessia Schivo, Annalisa Salviato, Erika Giorgia Quaranta, Ambra Pastori, Bianca Zecchin, Alice Fusaro, Isabella Monne, Calogero Terregino, Antonia Ricci |
| EPI_ISL_747482, EPI_ISL_747483 | Ospedale San Bonifacio | Istituto Zooprofilattico Sperimentale delle Venezie | Adelaide Milani, Alessia Schivo, Annalisa Salviato, Erika Giorgia Quaranta, Ambra Pastori, Bianca Zecchin, Alice Fusaro, Isabella Monne, Calogero Terregino, Antonia Ricci |
| EPI_ISL_747484, EPI_ISL_747485 | Ospedale Mater Salutis | Istituto Zooprofilattico Sperimentale delle Venezie | Adelaide Milani, Alessia Schivo, Annalisa Salviato, Erika Giorgia Quaranta, Ambra Pastori, Bianca Zecchin, Alice Fusaro, Isabella Monne, Calogero Terregino, Antonia Ricci |
| EPI_ISL_747486, EPI_ISL_747487, EPI_ISL_747488, EPI_ISL_747489 | ULSS 5 Polesana | Istituto Zooprofilattico Sperimentale delle Venezie | Adelaide Milani, Alessia Schivo, Annalisa Salviato, Erika Giorgia Quaranta, Ambra Pastori, Bianca Zecchin, Alice Fusaro, Isabella Monne, Calogero Terregino, Antonia Ricci |
| EPI_ISL_751318, EPI_ISL_751319, EPI_ISL_751320, EPI_ISL_751321, EPI_ISL_751322, EPI_ISL_751323, EPI_ISL_751324, EPI_ISL_751325, EPI_ISL_751326, EPI_ISL_751327, EPI_ISL_751328, EPI_ISL_751329, EPI_ISL_751330, EPI_ISL_751331, EPI_ISL_751332, EPI_ISL_751333, EPI_ISL_751334, EPI_ISL_751335, EPI_ISL_751336, EPI_ISL_751337, EPI_ISL_751338, EPI_ISL_751339, EPI_ISL_751340, EPI_ISL_751341, EPI_ISL_751342, EPI_ISL_751343, EPI_ISL_751344, EPI_ISL_751345, EPI_ISL_751346, EPI_ISL_751347, EPI_ISL_751348, EPI_ISL_751349, EPI_ISL_751350, EPI_ISL_751351, EPI_ISL_751352, EPI_ISL_751353, EPI_ISL_751354, EPI_ISL_751355, EPI_ISL_751356, EPI_ISL_751357, EPI_ISL_751358, EPI_ISL_751359, EPI_ISL_751360, EPI_ISL_751361, EPI_ISL_751362, EPI_ISL_751363, EPI_ISL_751364, EPI_ISL_751365, EPI_ISL_751366, EPI_ISL_751367, EPI_ISL_751368, EPI_ISL_751369, EPI_ISL_751370, EPI_ISL_751371, EPI_ISL_751372, EPI_ISL_751373, EPI_ISL_751374, EPI_ISL_751375, EPI_ISL_751376, EPI_ISL_751377, EPI_ISL_751378, EPI_ISL_751379, EPI_ISL_751380, EPI_ISL_751381, EPI_ISL_751382, EPI_ISL_751383, EPI_ISL_751384, EPI_ISL_751385, EPI_ISL_751386, EPI_ISL_751387, EPI_ISL_751388, EPI_ISL_751389, EPI_ISL_751390, EPI_ISL_751391, EPI_ISL_751392, EPI_ISL_751393, EPI_ISL_751394, EPI_ISL_751395, EPI_ISL_751396, EPI_ISL_751397, EPI_ISL_751398, EPI_ISL_751399, EPI_ISL_751400, EPI_ISL_751401, EPI_ISL_751402, EPI_ISL_751403, EPI_ISL_751404, EPI_ISL_751405, EPI_ISL_751406, EPI_ISL_751407, EPI_ISL_751408, EPI_ISL_751409, EPI_ISL_751410, EPI_ISL_751411, EPI_ISL_751412, EPI_ISL_751413, EPI_ISL_751414, EPI_ISL_751415, EPI_ISL_751416, EPI_ISL_751417, EPI_ISL_751418, EPI_ISL_751419, EPI_ISL_751420, EPI_ISL_751421, EPI_ISL_751422, EPI_ISL_751423, EPI_ISL_751424, EPI_ISL_751425, EPI_ISL_751426, EPI_ISL_751427, EPI_ISL_751428, EPI_ISL_751429, EPI_ISL_751430, EPI_ISL_751431, EPI_ISL_751432, EPI_ISL_751433, EPI_ISL_751434, EPI_ISL_751435, EPI_ISL_751436, EPI_ISL_751437, EPI_ISL_751438, EPI_ISL_751439, EPI_ISL_751440, EPI_ISL_751441, EPI_ISL_751442, EPI_ISL_751443, EPI_ISL_751444, EPI_ISL_751445, EPI_ISL_751446, EPI_ISL_751447 | | | |
| see above | IRCCS Sacro Cuore Don Calabria Hospital, Department of Infectious, Tropical Diseases & Microbiology | University of Verona, Department of Biotechnology | Antonio Mori, Michela Deiana, Elena Pomari, Chiara Piubelli; Giulia Lopatriello, Luca Marcolungo, Cristina Beltrami, Chiara Degli Esposti, Emanuela Cosentino, Massimo Delledonne |
| EPI_ISL_755572, EPI_ISL_755573, EPI_ISL_755574 | Center of Advanced Studies and Technology, CAST | Center of Advanced Studies and Technology, CAST | Ferrante,R., Mandatori,D., De Fabritiis,S. |
| EPI_ISL_763069, EPI_ISL_763071, EPI_ISL_763072, EPI_ISL_763073 | Istituto Zooprofilattico Sperimentale dell' Umbria e delle Marche -Togo Rosati | Istituto Superiore di Sanità | Massimo Biagetti , Monica Giammarioli, Luca De Sabato, Gabriele Vaccari, Ilaria Di Bartolo, Giovanni Ianiro |
| EPI_ISL_763078, EPI_ISL_763079, EPI_ISL_763080, EPI_ISL_763081, EPI_ISL_763082, EPI_ISL_763083, EPI_ISL_763084, EPI_ISL_763085 | Microbiologia e Virologia | Istituto Zooprofilattico Sperimentale delle Venezie | Adelaide Milani, Alessia Schivo, Annalisa Salviato, Erika Giorgia Quaranta, Ambra Pastori, Bianca Zecchin, Alice Fusaro, Isabella Monne, Calogero Terregino, Antonia Ricci |
| EPI_ISL_763094, EPI_ISL_763095, EPI_ISL_763096, EPI_ISL_763097, EPI_ISL_763098, EPI_ISL_763138, EPI_ISL_763318, EPI_ISL_763320, EPI_ISL_763322, EPI_ISL_763323, EPI_ISL_763324, EPI_ISL_763325, EPI_ISL_763326, EPI_ISL_763327, EPI_ISL_763328, EPI_ISL_763329, EPI_ISL_763330 | | | |
| see above | Istituto Zooprofilattico Sperimentale dell' Umbria e delle Marche -Togo Rosati | Istituto Superiore di Sanità | Massimo Biagetti , Monica Giammarioli, Luca De Sabato, Gabriele Vaccari, Ilaria Di Bartolo, Giovanni Ianiro |
| EPI_ISL_765567 | National Institute for Infectious Diseases, INMI, "L. Spallanzani" IRCCS | National Institute for Infectious Diseases, INMI, "L. Spallanzani" IRCCS | E Giombini, C.E.M Gruber, M Rueca, B Bartolini, O Butera, F Messina, A Di Caro, G Parisi, MR Capobianchi |
| EPI_ISL_765568 | National Institute for Infectious Diseases, INMI, "L. Spallanzani" IRCCS | National Institute for Infectious Diseases, INMI, "L. Spallanzani" IRCCS | O Butera, C.E.M Gruber, F Messina, M Rueca, B Bartolini, E Giombini, MR Capobianchi, A Di Caro |
| EPI_ISL_765569 | National Institute for Infectious Diseases, INMI, "L. Spallanzani" IRCCS | National Institute for Infectious Diseases, INMI, "L. Spallanzani" IRCCS | B Bartolini, M Rueca, O Butera, C.E.M Gruber, F Messina, E Giombini, A Di Caro, MR Capobianchi |
| EPI_ISL_765570 | National Institute for Infectious Diseases, INMI, "L. Spallanzani" IRCCS | National Institute for Infectious Diseases, INMI, "L. Spallanzani" IRCCS | M. Rueca, C.E.M Gruber, B Bartolini, O Butera, F Messina, E Giombini, A Di Caro, MR Capobianchi |
| EPI_ISL_765571 | National Institute for Infectious Diseases, INMI, "L. Spallanzani" IRCCS | National Institute for Infectious Diseases, INMI, "L. Spallanzani" IRCCS | E Giombini, B Bartolini, O Butera, C.E.M Gruber, M Rueca, F Messina, MR Capobianchi, A Di Caro |
| EPI_ISL_765572 | National Institute for Infectious Diseases, INMI, "L. Spallanzani" IRCCS | National Institute for Infectious Diseases, INMI, "L. Spallanzani" IRCCS | C.E.M Gruber, M Rueca, B Bartolini, F Messina, E Giombini,O Butera, A Di Caro, MR Capobianchi |
| EPI_ISL_765573 | National Institute for Infectious Diseases, INMI, "L. Spallanzani" IRCCS | National Institute for Infectious Diseases, INMI, "L. Spallanzani" IRCCS | F Messina, E Giombini, M Rueca, B Bartolini, C.E.M Gruber, MR Capobianchi, O Butera, A Di Caro |
| EPI_ISL_766571, EPI_ISL_766572, EPI_ISL_766573 | ULSS 7 Pedemontana - Distretto 1 | Istituto Zooprofilattico Sperimentale delle Venezie | Adelaide Milani, Alessia Schivo, Annalisa Salviato, Erika Giorgia Quaranta, Ambra Pastori, Bianca Zecchin, Alice Fusaro, Isabella Monne, Calogero Terregino, Antonia Ricci |

| | | | |
|---|---|---|---|
| EPI_ISL_766574, EPI_ISL_766575, EPI_ISL_766576, EPI_ISL_766577, EPI_ISL_766578, EPI_ISL_766579 | ULSS 8 Berica | Istituto Zooprofilattico Sperimentale delle Venezie | Adelaide Milani, Alessia Schivo, Annalisa Salviato, Erika Giorgia Quaranta, Ambra Pastori, Bianca Zecchin, Alice Fusaro, Isabella Monne, Calogero Terregino, Antonia Ricci |
| EPI_ISL_766580, EPI_ISL_766581 | ULSS 2 Treviso | Istituto Zooprofilattico Sperimentale delle Venezie | Adelaide Milani, Alessia Schivo, Annalisa Salviato, Erika Giorgia Quaranta, Ambra Pastori, Bianca Zecchin, Alice Fusaro, Isabella Monne, Calogero Terregino, Antonia Ricci |
| EPI_ISL_767013, EPI_ISL_767043 | University of Bari Biomedical Sciences and Human Oncology | University of Bari Biomedical Sciences and Human Oncology | Maria Chironna, Anna Sallustio, Daniela Loconsole, Marisa Accogli |
| EPI_ISL_775239, EPI_ISL_775240, EPI_ISL_775243, EPI_ISL_775244, EPI_ISL_775246, EPI_ISL_775257, EPI_ISL_775259, EPI_ISL_775261 | INT Fondazione Pascale | INT Fondazione Pascale | INT Fondazione Pascale |
| EPI_ISL_776879, EPI_ISL_776880, EPI_ISL_776881, EPI_ISL_776882, EPI_ISL_776883, EPI_ISL_776884, EPI_ISL_776885, EPI_ISL_776886, EPI_ISL_776887, EPI_ISL_776888, EPI_ISL_776889, EPI_ISL_776890, EPI_ISL_776891, EPI_ISL_776892, EPI_ISL_776893, EPI_ISL_776894, EPI_ISL_776895, EPI_ISL_776896, EPI_ISL_776897, EPI_ISL_776898, EPI_ISL_776899, EPI_ISL_776900, EPI_ISL_776901, EPI_ISL_776902, EPI_ISL_776903, EPI_ISL_776904, EPI_ISL_776905, EPI_ISL_776906, EPI_ISL_776907, EPI_ISL_776908, EPI_ISL_776909, EPI_ISL_776910, EPI_ISL_776911, EPI_ISL_776912, EPI_ISL_776913, EPI_ISL_776914, EPI_ISL_776915, EPI_ISL_776916, EPI_ISL_776917, EPI_ISL_776918, EPI_ISL_776919, EPI_ISL_776920, EPI_ISL_776921, EPI_ISL_776922, EPI_ISL_776923, EPI_ISL_776924, EPI_ISL_776925, EPI_ISL_776926, EPI_ISL_776927, EPI_ISL_776928, EPI_ISL_776929, EPI_ISL_776930, EPI_ISL_776931, EPI_ISL_776932, EPI_ISL_776933, EPI_ISL_776934, EPI_ISL_776935, EPI_ISL_776936, EPI_ISL_776937, EPI_ISL_776938, EPI_ISL_776939, EPI_ISL_776940, EPI_ISL_776941, EPI_ISL_776942, EPI_ISL_776943, EPI_ISL_776944, EPI_ISL_776945, EPI_ISL_776946, EPI_ISL_776947, EPI_ISL_776948, EPI_ISL_776949, EPI_ISL_776950, EPI_ISL_776951, EPI_ISL_776952, EPI_ISL_776953, EPI_ISL_776954, EPI_ISL_776955, EPI_ISL_776956, EPI_ISL_776957, EPI_ISL_776958, EPI_ISL_776959, EPI_ISL_776960, EPI_ISL_776961, EPI_ISL_776962, EPI_ISL_776963, EPI_ISL_776964, EPI_ISL_776965, EPI_ISL_776966, EPI_ISL_776967, EPI_ISL_776968, EPI_ISL_776969, EPI_ISL_776971, EPI_ISL_776972, EPI_ISL_776973, EPI_ISL_776974, EPI_ISL_776975, EPI_ISL_776976, EPI_ISL_776977, EPI_ISL_776978, EPI_ISL_776979, EPI_ISL_776980, EPI_ISL_776981, EPI_ISL_776982, EPI_ISL_776983, EPI_ISL_776984, EPI_ISL_776985, EPI_ISL_776986, EPI_ISL_776987, EPI_ISL_776988, EPI_ISL_776989, EPI_ISL_776990, EPI_ISL_776991, EPI_ISL_776992, EPI_ISL_776993, EPI_ISL_776995, EPI_ISL_776996, EPI_ISL_776997, EPI_ISL_776998, EPI_ISL_776999, EPI_ISL_777000, EPI_ISL_777001, EPI_ISL_777002, EPI_ISL_777003, EPI_ISL_777004, EPI_ISL_777005, EPI_ISL_777006, EPI_ISL_777007, EPI_ISL_777008, EPI_ISL_777009, EPI_ISL_778646, EPI_ISL_778647, EPI_ISL_778648, EPI_ISL_778649, EPI_ISL_778650, EPI_ISL_778651, EPI_ISL_778652, EPI_ISL_778653, EPI_ISL_778654, EPI_ISL_778655, EPI_ISL_778656, EPI_ISL_778657, EPI_ISL_778658, EPI_ISL_778659, EPI_ISL_778660, EPI_ISL_778661, EPI_ISL_778662, EPI_ISL_778663, EPI_ISL_778664, EPI_ISL_778665, EPI_ISL_778666, EPI_ISL_778667, EPI_ISL_778668, EPI_ISL_778669, EPI_ISL_778670, EPI_ISL_778671, EPI_ISL_778672, EPI_ISL_778673, EPI_ISL_778674, EPI_ISL_778675, EPI_ISL_778676, EPI_ISL_778677, EPI_ISL_778678, EPI_ISL_778679, EPI_ISL_778680, EPI_ISL_778681, EPI_ISL_778682, EPI_ISL_778683, EPI_ISL_778684, EPI_ISL_778685, EPI_ISL_778686, EPI_ISL_778687, EPI_ISL_778688, EPI_ISL_778689, EPI_ISL_778690, EPI_ISL_778691, EPI_ISL_778692, EPI_ISL_778693, EPI_ISL_778694, EPI_ISL_778695, EPI_ISL_778696, EPI_ISL_778697, EPI_ISL_778698, EPI_ISL_778699, EPI_ISL_778700, EPI_ISL_778701, EPI_ISL_778702, EPI_ISL_778703, EPI_ISL_778704, EPI_ISL_778705, EPI_ISL_778706, EPI_ISL_778707, EPI_ISL_778708, EPI_ISL_778709, EPI_ISL_778710, EPI_ISL_778711, EPI_ISL_778712, EPI_ISL_778713, EPI_ISL_778714, EPI_ISL_778715, EPI_ISL_778716, EPI_ISL_778717, EPI_ISL_778718, EPI_ISL_778719, EPI_ISL_778720, EPI_ISL_778721, EPI_ISL_778722, EPI_ISL_778723, EPI_ISL_778724, EPI_ISL_778725, EPI_ISL_778726, EPI_ISL_778727, EPI_ISL_778728, EPI_ISL_778729, EPI_ISL_778730, EPI_ISL_778731, EPI_ISL_778732, EPI_ISL_778733, EPI_ISL_778734, EPI_ISL_778735, EPI_ISL_778736, EPI_ISL_778737, EPI_ISL_778738, EPI_ISL_778739, EPI_ISL_778740, EPI_ISL_778741, EPI_ISL_778742, EPI_ISL_778743, EPI_ISL_778744, EPI_ISL_778745, EPI_ISL_778746, EPI_ISL_778747, EPI_ISL_778748, EPI_ISL_778749, EPI_ISL_778750, EPI_ISL_778751, EPI_ISL_778752, EPI_ISL_778753, EPI_ISL_778754, EPI_ISL_778755, EPI_ISL_778756, EPI_ISL_778757, EPI_ISL_778758, EPI_ISL_778759, EPI_ISL_778760, EPI_ISL_778761, EPI_ISL_778762, EPI_ISL_778763, EPI_ISL_778764, EPI_ISL_778765, EPI_ISL_778766, EPI_ISL_778767, EPI_ISL_778768, EPI_ISL_778769, EPI_ISL_778770, EPI_ISL_778771, EPI_ISL_778772, EPI_ISL_778773, EPI_ISL_778774, EPI_ISL_778775, EPI_ISL_778776, EPI_ISL_778777, EPI_ISL_778778, EPI_ISL_778779, EPI_ISL_778780, EPI_ISL_778781, EPI_ISL_778782, EPI_ISL_778783, EPI_ISL_778784, EPI_ISL_778785, EPI_ISL_778786, EPI_ISL_778787, EPI_ISL_778788, EPI_ISL_778789, EPI_ISL_778790, EPI_ISL_778791, EPI_ISL_778792, EPI_ISL_778793, EPI_ISL_778794, EPI_ISL_778795, EPI_ISL_778796, EPI_ISL_778797, EPI_ISL_778798, EPI_ISL_778799, EPI_ISL_778800, EPI_ISL_778801, EPI_ISL_778802, EPI_ISL_778803, EPI_ISL_778804, EPI_ISL_778805, EPI_ISL_778806, EPI_ISL_778807, EPI_ISL_778808, EPI_ISL_778809, EPI_ISL_778810, EPI_ISL_778811, EPI_ISL_778812, EPI_ISL_778813, EPI_ISL_778814, EPI_ISL_778815, EPI_ISL_778816, EPI_ISL_778817 | | | |
| see above | Istituto Zooprofilattico Sperimentale del Mezzogiorno | TIGEM | Antonio Grimaldi, Patrizia Annunziata, Francesco Panariello, Biancamaria Pierri, Valentina Bouche, Chiara Colantuono, Maria Concetta Cuomo, Denise Di Concilio, Lucio Di Filippo, Anna Manfredi, Marcello Salvi, Antonio Limone, Pellegrino Cerino, Andrea Ballabio, Davide Cacchiarelli. |
| EPI_ISL_778860 | Laboratory of Infectious Diseases, Department of Biomedical and Clinical Sciences L. Sacco, University of Milan | Laboratory of Infectious Diseases, Department of Biomedical and Clinical Sciences L. Sacco, University of Milan | Alessia Lai, Annalisa Bergna, Carla Della Ventura, Claudia Balotta, Massimo Galli, Gianguglielmo Zehender on behalf of SARS-CoV-2 ITALIAN RESEARCH ENTERPRISE-(SCIRE) Collaborative Group |
| EPI_ISL_778868 | Virology Lab, Ospedali Riuniti di Ancona | Dipartimento di Scienze Biomediche e Cliniche, L.Sacco, Università di Milano | Alessia Lai, Annalisa Bergna, Gianni Zehender, Claudia Balotta, Sara Caucci, Laura Di Sante, Roberta Longo, Sofia Maria Luigia Tiano, Massimo Galli, Patrizia Bagnarelli, Stefano Menzo |
| EPI_ISL_778869 | Virology Lab, Ospedali Riuniti, Ancona | Dipartimento di Scenze Biomediche e Cliniche, L. Sacco, Università di Milano | Alessia Lai, Annalisa Bergna, Gianni Zehender, Claudia Balotta, Sara Caucci, Laura Di Sante, Roberta Longo, Sofia Maria Luigia Tiano, Massimo Galli, Patrizia Bagnarelli, Stefano Menzo |
| EPI_ISL_779704, EPI_ISL_779707, EPI_ISL_779709, EPI_ISL_779712, EPI_ISL_779713, EPI_ISL_779714, EPI_ISL_780375, EPI_ISL_780376, EPI_ISL_780378, EPI_ISL_780381, EPI_ISL_780411, EPI_ISL_780412, EPI_ISL_780413 | | | |
| see above | Laboratory of Infectious Diseases, Department of Biomedical and Clinical Sciences L. Sacco, University of Milan | Laboratory of Infectious Diseases, Department of Biomedical and Clinical Sciences L. Sacco, University of Milan | Alessia Lai, Annalisa Bergna, Carla Della Ventura, Claudia Balotta, Massimo Galli, Gianguglielmo Zehender on behalf of SARS-CoV-2 ITALIAN RESEARCH ENTERPRISE-(SCIRE) Collaborative Group |
| EPI_ISL_788943, EPI_ISL_788944, EPI_ISL_788945, EPI_ISL_788946, EPI_ISL_788947, EPI_ISL_788948, EPI_ISL_788949, EPI_ISL_788950, EPI_ISL_788951, EPI_ISL_788952, EPI_ISL_788953, EPI_ISL_788954, EPI_ISL_788955, EPI_ISL_788956, EPI_ISL_788957, EPI_ISL_788958, EPI_ISL_788959, EPI_ISL_788960, EPI_ISL_788961, EPI_ISL_788962, EPI_ISL_788963, EPI_ISL_788964, EPI_ISL_788965, EPI_ISL_788966, EPI_ISL_788967, EPI_ISL_788968, EPI_ISL_788969, EPI_ISL_788970, EPI_ISL_788971, EPI_ISL_788972, EPI_ISL_788973, EPI_ISL_788974, EPI_ISL_788975, EPI_ISL_788976, EPI_ISL_788977, EPI_ISL_788978 | | | |
| see above | Ospedale "Di Venere" | Beaconlab (Bioinformatics, Evolution and Comparative Genomics lab), Dept of Biosciences, University on Milan | Iacobellis M, d'Avenia M, Piluscio R, Parisi A, Chiara M, Manzari C, Pesole G |
| EPI_ISL_794745 | Istituto Zooprofilattico Sperimentale della Puglia e della Basilicata | Istituto Zooprofilattico Sperimentale della Puglia e della Basilicata | Parisi A., Bianco A., Capozzi L., Del Sambro L., Manzulli V, Rondinone V., Pace L., Cipolletta D., Galante D. |
| EPI_ISL_794746 | SISP ASL BA AREA SUD | Istituto Zooprofilattico Sperimentale della Puglia e della Basilicata | Parisi A., Bianco A., Capozzi L., Del Sambro L., Manzulli V, Rondinone V., Pace L., Cipolletta D., Galante D. |
| EPI_ISL_794747 | Ospedale Vito Fazzi | Istituto Zooprofilattico Sperimentale della Puglia e della Basilicata | Parisi A., Bianco A., Capozzi L., Del Sambro L., Manzulli V, Rondinone V., Pace L., Cipolletta D., Galante D. |
| EPI_ISL_794748 | SISP ASL BA AREA SUD | Istituto Zooprofilattico Sperimentale della Puglia e della Basilicata | Parisi A., Bianco A., Capozzi L., Del Sambro L., Manzulli V, Rondinone V., Pace L., Cipolletta D., Galante D. |
| EPI_ISL_794749 | Istituto Zooprofilattico Sperimentale della Puglia e della Basilicata | Istituto Zooprofilattico Sperimentale della Puglia e della Basilicata | Parisi A., Bianco A., Capozzi L., Del Sambro L., Manzulli V, Rondinone V., Pace L., Cipolletta D., Galante D. |
| EPI_ISL_794750 | Ospedale Santa Caterina Novella | Istituto Zooprofilattico Sperimentale della Puglia e della Basilicata | Parisi A., Bianco A., Capozzi L., Del Sambro L., Manzulli V, Rondinone V., Pace L., Cipolletta D., Galante D. |
| EPI_ISL_794751 | Ospedale Vito Fazzi | Istituto Zooprofilattico Sperimentale della Puglia e della Basilicata | Parisi A., Bianco A., Capozzi L., Del Sambro L., Manzulli V, Rondinone V., Pace L., Cipolletta D., Galante D. |
| EPI_ISL_794752 | Istituto Zooprofilattico Sperimentale della Puglia e della Basilicata | Istituto Zooprofilattico Sperimentale della Puglia e della Basilicata | Parisi A., Bianco A., Capozzi L., Del Sambro L., Manzulli V, Rondinone V., Pace L., Cipolletta D., Galante D. |
| EPI_ISL_794753 | Ospedale Cardinale G. Panico | Istituto Zooprofilattico Sperimentale della Puglia e della Basilicata | Parisi A., Bianco A., Capozzi L., Del Sambro L., Manzulli V, Rondinone V., Pace L., Cipolletta D., Galante D. |
| EPI_ISL_794754, EPI_ISL_794755 | Istituto Zooprofilattico Sperimentale della Puglia e della Basilicata | Istituto Zooprofilattico Sperimentale della Puglia e della Basilicata | Parisi A., Bianco A., Capozzi L., Del Sambro L., Manzulli V, Rondinone V., Pace L., Cipolletta D., Galante D. |
| EPI_ISL_794756, EPI_ISL_794757, EPI_ISL_794758 | Presidio di Brindisi Di Summa - Perrino | Istituto Zooprofilattico Sperimentale della Puglia e della Basilicata | Parisi A., Bianco A., Capozzi L., Del Sambro L., Manzulli V, Rondinone V., Pace L., Cipolletta D., Galante D. |
| EPI_ISL_794759, EPI_ISL_794760, EPI_ISL_794761, EPI_ISL_794762, EPI_ISL_794763, EPI_ISL_794764, EPI_ISL_794765, EPI_ISL_794766, EPI_ISL_794767, EPI_ISL_794768, EPI_ISL_794769, EPI_ISL_794770, EPI_ISL_794771, EPI_ISL_794772, EPI_ISL_794773, EPI_ISL_794774, EPI_ISL_794775, EPI_ISL_794776, EPI_ISL_794777, EPI_ISL_794778, EPI_ISL_794779, EPI_ISL_794780, EPI_ISL_794781, EPI_ISL_794782, EPI_ISL_794783, EPI_ISL_794784, EPI_ISL_794785, EPI_ISL_794786, EPI_ISL_794787, EPI_ISL_794788, EPI_ISL_794789, EPI_ISL_794790, EPI_ISL_794791, EPI_ISL_794792, EPI_ISL_794793, EPI_ISL_794794, EPI_ISL_794795, EPI_ISL_794796, EPI_ISL_794797, EPI_ISL_794798, EPI_ISL_794799, EPI_ISL_794800, EPI_ISL_794801, EPI_ISL_794802, EPI_ISL_794803, EPI_ISL_794804, EPI_ISL_794805, EPI_ISL_794806, EPI_ISL_794807, EPI_ISL_794808, EPI_ISL_794809, EPI_ISL_794810, EPI_ISL_794811, EPI_ISL_794812, EPI_ISL_794813, EPI_ISL_794814, EPI_ISL_794815, EPI_ISL_794816, EPI_ISL_794817 | | | |
| see above | Istituto Zooprofilattico Sperimentale della Puglia e della Basilicata | Istituto Zooprofilattico Sperimentale della Puglia e della Basilicata | Parisi A., Bianco A., Capozzi L., Del Sambro L., Manzulli V, Rondinone V., Pace L., Cipolletta D., Galante D. |
| EPI_ISL_794826, EPI_ISL_794827, EPI_ISL_794828 | ULSS 7 Pedemontana - Distretto 2 | Istituto Zooprofilattico Sperimentale delle Venezie | Adelaide Milani, Alessia Schivo, Annalisa Salviato, Erika Giorgia Quaranta, Ambra Pastori, Bianca Zecchin, Alice Fusaro, Isabella Monne, Calogero Terregino, Antonia Ricci |
| EPI_ISL_803880, EPI_ISL_803881, EPI_ISL_803882, EPI_ISL_803883, EPI_ISL_803884, EPI_ISL_803885, EPI_ISL_803886, EPI_ISL_803887, EPI_ISL_803888, EPI_ISL_803891, EPI_ISL_803892, EPI_ISL_803893, EPI_ISL_803895 | | | |

| | | | |
|---|---|---|---|
| see above | Department of Medical Biotechnologies, University of Siena | Laboratory of Infectious Diseases, Department of Biomedical and Clinical Sciences L. Sacco, University of Milan | Ilaria Vicenti, Filippo Dragoni, Maurizio Zazzi, Maria Grazia Cusi, Alessia Lai, Annalisa Bergna, Carla Della Ventura, Claudia Balotta, Massimo Galli, Gianguglielmo Zehender on behalf of SARS-CoV-2 ITALIAN RESEARCH ENTERPRISE-(SCIRE) Collaborative Group |
| EPI_ISL_803896 | Virology Unit, Pisa University Hospital and Retrovirus Center, University of Pisa | National Institute for Infectious Diseases, INMI, "L. Spallanzani" IRCCS | M Rueca, E Giombini, C.E.M Gruber, B Bartolini, O Butera, F Messina, A Rosellini, P Mazzetti, M Pistello, A Di Caro, MR Capobianchi |
| EPI_ISL_803897 | National Institute for Infectious Diseases, INMI, "L. Spallanzani" IRCCS | National Institute for Infectious Diseases, INMI, "L. Spallanzani" IRCCS | F Messina, O Butera, E Giombini, M Rueca, B Bartolini, C.E.M Gruber, MR Capobianchi, A Di Caro |
| EPI_ISL_803898 | National Institute for Infectious Diseases, INMI, "L. Spallanzani" IRCCS | National Institute for Infectious Diseases, INMI, "L. Spallanzani" IRCCS | C.E.M Gruber, B Bartolini, E Giombini, M Rueca, O Butera, F Messina, A Di Caro, MR Capobianchi |
| EPI_ISL_803899 | Genomic Medicine Laboratory, IRCCS Santa Lucia Foundation | National Institute for Infectious Diseases, INMI, "L. Spallanzani" IRCCS | E Giombini, M. Rueca, B Bartolini, O Butera, C.E.M Gruber, F Messina, E Giardina, MR Capobianchi, A Di Caro |
| EPI_ISL_803900 | National Institute for Infectious Diseases, INMI, "L. Spallanzani" IRCCS | National Institute for Infectious Diseases, INMI, "L. Spallanzani" IRCCS | B Bartolini, O Butera, C.E.M Gruber, M Rueca, F Messina, E Giombini, MR Capobianchi, A Di Caro |
| EPI_ISL_804038, EPI_ISL_804040, EPI_ISL_804041, EPI_ISL_804042, EPI_ISL_804044, EPI_ISL_804048, EPI_ISL_804051, EPI_ISL_804052, EPI_ISL_804053, EPI_ISL_804054 | SC (UCO) Igiene e Sanità Pubblica (funzione integrata con SC Microbiologia e Virologia) e Laboratory of Molecular Virology of the International Centre for Genetic Engineering and Biotechnology (ICGEB) | ARGO Laboratorio Genomica ed Epigenomica | Licastro D, Dal Monego S, Degasperi M, Marcello A, D'Agaro P |
| EPI_ISL_806730 | Presidio Ospedaliero S.Liberatore Atri | Istituto Zooprofilattico Sperimentale dell'Abruzzo e Molise "G. Caporale" | Lorusso A, Marcacci M, Di Domenico M, Ancora M, Curini V, Mangone I, Rinaldi A, Di Pasquale A, Cammà C, Puglia I, Calistri P, Savini G |
| EPI_ISL_806731, EPI_ISL_806732 | SIESP DIPARTIMENTO DI PREVENZIONE CHIETI | Istituto Zooprofilattico Sperimentale dell'Abruzzo e Molise "G. Caporale" | Lorusso A, Marcacci M, Di Domenico M, Ancora M, Curini V, Mangone I, Rinaldi A, Di Pasquale A, Cammà C, Puglia I, Calistri P, Savini G |
| EPI_ISL_806733 | SIESP CHIETI - DRIVE IN CHIETI | Istituto Zooprofilattico Sperimentale dell'Abruzzo e Molise "G. Caporale" | Lorusso A, Marcacci M, Di Domenico M, Ancora M, Curini V, Mangone I, Rinaldi A, Di Pasquale A, Cammà C, Puglia I, Calistri P, Savini G |
| EPI_ISL_806734 | SIESP CHIETI - DRIVE IN LANCIANO | Istituto Zooprofilattico Sperimentale dell'Abruzzo e Molise "G. Caporale" | Lorusso A, Marcacci M, Di Domenico M, Ancora M, Curini V, Mangone I, Rinaldi A, Di Pasquale A, Cammà C, Puglia I, Calistri P, Savini G |
| EPI_ISL_806735, EPI_ISL_806736, EPI_ISL_806737, EPI_ISL_806738, EPI_ISL_806739, EPI_ISL_806740, EPI_ISL_806741, EPI_ISL_806742, EPI_ISL_806743, EPI_ISL_806744 | SIESP DIPARTIMENTO DI PREVENZIONE TERAMO | Istituto Zooprofilattico Sperimentale dell'Abruzzo e Molise "G. Caporale" | Lorusso A, Marcacci M, Di Domenico M, Ancora M, Curini V, Mangone I, Rinaldi A, Di Pasquale A, Cammà C, Puglia I, Calistri P, Savini G |
| EPI_ISL_806745 | RSA Giulianova | Istituto Zooprofilattico Sperimentale dell'Abruzzo e Molise "G. Caporale" | Lorusso A, Marcacci M, Di Domenico M, Ancora M, Curini V, Mangone I, Rinaldi A, Di Pasquale A, Cammà C, Puglia I, Calistri P, Savini G |
| EPI_ISL_806746, EPI_ISL_806747, EPI_ISL_806748 | SIESP DIPARTIMENTO DI PREVENZIONE TERAMO | Istituto Zooprofilattico Sperimentale dell'Abruzzo e Molise "G. Caporale" | Lorusso A, Marcacci M, Di Domenico M, Ancora M, Curini V, Mangone I, Rinaldi A, Di Pasquale A, Cammà C, Puglia I, Calistri P, Savini G |
| EPI_ISL_806749, EPI_ISL_806750, EPI_ISL_806751, EPI_ISL_806752, EPI_ISL_806753, EPI_ISL_806754, EPI_ISL_806755, EPI_ISL_806756, EPI_ISL_806757 | SIESP DIPARTIMENTO DI PREVENZIONE SULMONA | Istituto Zooprofilattico Sperimentale dell'Abruzzo e Molise "G. Caporale" | Lorusso A, Marcacci M, Di Domenico M, Ancora M, Curini V, Mangone I, Rinaldi A, Di Pasquale A, Cammà C, Puglia I, Calistri P, Savini G |
| EPI_ISL_806758, EPI_ISL_806759, EPI_ISL_806760, EPI_ISL_806761 | DIPARTIMENTO PREVENZIONE AVEZZANO-SERVIZIO DI IGIENE EPIDEMIOLOGIA E SANITA' PUBBLICA | Istituto Zooprofilattico Sperimentale dell'Abruzzo e Molise "G. Caporale" | Lorusso A, Marcacci M, Di Domenico M, Ancora M, Curini V, Mangone I, Rinaldi A, Di Pasquale A, Cammà C, Puglia I, Calistri P, Savini G |
| EPI_ISL_806762 | SIESP CHIETI - DRIVE IN ORTONA | Istituto Zooprofilattico Sperimentale dell'Abruzzo e Molise "G. Caporale" | Lorusso A, Marcacci M, Di Domenico M, Ancora M, Curini V, Mangone I, Rinaldi A, Di Pasquale A, Cammà C, Puglia I, Calistri P, Savini G |
| EPI_ISL_806763, EPI_ISL_806764, EPI_ISL_806765 | DIPARTIMENTO PREVENZIONE AVEZZANO-SERVIZIO DI IGIENE EPIDEMIOLOGIA E SANITA' PUBBLICA | Istituto Zooprofilattico Sperimentale dell'Abruzzo e Molise "G. Caporale" | Lorusso A, Marcacci M, Di Domenico M, Ancora M, Curini V, Mangone I, Rinaldi A, Di Pasquale A, Cammà C, Puglia I, Calistri P, Savini G |
| EPI_ISL_806766, EPI_ISL_806767, EPI_ISL_806768, EPI_ISL_806769, EPI_ISL_806770, EPI_ISL_806771, EPI_ISL_806772 | SIESP DIPARTIMENTO DI PREVENZIONE TERAMO | Istituto Zooprofilattico Sperimentale dell'Abruzzo e Molise "G. Caporale" | Lorusso A, Marcacci M, Di Domenico M, Ancora M, Curini V, Mangone I, Rinaldi A, Di Pasquale A, Cammà C, Puglia I, Calistri P, Savini G |
| EPI_ISL_806773, EPI_ISL_806774 | Ospedale S.Salvatore-Medicina Interna L'Aquila | Istituto Zooprofilattico Sperimentale dell'Abruzzo e Molise "G. Caporale" | Lorusso A, Marcacci M, Di Domenico M, Ancora M, Curini V, Mangone I, Rinaldi A, Di Pasquale A, Cammà C, Puglia I, Calistri P, Savini G |
| EPI_ISL_806775, EPI_ISL_806776 | SIESP CHIETI - DRIVE IN LANCIANO | Istituto Zooprofilattico Sperimentale dell'Abruzzo e Molise "G. Caporale" | Lorusso A, Marcacci M, Di Domenico M, Ancora M, Curini V, Mangone I, Rinaldi A, Di Pasquale A, Cammà C, Puglia I, Calistri P, Savini G |
| EPI_ISL_806777, EPI_ISL_806778, EPI_ISL_806779, EPI_ISL_806780 | SIESP DIPARTIMENTO DI PREVENZIONE TERAMO | Istituto Zooprofilattico Sperimentale dell'Abruzzo e Molise "G. Caporale" | Lorusso A, Marcacci M, Di Domenico M, Ancora M, Curini V, Mangone I, Rinaldi A, Di Pasquale A, Cammà C, Puglia I, Calistri P, Savini G |
| EPI_ISL_806781, EPI_ISL_806782 | DIPARTIMENTO PREVENZIONE AVEZZANO-SERVIZIO DI IGIENE EPIDEMIOLOGIA E SANITA' PUBBLICA | Istituto Zooprofilattico Sperimentale dell'Abruzzo e Molise "G. Caporale" | Lorusso A, Marcacci M, Di Domenico M, Ancora M, Curini V, Mangone I, Rinaldi A, Di Pasquale A, Cammà C, Puglia I, Calistri P, Savini G |
| EPI_ISL_806783 | Casa di cura Di Lorenzo- Avezzano | Istituto Zooprofilattico Sperimentale dell'Abruzzo e Molise "G. Caporale" | Lorusso A, Marcacci M, Di Domenico M, Ancora M, Curini V, Mangone I, Rinaldi A, Di Pasquale A, Cammà C, Puglia I, Calistri P, Savini G |
| EPI_ISL_806784, EPI_ISL_806785 | Ospedale Civile G.Mazzini-Teramo | Istituto Zooprofilattico Sperimentale dell'Abruzzo e Molise "G. Caporale" | Lorusso A, Marcacci M, Di Domenico M, Ancora M, Curini V, Mangone I, Rinaldi A, Di Pasquale A, Cammà C, Puglia I, Calistri P, Savini G |
| EPI_ISL_806786, EPI_ISL_806787, EPI_ISL_806788 | SIESP DIPARTIMENTO DI PREVENZIONE TERAMO | Istituto Zooprofilattico Sperimentale dell'Abruzzo e Molise "G. Caporale" | Lorusso A, Marcacci M, Di Domenico M, Ancora M, Curini V, Mangone I, Rinaldi A, Di Pasquale A, Cammà C, Puglia I, Calistri P, Savini G |
| EPI_ISL_806789 | Ospedale Civile Giulianova | Istituto Zooprofilattico Sperimentale dell'Abruzzo e Molise "G. Caporale" | Lorusso A, Marcacci M, Di Domenico M, Ancora M, Curini V, Mangone I, Rinaldi A, Di Pasquale A, Cammà C, Puglia I, Calistri P, Savini G |
| EPI_ISL_806790 | SIESP DIPARTIMENTO DI PREVENZIONE DELL'AQUILA | Istituto Zooprofilattico Sperimentale dell'Abruzzo e Molise "G. Caporale" | Lorusso A, Marcacci M, Di Domenico M, Ancora M, Curini V, Mangone I, Rinaldi A, Di Pasquale A, Cammà C, Puglia I, Calistri P, Savini G |
| EPI_ISL_806791, EPI_ISL_806792, EPI_ISL_806793, EPI_ISL_806794, EPI_ISL_806795, EPI_ISL_806796, EPI_ISL_806797, EPI_ISL_806798 | SIESP CHIETI - DRIVE IN ORTONA | Istituto Zooprofilattico Sperimentale dell'Abruzzo e Molise "G. Caporale" | Lorusso A, Marcacci M, Di Domenico M, Ancora M, Curini V, Mangone I, Rinaldi A, Di Pasquale A, Cammà C, Puglia I, Calistri P, Savini G |
| EPI_ISL_806799 | SIESP CHIETI - DRIVE IN LANCIANO | Istituto Zooprofilattico Sperimentale dell'Abruzzo e Molise "G. Caporale" | Lorusso A, Marcacci M, Di Domenico M, Ancora M, Curini V, Mangone I, Rinaldi A, Di Pasquale A, Cammà C, Puglia I, Calistri P, Savini G |

| | | | |
|---|---|---|---|
| EPI_ISL_806800, EPI_ISL_806801, EPI_ISL_806802, EPI_ISL_806803 | SIESP CHIETI - DRIVE IN GISSI | Istituto Zooprofilattico Sperimentale dell'Abruzzo e Molise "G. Caporale" | Lorusso A, Marcacci M, Di Domenico M, Ancora M, Curini V, Mangone I, Rinaldi A, Di Pasquale A, Cammà C, Puglia I, Calistri P, Savini G |
| EPI_ISL_806804, EPI_ISL_806805 | SIESP CHIETI - DISTRETTO SANITARIO CHIETI | Istituto Zooprofilattico Sperimentale dell'Abruzzo e Molise "G. Caporale" | Lorusso A, Marcacci M, Di Domenico M, Ancora M, Curini V, Mangone I, Rinaldi A, Di Pasquale A, Cammà C, Puglia I, Calistri P, Savini G |
| EPI_ISL_806806 | SIESP DIPARTIMENTO DI PREVENZIONE CHIETI | Istituto Zooprofilattico Sperimentale dell'Abruzzo e Molise "G. Caporale" | Lorusso A, Marcacci M, Di Domenico M, Ancora M, Curini V, Mangone I, Rinaldi A, Di Pasquale A, Cammà C, Puglia I, Calistri P, Savini G |
| EPI_ISL_812968 | Department of Molecular Medicine, University of Padova | Department of Molecular Medicine, University of Padova | Lavezzo,E., Franchin,E., Barzon,L., Del Vecchio,C., Rossi,L.,Manganelli,R., Loregian,A., Abate,D., Sciro,M., De Canale,E.,Vanuzzo,M.C., Besutti,V., Saluzzo,F., Onelia,F., Pacenti,M.,Manuto,L., Parisi,S.G., Masi,G., Trevisan,M., Toppo,S. and Crisanti,A. |
| EPI_ISL_824407 | Istituto Nazionale Malattie Infettive Lazzaro Spallanzani IRCCS | Istituto Nazionale Malattie Infettive Lazzaro Spallanzani IRCCS | Emanuela Giombini, Ornella Butera, Cesare E.M. Gruber, Martina Rueca, Francesco Messina, Barbara Bartolini, Silvia Meschi, Francesca Colavita, Concetta Castilletti, Antonino Di Caro, Maria R. Capobianchi |
| EPI_ISL_824408 | Istituto Nazionale Malattie Infettive Lazzaro Spallanzani IRCCS | Istituto Nazionale Malattie Infettive Lazzaro Spallanzani IRCCS | Barbara Bartolini, Ornella Butera, Cesare E.M. Gruber, Martina Rueca, Francesco Messina, Emanuela Giombini, Silvia Meschi, Francesca Colavita, Concetta Castilletti, Maria R. Capobianchi, Antonino Di Caro |
| EPI_ISL_825155, EPI_ISL_825156 | Microbiology and Virology Unit, Florence Careggi University Hospital | Microbiology and Virology Unit, Florence Careggi University Hospital | Vincenzo Di Pilato, Marco Coppi, Fabio Morecchiato, Alberto Antonelli, Emanuele Gori, Gian Maria Rossolini |
| EPI_ISL_826284, EPI_ISL_826458 | University of Bari Biomedical Sciences and Human Oncology | University of Bari Biomedical Sciences and Human Oncology | Chironna Maria, Sallustio Anna, Loconsole Daniela, Accogli Marisa |
| EPI_ISL_826520 | University of Bari Biomedical Sciences and Human Oncology | University of Bari Biomedical Sciences and Human Oncology | Chironna M, Sallustio A., Loconsole D., Accogli M. |
| EPI_ISL_832815, EPI_ISL_832816 | SIESP CHIETI - DRIVE IN LANCIANO | Istituto Zooprofilattico Sperimentale dell'Abruzzo e Molise "G.Caporale" | Lorusso A, Marcacci M, Di Domenico M, Curini V, Ancora M, Cammà C, Rinaldi A, Mangone I, Di Pasquale A, Puglia I, Calistri P, Savini G. |
| EPI_ISL_832817 | SIESP DIPARTIMENTO DI PREVENZIONE TERAMO | Istituto Zooprofilattico Sperimentale dell'Abruzzo e Molise "G.Caporale" | Lorusso A, Marcacci M, Di Domenico M, Curini V, Ancora M, Cammà C, Rinaldi A, Mangone I, Di Pasquale A, Puglia I, Calistri P, Savini G. |
| EPI_ISL_832818 | Ospedale Civile Atri | Istituto Zooprofilattico Sperimentale dell'Abruzzo e Molise "G.Caporale" | Lorusso A, Marcacci M, Di Domenico M, Curini V, Ancora M, Cammà C, Rinaldi A, Mangone I, Di Pasquale A, Puglia I, Calistri P, Savini G. |
| EPI_ISL_832819 | SIESP DIPARTIMENTO DI PREVENZIONE SULMONA | Istituto Zooprofilattico Sperimentale dell'Abruzzo e Molise "G.Caporale" | Lorusso A, Marcacci M, Di Domenico M, Curini V, Ancora M, Cammà C, Rinaldi A, Mangone I, Di Pasquale A, Puglia I, Calistri P, Savini G. |
| EPI_ISL_832820 | Medico Competente P.O. L'Aquila | Istituto Zooprofilattico Sperimentale dell'Abruzzo e Molise "G.Caporale" | Lorusso A, Marcacci M, Di Domenico M, Curini V, Ancora M, Cammà C, Rinaldi A, Mangone I, Di Pasquale A, Puglia I, Calistri P, Savini G. |
| EPI_ISL_832821 | SIESP CHIETI - DRIVE IN CHIETI | Istituto Zooprofilattico Sperimentale dell'Abruzzo e Molise "G.Caporale" | Lorusso A, Marcacci M, Di Domenico M, Curini V, Ancora M, Cammà C, Rinaldi A, Mangone I, Di Pasquale A, Puglia I, Calistri P, Savini G. |
| EPI_ISL_832822 | SIESP CHIETI - DRIVE IN LANCIANO | Istituto Zooprofilattico Sperimentale dell'Abruzzo e Molise "G.Caporale" | Lorusso A, Marcacci M, Di Domenico M, Curini V, Ancora M, Cammà C, Rinaldi A, Mangone I, Di Pasquale A, Puglia I, Calistri P, Savini G. |
| EPI_ISL_833042 | OSPEDALE SAN SALVATORE L'AQUILA UOC PNEUMOLOGIAE UTSIR | Istituto Zooprofilattico Sperimentale dell'Abruzzo e Molise "G. Caporale" | Lorusso A, Marcacci M, Di Domenico M, Ancora M, Curini V, Mangone I, Rinaldi A, Delli Compagni E, Di Pasquale A, Cammà C, Puglia I, Calistri P, Savini G |
| EPI_ISL_833043 | SIESP DIPARTIMENTO DI PREVENZIONE CHIETI | Istituto Zooprofilattico Sperimentale dell'Abruzzo e Molise "G. Caporale" | Lorusso A, Marcacci M, Di Domenico M, Ancora M, Curini V, Mangone I, Rinaldi A, Delli Compagni E, Di Pasquale A, Cammà C, Puglia I, Calistri P, Savini G |
| EPI_ISL_833044 | SIESP CHIETI - DRIVE IN CHIETI | Istituto Zooprofilattico Sperimentale dell'Abruzzo e Molise "G. Caporale" | Lorusso A, Marcacci M, Di Domenico M, Ancora M, Curini V, Mangone I, Rinaldi A, Delli Compagni E, Di Pasquale A, Cammà C, Puglia I, Calistri P, Savini G |
| EPI_ISL_833045 | SIESP CHIETI - DRIVE IN ORTONA | Istituto Zooprofilattico Sperimentale dell'Abruzzo e Molise "G. Caporale" | Lorusso A, Marcacci M, Di Domenico M, Ancora M, Curini V, Mangone I, Rinaldi A, Delli Compagni E, Di Pasquale A, Cammà C, Puglia I, Calistri P, Savini G |
| EPI_ISL_833046 | FPAM | Istituto Zooprofilattico Sperimentale dell'Abruzzo e Molise "G. Caporale" | Lorusso A, Marcacci M, Di Domenico M, Ancora M, Curini V, Mangone I, Rinaldi A, Delli Compagni E, Di Pasquale A, Cammà C, Puglia I, Calistri P, Savini G |
| EPI_ISL_833047, EPI_ISL_833048 | OSPEDALE CIVILE ATRI | Istituto Zooprofilattico Sperimentale dell'Abruzzo e Molise "G. Caporale" | Lorusso A, Marcacci M, Di Domenico M, Ancora M, Curini V, Mangone I, Rinaldi A, Delli Compagni E, Di Pasquale A, Cammà C, Puglia I, Calistri P, Savini G |
| EPI_ISL_833049 | SIESP CHIETI - DRIVE IN LANCIANO | Istituto Zooprofilattico Sperimentale dell'Abruzzo e Molise "G. Caporale" | Lorusso A, Marcacci M, Di Domenico M, Ancora M, Curini V, Mangone I, Rinaldi A, Delli Compagni E, Di Pasquale A, Cammà C, Puglia I, Calistri P, Savini G |
| EPI_ISL_833050, EPI_ISL_833051 | DIP. PREV. AVEZZANO SERVIZIO DI IGIENE EPIDEMIOLOGIAE SANITA' PUBBLICA | Istituto Zooprofilattico Sperimentale dell'Abruzzo e Molise "G. Caporale" | Lorusso A, Marcacci M, Di Domenico M, Ancora M, Curini V, Mangone I, Rinaldi A, Delli Compagni E, Di Pasquale A, Cammà C, Puglia I, Calistri P, Savini G |
| EPI_ISL_833052, EPI_ISL_833053, EPI_ISL_833054 | "Presidio Ospedaliero "San Liberatore" Atri | Istituto Zooprofilattico Sperimentale dell'Abruzzo e Molise "G. Caporale" | Lorusso A, Marcacci M, Di Domenico M, Ancora M, Curini V, Mangone I, Rinaldi A, Delli Compagni E, Di Pasquale A, Cammà C, Puglia I, Calistri P, Savini G |
| EPI_ISL_833055, EPI_ISL_833056 | "SIESP DIPARTIMENTO DI PREVENZIONE CHIETi | Istituto Zooprofilattico Sperimentale dell'Abruzzo e Molise "G. Caporale" | Lorusso A, Marcacci M, Di Domenico M, Ancora M, Curini V, Mangone I, Rinaldi A, Delli Compagni E, Di Pasquale A, Cammà C, Puglia I, Calistri P, Savini G |
| EPI_ISL_833057, EPI_ISL_833058, EPI_ISL_833059, EPI_ISL_833060, EPI_ISL_833061, EPI_ISL_833062, EPI_ISL_833063, EPI_ISL_833064, EPI_ISL_833065, EPI_ISL_833066, EPI_ISL_833067 | | | |
| see above | DIP. PREV. AVEZZANO SERVIZIO DI IGIENE EPIDEMIOLOGIAE SANITA' PUBBLICA | Istituto Zooprofilattico Sperimentale dell'Abruzzo e Molise "G. Caporale" | Lorusso A, Marcacci M, Di Domenico M, Ancora M, Curini V, Mangone I, Rinaldi A, Delli Compagni E, Di Pasquale A, Cammà C, Puglia I, Calistri P, Savini G |
| EPI_ISL_833068, EPI_ISL_833069, EPI_ISL_833070, EPI_ISL_833071, EPI_ISL_833072, EPI_ISL_833073, EPI_ISL_833074, EPI_ISL_833075 | FPAM | Istituto Zooprofilattico Sperimentale dell'Abruzzo e Molise "G. Caporale" | Lorusso A, Marcacci M, Di Domenico M, Ancora M, Curini V, Mangone I, Rinaldi A, Delli Compagni E, Di Pasquale A, Cammà C, Puglia I, Calistri P, Savini G |
| EPI_ISL_833076 | IZSAM | Istituto Zooprofilattico Sperimentale dell'Abruzzo e Molise "G. Caporale" | Lorusso A, Marcacci M, Di Domenico M, Ancora M, Curini V, Mangone I, Rinaldi A, Delli Compagni E, Di Pasquale A, Cammà C, Puglia I, Calistri P, Savini G |
| EPI_ISL_833077, EPI_ISL_833078 | OSPEDALE CIVILE ATRI | Istituto Zooprofilattico Sperimentale dell'Abruzzo e Molise "G. Caporale" | Lorusso A, Marcacci M, Di Domenico M, Ancora M, Curini V, Mangone I, Rinaldi A, Delli Compagni E, Di Pasquale A, Cammà C, Puglia I, Calistri P, Savini G |
| EPI_ISL_833079, EPI_ISL_833080 | OSPEDALE CIVILE TERAMO | Istituto Zooprofilattico Sperimentale dell'Abruzzo e Molise "G. Caporale" | Lorusso A, Marcacci M, Di Domenico M, Ancora M, Curini V, Mangone I, Rinaldi A, Delli Compagni E, Di Pasquale A, Cammà C, Puglia I, Calistri P, Savini G |
| EPI_ISL_833081 | OSPEDALE S.S. ANNUNZIATA CHIETI | Istituto Zooprofilattico Sperimentale dell'Abruzzo e Molise "G. Caporale" | Lorusso A, Marcacci M, Di Domenico M, Ancora M, Curini V, Mangone I, Rinaldi A, Delli Compagni E, Di Pasquale A, Cammà C, Puglia I, Calistri P, Savini G |
| EPI_ISL_833082 | OSPEDALE SAN SALVATORE | Istituto Zooprofilattico Sperimentale dell'Abruzzo e Molise "G. Caporale" | Lorusso A, Marcacci M, Di Domenico M, Ancora M, Curini V, Mangone I, Rinaldi A, Delli Compagni E, Di Pasquale A, Cammà C, Puglia I, Calistri P, Savini G |
| EPI_ISL_833083, EPI_ISL_833084 | RPS | Istituto Zooprofilattico Sperimentale dell'Abruzzo e Molise "G. Caporale" | Lorusso A, Marcacci M, Di Domenico M, Ancora M, Curini V, Mangone I, Rinaldi A, Delli Compagni E, Di Pasquale A, Cammà C, Puglia I, Calistri P, Savini G |
| EPI_ISL_833085, EPI_ISL_833086 | Servizio di Igiene Epidemiologia e Sanità Pubblica (SIESP) | Istituto Zooprofilattico Sperimentale dell'Abruzzo e Molise "G. | Lorusso A, Marcacci M, Di Domenico M, Ancora M, Curini V, Mangone I, Rinaldi A, Delli Compagni E, Di Pasquale A, Cammà C, Puglia I, Calistri P, Savini |

| | | | |
|---|---|---|---|
| | CHIETI - DRIVE IN GISSI | Caporale" | G |
| EPI_ISL_833087 | Servizio di Igiene Epidemiologia e Sanità Pubblica (SIESP) L'AQUILA | Istituto Zooprofilattico Sperimentale dell'Abruzzo e Molise "G. Caporale" | Lorusso A, Marcacci M, Di Domenico M, Ancora M, Curini V, Mangone I, Rinaldi A, Delli Compagni E, Di Pasquale A, Cammà C, Puglia I, Calistri P, Savini G |
| EPI_ISL_833088, EPI_ISL_833089 | Servizio di Igiene Epidemiologia e Sanità Pubblica (SIESP) SULMONA | Istituto Zooprofilattico Sperimentale dell'Abruzzo e Molise "G. Caporale" | Lorusso A, Marcacci M, Di Domenico M, Ancora M, Curini V, Mangone I, Rinaldi A, Delli Compagni E, Di Pasquale A, Cammà C, Puglia I, Calistri P, Savini G |
| EPI_ISL_833090, EPI_ISL_833091, EPI_ISL_833092, EPI_ISL_833093, EPI_ISL_833094, EPI_ISL_833095, EPI_ISL_833096 | SIESP CHIETI - DRIVE IN CHIETI | Istituto Zooprofilattico Sperimentale dell'Abruzzo e Molise "G. Caporale" | Lorusso A, Marcacci M, Di Domenico M, Ancora M, Curini V, Mangone I, Rinaldi A, Delli Compagni E, Di Pasquale A, Cammà C, Puglia I, Calistri P, Savini G |
| EPI_ISL_833097, EPI_ISL_833098 | SIESP CHIETI - DRIVE IN LANCIANO | Istituto Zooprofilattico Sperimentale dell'Abruzzo e Molise "G. Caporale" | Lorusso A, Marcacci M, Di Domenico M, Ancora M, Curini V, Mangone I, Rinaldi A, Delli Compagni E, Di Pasquale A, Cammà C, Puglia I, Calistri P, Savini G |
| EPI_ISL_833099, EPI_ISL_833100 | SIESP CHIETI - DRIVE IN ORTONA | Istituto Zooprofilattico Sperimentale dell'Abruzzo e Molise "G. Caporale" | Lorusso A, Marcacci M, Di Domenico M, Ancora M, Curini V, Mangone I, Rinaldi A, Delli Compagni E, Di Pasquale A, Cammà C, Puglia I, Calistri P, Savini G |
| EPI_ISL_833101, EPI_ISL_833102 | SIESP DIPARTIMENTO DI PREVENZIONE CHIETI | Istituto Zooprofilattico Sperimentale dell'Abruzzo e Molise "G. Caporale" | Lorusso A, Marcacci M, Di Domenico M, Ancora M, Curini V, Mangone I, Rinaldi A, Delli Compagni E, Di Pasquale A, Cammà C, Puglia I, Calistri P, Savini G |
| EPI_ISL_833103, EPI_ISL_833104, EPI_ISL_833105, EPI_ISL_833106, EPI_ISL_833107, EPI_ISL_833108, EPI_ISL_833109, EPI_ISL_833110, EPI_ISL_833111, EPI_ISL_833112, EPI_ISL_833113, EPI_ISL_833114, EPI_ISL_833115, EPI_ISL_833116, EPI_ISL_833117, EPI_ISL_833118 | | | |
| see above | SIESP DIPARTIMENTO DI PREVENZIONE TERAMO C.DA CASALENA | Istituto Zooprofilattico Sperimentale dell'Abruzzo e Molise "G. Caporale" | Lorusso A, Marcacci M, Di Domenico M, Ancora M, Curini V, Mangone I, Rinaldi A, Delli Compagni E, Di Pasquale A, Cammà C, Puglia I, Calistri P, Savini G |
| EPI_ISL_833119 | SIESP L'AQUILA | Istituto Zooprofilattico Sperimentale dell'Abruzzo e Molise "G. Caporale" | Lorusso A, Marcacci M, Di Domenico M, Ancora M, Curini V, Mangone I, Rinaldi A, Delli Compagni E, Di Pasquale A, Cammà C, Puglia I, Calistri P, Savini G |
| EPI_ISL_833120, EPI_ISL_833121, EPI_ISL_833122 | SIESP SULMONA | Istituto Zooprofilattico Sperimentale dell'Abruzzo e Molise "G. Caporale" | Lorusso A, Marcacci M, Di Domenico M, Ancora M, Curini V, Mangone I, Rinaldi A, Delli Compagni E, Di Pasquale A, Cammà C, Puglia I, Calistri P, Savini G |
| EPI_ISL_833123 | Usca Pizzoli | Istituto Zooprofilattico Sperimentale dell'Abruzzo e Molise "G. Caporale" | Lorusso A, Marcacci M, Di Domenico M, Ancora M, Curini V, Mangone I, Rinaldi A, Delli Compagni E, Di Pasquale A, Cammà C, Puglia I, Calistri P, Savini G |
| EPI_ISL_833124, EPI_ISL_833125, EPI_ISL_833126, EPI_ISL_833127, EPI_ISL_833128 | USCA Sulmona | Istituto Zooprofilattico Sperimentale dell'Abruzzo e Molise "G. Caporale" | Lorusso A, Marcacci M, Di Domenico M, Ancora M, Curini V, Mangone I, Rinaldi A, Delli Compagni E, Di Pasquale A, Cammà C, Puglia I, Calistri P, Savini G |
| EPI_ISL_833129 | USCA Tagliacozzo | Istituto Zooprofilattico Sperimentale dell'Abruzzo e Molise "G. Caporale" | Lorusso A, Marcacci M, Di Domenico M, Ancora M, Curini V, Mangone I, Rinaldi A, Delli Compagni E, Di Pasquale A, Cammà C, Puglia I, Calistri P, Savini G |
| EPI_ISL_833228, EPI_ISL_833229 | R.P. GUARDIAGRELE Ospedale di Comunità | Istitituto Zooprofilattico Sperimentale dell'Abruzzo e Molise "G.Caporale" | Lorusso A, Marcacci M, Di Domenico M, Curini V, Ancora M, Cammà C, Rinaldi A, Mangone I, Di Pasquale A, Puglia I, Calistri P, Savini G. |
| EPI_ISL_833230 | OSPEDALE S.S. ANNUNZIATA CHIETI - CLINICA MEDICA (MEDICINA GENERALE 1) | Istitituto Zooprofilattico Sperimentale dell'Abruzzo e Molise "G.Caporale" | Lorusso A, Marcacci M, Di Domenico M, Curini V, Ancora M, Cammà C, Rinaldi A, Mangone I, Di Pasquale A, Puglia I, Calistri P, Savini G. |
| EPI_ISL_833231 | SIESP CHIETI - DRIVE IN LANCIANO | Istitituto Zooprofilattico Sperimentale dell'Abruzzo e Molise "G.Caporale" | Lorusso A, Marcacci M, Di Domenico M, Curini V, Ancora M, Cammà C, Rinaldi A, Mangone I, Di Pasquale A, Puglia I, Calistri P, Savini G. |
| EPI_ISL_833232 | SIESP CHIETI - DRIVE IN CHIETI | Istitituto Zooprofilattico Sperimentale dell'Abruzzo e Molise "G.Caporale" | Lorusso A, Marcacci M, Di Domenico M, Curini V, Ancora M, Cammà C, Rinaldi A, Mangone I, Di Pasquale A, Puglia I, Calistri P, Savini G. |
| EPI_ISL_833233, EPI_ISL_833234, EPI_ISL_833235, EPI_ISL_833236, EPI_ISL_833237, EPI_ISL_833238 | SIESP DIPARTIMENTO DI PREVENZIONE CHIETI | Istitituto Zooprofilattico Sperimentale dell'Abruzzo e Molise "G.Caporale" | Lorusso A, Marcacci M, Di Domenico M, Curini V, Ancora M, Cammà C, Rinaldi A, Mangone I, Di Pasquale A, Puglia I, Calistri P, Savini G. |
| EPI_ISL_833239, EPI_ISL_833240 | SIESP DIPARTIMENTO DI PREVENZIONE TERAMO | Istitituto Zooprofilattico Sperimentale dell'Abruzzo e Molise "G.Caporale" | Lorusso A, Marcacci M, Di Domenico M, Curini V, Ancora M, Cammà C, Rinaldi A, Mangone I, Di Pasquale A, Puglia I, Calistri P, Savini G. |
| EPI_ISL_833241 | SIESP DIPARTIMENTO DI PREVENZIONE CHIETI | Istitituto Zooprofilattico Sperimentale dell'Abruzzo e Molise "G.Caporale" | Lorusso A, Marcacci M, Di Domenico M, Curini V, Ancora M, Cammà C, Rinaldi A, Mangone I, Di Pasquale A, Puglia I, Calistri P, Savini G. |
| EPI_ISL_833242 | SIESP CHIETI - DRIVE IN LANCIANO | Istitituto Zooprofilattico Sperimentale dell'Abruzzo e Molise "G.Caporale" | Lorusso A, Marcacci M, Di Domenico M, Curini V, Ancora M, Cammà C, Rinaldi A, Mangone I, Di Pasquale A, Puglia I, Calistri P, Savini G. |
| EPI_ISL_833243, EPI_ISL_833244, EPI_ISL_833245, EPI_ISL_833246, EPI_ISL_833247 | SIESP DIPARTIMENTO DI PREVENZIONE CHIETI | Istitituto Zooprofilattico Sperimentale dell'Abruzzo e Molise "G.Caporale" | Lorusso A, Marcacci M, Di Domenico M, Curini V, Ancora M, Cammà C, Rinaldi A, Mangone I, Di Pasquale A, Puglia I, Calistri P, Savini G. |
| EPI_ISL_833250 | SIESP CHIETI - DRIVE IN ORTONA | Istituto Zooprofilattico Sperimentale dell'Abruzzo e Molise "G. Caporale" | Lorusso A, Marcacci M, Di Domenico M, Ancora M, Curini V, Mangone I, Rinaldi A, Di Pasquale A, Cammà C, Puglia I, Calistri P, Savini G |
| EPI_ISL_833251 | Ospedale Civile Atri-Medicina Interna | Istituto Zooprofilattico Sperimentale dell'Abruzzo e Molise "G. Caporale" | Lorusso A, Marcacci M, Di Domenico M, Ancora M, Curini V, Mangone I, Rinaldi A, Di Pasquale A, Cammà C, Puglia I, Calistri P, Savini G |
| EPI_ISL_833252 | Ospedale Civile Giulianova-Pronto Soccorso | Istituto Zooprofilattico Sperimentale dell'Abruzzo e Molise "G. Caporale" | Lorusso A, Marcacci M, Di Domenico M, Ancora M, Curini V, Mangone I, Rinaldi A, Di Pasquale A, Cammà C, Puglia I, Calistri P, Savini G |
| EPI_ISL_833253 | USCA-Avezzano | Istituto Zooprofilattico Sperimentale dell'Abruzzo e Molise "G. Caporale" | Lorusso A, Marcacci M, Di Domenico M, Ancora M, Curini V, Mangone I, Rinaldi A, Di Pasquale A, Cammà C, Puglia I, Calistri P, Savini G |
| EPI_ISL_833254 | USCA-Pescina | Istituto Zooprofilattico Sperimentale dell'Abruzzo e Molise "G. Caporale" | Lorusso A, Marcacci M, Di Domenico M, Ancora M, Curini V, Mangone I, Rinaldi A, Di Pasquale A, Cammà C, Puglia I, Calistri P, Savini G |
| EPI_ISL_833255, EPI_ISL_833256, EPI_ISL_833257 | Ospedale Civile Atri | Istituto Zooprofilattico Sperimentale dell'Abruzzo e Molise "G. Caporale" | Lorusso A, Marcacci M, Di Domenico M, Ancora M, Curini V, Mangone I, Rinaldi A, Di Pasquale A, Cammà C, Puglia I, Calistri P, Savini G |
| EPI_ISL_833258, EPI_ISL_833259 | SIESP DIPARTIMENTO DI PREVENZIONE TERAMO | Istituto Zooprofilattico Sperimentale dell'Abruzzo e Molise "G. Caporale" | Lorusso A, Marcacci M, Di Domenico M, Ancora M, Curini V, Mangone I, Rinaldi A, Di Pasquale A, Cammà C, Puglia I, Calistri P, Savini G |
| EPI_ISL_833260, EPI_ISL_833261 | Giulianova | Istituto Zooprofilattico Sperimentale dell'Abruzzo e Molise "G. Caporale" | Lorusso A, Marcacci M, Di Domenico M, Ancora M, Curini V, Mangone I, Rinaldi A, Di Pasquale A, Cammà C, Puglia I, Calistri P, Savini G |
| EPI_ISL_833262, EPI_ISL_833263, EPI_ISL_833264 | SIESP DIPARTIMENTO DI PREVENZIONE CHIETI | Istituto Zooprofilattico Sperimentale dell'Abruzzo e Molise "G. Caporale" | Lorusso A, Marcacci M, Di Domenico M, Ancora M, Curini V, Mangone I, Rinaldi A, Di Pasquale A, Cammà C, Puglia I, Calistri P, Savini G |
| EPI_ISL_833265, EPI_ISL_833266, EPI_ISL_833267, EPI_ISL_833268, EPI_ISL_833269 | Ospedale SS Annunziata Chieti | Istituto Zooprofilattico Sperimentale dell'Abruzzo e Molise "G. Caporale" | Lorusso A, Marcacci M, Di Domenico M, Ancora M, Curini V, Mangone I, Rinaldi A, Di Pasquale A, Cammà C, Puglia I, Calistri P, Savini G |
| EPI_ISL_833270 | SIESP CHIETI - DRIVE IN ORTONA | Istituto Zooprofilattico Sperimentale dell'Abruzzo e Molise "G. | Lorusso A, Marcacci M, Di Domenico M, Ancora M, Curini V, Mangone I, Rinaldi A, Di Pasquale A, Cammà C, Puglia I, Calistri P, Savini G |

| | | | |
|---|---|---|---|
| | | Caporale" | |
| EPI_ISL_833271 | SIESP CHIETI - DRIVE IN GISSI | Istituto Zooprofilattico Sperimentale dell'Abruzzo e Molise "G. Caporale" | Lorusso A, Marcacci M, Di Domenico M, Ancora M, Curini V, Mangone I, Rinaldi A, Di Pasquale A, Cammà C, Puglia I, Calistri P, Savini G |
| EPI_ISL_833272, EPI_ISL_833273, EPI_ISL_833274, EPI_ISL_833275 | SIESP CHIETI - DRIVE IN ORTONA | Istituto Zooprofilattico Sperimentale dell'Abruzzo e Molise "G. Caporale" | Lorusso A, Marcacci M, Di Domenico M, Ancora M, Curini V, Mangone I, Rinaldi A, Di Pasquale A, Cammà C, Puglia I, Calistri P, Savini G |
| EPI_ISL_833276 | Ospedale Civile G.Mazzini-Teramo | Istituto Zooprofilattico Sperimentale dell'Abruzzo e Molise "G. Caporale" | Lorusso A, Marcacci M, Di Domenico M, Ancora M, Curini V, Mangone I, Rinaldi A, Di Pasquale A, Cammà C, Puglia I, Calistri P, Savini G |
| EPI_ISL_833277 | USCA-Sulmona | Istituto Zooprofilattico Sperimentale dell'Abruzzo e Molise "G. Caporale" | Lorusso A, Marcacci M, Di Domenico M, Ancora M, Curini V, Mangone I, Rinaldi A, Di Pasquale A, Cammà C, Puglia I, Calistri P, Savini G |
| EPI_ISL_833278, EPI_ISL_833279, EPI_ISL_833280 | SIESP CHIETI- DRIVE IN CHIETI | Istituto Zooprofilattico Sperimentale dell'Abruzzo e Molise "G. Caporale" | Lorusso A, Marcacci M, Di Domenico M, Ancora M, Curini V, Mangone I, Rinaldi A, Di Pasquale A, Cammà C, Puglia I, Calistri P, Savini G |
| EPI_ISL_833281, EPI_ISL_833282 | SIESP DIPARTIMENTO DI PREVENZIONE TERAMO | Istituto Zooprofilattico Sperimentale dell'Abruzzo e Molise "G. Caporale" | Lorusso A, Marcacci M, Di Domenico M, Ancora M, Curini V, Mangone I, Rinaldi A, Di Pasquale A, Cammà C, Puglia I, Calistri P, Savini G |
| EPI_ISL_833283 | DIPARTIMENTO PREVENZIONE AVEZZANO-SERVIZIO DI IGIENE EPIDEMIOLOGIA E SANITA' PUBBLICA | Istituto Zooprofilattico Sperimentale dell'Abruzzo e Molise "G. Caporale" | Lorusso A, Marcacci M, Di Domenico M, Ancora M, Curini V, Mangone I, Rinaldi A, Di Pasquale A, Cammà C, Puglia I, Calistri P, Savini G |
| EPI_ISL_833284, EPI_ISL_833285, EPI_ISL_833286, EPI_ISL_833287 | SIESP CHIETI - DRIVE IN ORTONA | Istituto Zooprofilattico Sperimentale dell'Abruzzo e Molise "G. Caporale" | Lorusso A, Marcacci M, Di Domenico M, Ancora M, Curini V, Mangone I, Rinaldi A, Di Pasquale A, Cammà C, Puglia I, Calistri P, Savini G |
| EPI_ISL_833288 | PRESIDIO TERRITORIALE CASOLI CH | Istituto Zooprofilattico Sperimentale dell'Abruzzo e Molise "G. Caporale" | Lorusso A, Marcacci M, Di Domenico M, Ancora M, Curini V, Mangone I, Rinaldi A, Di Pasquale A, Cammà C, Puglia I, Calistri P, Savini G |
| EPI_ISL_833289 | Ospedale SS Annunziata Chieti | Istituto Zooprofilattico Sperimentale dell'Abruzzo e Molise "G. Caporale" | Lorusso A, Marcacci M, Di Domenico M, Ancora M, Curini V, Mangone I, Rinaldi A, Di Pasquale A, Cammà C, Puglia I, Calistri P, Savini G |
| EPI_ISL_833290 | PRESIDIO TERRITORIALE CASOLI CH | Istituto Zooprofilattico Sperimentale dell'Abruzzo e Molise "G. Caporale" | Lorusso A, Marcacci M, Di Domenico M, Ancora M, Curini V, Mangone I, Rinaldi A, Di Pasquale A, Cammà C, Puglia I, Calistri P, Savini G |
| EPI_ISL_833291 | SIESP DIPARTIMENTO DI PREVENZIONE DELL'AQUILA | Istituto Zooprofilattico Sperimentale dell'Abruzzo e Molise "G. Caporale" | Lorusso A, Marcacci M, Di Domenico M, Ancora M, Curini V, Mangone I, Rinaldi A, Di Pasquale A, Cammà C, Puglia I, Calistri P, Savini G |
| EPI_ISL_833292, EPI_ISL_833293, EPI_ISL_833294 | SIESP CHIETI - DRIVE IN LANCIANO | Istituto Zooprofilattico Sperimentale dell'Abruzzo e Molise "G. Caporale" | Lorusso A, Marcacci M, Di Domenico M, Ancora M, Curini V, Mangone I, Rinaldi A, Di Pasquale A, Cammà C, Puglia I, Calistri P, Savini G |
| EPI_ISL_833295, EPI_ISL_833296 | SIESP DIPARTIMENTO DI PREVENZIONE TERAMO | Istituto Zooprofilattico Sperimentale dell'Abruzzo e Molise "G. Caporale" | Lorusso A, Marcacci M, Di Domenico M, Ancora M, Curini V, Mangone I, Rinaldi A, Di Pasquale A, Cammà C, Puglia I, Calistri P, Savini G |
| EPI_ISL_833297 | Ospedale Civile Giulianova | Istituto Zooprofilattico Sperimentale dell'Abruzzo e Molise "G. Caporale" | Lorusso A, Marcacci M, Di Domenico M, Ancora M, Curini V, Mangone I, Rinaldi A, Di Pasquale A, Cammà C, Puglia I, Calistri P, Savini G |
| EPI_ISL_833298 | SIESP CHIETI - DRIVE IN LANCIANO | Istituto Zooprofilattico Sperimentale dell'Abruzzo e Molise "G. Caporale" | Lorusso A, Marcacci M, Di Domenico M, Ancora M, Curini V, Mangone I, Rinaldi A, Di Pasquale A, Cammà C, Puglia I, Calistri P, Savini G |
| EPI_ISL_833299 | SIESP DIPARTIMENTO DI PREVENZIONE CHIETI | Istituto Zooprofilattico Sperimentale dell'Abruzzo e Molise "G. Caporale" | Lorusso A, Marcacci M, Di Domenico M, Ancora M, Curini V, Mangone I, Rinaldi A, Di Pasquale A, Cammà C, Puglia I, Calistri P, Savini G |
| EPI_ISL_833300 | SIESP CHIETI - DRIVE IN LANCIANO | Istituto Zooprofilattico Sperimentale dell'Abruzzo e Molise "G. Caporale" | Lorusso A, Marcacci M, Di Domenico M, Ancora M, Curini V, Mangone I, Rinaldi A, Di Pasquale A, Cammà C, Puglia I, Calistri P, Savini G |
| EPI_ISL_833301 | DIPARTIMENTO PREVENZIONE AVEZZANO-SERVIZIO DI IGIENE EPIDEMIOLOGIA E SANITA' PUBBLICA | Istituto Zooprofilattico Sperimentale dell'Abruzzo e Molise "G. Caporale" | Lorusso A, Marcacci M, Di Domenico M, Ancora M, Curini V, Mangone I, Rinaldi A, Di Pasquale A, Cammà C, Puglia I, Calistri P, Savini G |
| EPI_ISL_833302, EPI_ISL_833303, EPI_ISL_833304, EPI_ISL_833305 | SIESP DIPARTIMENTO DI PREVENZIONE TERAMO | Istituto Zooprofilattico Sperimentale dell'Abruzzo e Molise "G. Caporale" | Lorusso A, Marcacci M, Di Domenico M, Ancora M, Curini V, Mangone I, Rinaldi A, Di Pasquale A, Cammà C, Puglia I, Calistri P, Savini G |
| EPI_ISL_833306, EPI_ISL_833307, EPI_ISL_833308, EPI_ISL_833309, EPI_ISL_833310 | SIESP CHIETI - DRIVE IN ORTONA | Istituto Zooprofilattico Sperimentale dell'Abruzzo e Molise "G. Caporale" | Lorusso A, Marcacci M, Di Domenico M, Ancora M, Curini V, Mangone I, Rinaldi A, Di Pasquale A, Cammà C, Puglia I, Calistri P, Savini G |
| EPI_ISL_833311, EPI_ISL_833312 | SIESP CHIETI- DRIVE IN CHIETI | Istituto Zooprofilattico Sperimentale dell'Abruzzo e Molise "G. Caporale" | Lorusso A, Marcacci M, Di Domenico M, Ancora M, Curini V, Mangone I, Rinaldi A, Di Pasquale A, Cammà C, Puglia I, Calistri P, Savini G |
| EPI_ISL_833313, EPI_ISL_833314, EPI_ISL_833315 | SIESP CHIETI - DRIVE IN LANCIANO | Istituto Zooprofilattico Sperimentale dell'Abruzzo e Molise "G. Caporale" | Lorusso A, Marcacci M, Di Domenico M, Ancora M, Curini V, Mangone I, Rinaldi A, Di Pasquale A, Cammà C, Puglia I, Calistri P, Savini G |
| EPI_ISL_833316 | SIESP CHIETI - DRIVE IN GISSI | Istituto Zooprofilattico Sperimentale dell'Abruzzo e Molise "G. Caporale" | Lorusso A, Marcacci M, Di Domenico M, Ancora M, Curini V, Mangone I, Rinaldi A, Di Pasquale A, Cammà C, Puglia I, Calistri P, Savini G |
| EPI_ISL_833317, EPI_ISL_833318 | SIESP CHIETI - DRIVE IN CHIETI | Istituto Zooprofilattico Sperimentale dell'Abruzzo e Molise "G. Caporale" | Lorusso A, Marcacci M, Di Domenico M, Ancora M, Curini V, Mangone I, Rinaldi A, Di Pasquale A, Cammà C, Puglia I, Calistri P, Savini G |
| EPI_ISL_833319, EPI_ISL_833320 | SIESP DIPARTIMENTO DI PREVENZIONE SULMONA | Istituto Zooprofilattico Sperimentale dell'Abruzzo e Molise "G. Caporale" | Lorusso A, Marcacci M, Di Domenico M, Ancora M, Curini V, Mangone I, Rinaldi A, Di Pasquale A, Cammà C, Puglia I, Calistri P, Savini G |
| EPI_ISL_833321 | SIESP DIPARTIMENTO DI PREVENZIONE CHIETI | Istituto Zooprofilattico Sperimentale dell'Abruzzo e Molise "G. Caporale" | Lorusso A, Marcacci M, Di Domenico M, Ancora M, Curini V, Mangone I, Rinaldi A, Di Pasquale A, Cammà C, Puglia I, Calistri P, Savini G |
| EPI_ISL_833322 | SIESP DIPARTIMENTO DI PREVENZIONE SULMONA | Istituto Zooprofilattico Sperimentale dell'Abruzzo e Molise "G. Caporale" | Lorusso A, Marcacci M, Di Domenico M, Ancora M, Curini V, Mangone I, Rinaldi A, Di Pasquale A, Cammà C, Puglia I, Calistri P, Savini G |
| EPI_ISL_833323 | SIESP CHIETI - DRIVE IN CHIETI | Istituto Zooprofilattico Sperimentale dell'Abruzzo e Molise "G. Caporale" | Lorusso A, Marcacci M, Di Domenico M, Ancora M, Curini V, Mangone I, Rinaldi A, Di Pasquale A, Cammà C, Puglia I, Calistri P, Savini G |
| EPI_ISL_833324 | SIESP DIPARTIMENTO DI PREVENZIONE CHIETI | Istituto Zooprofilattico Sperimentale dell'Abruzzo e Molise "G. Caporale" | Lorusso A, Marcacci M, Di Domenico M, Ancora M, Curini V, Mangone I, Rinaldi A, Di Pasquale A, Cammà C, Puglia I, Calistri P, Savini G |
| EPI_ISL_833325 | Ospedale SS Annunziata Chieti | Istituto Zooprofilattico Sperimentale dell'Abruzzo e Molise "G. Caporale" | Lorusso A, Marcacci M, Di Domenico M, Ancora M, Curini V, Mangone I, Rinaldi A, Di Pasquale A, Cammà C, Puglia I, Calistri P, Savini G |
| EPI_ISL_833326 | SIESP DIPARTIMENTO DI PREVENZIONE CHIETI | Istituto Zooprofilattico Sperimentale dell'Abruzzo e Molise "G. Caporale" | Lorusso A, Marcacci M, Di Domenico M, Ancora M, Curini V, Mangone I, Rinaldi A, Di Pasquale A, Cammà C, Puglia I, Calistri P, Savini G |
| EPI_ISL_833327 | Ospedale SS Annunziata Chieti | Istituto Zooprofilattico Sperimentale dell'Abruzzo e Molise "G. Caporale" | Lorusso A, Marcacci M, Di Domenico M, Ancora M, Curini V, Mangone I, Rinaldi A, Di Pasquale A, Cammà C, Puglia I, Calistri P, Savini G |
| EPI_ISL_833328, EPI_ISL_833329, EPI_ISL_833330 | SIESP CHIETI - DRIVE IN ORTONA | Istituto Zooprofilattico Sperimentale dell'Abruzzo e Molise "G. Caporale" | Lorusso A, Marcacci M, Di Domenico M, Ancora M, Curini V, Mangone I, Rinaldi A, Di Pasquale A, Cammà C, Puglia I, Calistri P, Savini G |

| | | | |
|---|---|---|---|
| EPI_ISL_833331 | Ospedale Civile Atri | Istituto Zooprofilattico Sperimentale dell'Abruzzo e Molise "G. Caporale" | Lorusso A, Marcacci M, Di Domenico M, Ancora M, Curini V, Mangone I, Rinaldi A, Di Pasquale A, Cammà C, Puglia I, Calistri P, Savini G |
| EPI_ISL_833435, EPI_ISL_833436, EPI_ISL_833437, EPI_ISL_833438, EPI_ISL_833439, EPI_ISL_833440, EPI_ISL_833441, EPI_ISL_833442, EPI_ISL_833443, EPI_ISL_833444, EPI_ISL_833445, EPI_ISL_833446, EPI_ISL_833447, EPI_ISL_833448, EPI_ISL_833449, EPI_ISL_833450, EPI_ISL_833451, EPI_ISL_833452, EPI_ISL_833453, EPI_ISL_833454, EPI_ISL_833455 | | | |
| see above | SC (UCO) Igiene e Sanità Pubblica (funzione integrata con SC Microbiologia e Virologia) e Laboratory of Molecular Virology of the International Centre for Genetic Engineering and Biotechnology (ICGEB) | ARGO Laboratorio Genomica ed Epigenomica | Licastro D, Dal Monego S, Degasperi M, Marcello A, D'Agaro P |
| EPI_ISL_833522, EPI_ISL_833523, EPI_ISL_833524, EPI_ISL_833525, EPI_ISL_833526, EPI_ISL_833527, EPI_ISL_833528, EPI_ISL_833529, EPI_ISL_833530, EPI_ISL_833531, EPI_ISL_833532, EPI_ISL_833533, EPI_ISL_833534, EPI_ISL_833535, EPI_ISL_833536, EPI_ISL_833537, EPI_ISL_833538, EPI_ISL_833539, EPI_ISL_833540, EPI_ISL_833541, EPI_ISL_833542, EPI_ISL_833543, EPI_ISL_833544, EPI_ISL_833545, EPI_ISL_833546, EPI_ISL_833547, EPI_ISL_833548, EPI_ISL_833549, EPI_ISL_833550, EPI_ISL_833551, EPI_ISL_833552, EPI_ISL_833553, EPI_ISL_833554, EPI_ISL_833555, EPI_ISL_833556, EPI_ISL_833557, EPI_ISL_833558, EPI_ISL_833559, EPI_ISL_833560, EPI_ISL_833561, EPI_ISL_833562, EPI_ISL_833563, EPI_ISL_833564, EPI_ISL_833565, EPI_ISL_833566, EPI_ISL_833567, EPI_ISL_833568, EPI_ISL_833569, EPI_ISL_833570, EPI_ISL_837248, EPI_ISL_837249, EPI_ISL_837250, EPI_ISL_837251, EPI_ISL_837252, EPI_ISL_837253, EPI_ISL_837254, EPI_ISL_837255, EPI_ISL_837256, EPI_ISL_837257, EPI_ISL_837258, EPI_ISL_837259, EPI_ISL_837260, EPI_ISL_837261, EPI_ISL_837262, EPI_ISL_837263, EPI_ISL_837264, EPI_ISL_837265, EPI_ISL_837266, EPI_ISL_837267, EPI_ISL_837268, EPI_ISL_837269, EPI_ISL_837270, EPI_ISL_837271, EPI_ISL_837272, EPI_ISL_837273, EPI_ISL_837274, EPI_ISL_837275, EPI_ISL_837276, EPI_ISL_837277, EPI_ISL_837278, EPI_ISL_837279, EPI_ISL_837280, EPI_ISL_837281, EPI_ISL_837282, EPI_ISL_837283, EPI_ISL_837284, EPI_ISL_837285, EPI_ISL_837286, EPI_ISL_837287, EPI_ISL_837288, EPI_ISL_837289, EPI_ISL_837290, EPI_ISL_837291, EPI_ISL_837292, EPI_ISL_837293, EPI_ISL_837294, EPI_ISL_837295, EPI_ISL_837296, EPI_ISL_837297, EPI_ISL_837298, EPI_ISL_837299, EPI_ISL_837300, EPI_ISL_837301, EPI_ISL_837302, EPI_ISL_837303, EPI_ISL_837304, EPI_ISL_837305, EPI_ISL_837306, EPI_ISL_837307, EPI_ISL_837308, EPI_ISL_837309, EPI_ISL_837310, EPI_ISL_837311, EPI_ISL_837312, EPI_ISL_837313, EPI_ISL_837314, EPI_ISL_837315, EPI_ISL_837316, EPI_ISL_837317, EPI_ISL_837318, EPI_ISL_837319, EPI_ISL_837320, EPI_ISL_837321, EPI_ISL_837322, EPI_ISL_837323, EPI_ISL_837324, EPI_ISL_837325, EPI_ISL_837326, EPI_ISL_837327, EPI_ISL_837328, EPI_ISL_837329, EPI_ISL_837330, EPI_ISL_837433, EPI_ISL_837434, EPI_ISL_837435, EPI_ISL_837436, EPI_ISL_837437, EPI_ISL_837438, EPI_ISL_837439, EPI_ISL_837440, EPI_ISL_837441, EPI_ISL_837442, EPI_ISL_837443, EPI_ISL_837444, EPI_ISL_837445, EPI_ISL_837446, EPI_ISL_837447, EPI_ISL_837448, EPI_ISL_837449, EPI_ISL_837450, EPI_ISL_837451, EPI_ISL_837452, EPI_ISL_837453, EPI_ISL_837454, EPI_ISL_837455, EPI_ISL_837456, EPI_ISL_837457, EPI_ISL_837458, EPI_ISL_837459, EPI_ISL_837460, EPI_ISL_837461, EPI_ISL_837462, EPI_ISL_837463, EPI_ISL_837464, EPI_ISL_837465, EPI_ISL_837466, EPI_ISL_837467, EPI_ISL_837468, EPI_ISL_837469, EPI_ISL_837470, EPI_ISL_837471, EPI_ISL_837472, EPI_ISL_837473, EPI_ISL_837474, EPI_ISL_837475, EPI_ISL_837476, EPI_ISL_837477, EPI_ISL_837478, EPI_ISL_837479, EPI_ISL_837480, EPI_ISL_837481, EPI_ISL_837482, EPI_ISL_837483, EPI_ISL_837484, EPI_ISL_837485, EPI_ISL_837486, EPI_ISL_837487, EPI_ISL_837488, EPI_ISL_837489, EPI_ISL_837490, EPI_ISL_837491, EPI_ISL_837492, EPI_ISL_837493, EPI_ISL_837494, EPI_ISL_837495, EPI_ISL_837496, EPI_ISL_837497, EPI_ISL_837498, EPI_ISL_837499, EPI_ISL_837500, EPI_ISL_837501, EPI_ISL_837502, EPI_ISL_837503, EPI_ISL_837504 | | | |
| see above | Istituto Zooprofilattico Sperimentale del Mezzogiorno | TIGEM | Antonio Grimaldi, Patrizia Annunziata, Francesco Panariello, Biancamaria Pierri, Valentina Bouche, Chiara Colantuono, Maria Concetta Cuomo, Denise Di Concilio, Lucio Di Filippo, Anna Manfredi, Marcello Salvi, Antonio Limone, Pellegrino Cerino, Andrea Ballabio, Davide Cacchiarelli. |
| EPI_ISL_847825 | Lab. Microbiologia e Virologia, Cotugno, A.O. dei Colli | Lab. Microbiologia e Virologia, Cotugno, A.O. dei Colli | Luigi Atripaldi, Claudia Tiberio, Anna Perfetti |
| EPI_ISL_849634 | SIESP DIPARTIMENTO DI PREVENZIONE CHIETI | Istitituto Zooprofilattico Sperimentale dell'Abruzzo e Molise "G.Caporale" | Lorusso A, Marcacci M, Di Domenico M, Curini V, Ancora M, Cammà C, Rinaldi A, Mangone I, Di Pasquale A, Puglia I, Savini G. |
| EPI_ISL_849635, EPI_ISL_849636 | DIPARTIMENTO PREVENZIONE AVEZZANO-SERVIZIO DI IGIENE EPIDEMIOLOGIA E SANITA' PUBBLICA | Istitituto Zooprofilattico Sperimentale dell'Abruzzo e Molise "G.Caporale" | Lorusso A, Marcacci M, Di Domenico M, Curini V, Ancora M, Cammà C, Rinaldi A, Mangone I, Di Pasquale A, Puglia I, Savini G. |
| EPI_ISL_849637, EPI_ISL_849638, EPI_ISL_849639, EPI_ISL_849640, EPI_ISL_849641, EPI_ISL_849642, EPI_ISL_849643, EPI_ISL_849644, EPI_ISL_849645, EPI_ISL_849646 | SIESP DIPARTIMENTO DI PREVENZIONE CHIETI | Istitituto Zooprofilattico Sperimentale dell'Abruzzo e Molise "G.Caporale" | Lorusso A, Marcacci M, Di Domenico M, Curini V, Ancora M, Cammà C, Rinaldi A, Mangone I, Di Pasquale A, Puglia I, Savini G. |
| EPI_ISL_849647, EPI_ISL_849648, EPI_ISL_849649 | SIESP- DRIVE IN CHIETI | Istitituto Zooprofilattico Sperimentale dell'Abruzzo e Molise "G.Caporale" | Lorusso A, Marcacci M, Di Domenico M, Curini V, Ancora M, Cammà C, Rinaldi A, Mangone I, Di Pasquale A, Puglia I, Savini G. |
| EPI_ISL_849650, EPI_ISL_849651, EPI_ISL_849652 | Servizio di igiene epidemiologia e sanità pubblica (SIESP)-Chieti | Istitituto Zooprofilattico Sperimentale dell'Abruzzo e Molise "G.Caporale" | Lorusso A, Marcacci M, Di Domenico M, Curini V, Ancora M, Cammà C, Rinaldi A, Mangone I, Di Pasquale A, Puglia I, Savini G. |
| EPI_ISL_849653 | Servizio di igiene e sanità pubblica (SIESP)-Teramo | Istitituto Zooprofilattico Sperimentale dell'Abruzzo e Molise "G.Caporale" | Lorusso A, Marcacci M, Di Domenico M, Curini V, Ancora M, Cammà C, Rinaldi A, Mangone I, Di Pasquale A, Puglia I, Savini G. |
| EPI_ISL_849654 | Servizio Igiene Epidemiologia e Sanità Pubblica (SIESP)-L'Aquila | Istitituto Zooprofilattico Sperimentale dell'Abruzzo e Molise "G.Caporale" | Lorusso A, Marcacci M, Di Domenico M, Curini V, Ancora M, Cammà C, Rinaldi A, Mangone I, Di Pasquale A, Puglia I, Savini G. |
| EPI_ISL_849655 | Servizio di igiene e sanità pubblica (SIESP)-Teramo | Istitituto Zooprofilattico Sperimentale dell'Abruzzo e Molise "G.Caporale" | Lorusso A, Marcacci M, Di Domenico M, Curini V, Ancora M, Cammà C, Rinaldi A, Mangone I, Di Pasquale A, Puglia I, Savini G. |
| EPI_ISL_849656, EPI_ISL_849657, EPI_ISL_849658, EPI_ISL_849659, EPI_ISL_849660, EPI_ISL_849661 | Servizio Igiene Epidemiologia e Sanità Pubblica (SIESP)-L'Aquila | Istitituto Zooprofilattico Sperimentale dell'Abruzzo e Molise "G.Caporale" | Lorusso A, Marcacci M, Di Domenico M, Curini V, Ancora M, Cammà C, Rinaldi A, Mangone I, Di Pasquale A, Puglia I, Savini G. |
| EPI_ISL_849662, EPI_ISL_849663 | Servizio di igiene epidemiologia e sanità pubblica (SIESP)-Chieti | Istitituto Zooprofilattico Sperimentale dell'Abruzzo e Molise "G.Caporale" | Lorusso A, Marcacci M, Di Domenico M, Curini V, Ancora M, Cammà C, Rinaldi A, Mangone I, Di Pasquale A, Puglia I, Savini G. |
| EPI_ISL_849763 | Molecular Biology, IZS Sicilia | Molecular Biology, IZS Sicilia | Reale,S., Seidita,G., Di Gaudio,F., Scibetta,S. and Vitale,F. |
| EPI_ISL_852619, EPI_ISL_852622, EPI_ISL_852628, EPI_ISL_852629, EPI_ISL_852630, EPI_ISL_852631, EPI_ISL_852632, EPI_ISL_852813, EPI_ISL_852819, EPI_ISL_852821, EPI_ISL_852823, EPI_ISL_852825, EPI_ISL_852828 | | | |
| see above | Lab. Microbiologia e Virologia, Cotugno, A.O. dei Colli | Lab. Microbiologia e Virologia, Cotugno, A.O. dei Colli | Luigi Atripaldi, Claudia Tiberio, Anna Perfetti |
| EPI_ISL_854303 | Pharmgenetix GmbH | Bergthaler laboratory, CeMM Research Center for Molecular Medicine of the Austrian Academy of Sciences | Lukas Endler, Alexandra Popa, Benedikt Agerer, Jakob-Wendelin Genger, Alexander Lercher, Anna Schedl, Thomas Penz, Michael Schuster, Jan Laine, Martin Senekowitsch, Christoph Bock, Andreas Bergthaler |
| EPI_ISL_855550 | Department of Infectious Diseases, Istituto Superiore di Sanità, Rome, Italy | Istituto Superiore di Sanità (ISS) | Paola Stefanelli, Angela Di Martino, Alessandra Lo Presti, Stefano Fiore, Gabriele Vaccari, Luca De Sabato, Ilaria Di Bartolo, Giovanni Ianiro, Manuela Marra, Maria Carollo, Marco Crescenzi |
| EPI_ISL_855552 | Department of Infectious Diseases, Istituto Superiore di Sanità, Rome, Italy; Ospedali Riuniti, Laboratorio Virologia, Ancona, Italy | Istituto Superiore di Sanità (ISS) | Paola Stefanelli, Angela Di Martino, Alessandra Lo Presti, Stefano Fiore, Patrizia Bagnarelli, Gabriele Vaccari, Luca De Sabato, Ilaria Di Bartolo, Giovanni Ianiro, Manuela Marra, Maria Carollo, Marco Crescenzi |
| EPI_ISL_855553 | Department of Infectious Diseases, Istituto Superiore di Sanità, Rome, Italy | Istituto Superiore di Sanità (ISS) | Paola Stefanelli, Angela Di Martino, Alessandra Lo Presti, Stefano Fiore, Gabriele Vaccari, Luca De Sabato, Ilaria Di Bartolo, Giovanni Ianiro, Manuela Marra, Maria Carollo, Marco Crescenzi |
| EPI_ISL_855554, EPI_ISL_855555 | Department of Infectious Diseases, Istituto Superiore di Sanità, Rome, Italy; ASST G.O.M. Niguarda, Sezione Microbiologia e Virologia, Milano, Italy | Istituto Superiore di Sanità (ISS) | Paola Stefanelli, Angela Di Martino, Alessandra Lo Presti, Stefano Fiore, Carlo Federico Perno, Gabriele Vaccari, Luca De Sabato, Ilaria Di Bartolo, Giovanni Ianiro, Manuela Marra, Maria Carollo, Marco Crescenzi |
| EPI_ISL_855556, EPI_ISL_855573, EPI_ISL_855610, EPI_ISL_855611, EPI_ISL_855612, EPI_ISL_855613, EPI_ISL_855614, EPI_ISL_856679, EPI_ISL_856682, EPI_ISL_856694, EPI_ISL_856698 | | | |
| see above | Lab. Microbiologia e Virologia, Cotugno, A.O. dei Colli | Lab. Microbiologia e Virologia, Cotugno, A.O. dei Colli | Luigi Atripaldi, Claudia Tiberio, Anna Perfetti |
| EPI_ISL_856712, EPI_ISL_856713, EPI_ISL_856714, EPI_ISL_856715, EPI_ISL_856716, EPI_ISL_856717, EPI_ISL_856718, EPI_ISL_856719, EPI_ISL_856720, EPI_ISL_856721 | Infectious Diseases Unit, Department of Internal Medicine, Azienda Ospedaliera-Universitaria di Padova | Laboratory of Infectious Diseases, Department of Biomedical and Clinical Sciences L. Sacco, University of Milan | Anna Maria Cattelan, Lolita Sasset, Davide Leoni, Alessia Lai, Annalisa Bergna, Carla Della Ventura, Claudia Balotta, Massimo Galli, Gianguglielmo Zehender on behalf of SARS-CoV-2 ITALIAN RESEARCH ENTERPRISE-(SCIRE) Collaborative Group |
| EPI_ISL_856869 | Virology Laboratory, Scientific Department, Army Medical Center | Virology Laboratory, Scientific Department, Army Medical Center | Silvia Fillo, Giovanni Faggioni, Riccardo De Sanctis, Antonella Fortunato, Anella Monte, Anna Anselmo, Vanessa Vera Fain, Francesco Giordani, Nino D'Amore, Filippo Molinari, Giancarlo Petralito, Florigio Lista |
| EPI_ISL_856870 | Virology Laboratory, Scientific Department, Army Medical Center | Virology Laboratory, Scientific Department, Army Medical Center | Silvia Fillo, Giovanni Faggioni, Riccardo De Sanctis, Antonella Fortunato, Anella Monte, Anna Anselmo, Vanessa Vera Fain, Francesco Giordani, Nino D'Amore, Filippo Molinari, Giancarlo Petralito, Florigio Lista |

| | | | |
|---|---|---|---|
| EPI_ISL_856871 | Department of Infectious Diseases, Istituto Superiore di Sanità, Roma, Italy; AORN dei Colli, UOC Microbiologia Virologia, Napoli, Italy | Virology Laboratory, Scientific Department, Army Medical Center | Paola Stefanelli, Angela Di Martino, Alessandra Lo Presti, Stefano Fiore, Luigi Atripaldi, Silvia Fillo, Giovanni Faggioni, Riccardo De Sanctis, Antonella Fortunato, Anna Anselmo, Francesco Giordani, Vanessa Vera Fain, Nino D'Amore, Florigio Lista |
| EPI_ISL_856872, EPI_ISL_856873, EPI_ISL_856874, EPI_ISL_856875 | Virology Laboratory, Scientific Department, Army Medical Center | Virology Laboratory, Scientific Department, Army Medical Center | Silvia Fillo, Giovanni Faggioni, Riccardo De Sanctis, Antonella Fortunato, Anella Monte, Anna Anselmo, Vanessa Vera Fain, Francesco Giordani, Nino D'Amore, Filippo Molinari, Giancarlo Petralito, Florigio Lista |
| EPI_ISL_856876 | Virology Laboratory, Scientific Department, Army Medical Center | Virology Laboratory, Scientific Department, Army Medical Center | Silvia Fillo, Giovanni Faggioni, Riccardo De Sanctis, Antonella Fortunato, Anella Monte, Anna Anselmo, Vanessa Vera Fain, Francesco Giordani, Nino D'Amore, Filippo Molinari, Giancarlo Petralito, Florigio Lista |
| EPI_ISL_856877, EPI_ISL_856878 | Virology Laboratory, Scientific Department, Army Medical Center | Virology Laboratory, Scientific Department, Army Medical Center | Silvia Fillo, Giovanni Faggioni, Riccardo De Sanctis, Antonella Fortunato, Anella Monte, Anna Anselmo, Vanessa Vera Fain, Francesco Giordani, Nino D'Amore, Filippo Molinari, Giancarlo Petralito, Florigio Lista |
| EPI_ISL_856879 | Virology Laboratory, Scientific Department, Army Medical Center | Virology Laboratory, Scientific Department, Army Medical Center | Silvia Fillo, Giovanni Faggioni, Riccardo De Sanctis, Antonella Fortunato, Anella Monte, Anna Anselmo, Vanessa Vera Fain, Francesco Giordani, Nino D'Amore, Filippo Molinari, Giancarlo Petralito, Florigio Lista |
| EPI_ISL_856880 | Department of Infectious Diseases, Istituto Superiore di Sanità, Roma, Italy; Ospedali Riuniti, Laboratorio Virologia, Ancona, Italy | Virology Laboratory, Scientific Department, Army Medical Center | Paola Stefanelli, Angela Di Martino, Alessandra Lo Presti, Stefano Fiore, Patrizia Bagnarelli, Silvia Fillo, Giovanni Faggioni, Riccardo De Sanctis, Antonella Fortunato, Anna Anselmo, Francesco Giordani, Vanessa Vera Fain, Nino D'Amore, Florigio Lista |
| EPI_ISL_856881 | Department of Infectious Diseases, Istituto Superiore di Sanità, Roma, Italy; PO Cardarelli, Laboratorio Analisi Microbiologia e Virologia, Campobasso, Italy | Virology Laboratory, Scientific Department, Army Medical Center | Paola Stefanelli, Angela Di Martino, Alessandra Lo Presti, Stefano Fiore, Massimiliano Scutellà , Silvia Fillo, Giovanni Faggioni, Riccardo De Sanctis, Antonella Fortunato, Anna Anselmo, Francesco Giordani, Vanessa Vera Fain, Nino D'Amore, Florigio Lista |
| EPI_ISL_856882 | Virology Laboratory, Scientific Department, Army Medical Center | Virology Laboratory, Scientific Department, Army Medical Center | Silvia Fillo, Giovanni Faggioni, Riccardo De Sanctis, Antonella Fortunato, Anella Monte, Anna Anselmo, Vanessa Vera Fain, Francesco Giordani, Nino D'Amore, Filippo Molinari, Giancarlo Petralito, Florigio Lista |
| EPI_ISL_856883 | Virology Laboratory, Scientific Department, Army Medical Center | Virology Laboratory, Scientific Department, Army Medical Center | Silvia Fillo, Giovanni Faggioni, Riccardo De Sanctis, Antonella Fortunato, Anella Monte, Anna Anselmo, Vanessa Vera Fain, Francesco Giordani, Nino D'Amore, Filippo Molinari, Giancarlo Petralito, Florigio Lista |
| EPI_ISL_856884 | Department of Infectious Diseases, Istituto Superiore di Sanità, Roma, Italy; PO Santa Chiara, Microbiologia e Virologia, Trento, Italy | Virology Laboratory, Scientific Department, Army Medical Center | Paola Stefanelli, Angela Di Martino, Alessandra Lo Presti, Stefano Fiore, Paolo Lanzafame , Silvia Fillo, Giovanni Faggioni, Riccardo De Sanctis, Antonella Fortunato, Anna Anselmo, Francesco Giordani, Vanessa Vera Fain, Nino D'Amore, Florigio Lista |
| EPI_ISL_856885 | Virology Laboratory, Scientific Department, Army Medical Center | Virology Laboratory, Scientific Department, Army Medical Center | Silvia Fillo, Giovanni Faggioni, Riccardo De Sanctis, Antonella Fortunato, Anella Monte, Anna Anselmo, Vanessa Vera Fain, Francesco Giordani, Nino D'Amore, Filippo Molinari, Giancarlo Petralito, Florigio Lista |
| EPI_ISL_856886 | Department of Infectious Diseases, Istituto Superiore di Sanità, Roma, Italy; PO Spirito Santo, UOC Microbiologia e Virologia Clinica, Pescara, Italy | Virology Laboratory, Scientific Department, Army Medical Center | Paola Stefanelli, Angela Di Martino, Alessandra Lo Presti, Stefano Fiore, Paolo Fazii, Silvia Fillo, Giovanni Faggioni, Riccardo De Sanctis, Antonella Fortunato, Anna Anselmo, Francesco Giordani, Vanessa Vera Fain, Nino D'Amore, Florigio Lista |
| EPI_ISL_856887 | Department of Infectious Diseases, Istituto Superiore di Sanità, Roma, Italy; AUOC Policlinico Bari, UOC Igiene, Bari, Italy | Virology Laboratory, Scientific Department, Army Medical Center | Paola Stefanelli, Angela Di Martino, Alessandra Lo Presti, Stefano Fiore, Maria Chironna, Silvia Fillo, Giovanni Faggioni, Riccardo De Sanctis, Antonella Fortunato, Anna Anselmo, Francesco Giordani, Vanessa Vera Fain, Nino D'Amore, Florigio Lista |
| EPI_ISL_856888 | Department of Infectious Diseases, Istituto Superiore di Sanità, Roma, Italy; A.O.R. San Carlo di Potenza, Laboratorio Microbiologia e Virologia, Potenza, Italy | Virology Laboratory, Scientific Department, Army Medical Center | Paola Stefanelli, Angela Di Martino, Alessandra Lo Presti, Stefano Fiore, Antonio Picerno, Silvia Fillo, Giovanni Faggioni, Riccardo De Sanctis, Antonella Fortunato, Anna Anselmo, Francesco Giordani, Vanessa Vera Fain, Nino D'Amore, Florigio Lista |
| EPI_ISL_856889 | Department of Infectious Diseases, Istituto Superiore di Sanità, Roma, Italy | Virology Laboratory, Scientific Department, Army Medical Center | Paola Stefanelli, Angela Di Martino, Alessandra Lo Presti, Stefano Fiore, Silvia Fillo, Giovanni Faggioni, Riccardo De Sanctis, Antonella Fortunato, Anna Anselmo, Francesco Giordani, Vanessa Vera Fain, Nino D'Amore, Florigio Lista |
| EPI_ISL_856890 | Department of Infectious Diseases, Istituto Superiore di Sanità, Roma, Italy; AO Annunziata, UOC Microbiologia e Virologia, Cosenza, Italy | Virology Laboratory, Scientific Department, Army Medical Center | Paola Stefanelli, Angela Di Martino, Alessandra Lo Presti, Stefano Fiore, Cristina Giraldi, Silvia Fillo, Giovanni Faggioni, Riccardo De Sanctis, Antonella Fortunato, Anna Anselmo, Francesco Giordani, Vanessa Vera Fain, Nino D'Amore, Florigio Lista |
| EPI_ISL_856891, EPI_ISL_856892 | Department of Infectious Diseases, Istituto Superiore di Sanità, Roma, Italy; AORN dei Colli, UOC Microbiologia Virologia, Napoli, Italy | Virology Laboratory, Scientific Department, Army Medical Center | Paola Stefanelli, Angela Di Martino, Alessandra Lo Presti, Stefano Fiore, Luigi Atripaldi, Silvia Fillo, Giovanni Faggioni, Riccardo De Sanctis, Antonella Fortunato, Anna Anselmo, Francesco Giordani, Vanessa Vera Fain, Nino D'Amore, Florigio Lista |
| EPI_ISL_856893, EPI_ISL_856894 | Department of Infectious Diseases, Istituto Superiore di Sanità, Roma, Italy; ASUITS - Ospedale Maggiore, S.C. UOC Igiene e Sanità Pubblica, Trieste, Italy | Virology Laboratory, Scientific Department, Army Medical Center | Paola Stefanelli, Angela Di Martino, Alessandra Lo Presti, Stefano Fiore, Pierlanfranco D'Agaro, Silvia Fillo, Giovanni Faggioni, Riccardo De Sanctis, Antonella Fortunato, Anna Anselmo, Francesco Giordani, Vanessa Vera Fain, Nino D'Amore, Florigio Lista |
| EPI_ISL_856895, EPI_ISL_856896 | Department of Infectious Diseases, Istituto Superiore di Sanità, Roma, Italy; Ospedali Riuniti, Laboratorio Virologia, Ancona, Italy | Virology Laboratory, Scientific Department, Army Medical Center | Paola Stefanelli, Angela Di Martino, Alessandra Lo Presti, Stefano Fiore, Patrizia Bagnarelli, Silvia Fillo, Giovanni Faggioni, Riccardo De Sanctis, Antonella Fortunato, Anna Anselmo, Francesco Giordani, Vanessa Vera Fain, Nino D'Amore, Florigio Lista |
| EPI_ISL_856897 | Department of Infectious Diseases, Istituto Superiore di Sanità, Roma, Italy; PO Cardarelli, Laboratorio Analisi Microbiologia e Virologia, Campobasso, Italy | Virology Laboratory, Scientific Department, Army Medical Center | Paola Stefanelli, Angela Di Martino, Alessandra Lo Presti, Stefano Fiore, Massimiliano Scutellà, Silvia Fillo, Giovanni Faggioni, Riccardo De Sanctis, Antonella Fortunato, Anna Anselmo, Francesco Giordani, Vanessa Vera Fain, Nino D'Amore, Florigio Lista |
| EPI_ISL_856898, EPI_ISL_856899 | Department of Infectious Diseases, Istituto Superiore di Sanità, Roma, Italy; ASL Città di Torino, S.C. Microbiologia e Virologia, Torino, Italy | Virology Laboratory, Scientific Department, Army Medical Center | Paola Stefanelli, Angela Di Martino, Alessandra Lo Presti, Stefano Fiore, Valeria Ghisetti, Silvia Fillo, Giovanni Faggioni, Riccardo De Sanctis, Antonella Fortunato, Anna Anselmo, Francesco Giordani, Vanessa Vera Fain, Nino D'Amore, Florigio Lista |
| EPI_ISL_856900, EPI_ISL_856901 | Department of Infectious Diseases, Istituto Superiore di Sanità, Roma, Italy; AOUP V. Emanuele di Catania, P.O. Gaspare Rodolico, Catania, Italy | Virology Laboratory, Scientific Department, Army Medical Center | Paola Stefanelli, Angela Di Martino, Alessandra Lo Presti, Stefano Fiore, Guido Scalia, Silvia Fillo, Giovanni Faggioni, Riccardo De Sanctis, Antonella Fortunato, Anna Anselmo, Francesco Giordani, Vanessa Vera Fain, Nino D'Amore, Florigio Lista |
| EPI_ISL_856902 | Department of Infectious Diseases, Istituto Superiore di Sanità, Roma, Italy; AS Alto Adige, Microbiologia e Virologia, Bolzano, Italy | Virology Laboratory, Scientific Department, Army Medical Center | Paola Stefanelli, Angela Di Martino, Alessandra Lo Presti, Stefano Fiore, Elisabetta Pagani, Silvia Fillo, Giovanni Faggioni, Riccardo De Sanctis, Antonella Fortunato, Anna Anselmo, Francesco Giordani, Vanessa Vera Fain, Nino D'Amore, Florigio Lista |
| EPI_ISL_856903 | Department of Infectious Diseases, Istituto Superiore di Sanità, Roma, Italy; PO Santa Chiara, Microbiologia e Virologia, Trento, Italy | Virology Laboratory, Scientific Department, Army Medical Center | Paola Stefanelli, Angela Di Martino, Alessandra Lo Presti, Stefano Fiore, Paolo Lanzafame, Silvia Fillo, Giovanni Faggioni, Riccardo De Sanctis, Antonella Fortunato, Anna Anselmo, Francesco Giordani, Vanessa Vera Fain, Nino D'Amore, Florigio Lista |
| EPI_ISL_856904 | Department of Infectious Diseases, Istituto Superiore di Sanità, Roma, Italy | Virology Laboratory, Scientific Department, Army Medical Center | Paola Stefanelli, Angela Di Martino, Alessandra Lo Presti, Stefano Fiore, Silvia Fillo, Giovanni Faggioni, Riccardo De Sanctis, Antonella Fortunato, Anna Anselmo, Francesco Giordani, Vanessa Vera Fain, Nino D'Amore, Florigio Lista |
| EPI_ISL_856905, EPI_ISL_856906 | Department of Infectious Diseases, Istituto Superiore di Sanità, Roma, Italy; AO S.M. della Misericordia, S.C. Microbiologia, Perugia, Italy | Virology Laboratory, Scientific Department, Army Medical Center | Paola Stefanelli, Angela Di Martino, Alessandra Lo Presti, Stefano Fiore, Barbara Camilloni, Silvia Fillo, Giovanni Faggioni, Riccardo De Sanctis, Antonella Fortunato, Anna Anselmo, Francesco Giordani, Vanessa Vera Fain, Nino D'Amore, Florigio Lista |
| EPI_ISL_856907 | Department of Infectious Diseases, Istituto Superiore di Sanità, Roma, Italy; AUSL Valle d'Aosta, Aosta, Italy | Virology Laboratory, Scientific Department, Army Medical Center | Paola Stefanelli, Angela Di Martino, Alessandra Lo Presti, Stefano Fiore, Massimo Di Benedetto, Silvia Fillo, Giovanni Faggioni, Riccardo De Sanctis, Antonella Fortunato, Anna Anselmo, Francesco Giordani, Vanessa Vera Fain, Nino D'Amore, Florigio Lista |
| EPI_ISL_856908 | Department of Infectious Diseases, Istituto Superiore di Sanità, Roma, Italy; PO Madre Teresa di Calcutta, UOC. Laboratorio Analisi ULSS 6 Euganea, Padova, Italy | Virology Laboratory, Scientific Department, Army Medical Center | Paola Stefanelli, Angela Di Martino, Alessandra Lo Presti, Stefano Fiore, Giacomo Mezzapelle, Silvia Fillo, Giovanni Faggioni, Riccardo De Sanctis, Antonella Fortunato, Anna Anselmo, Francesco Giordani, Vanessa Vera Fain, Nino D'Amore, Florigio Lista |

| | | | |
|---|---|---|---|
| EPI_ISL_856909 | Department of Infectious Diseases, Istituto Superiore di Sanità, Roma, Italy; ULSS 8 Berica Vicenza, UOC Microbiologia, Vicenza, Italy | Virology Laboratory, Scientific Department, Army Medical Center | Paola Stefanelli, Angela Di Martino, Alessandra Lo Presti, Stefano Fiore, Mario Rassu, Silvia Fillo, Giovanni Faggioni, Riccardo De Sanctis, Antonella Fortunato, Anna Anselmo, Francesco Giordani, Vanessa Vera Fain, Nino D'Amore, Florigio Lista |
| EPI_ISL_869166 | Laboratory of Microbiology,ASST Settelaghi, Varese, Italy | Laboratory of Microbiology,ASST Settelaghi, Varese, Italy | Maggi,F., Novazzi,F., Genoni,A., Baj,A., Spezia,P.G., Focosi,D., Zago,C., Colombo,A., Cassani,G., Pasciuta,R., Tamborini,A.,Rossi,A., Prestia,M., Capuano,R., Azzi,L., Donadini,A., Catanoso,G., Grossi,P., Maffioli,L. and Bonelli,G. |
| EPI_ISL_869240 | Laboratory of Microbiology, ASST Settelaghi, Varese, Italy | Laboratory of Microbiology, ASST Settelaghi, Varese, Italy | Novazzi,F., Genoni,A., Focosi,D., Baj,A., Spezia,P.G., Zago,C., Colombo,A., Cassani,G., Pasciuta,R., Tamborini,A.,Rossi,A., Prestia,M., Capuano,R., Azzi,L., Donadini,A.,Catanoso,G., Maggi,F. |
| EPI_ISL_873209 | Medicine and Surgery, University of Insubria | Medicine and Surgery, University of Insubria | Maggi,F., Novazzi,F., Genoni,A., Baj,A., Spezia,P.G., Focosi,D.,Zago,C., Colombo,A., Cassani,G., Pasciuta,R., Tamborini,A.,Rossi,A., Prestia,M., Capuano,R., Azzi,L., Donadini,A.,Catanoso,G., Grossi,P., Maffioli,L. and Bonelli,G. |
| EPI_ISL_875566, EPI_ISL_875567, EPI_ISL_875568 | SIESP L'AQUILA | Istituto Zooprofilattico Sperimentale dell'Abruzzo e Molise "G. Caporale" | Lorusso A, Marcacci M, Di Domenico M, Ancora M, Curini V, Mangone I, Rinaldi A, Scialabba S, Di Pasquale A, Cammà C, Puglia I, Calistri P, Savini G |
| EPI_ISL_883155 | San Donato Arezzo Analysis Laboratory - Clinical Molecular Pathology sector | San Donato Arezzo Analysis Laboratory - Clinical Molecular Pathology sector | Alessandro Pancrazzi and Alice Moncada |
| EPI_ISL_883286, EPI_ISL_883287 | SIESP DIPARTIMENTO DI PREVENZIONE CHIE | Istituto Zooprofilattico Sperimentale dell'Abruzzo e Molise "G. Caporale" | Lorusso A, Marcacci M, Di Domenico M, Ancora M, Curini V, Mangone I, Rinaldi A, Scialabba S, Di Pasquale A, Cammà C, Puglia I, Calistri P, Savini G |
| EPI_ISL_883288, EPI_ISL_883289 | SIESP CHIETI-DRIVE IN ORTONA | Istituto Zooprofilattico Sperimentale dell'Abruzzo e Molise "G. Caporale" | Lorusso A, Marcacci M, Di Domenico M, Ancora M, Curini V, Mangone I, Rinaldi A, Scialabba S, Di Pasquale A, Cammà C, Puglia I, Calistri P, Savini G |
| EPI_ISL_883290 | SIESP CHIETI-DRIVE IN CHIETI | Istituto Zooprofilattico Sperimentale dell'Abruzzo e Molise "G. Caporale" | Lorusso A, Marcacci M, Di Domenico M, Ancora M, Curini V, Mangone I, Rinaldi A, Scialabba S, Di Pasquale A, Cammà C, Puglia I, Calistri P, Savini G |
| EPI_ISL_883291 | RP Guardiagrele-Ospedale di Comunità | Istituto Zooprofilattico Sperimentale dell'Abruzzo e Molise "G. Caporale" | Lorusso A, Marcacci M, Di Domenico M, Ancora M, Curini V, Mangone I, Rinaldi A, Scialabba S, Di Pasquale A, Cammà C, Puglia I, Calistri P, Savini G |
| EPI_ISL_883292 | SIESP CHIETI-DRIVE IN ORTONA | Istituto Zooprofilattico Sperimentale dell'Abruzzo e Molise "G. Caporale" | Lorusso A, Marcacci M, Di Domenico M, Ancora M, Curini V, Mangone I, Rinaldi A, Scialabba S, Di Pasquale A, Cammà C, Puglia I, Calistri P, Savini G |
| EPI_ISL_883293 | SIESP CHIETI-DRIVE IN LANCIANO | Istituto Zooprofilattico Sperimentale dell'Abruzzo e Molise "G. Caporale" | Lorusso A, Marcacci M, Di Domenico M, Ancora M, Curini V, Mangone I, Rinaldi A, Scialabba S, Di Pasquale A, Cammà C, Puglia I, Calistri P, Savini G |
| EPI_ISL_883294, EPI_ISL_883295 | RP Guardiagrele-Ospedale di Comunità | Istituto Zooprofilattico Sperimentale dell'Abruzzo e Molise "G. Caporale" | Lorusso A, Marcacci M, Di Domenico M, Ancora M, Curini V, Mangone I, Rinaldi A, Scialabba S, Di Pasquale A, Cammà C, Puglia I, Calistri P, Savini G |
| EPI_ISL_883296 | SIESP CHIETI-DRIVE IN ORTONA | Istituto Zooprofilattico Sperimentale dell'Abruzzo e Molise "G. Caporale" | Lorusso A, Marcacci M, Di Domenico M, Ancora M, Curini V, Mangone I, Rinaldi A, Scialabba S, Di Pasquale A, Cammà C, Puglia I, Calistri P, Savini G |
| EPI_ISL_883297 | RP Guardiagrele-Ospedale di Comunità | Istituto Zooprofilattico Sperimentale dell'Abruzzo e Molise "G. Caporale" | Lorusso A, Marcacci M, Di Domenico M, Ancora M, Curini V, Mangone I, Rinaldi A, Scialabba S, Di Pasquale A, Cammà C, Puglia I, Calistri P, Savini G |
| EPI_ISL_883298, EPI_ISL_883299 | SIESP DIPARTIMENTO DI PREVENZIONE CHIE | Istituto Zooprofilattico Sperimentale dell'Abruzzo e Molise "G. Caporale" | Lorusso A, Marcacci M, Di Domenico M, Ancora M, Curini V, Mangone I, Rinaldi A, Scialabba S, Di Pasquale A, Cammà C, Puglia I, Calistri P, Savini G |
| EPI_ISL_883300 | SIESP CHIETI-DRIVE IN CHIETI | Istituto Zooprofilattico Sperimentale dell'Abruzzo e Molise "G. Caporale" | Lorusso A, Marcacci M, Di Domenico M, Ancora M, Curini V, Mangone I, Rinaldi A, Scialabba S, Di Pasquale A, Cammà C, Puglia I, Calistri P, Savini G |
| EPI_ISL_883301, EPI_ISL_883302 | RP Guardiagrele-Ospedale di Comunità | Istituto Zooprofilattico Sperimentale dell'Abruzzo e Molise "G. Caporale" | Lorusso A, Marcacci M, Di Domenico M, Ancora M, Curini V, Mangone I, Rinaldi A, Scialabba S, Di Pasquale A, Cammà C, Puglia I, Calistri P, Savini G |
| EPI_ISL_883303, EPI_ISL_883304, EPI_ISL_883305 | SIESP DIPARTIMENTO DI PREVENZIONE CHIE | Istituto Zooprofilattico Sperimentale dell'Abruzzo e Molise "G. Caporale" | Lorusso A, Marcacci M, Di Domenico M, Ancora M, Curini V, Mangone I, Rinaldi A, Scialabba S, Di Pasquale A, Cammà C, Puglia I, Calistri P, Savini G |
| EPI_ISL_884865 | Medicine and Surgery, University of Insubria | University of Insubria | Novazzi,F., Genoni,A., Focosi,D., Baj,A., Spezia,P.G., Zago,C.,Colombo,A., Cassani,G., Pasciuta,R., Tamborini,A., Rossi,A.,Prestia,M., Capuano,R., Azzi,L., Donadini,A., Catanoso,G. and Maggi,F. |
| EPI_ISL_902754 | University Hospital Sant'Andrea-Sapienza | INMI Lazzaro Spallanzani IRCCS | B Bartolini, E Giombini, M Rueca, O Butera, F Messina, C.E.M Gruber, M Simmaco, I Santino, A Di Caro, MR Capobianchi |
| EPI_ISL_902755 | IRCCS San Raffaele | INMI Lazzaro Spallanzani IRCCS | E Giombini, M Rueca, O Butera, F Messina, C.E.M Gruber, B Bartolini, D Russo, D Limongi, MR Capobianchi, A Di Caro |
| EPI_ISL_902756 | Fondazione Policlinico Universitario "A. Gemelli" IRCCS | INMI Lazzaro Spallanzani IRCCS | M Rueca, O Butera, F Messina, C.E.M Gruber, B Bartolini, E Giombini, P Cattani, M Sanguinetti, MR Capobianchi, A Di Caro |
| EPI_ISL_911525 | Microbiology and Virology Unit, Florence Careggi University Hospital | Microbiology and Virology Unit, Florence Careggi University Hospital | Vincenzo Di Pilato, Marco Coppi, Fabio Morecchiato, Noemi Aiezza, Ilaria Baccani, Alberto Antonelli, Emanuele Gori, Gian Maria Rossolini |
| EPI_ISL_913446 | Laboratorio Genzano - ASL RM 6 | INMI Lazzaro Spallanzani IRCCS | Emanuela Giombini, Martina Rueca, Barbara Bartolini, Ornella Butera, Cesare E.M. Gruber, Francesco Messina, Grazia Tramini, Emanuela Conti, Antonino Di Caro, Maria R. Capobianchi |
| EPI_ISL_913447 | Laboratorio Genzano - ASL RM 6 | INMI Lazzaro Spallanzani IRCCS | Francesco Messina, Emanuela Giombini, Ornella Butera, Cesare EM Gruber, Martina Rueca, Barbara Bartolini, Grazia Tramini, Emanuela Conti, Maria R Capobianchi, Antonino Di Caro |
| EPI_ISL_918269 | SIESP DIPARTIMENTO DI PREVENZIONE TERAMO | Istituto Zooprofilattico Sperimentale dell'Abruzzo e Molise "G. Caporale" | Lorusso A, Marcacci M, Di Domenico M, Ancora M, Curini V, Mangone I, Rinaldi A, Scialabba S, Di Pasquale A, Cammà C, Puglia I, Calistri P, Savini G |
| EPI_ISL_918410 | Ospedale Di Venere - Carbonara | Istituto Zooprofilattico Sperimentale della Puglia e della Basilicata | Parisi A., Bianco A., Capozzi L., Del Sambro L., Simone D., Manzulli V, Rondinone V., Pace L., Cipolletta D., Galante D. |
| EPI_ISL_918483 | Laboratory of Microbiology, ASST Settelaghi, Varese, Italy | Laboratory of Microbiology, ASST Settelaghi, Varese, Italy | Novazzi,F., Genoni,A., Baj,A., Focosi,D., Spezia,P.G., Zago,C., Colombo,A., Cassani,G., Pasciuta,R., Tamborini,A., Rossi,A., Prestia,M., Capuano,R., Maggi,F. |
| EPI_ISL_936486 | Medicine and Surgery, University of Insubria | Medicine and Surgery, University of Insubria | Novazzi,F., Genoni,A., Baj,A., Spezia,P.G., Focosi,D., Zago,C.,Colombo,A., Cassani,G., Pasciuta,R., Tamborini,A., Rossi,A.,Prestia,M., Capuano,R. and Maggi,F. |
| EPI_ISL_940565, EPI_ISL_940631, EPI_ISL_940632, EPI_ISL_940739, EPI_ISL_949182 | University of Bari Biomedical Sciences and Human Oncology | University of Bari Biomedical Sciences and Human Oncology | Chironna M., Sallustio A., Loconsole D., Accogli M. |
| EPI_ISL_949184 | University of Bari Biomedical Sciences and Human Oncology | University of Bari Biomedical Sciences and Human Oncology | Chironna M., Sallustio A., Loconsole D., Accogli A. |
| EPI_ISL_949185 | University of Bari Biomedical Sciences and Human Oncology | University of Bari Biomedical Sciences and Human Oncology | Chironna M., Sallustio A., Loconsole D., Accogli M. |
| EPI_ISL_949188 | University of Bari Biomedical Sciences and Human Oncology | University of Bari Biomedical Sciences and Human Oncology | Chironna A., Sallustio A., Loconsole D., Accogli M. |
| EPI_ISL_949191 | University of Bari Biomedical Sciences and Human Oncology | University of Bari Biomedical Sciences and Human Oncology | Chironna M., Sallustio A., Loconsole D., Accogli M. |
| EPI_ISL_961018 | SIESP CHIETI - DRIVE IN CHIETI | Istituto Zooprofilattico Sperimentale dell'Abruzzo e Molise "G. Caporale" | Lorusso A, Marcacci M, Di Domenico M, Ancora M, Curini V, Mangone I, Rinaldi A, Scialabba S, Di Pasquale A, Cammà C, Puglia I, Calistri P, Savini G |
| EPI_ISL_961019, EPI_ISL_961020 | SIESP CHIETI - DRIVE IN ORTONA | Istituto Zooprofilattico Sperimentale dell'Abruzzo e Molise "G. Caporale" | Lorusso A, Marcacci M, Di Domenico M, Ancora M, Curini V, Mangone I, Rinaldi A, Scialabba S, Di Pasquale A, Cammà C, Puglia I, Calistri P, Savini G |

| | | | |
|---|---|---|---|
| EPI_ISL_961021, EPI_ISL_961022, EPI_ISL_961023, EPI_ISL_961024, EPI_ISL_961025, EPI_ISL_961026 | SIESP CHIETI - DRIVE IN CHIETI | Istituto Zooprofilattico Sperimentale dell'Abruzzo e Molise "G. Caporale" | Lorusso A, Marcacci M, Di Domenico M, Ancora M, Curini V, Mangone I, Rinaldi A, Scialabba S, Di Pasquale A, Cammà C, Puglia I, Calistri P, Savini G |
| EPI_ISL_961027 | SIESP CHIETI-DRIVE IN VASTO | Istituto Zooprofilattico Sperimentale dell'Abruzzo e Molise "G. Caporale" | Lorusso A, Marcacci M, Di Domenico M, Ancora M, Curini V, Mangone I, Rinaldi A, Scialabba S, Di Pasquale A, Cammà C, Puglia I, Calistri P, Savini G |
| EPI_ISL_961028, EPI_ISL_961029, EPI_ISL_961030, EPI_ISL_961031 | SIESP CHIETI - DRIVE IN CHIETI | Istituto Zooprofilattico Sperimentale dell'Abruzzo e Molise "G. Caporale" | Lorusso A, Marcacci M, Di Domenico M, Ancora M, Curini V, Mangone I, Rinaldi A, Scialabba S, Di Pasquale A, Cammà C, Puglia I, Calistri P, Savini G |
| EPI_ISL_961032 | SIESP DIPARTIMENTO DI PREVENZIONE CHIETI | Istituto Zooprofilattico Sperimentale dell'Abruzzo e Molise "G. Caporale" | Lorusso A, Marcacci M, Di Domenico M, Ancora M, Curini V, Mangone I, Rinaldi A, Scialabba S, Di Pasquale A, Cammà C, Puglia I, Calistri P, Savini G |
| EPI_ISL_961033 | SIESP CHIETI - DRIVE IN ORTONA | Istituto Zooprofilattico Sperimentale dell'Abruzzo e Molise "G. Caporale" | Lorusso A, Marcacci M, Di Domenico M, Ancora M, Curini V, Mangone I, Rinaldi A, Scialabba S, Di Pasquale A, Cammà C, Puglia I, Calistri P, Savini G |
| EPI_ISL_961034, EPI_ISL_961035 | SIESP CHIETI - DRIVE IN CHIETI | Istituto Zooprofilattico Sperimentale dell'Abruzzo e Molise "G. Caporale" | Lorusso A, Marcacci M, Di Domenico M, Ancora M, Curini V, Mangone I, Rinaldi A, Scialabba S, Di Pasquale A, Cammà C, Puglia I, Calistri P, Savini G |
| EPI_ISL_961036 | SIESP CHIETI - DRIVE IN ORTONA | Istituto Zooprofilattico Sperimentale dell'Abruzzo e Molise "G. Caporale" | Lorusso A, Marcacci M, Di Domenico M, Ancora M, Curini V, Mangone I, Rinaldi A, Scialabba S, Di Pasquale A, Cammà C, Puglia I, Calistri P, Savini G |
| EPI_ISL_961037, EPI_ISL_961038, EPI_ISL_961039, EPI_ISL_961040, EPI_ISL_961041 | SIESP CHIETI - DRIVE IN CHIETI | Istituto Zooprofilattico Sperimentale dell'Abruzzo e Molise "G. Caporale" | Lorusso A, Marcacci M, Di Domenico M, Ancora M, Curini V, Mangone I, Rinaldi A, Scialabba S, Di Pasquale A, Cammà C, Puglia I, Calistri P, Savini G |
| EPI_ISL_961042 | SIESP TERAMO | Istituto Zooprofilattico Sperimentale dell'Abruzzo e Molise "G. Caporale" | Lorusso A, Marcacci M, Di Domenico M, Ancora M, Curini V, Mangone I, Rinaldi A, Scialabba S, Di Pasquale A, Cammà C, Puglia I, Calistri P, Savini G |
| EPI_ISL_961043 | SIESP CHIETI - DRIVE IN CHIETI | Istituto Zooprofilattico Sperimentale dell'Abruzzo e Molise "G. Caporale" | Lorusso A, Marcacci M, Di Domenico M, Ancora M, Curini V, Mangone I, Rinaldi A, Scialabba S, Di Pasquale A, Cammà C, Puglia I, Calistri P, Savini G |
| EPI_ISL_961044, EPI_ISL_961045, EPI_ISL_961046 | SIESP CHIETI - DRIVE IN ORTONA | Istituto Zooprofilattico Sperimentale dell'Abruzzo e Molise "G. Caporale" | Lorusso A, Marcacci M, Di Domenico M, Ancora M, Curini V, Mangone I, Rinaldi A, Scialabba S, Di Pasquale A, Cammà C, Puglia I, Calistri P, Savini G |
| EPI_ISL_961047 | SIESP CHIETI - DRIVE IN CHIETI | Istituto Zooprofilattico Sperimentale dell'Abruzzo e Molise "G. Caporale" | Lorusso A, Marcacci M, Di Domenico M, Ancora M, Curini V, Mangone I, Rinaldi A, Scialabba S, Di Pasquale A, Cammà C, Puglia I, Calistri P, Savini G |
| EPI_ISL_961048, EPI_ISL_961049 | SIESP CHIETI - DRIVE IN ORTONA | Istituto Zooprofilattico Sperimentale dell'Abruzzo e Molise "G. Caporale" | Lorusso A, Marcacci M, Di Domenico M, Ancora M, Curini V, Mangone I, Rinaldi A, Scialabba S, Di Pasquale A, Cammà C, Puglia I, Calistri P, Savini G |
| EPI_ISL_961050, EPI_ISL_961051, EPI_ISL_961052, EPI_ISL_961053, EPI_ISL_961054 | SIESP DIPARTIMENTO DI PREVENZIONE CHIETI | Istituto Zooprofilattico Sperimentale dell'Abruzzo e Molise "G. Caporale" | Lorusso A, Marcacci M, Di Domenico M, Ancora M, Curini V, Mangone I, Rinaldi A, Scialabba S, Di Pasquale A, Cammà C, Puglia I, Calistri P, Savini G |
| EPI_ISL_961055, EPI_ISL_961056 | SIESP CHIETI - DRIVE IN ORTONA | Istituto Zooprofilattico Sperimentale dell'Abruzzo e Molise "G. Caporale" | Lorusso A, Marcacci M, Di Domenico M, Ancora M, Curini V, Mangone I, Rinaldi A, Scialabba S, Di Pasquale A, Cammà C, Puglia I, Calistri P, Savini G |
| EPI_ISL_961057, EPI_ISL_961058, EPI_ISL_961059, EPI_ISL_961060 | SIESP CHIETI - DRIVE IN CHIETI | Istituto Zooprofilattico Sperimentale dell'Abruzzo e Molise "G. Caporale" | Lorusso A, Marcacci M, Di Domenico M, Ancora M, Curini V, Mangone I, Rinaldi A, Scialabba S, Di Pasquale A, Cammà C, Puglia I, Calistri P, Savini G |
| EPI_ISL_961061 | SIESP CHIETI - DRIVE IN ORTONA | Istituto Zooprofilattico Sperimentale dell'Abruzzo e Molise "G. Caporale" | Lorusso A, Marcacci M, Di Domenico M, Ancora M, Curini V, Mangone I, Rinaldi A, Scialabba S, Di Pasquale A, Cammà C, Puglia I, Calistri P, Savini G |
| EPI_ISL_961062 | SIESP CHIETI - DRIVE IN CHIETI | Istituto Zooprofilattico Sperimentale dell'Abruzzo e Molise "G. Caporale" | Lorusso A, Marcacci M, Di Domenico M, Ancora M, Curini V, Mangone I, Rinaldi A, Scialabba S, Di Pasquale A, Cammà C, Puglia I, Calistri P, Savini G |
| EPI_ISL_961063 | SIESP DIPARTIMENTO DI PREVENZIONE CHIETI | Istituto Zooprofilattico Sperimentale dell'Abruzzo e Molise "G. Caporale" | Lorusso A, Marcacci M, Di Domenico M, Ancora M, Curini V, Mangone I, Rinaldi A, Scialabba S, Di Pasquale A, Cammà C, Puglia I, Calistri P, Savini G |
| EPI_ISL_961064, EPI_ISL_961065 | SIESP CHIETI - DRIVE IN ORTONA | Istituto Zooprofilattico Sperimentale dell'Abruzzo e Molise "G. Caporale" | Lorusso A, Marcacci M, Di Domenico M, Ancora M, Curini V, Mangone I, Rinaldi A, Scialabba S, Di Pasquale A, Cammà C, Puglia I, Calistri P, Savini G |
| EPI_ISL_961066 | SIESP CHIETI - DRIVE IN LANCIANO | Istituto Zooprofilattico Sperimentale dell'Abruzzo e Molise "G. Caporale" | Lorusso A, Marcacci M, Di Domenico M, Ancora M, Curini V, Mangone I, Rinaldi A, Scialabba S, Di Pasquale A, Cammà C, Puglia I, Calistri P, Savini G |
| EPI_ISL_961067, EPI_ISL_961068 | SIESP DIPARTIMENTO DI PREVENZIONE CHIETI | Istituto Zooprofilattico Sperimentale dell'Abruzzo e Molise "G. Caporale" | Lorusso A, Marcacci M, Di Domenico M, Ancora M, Curini V, Mangone I, Rinaldi A, Scialabba S, Di Pasquale A, Cammà C, Puglia I, Calistri P, Savini G |
| EPI_ISL_961069 | SIESP CHIETI - DRIVE IN CHIETI | Istituto Zooprofilattico Sperimentale dell'Abruzzo e Molise "G. Caporale" | Lorusso A, Marcacci M, Di Domenico M, Ancora M, Curini V, Mangone I, Rinaldi A, Scialabba S, Di Pasquale A, Cammà C, Puglia I, Calistri P, Savini G |
| EPI_ISL_961070 | SIESP DIPARTIMENTO DI PREVENZIONE CHIETI | Istituto Zooprofilattico Sperimentale dell'Abruzzo e Molise "G. Caporale" | Lorusso A, Marcacci M, Di Domenico M, Ancora M, Curini V, Mangone I, Rinaldi A, Scialabba S, Di Pasquale A, Cammà C, Puglia I, Calistri P, Savini G |
| EPI_ISL_961071, EPI_ISL_961072, EPI_ISL_961073 | SIESP CHIETI - DRIVE IN CHIETI | Istituto Zooprofilattico Sperimentale dell'Abruzzo e Molise "G. Caporale" | Lorusso A, Marcacci M, Di Domenico M, Ancora M, Curini V, Mangone I, Rinaldi A, Scialabba S, Di Pasquale A, Cammà C, Puglia I, Calistri P, Savini G |
| EPI_ISL_961074 | SIESP CHIETI - DRIVE IN ORTONA | Istituto Zooprofilattico Sperimentale dell'Abruzzo e Molise "G. Caporale" | Lorusso A, Marcacci M, Di Domenico M, Ancora M, Curini V, Mangone I, Rinaldi A, Scialabba S, Di Pasquale A, Cammà C, Puglia I, Calistri P, Savini G |
| EPI_ISL_961075 | SIESP DIPARTIMENTO DI PREVENZIONE CHIETI | Istituto Zooprofilattico Sperimentale dell'Abruzzo e Molise "G. Caporale" | Lorusso A, Marcacci M, Di Domenico M, Ancora M, Curini V, Mangone I, Rinaldi A, Scialabba S, Di Pasquale A, Cammà C, Puglia I, Calistri P, Savini G |
| EPI_ISL_961076 | SIESP CHIETI - DRIVE IN ORTONA | Istituto Zooprofilattico Sperimentale dell'Abruzzo e Molise "G. Caporale" | Lorusso A, Marcacci M, Di Domenico M, Ancora M, Curini V, Mangone I, Rinaldi A, Scialabba S, Di Pasquale A, Cammà C, Puglia I, Calistri P, Savini G |
| EPI_ISL_961077 | SIESP TERAMO | Istituto Zooprofilattico Sperimentale dell'Abruzzo e Molise "G. Caporale" | Lorusso A, Marcacci M, Di Domenico M, Ancora M, Curini V, Mangone I, Rinaldi A, Scialabba S, Di Pasquale A, Cammà C, Puglia I, Calistri P, Savini G |
| EPI_ISL_961078, EPI_ISL_961079, EPI_ISL_961080 | DIP. PREV. AVEZZANO SERVIZIO DI IGIENE EPIDEMIOLOGIAE SANITA' PUBBLICA | Istituto Zooprofilattico Sperimentale dell'Abruzzo e Molise "G. Caporale" | Lorusso A, Marcacci M, Di Domenico M, Ancora M, Curini V, Mangone I, Rinaldi A, Scialabba S, Di Pasquale A, Cammà C, Puglia I, Calistri P, Savini G |
| EPI_ISL_961081 | SIESP CHIETI - DRIVE IN CHIETI | Istituto Zooprofilattico Sperimentale dell'Abruzzo e Molise "G. Caporale" | Lorusso A, Marcacci M, Di Domenico M, Ancora M, Curini V, Mangone I, Rinaldi A, Scialabba S, Di Pasquale A, Cammà C, Puglia I, Calistri P, Savini G |
| EPI_ISL_961082 | SIESP DIPARTIMENTO DI PREVENZIONE CHIETI | Istituto Zooprofilattico Sperimentale dell'Abruzzo e Molise "G. Caporale" | Lorusso A, Marcacci M, Di Domenico M, Ancora M, Curini V, Mangone I, Rinaldi A, Scialabba S, Di Pasquale A, Cammà C, Puglia I, Calistri P, Savini G |
| EPI_ISL_961083 | DIP. PREV. AVEZZANO SERVIZIO DI IGIENE EPIDEMIOLOGIAE SANITA' PUBBLICA | Istituto Zooprofilattico Sperimentale dell'Abruzzo e Molise "G. Caporale" | Lorusso A, Marcacci M, Di Domenico M, Ancora M, Curini V, Mangone I, Rinaldi A, Scialabba S, Di Pasquale A, Cammà C, Puglia I, Calistri P, Savini G |
| EPI_ISL_961084 | SIESP TERAMO | Istituto Zooprofilattico Sperimentale dell'Abruzzo e Molise "G. Caporale" | Lorusso A, Marcacci M, Di Domenico M, Ancora M, Curini V, Mangone I, Rinaldi A, Scialabba S, Di Pasquale A, Cammà C, Puglia I, Calistri P, Savini G |

| | | | |
|---|---|---|---|
| EPI_ISL_961085 | SIESP CHIETI - DRIVE IN ORTONA | Istituto Zooprofilattico Sperimentale dell'Abruzzo e Molise "G. Caporale" | Lorusso A, Marcacci M, Di Domenico M, Ancora M, Curini V, Mangone I, Rinaldi A, Scialabba S, Di Pasquale A, Cammà C, Puglia I, Calistri P, Savini G |
| EPI_ISL_961086 | SIESP CHIETI - DRIVE IN CHIETI | Istituto Zooprofilattico Sperimentale dell'Abruzzo e Molise "G. Caporale" | Lorusso A, Marcacci M, Di Domenico M, Ancora M, Curini V, Mangone I, Rinaldi A, Scialabba S, Di Pasquale A, Cammà C, Puglia I, Calistri P, Savini G |
| EPI_ISL_961087, EPI_ISL_961088, EPI_ISL_961089 | SIESP DIPARTIMENTO DI PREVENZIONE CHIETI | Istituto Zooprofilattico Sperimentale dell'Abruzzo e Molise "G. Caporale" | Lorusso A, Marcacci M, Di Domenico M, Ancora M, Curini V, Mangone I, Rinaldi A, Scialabba S, Di Pasquale A, Cammà C, Puglia I, Calistri P, Savini G |
| EPI_ISL_961090, EPI_ISL_961091, EPI_ISL_961092 | SIESP CHIETI - DRIVE IN CHIETI | Istituto Zooprofilattico Sperimentale dell'Abruzzo e Molise "G. Caporale" | Lorusso A, Marcacci M, Di Domenico M, Ancora M, Curini V, Mangone I, Rinaldi A, Scialabba S, Di Pasquale A, Cammà C, Puglia I, Calistri P, Savini G |
| EPI_ISL_961093 | Ospedale SS Annunziata | Istituto Zooprofilattico Sperimentale dell'Abruzzo e Molise "G. Caporale" | Lorusso A, Marcacci M, Di Domenico M, Ancora M, Curini V, Mangone I, Rinaldi A, Scialabba S, Di Pasquale A, Cammà C, Puglia I, Calistri P, Savini G |
| EPI_ISL_961094, EPI_ISL_961095 | DIP. PREV. AVEZZANO SERVIZIO DI IGIENE EPIDEMIOLOGIAE SANITA' PUBBLICA | Istituto Zooprofilattico Sperimentale dell'Abruzzo e Molise "G. Caporale" | Lorusso A, Marcacci M, Di Domenico M, Ancora M, Curini V, Mangone I, Rinaldi A, Scialabba S, Di Pasquale A, Cammà C, Puglia I, Calistri P, Savini G |
| EPI_ISL_961096 | SIESP L'AQUILA | Istituto Zooprofilattico Sperimentale dell'Abruzzo e Molise "G. Caporale" | Lorusso A, Marcacci M, Di Domenico M, Ancora M, Curini V, Mangone I, Rinaldi A, Scialabba S, Di Pasquale A, Cammà C, Puglia I, Calistri P, Savini G |
| EPI_ISL_961097, EPI_ISL_961098, EPI_ISL_961099, EPI_ISL_961100, EPI_ISL_961101, EPI_ISL_961102, EPI_ISL_961103 | SIESP TERAMO | Istituto Zooprofilattico Sperimentale dell'Abruzzo e Molise "G. Caporale" | Lorusso A, Marcacci M, Di Domenico M, Ancora M, Curini V, Mangone I, Rinaldi A, Scialabba S, Di Pasquale A, Cammà C, Puglia I, Calistri P, Savini G |
| EPI_ISL_961673 | SIESP CHIETI DRIVE IN LANCIANO | Istituto Zooprofilattico Sperimentale dell'Abruzzo e Molise "G. Caporale" | Lorusso A, Marcacci M, Di Domenico M, Ancora M, Curini V, Mangone I, Rinaldi A, Scialabba S, Di Pasquale A, Cammà C, Puglia I, Calistri P, Savini G |
| EPI_ISL_961674, EPI_ISL_961675, EPI_ISL_961676, EPI_ISL_961677 | SIESP CHIETI - DRIVE IN CHIETI | Istituto Zooprofilattico Sperimentale dell'Abruzzo e Molise "G. Caporale" | Lorusso A, Marcacci M, Di Domenico M, Ancora M, Curini V, Mangone I, Rinaldi A, Scialabba S, Di Pasquale A, Cammà C, Puglia I, Calistri P, Savini G |
| EPI_ISL_961678 | SIESP TERAMO | Istituto Zooprofilattico Sperimentale dell'Abruzzo e Molise "G. Caporale" | Lorusso A, Marcacci M, Di Domenico M, Ancora M, Curini V, Mangone I, Rinaldi A, Scialabba S, Di Pasquale A, Cammà C, Puglia I, Calistri P, Savini G |
| EPI_ISL_961679, EPI_ISL_961680, EPI_ISL_961681, EPI_ISL_961682 | SIESP CHIETI - DRIVE IN CHIETI | Istituto Zooprofilattico Sperimentale dell'Abruzzo e Molise "G. Caporale" | Lorusso A, Marcacci M, Di Domenico M, Ancora M, Curini V, Mangone I, Rinaldi A, Scialabba S, Di Pasquale A, Cammà C, Puglia I, Calistri P, Savini G |
| EPI_ISL_961683 | SIESP TERAMO | Istituto Zooprofilattico Sperimentale dell'Abruzzo e Molise "G. Caporale" | Lorusso A, Marcacci M, Di Domenico M, Ancora M, Curini V, Mangone I, Rinaldi A, Scialabba S, Di Pasquale A, Cammà C, Puglia I, Calistri P, Savini G |
| EPI_ISL_961684 | SIESP CHIETI - DRIVE IN CHIETI | Istituto Zooprofilattico Sperimentale dell'Abruzzo e Molise "G. Caporale" | Lorusso A, Marcacci M, Di Domenico M, Ancora M, Curini V, Mangone I, Rinaldi A, Scialabba S, Di Pasquale A, Cammà C, Puglia I, Calistri P, Savini G |
| EPI_ISL_961685 | SIESP DIPARTIMENTO DI PREVENZIONE CHIETI | Istituto Zooprofilattico Sperimentale dell'Abruzzo e Molise "G. Caporale" | Lorusso A, Marcacci M, Di Domenico M, Ancora M, Curini V, Mangone I, Rinaldi A, Scialabba S, Di Pasquale A, Cammà C, Puglia I, Calistri P, Savini G |
| EPI_ISL_961686, EPI_ISL_961687 | SIESP CHIETI - DRIVE IN ORTONA | Istituto Zooprofilattico Sperimentale dell'Abruzzo e Molise "G. Caporale" | Lorusso A, Marcacci M, Di Domenico M, Ancora M, Curini V, Mangone I, Rinaldi A, Scialabba S, Di Pasquale A, Cammà C, Puglia I, Calistri P, Savini G |
| EPI_ISL_961688 | SIESP TERAMO | Istituto Zooprofilattico Sperimentale dell'Abruzzo e Molise "G. Caporale" | Lorusso A, Marcacci M, Di Domenico M, Ancora M, Curini V, Mangone I, Rinaldi A, Scialabba S, Di Pasquale A, Cammà C, Puglia I, Calistri P, Savini G |
| EPI_ISL_961689, EPI_ISL_961690 | SIESP CHIETI - DRIVE IN CHIETI | Istituto Zooprofilattico Sperimentale dell'Abruzzo e Molise "G. Caporale" | Lorusso A, Marcacci M, Di Domenico M, Ancora M, Curini V, Mangone I, Rinaldi A, Scialabba S, Di Pasquale A, Cammà C, Puglia I, Calistri P, Savini G |
| EPI_ISL_961691, EPI_ISL_961692, EPI_ISL_961693 | SIESP CHIETI - DRIVE IN ORTONA | Istituto Zooprofilattico Sperimentale dell'Abruzzo e Molise "G. Caporale" | Lorusso A, Marcacci M, Di Domenico M, Ancora M, Curini V, Mangone I, Rinaldi A, Scialabba S, Di Pasquale A, Cammà C, Puglia I, Calistri P, Savini G |
| EPI_ISL_961694 | SIESP CHIETI - DRIVE IN CHIETI | Istituto Zooprofilattico Sperimentale dell'Abruzzo e Molise "G. Caporale" | Lorusso A, Marcacci M, Di Domenico M, Ancora M, Curini V, Mangone I, Rinaldi A, Scialabba S, Di Pasquale A, Cammà C, Puglia I, Calistri P, Savini G |
| EPI_ISL_961695 | SIESP DIPARTIMENTO DI PREVENZIONE CHIETI | Istituto Zooprofilattico Sperimentale dell'Abruzzo e Molise "G. Caporale" | Lorusso A, Marcacci M, Di Domenico M, Ancora M, Curini V, Mangone I, Rinaldi A, Scialabba S, Di Pasquale A, Cammà C, Puglia I, Calistri P, Savini G |
| EPI_ISL_961696 | SIESP SULMONA | Istituto Zooprofilattico Sperimentale dell'Abruzzo e Molise "G. Caporale" | Lorusso A, Marcacci M, Di Domenico M, Ancora M, Curini V, Mangone I, Rinaldi A, Scialabba S, Di Pasquale A, Cammà C, Puglia I, Calistri P, Savini G |
| EPI_ISL_961697, EPI_ISL_961698 | SIESP CHIETI - DRIVE IN CHIETI | Istituto Zooprofilattico Sperimentale dell'Abruzzo e Molise "G. Caporale" | Lorusso A, Marcacci M, Di Domenico M, Ancora M, Curini V, Mangone I, Rinaldi A, Scialabba S, Di Pasquale A, Cammà C, Puglia I, Calistri P, Savini G |
| EPI_ISL_961699 | SIESP CHIETI - DRIVE IN ORTONA | Istituto Zooprofilattico Sperimentale dell'Abruzzo e Molise "G. Caporale" | Lorusso A, Marcacci M, Di Domenico M, Ancora M, Curini V, Mangone I, Rinaldi A, Scialabba S, Di Pasquale A, Cammà C, Puglia I, Calistri P, Savini G |
| EPI_ISL_961700 | SIESP CHIETI - DRIVE IN CHIETI | Istituto Zooprofilattico Sperimentale dell'Abruzzo e Molise "G. Caporale" | Lorusso A, Marcacci M, Di Domenico M, Ancora M, Curini V, Mangone I, Rinaldi A, Scialabba S, Di Pasquale A, Cammà C, Puglia I, Calistri P, Savini G |
| EPI_ISL_961701 | Ospedale Civile Teramo | Istituto Zooprofilattico Sperimentale dell'Abruzzo e Molise "G. Caporale" | Lorusso A, Marcacci M, Di Domenico M, Ancora M, Curini V, Mangone I, Rinaldi A, Scialabba S, Di Pasquale A, Cammà C, Puglia I, Calistri P, Savini G |
| EPI_ISL_961702 | SIESP CHIETI - DRIVE IN CHIETI | Istituto Zooprofilattico Sperimentale dell'Abruzzo e Molise "G. Caporale" | Lorusso A, Marcacci M, Di Domenico M, Ancora M, Curini V, Mangone I, Rinaldi A, Scialabba S, Di Pasquale A, Cammà C, Puglia I, Calistri P, Savini G |
| EPI_ISL_961703 | SIESP DIPARTIMENTO DI PREVENZIONE CHIETI | Istituto Zooprofilattico Sperimentale dell'Abruzzo e Molise "G. Caporale" | Lorusso A, Marcacci M, Di Domenico M, Ancora M, Curini V, Mangone I, Rinaldi A, Scialabba S, Di Pasquale A, Cammà C, Puglia I, Calistri P, Savini G |
| EPI_ISL_961704 | SIESP SULMONA | Istituto Zooprofilattico Sperimentale dell'Abruzzo e Molise "G. Caporale" | Lorusso A, Marcacci M, Di Domenico M, Ancora M, Curini V, Mangone I, Rinaldi A, Scialabba S, Di Pasquale A, Cammà C, Puglia I, Calistri P, Savini G |
| EPI_ISL_961705 | Ospedale San Salvatore UOC Malattie Infettive | Istituto Zooprofilattico Sperimentale dell'Abruzzo e Molise "G. Caporale" | Lorusso A, Marcacci M, Di Domenico M, Ancora M, Curini V, Mangone I, Rinaldi A, Scialabba S, Di Pasquale A, Cammà C, Puglia I, Calistri P, Savini G |
| EPI_ISL_961706 | SIESP CHIETI - DRIVE IN CHIETI | Istituto Zooprofilattico Sperimentale dell'Abruzzo e Molise "G. Caporale" | Lorusso A, Marcacci M, Di Domenico M, Ancora M, Curini V, Mangone I, Rinaldi A, Scialabba S, Di Pasquale A, Cammà C, Puglia I, Calistri P, Savini G |
| EPI_ISL_961707, EPI_ISL_961708 | SIESP DIPARTIMENTO DI PREVENZIONE CHIETI | Istituto Zooprofilattico Sperimentale dell'Abruzzo e Molise "G. Caporale" | Lorusso A, Marcacci M, Di Domenico M, Ancora M, Curini V, Mangone I, Rinaldi A, Scialabba S, Di Pasquale A, Cammà C, Puglia I, Calistri P, Savini G |
| EPI_ISL_961709, EPI_ISL_961710, EPI_ISL_961711 | SIESP CHIETI - DRIVE IN CHIETI | Istituto Zooprofilattico Sperimentale dell'Abruzzo e Molise "G. Caporale" | Lorusso A, Marcacci M, Di Domenico M, Ancora M, Curini V, Mangone I, Rinaldi A, Scialabba S, Di Pasquale A, Cammà C, Puglia I, Calistri P, Savini G |
| EPI_ISL_961712, EPI_ISL_961713 | SIESP CHIETI DRIVE IN LANCIANO | Istituto Zooprofilattico Sperimentale dell'Abruzzo e Molise "G. Caporale" | Lorusso A, Marcacci M, Di Domenico M, Ancora M, Curini V, Mangone I, Rinaldi A, Scialabba S, Di Pasquale A, Cammà C, Puglia I, Calistri P, Savini G |
| EPI_ISL_961714 | SIESP CHIETI - DRIVE IN CHIETI | Istituto Zooprofilattico Sperimentale dell'Abruzzo e Molise "G. | Lorusso A, Marcacci M, Di Domenico M, Ancora M, Curini V, Mangone I, Rinaldi A, Scialabba S, Di Pasquale A, Cammà C, Puglia I, Calistri P, Savini G |

| | | | |
|---|---|---|---|
| | | Caporale" | |
| EPI_ISL_961715, EPI_ISL_961716 | SIESP CHIETI - DRIVE IN ORTONA | Istituto Zooprofilattico Sperimentale dell'Abruzzo e Molise "G. Caporale" | Lorusso A, Marcacci M, Di Domenico M, Ancora M, Curini V, Mangone I, Rinaldi A, Scialabba S, Di Pasquale A, Cammà C, Puglia I, Calistri P, Savini G |
| EPI_ISL_961717 | SIESP CHIETI - DRIVE IN CHIETI | Istituto Zooprofilattico Sperimentale dell'Abruzzo e Molise "G. Caporale" | Lorusso A, Marcacci M, Di Domenico M, Ancora M, Curini V, Mangone I, Rinaldi A, Scialabba S, Di Pasquale A, Cammà C, Puglia I, Calistri P, Savini G |
| EPI_ISL_961718, EPI_ISL_961719, EPI_ISL_961720, EPI_ISL_961721, EPI_ISL_961722 | SIESP CHIETI - DRIVE IN ORTONA | Istituto Zooprofilattico Sperimentale dell'Abruzzo e Molise "G. Caporale" | Lorusso A, Marcacci M, Di Domenico M, Ancora M, Curini V, Mangone I, Rinaldi A, Scialabba S, Di Pasquale A, Cammà C, Puglia I, Calistri P, Savini G |
| EPI_ISL_961723 | SIESP CHIETI DRIVE IN LANCIANO | Istituto Zooprofilattico Sperimentale dell'Abruzzo e Molise "G. Caporale" | Lorusso A, Marcacci M, Di Domenico M, Ancora M, Curini V, Mangone I, Rinaldi A, Scialabba S, Di Pasquale A, Cammà C, Puglia I, Calistri P, Savini G |
| EPI_ISL_961724 | SIESP CHIETI - DRIVE IN ORTONA | Istituto Zooprofilattico Sperimentale dell'Abruzzo e Molise "G. Caporale" | Lorusso A, Marcacci M, Di Domenico M, Ancora M, Curini V, Mangone I, Rinaldi A, Scialabba S, Di Pasquale A, Cammà C, Puglia I, Calistri P, Savini G |
| EPI_ISL_961725 | SIESP CHIETI DRIVE IN GISSI | Istituto Zooprofilattico Sperimentale dell'Abruzzo e Molise "G. Caporale" | Lorusso A, Marcacci M, Di Domenico M, Ancora M, Curini V, Mangone I, Rinaldi A, Scialabba S, Di Pasquale A, Cammà C, Puglia I, Calistri P, Savini G |
| EPI_ISL_961726, EPI_ISL_961727 | SIESP CHIETI - DRIVE IN CHIETI | Istituto Zooprofilattico Sperimentale dell'Abruzzo e Molise "G. Caporale" | Lorusso A, Marcacci M, Di Domenico M, Ancora M, Curini V, Mangone I, Rinaldi A, Scialabba S, Di Pasquale A, Cammà C, Puglia I, Calistri P, Savini G |
| EPI_ISL_961728 | SIESP DIPARTIMENTO DI PREVENZIONE CHIETI | Istituto Zooprofilattico Sperimentale dell'Abruzzo e Molise "G. Caporale" | Lorusso A, Marcacci M, Di Domenico M, Ancora M, Curini V, Mangone I, Rinaldi A, Scialabba S, Di Pasquale A, Cammà C, Puglia I, Calistri P, Savini G |
| EPI_ISL_961729 | SIESP CHIETI - DRIVE IN CHIETI | Istituto Zooprofilattico Sperimentale dell'Abruzzo e Molise "G. Caporale" | Lorusso A, Marcacci M, Di Domenico M, Ancora M, Curini V, Mangone I, Rinaldi A, Scialabba S, Di Pasquale A, Cammà C, Puglia I, Calistri P, Savini G |
| EPI_ISL_961730 | Dipartimento Prevenzione Avezzano-Servizio Igiene epidemiologia Sanità Pubblica | Istituto Zooprofilattico Sperimentale dell'Abruzzo e Molise "G. Caporale" | Lorusso A, Marcacci M, Di Domenico M, Ancora M, Curini V, Mangone I, Rinaldi A, Scialabba S, Di Pasquale A, Cammà C, Puglia I, Calistri P, Savini G |
| EPI_ISL_961731, EPI_ISL_961732 | SIESP CHIETI - DRIVE IN CHIETI | Istituto Zooprofilattico Sperimentale dell'Abruzzo e Molise "G. Caporale" | Lorusso A, Marcacci M, Di Domenico M, Ancora M, Curini V, Mangone I, Rinaldi A, Scialabba S, Di Pasquale A, Cammà C, Puglia I, Calistri P, Savini G |
| EPI_ISL_961733 | SIESP CHIETI - DRIVE IN ORTONA | Istituto Zooprofilattico Sperimentale dell'Abruzzo e Molise "G. Caporale" | Lorusso A, Marcacci M, Di Domenico M, Ancora M, Curini V, Mangone I, Rinaldi A, Scialabba S, Di Pasquale A, Cammà C, Puglia I, Calistri P, Savini G |
| EPI_ISL_961734 | Dipartimento Prevenzione Avezzano-Servizio Igiene epidemiologia Sanità Pubblica | Istituto Zooprofilattico Sperimentale dell'Abruzzo e Molise "G. Caporale" | Lorusso A, Marcacci M, Di Domenico M, Ancora M, Curini V, Mangone I, Rinaldi A, Scialabba S, Di Pasquale A, Cammà C, Puglia I, Calistri P, Savini G |
| EPI_ISL_961735, EPI_ISL_961736 | SIESP CHIETI - DRIVE IN CHIETI | Istituto Zooprofilattico Sperimentale dell'Abruzzo e Molise "G. Caporale" | Lorusso A, Marcacci M, Di Domenico M, Ancora M, Curini V, Mangone I, Rinaldi A, Scialabba S, Di Pasquale A, Cammà C, Puglia I, Calistri P, Savini G |
| EPI_ISL_961737 | SIESP CHIETI- DRIVE IN VASTO | Istituto Zooprofilattico Sperimentale dell'Abruzzo e Molise "G. Caporale" | Lorusso A, Marcacci M, Di Domenico M, Ancora M, Curini V, Mangone I, Rinaldi A, Scialabba S, Di Pasquale A, Cammà C, Puglia I, Calistri P, Savini G |
| EPI_ISL_961738, EPI_ISL_961739 | SIESP CHIETI DRIVE IN LANCIANO | Istituto Zooprofilattico Sperimentale dell'Abruzzo e Molise "G. Caporale" | Lorusso A, Marcacci M, Di Domenico M, Ancora M, Curini V, Mangone I, Rinaldi A, Scialabba S, Di Pasquale A, Cammà C, Puglia I, Calistri P, Savini G |
| EPI_ISL_961740 | SIESP CHIETI - DRIVE IN CHIETI | Istituto Zooprofilattico Sperimentale dell'Abruzzo e Molise "G. Caporale" | Lorusso A, Marcacci M, Di Domenico M, Ancora M, Curini V, Mangone I, Rinaldi A, Scialabba S, Di Pasquale A, Cammà C, Puglia I, Calistri P, Savini G |
| EPI_ISL_961741 | SIESP CHIETI- DRIVE IN VASTO | Istituto Zooprofilattico Sperimentale dell'Abruzzo e Molise "G. Caporale" | Lorusso A, Marcacci M, Di Domenico M, Ancora M, Curini V, Mangone I, Rinaldi A, Scialabba S, Di Pasquale A, Cammà C, Puglia I, Calistri P, Savini G |
| EPI_ISL_961742 | SIESP CHIETI - DRIVE IN CHIETI | Istituto Zooprofilattico Sperimentale dell'Abruzzo e Molise "G. Caporale" | Lorusso A, Marcacci M, Di Domenico M, Ancora M, Curini V, Mangone I, Rinaldi A, Scialabba S, Di Pasquale A, Cammà C, Puglia I, Calistri P, Savini G |
| EPI_ISL_961743 | SIESP CHIETI DRIVE IN GISSI | Istituto Zooprofilattico Sperimentale dell'Abruzzo e Molise "G. Caporale" | Lorusso A, Marcacci M, Di Domenico M, Ancora M, Curini V, Mangone I, Rinaldi A, Scialabba S, Di Pasquale A, Cammà C, Puglia I, Calistri P, Savini G |
| EPI_ISL_961744 | SIESP CHIETI - DRIVE IN ORTONA | Istituto Zooprofilattico Sperimentale dell'Abruzzo e Molise "G. Caporale" | Lorusso A, Marcacci M, Di Domenico M, Ancora M, Curini V, Mangone I, Rinaldi A, Scialabba S, Di Pasquale A, Cammà C, Puglia I, Calistri P, Savini G |
| EPI_ISL_961745 | SIESP CHIETI DRIVE IN GISSI | Istituto Zooprofilattico Sperimentale dell'Abruzzo e Molise "G. Caporale" | Lorusso A, Marcacci M, Di Domenico M, Ancora M, Curini V, Mangone I, Rinaldi A, Scialabba S, Di Pasquale A, Cammà C, Puglia I, Calistri P, Savini G |
| EPI_ISL_961746 | SIESP CHIETI - DRIVE IN CHIETI | Istituto Zooprofilattico Sperimentale dell'Abruzzo e Molise "G. Caporale" | Lorusso A, Marcacci M, Di Domenico M, Ancora M, Curini V, Mangone I, Rinaldi A, Scialabba S, Di Pasquale A, Cammà C, Puglia I, Calistri P, Savini G |
| EPI_ISL_961747 | Dipartimento Prevenzione Avezzano-Servizio Igiene epidemiologia Sanità Pubblica | Istituto Zooprofilattico Sperimentale dell'Abruzzo e Molise "G. Caporale" | Lorusso A, Marcacci M, Di Domenico M, Ancora M, Curini V, Mangone I, Rinaldi A, Scialabba S, Di Pasquale A, Cammà C, Puglia I, Calistri P, Savini G |
| EPI_ISL_961748 | SIESP CHIETI DRIVE IN LANCIANO | Istituto Zooprofilattico Sperimentale dell'Abruzzo e Molise "G. Caporale" | Lorusso A, Marcacci M, Di Domenico M, Ancora M, Curini V, Mangone I, Rinaldi A, Scialabba S, Di Pasquale A, Cammà C, Puglia I, Calistri P, Savini G |
| EPI_ISL_961749 | SIESP DIPARTIMENTO DI PREVENZIONE CHIETI | Istituto Zooprofilattico Sperimentale dell'Abruzzo e Molise "G. Caporale" | Lorusso A, Marcacci M, Di Domenico M, Ancora M, Curini V, Mangone I, Rinaldi A, Scialabba S, Di Pasquale A, Cammà C, Puglia I, Calistri P, Savini G |
| EPI_ISL_961750 | Ospedale Cvile Atri Med. Interna | Istituto Zooprofilattico Sperimentale dell'Abruzzo e Molise "G. Caporale" | Lorusso A, Marcacci M, Di Domenico M, Ancora M, Curini V, Mangone I, Rinaldi A, Scialabba S, Di Pasquale A, Cammà C, Puglia I, Calistri P, Savini G |
| EPI_ISL_961751 | SIESP DIPARTIMENTO DI PREVENZIONE CHIETI | Istituto Zooprofilattico Sperimentale dell'Abruzzo e Molise "G. Caporale" | Lorusso A, Marcacci M, Di Domenico M, Ancora M, Curini V, Mangone I, Rinaldi A, Scialabba S, Di Pasquale A, Cammà C, Puglia I, Calistri P, Savini G |
| EPI_ISL_961752, EPI_ISL_961753 | SIESP SULMONA | Istituto Zooprofilattico Sperimentale dell'Abruzzo e Molise "G. Caporale" | Lorusso A, Marcacci M, Di Domenico M, Ancora M, Curini V, Mangone I, Rinaldi A, Scialabba S, Di Pasquale A, Cammà C, Puglia I, Calistri P, Savini G |
| EPI_ISL_961754 | Presidio Ospedaliero Sulmona | Istituto Zooprofilattico Sperimentale dell'Abruzzo e Molise "G. Caporale" | Lorusso A, Marcacci M, Di Domenico M, Ancora M, Curini V, Mangone I, Rinaldi A, Scialabba S, Di Pasquale A, Cammà C, Puglia I, Calistri P, Savini G |
| EPI_ISL_961755, EPI_ISL_961756, EPI_ISL_961757, EPI_ISL_961758 | SIESP SULMONA | Istituto Zooprofilattico Sperimentale dell'Abruzzo e Molise "G. Caporale" | Lorusso A, Marcacci M, Di Domenico M, Ancora M, Curini V, Mangone I, Rinaldi A, Scialabba S, Di Pasquale A, Cammà C, Puglia I, Calistri P, Savini G |
| EPI_ISL_965025 | S.C. Microbiologia e Virologia Laboratorio Virologia -Speciale Centro Influenza - AOU di Sassari - Viale san Pietro 43/B Palazzo Infettivologia | Laboratorio specialistico UOC Ematologia - Ospedale "San Francesco" - ATS-ASSL Nuoro | Piras Giovanna, Malune Paolo, Asproni Rosanna, Monne Maria Itria, Palmas Angelo Domenico Serra Caterina, Rimini Elena, Rubino Salvatore |
| EPI_ISL_965028, EPI_ISL_965031, EPI_ISL_965114 | Laboratorio Biologia Molecolare Sars Cov2 - UOC Laboratorio Analisi - Servizio Medicina di Laboratorio, Ospedale "San Francesco" - ATS-ASSL Nuoro | Laboratorio specialistico UOC Ematologia - Ospedale "San Francesco" - ATS-ASSL Nuoro | Piras Giovanna, Asproni Rosanna, Malune Paolo, Fiamma Maura, Monne Maria Itria, Palmas Angelo Domenico, Lo Maglio Iana, Mameli Giuseppe |
| EPI_ISL_965127 | INMI Lazzaro Spallanzani IRCCS | INMI Lazzaro Spallanzani IRCCS | CEM Gruber, B Bartolini, E Giombini, M Rueca, O Butera, F Messina, A Di Caro, MR Capobianchi |

| | | | |
|---|---|---|---|
| EPI_ISL_965128 | INMI Lazzaro Spallanzani IRCCS | INMI Lazzaro Spallanzani IRCCS | M. Rueca, E Giombini, B Bartolini, O Butera, C.E.M Gruber, F Messina, A Di Caro, MR Capobianchi |
| EPI_ISL_965140 | UOC "Microbiologia e Virologia" AOU "Policlinico Umberto I", Roma; Dipartimento di Medicina Molecolare Sapienza Università di Roma | INMI Lazzaro Spallanzani IRCCS | F Messina, C.E.M Gruber, B Bartolini, E Giombini, M Rueca, O Butera, G Antonelli, O Turriziani, MR Capobianchi, A Di Caro |
| EPI_ISL_965223 | SIESP SULMONA | Istituto Zooprofilattico Sperimentale dell'Abruzzo e Molise "G. Caporale" | Lorusso A, Marcacci M, Di Domenico M, Ancora M, Curini V, Mangone I, Rinaldi A, Scialabba S, Di Pasquale A, Cammà C, Puglia I, Calistri P, Savini G |
| EPI_ISL_965224 | OSPEDALE SS. ANNUNZIATA CHIETI | Istituto Zooprofilattico Sperimentale dell'Abruzzo e Molise "G. Caporale" | Lorusso A, Marcacci M, Di Domenico M, Ancora M, Curini V, Mangone I, Rinaldi A, Scialabba S, Di Pasquale A, Cammà C, Puglia I, Calistri P, Savini G |
| EPI_ISL_965225, EPI_ISL_965226 | USCA SULMONA | Istituto Zooprofilattico Sperimentale dell'Abruzzo e Molise "G. Caporale" | Lorusso A, Marcacci M, Di Domenico M, Ancora M, Curini V, Mangone I, Rinaldi A, Scialabba S, Di Pasquale A, Cammà C, Puglia I, Calistri P, Savini G |
| EPI_ISL_965227, EPI_ISL_965228, EPI_ISL_965229 | SIESP CHIETI - DRIVE IN CHIETI | Istituto Zooprofilattico Sperimentale dell'Abruzzo e Molise "G. Caporale" | Lorusso A, Marcacci M, Di Domenico M, Ancora M, Curini V, Mangone I, Rinaldi A, Scialabba S, Di Pasquale A, Cammà C, Puglia I, Calistri P, Savini G |
| EPI_ISL_965230 | SIESP TERAMO DIP PREV | Istituto Zooprofilattico Sperimentale dell'Abruzzo e Molise "G. Caporale" | Lorusso A, Marcacci M, Di Domenico M, Ancora M, Curini V, Mangone I, Rinaldi A, Scialabba S, Di Pasquale A, Cammà C, Puglia I, Calistri P, Savini G |
| EPI_ISL_965231 | Ospedale San Salvatore UOC MED INT. | Istituto Zooprofilattico Sperimentale dell'Abruzzo e Molise "G. Caporale" | Lorusso A, Marcacci M, Di Domenico M, Ancora M, Curini V, Mangone I, Rinaldi A, Scialabba S, Di Pasquale A, Cammà C, Puglia I, Calistri P, Savini G |
| EPI_ISL_965232, EPI_ISL_965233 | SIESP CHIETI - DRIVE IN CHIETI | Istituto Zooprofilattico Sperimentale dell'Abruzzo e Molise "G. Caporale" | Lorusso A, Marcacci M, Di Domenico M, Ancora M, Curini V, Mangone I, Rinaldi A, Scialabba S, Di Pasquale A, Cammà C, Puglia I, Calistri P, Savini G |
| EPI_ISL_965234 | SIESP SULMONA | Istituto Zooprofilattico Sperimentale dell'Abruzzo e Molise "G. Caporale" | Lorusso A, Marcacci M, Di Domenico M, Ancora M, Curini V, Mangone I, Rinaldi A, Scialabba S, Di Pasquale A, Cammà C, Puglia I, Calistri P, Savini G |
| EPI_ISL_965235 | SIESP TERAMO DIP PREV | Istituto Zooprofilattico Sperimentale dell'Abruzzo e Molise "G. Caporale" | Lorusso A, Marcacci M, Di Domenico M, Ancora M, Curini V, Mangone I, Rinaldi A, Scialabba S, Di Pasquale A, Cammà C, Puglia I, Calistri P, Savini G |
| EPI_ISL_965236, EPI_ISL_965237, EPI_ISL_965238, EPI_ISL_965239 | SIESP CHIETI - DRIVE IN CHIETI | Istituto Zooprofilattico Sperimentale dell'Abruzzo e Molise "G. Caporale" | Lorusso A, Marcacci M, Di Domenico M, Ancora M, Curini V, Mangone I, Rinaldi A, Scialabba S, Di Pasquale A, Cammà C, Puglia I, Calistri P, Savini G |
| EPI_ISL_965240 | SIESP CHIETI | Istituto Zooprofilattico Sperimentale dell'Abruzzo e Molise "G. Caporale" | Lorusso A, Marcacci M, Di Domenico M, Ancora M, Curini V, Mangone I, Rinaldi A, Scialabba S, Di Pasquale A, Cammà C, Puglia I, Calistri P, Savini G |
| EPI_ISL_965241, EPI_ISL_965242 | USCA SULMONA | Istituto Zooprofilattico Sperimentale dell'Abruzzo e Molise "G. Caporale" | Lorusso A, Marcacci M, Di Domenico M, Ancora M, Curini V, Mangone I, Rinaldi A, Scialabba S, Di Pasquale A, Cammà C, Puglia I, Calistri P, Savini G |
| EPI_ISL_965243, EPI_ISL_965244 | SIESP CHIETI-DRIVE IN ORTONA | Istituto Zooprofilattico Sperimentale dell'Abruzzo e Molise "G. Caporale" | Lorusso A, Marcacci M, Di Domenico M, Ancora M, Curini V, Mangone I, Rinaldi A, Scialabba S, Di Pasquale A, Cammà C, Puglia I, Calistri P, Savini G |
| EPI_ISL_965245 | OSPEDALE CIVILE TERAMO, DIV. PNEUMOLOGIA, PIA | Istituto Zooprofilattico Sperimentale dell'Abruzzo e Molise "G. Caporale" | Lorusso A, Marcacci M, Di Domenico M, Ancora M, Curini V, Mangone I, Rinaldi A, Scialabba S, Di Pasquale A, Cammà C, Puglia I, Calistri P, Savini G |
| EPI_ISL_965246 | OSP CIV GIULIANOVA, MED INT | Istituto Zooprofilattico Sperimentale dell'Abruzzo e Molise "G. Caporale" | Lorusso A, Marcacci M, Di Domenico M, Ancora M, Curini V, Mangone I, Rinaldi A, Scialabba S, Di Pasquale A, Cammà C, Puglia I, Calistri P, Savini G |
| EPI_ISL_965247, EPI_ISL_965248, EPI_ISL_965249, EPI_ISL_965250, EPI_ISL_965251, EPI_ISL_965252, EPI_ISL_965253 | SIESP CHIETI | Istituto Zooprofilattico Sperimentale dell'Abruzzo e Molise "G. Caporale" | Lorusso A, Marcacci M, Di Domenico M, Ancora M, Curini V, Mangone I, Rinaldi A, Scialabba S, Di Pasquale A, Cammà C, Puglia I, Calistri P, Savini G |
| EPI_ISL_965254, EPI_ISL_965255 | SIESP CHIETI - DRIVE IN CHIETI | Istituto Zooprofilattico Sperimentale dell'Abruzzo e Molise "G. Caporale" | Lorusso A, Marcacci M, Di Domenico M, Ancora M, Curini V, Mangone I, Rinaldi A, Scialabba S, Di Pasquale A, Cammà C, Puglia I, Calistri P, Savini G |
| EPI_ISL_965256 | SIESP CHIETI-DRIVE IN VASTO | Istituto Zooprofilattico Sperimentale dell'Abruzzo e Molise "G. Caporale" | Lorusso A, Marcacci M, Di Domenico M, Ancora M, Curini V, Mangone I, Rinaldi A, Scialabba S, Di Pasquale A, Cammà C, Puglia I, Calistri P, Savini G |
| EPI_ISL_965257 | SIESP CHIETI-DRIVE IN GISSI | Istituto Zooprofilattico Sperimentale dell'Abruzzo e Molise "G. Caporale" | Lorusso A, Marcacci M, Di Domenico M, Ancora M, Curini V, Mangone I, Rinaldi A, Scialabba S, Di Pasquale A, Cammà C, Puglia I, Calistri P, Savini G |
| EPI_ISL_965258, EPI_ISL_965259 | SIESP CHIETI-DRIVE IN VASTO | Istituto Zooprofilattico Sperimentale dell'Abruzzo e Molise "G. Caporale" | Lorusso A, Marcacci M, Di Domenico M, Ancora M, Curini V, Mangone I, Rinaldi A, Scialabba S, Di Pasquale A, Cammà C, Puglia I, Calistri P, Savini G |
| EPI_ISL_965260 | SIESP CHIETI-DRIVE IN GISSI | Istituto Zooprofilattico Sperimentale dell'Abruzzo e Molise "G. Caporale" | Lorusso A, Marcacci M, Di Domenico M, Ancora M, Curini V, Mangone I, Rinaldi A, Scialabba S, Di Pasquale A, Cammà C, Puglia I, Calistri P, Savini G |
| EPI_ISL_965261 | SIESP CHIETI | Istituto Zooprofilattico Sperimentale dell'Abruzzo e Molise "G. Caporale" | Lorusso A, Marcacci M, Di Domenico M, Ancora M, Curini V, Mangone I, Rinaldi A, Scialabba S, Di Pasquale A, Cammà C, Puglia I, Calistri P, Savini G |
| EPI_ISL_965262 | SIESP CHIETI, DISTRETTO SAN CHIETI | Istituto Zooprofilattico Sperimentale dell'Abruzzo e Molise "G. Caporale" | Lorusso A, Marcacci M, Di Domenico M, Ancora M, Curini V, Mangone I, Rinaldi A, Scialabba S, Di Pasquale A, Cammà C, Puglia I, Calistri P, Savini G |
| EPI_ISL_965263 | SIESP CHIETI-DRIVE IN ORTONA | Istituto Zooprofilattico Sperimentale dell'Abruzzo e Molise "G. Caporale" | Lorusso A, Marcacci M, Di Domenico M, Ancora M, Curini V, Mangone I, Rinaldi A, Scialabba S, Di Pasquale A, Cammà C, Puglia I, Calistri P, Savini G |
| EPI_ISL_965265 | SIESP CHIETI - DRIVE IN CHIETI | Istituto Zooprofilattico Sperimentale dell'Abruzzo e Molise "G. Caporale" | Lorusso A, Marcacci M, Di Domenico M, Ancora M, Curini V, Mangone I, Rinaldi A, Scialabba S, Di Pasquale A, Cammà C, Puglia I, Calistri P, Savini G |
| EPI_ISL_965266 | SIESP CHIETI | Istituto Zooprofilattico Sperimentale dell'Abruzzo e Molise "G. Caporale" | Lorusso A, Marcacci M, Di Domenico M, Ancora M, Curini V, Mangone I, Rinaldi A, Scialabba S, Di Pasquale A, Cammà C, Puglia I, Calistri P, Savini G |
| EPI_ISL_965267 | SIESP CHIETI, DISTRETTON SAN CHIETI | Istituto Zooprofilattico Sperimentale dell'Abruzzo e Molise "G. Caporale" | Lorusso A, Marcacci M, Di Domenico M, Ancora M, Curini V, Mangone I, Rinaldi A, Scialabba S, Di Pasquale A, Cammà C, Puglia I, Calistri P, Savini G |
| EPI_ISL_965268, EPI_ISL_965269 | SIESP CHIETI, DISTRETTO SAN CHIETI | Istituto Zooprofilattico Sperimentale dell'Abruzzo e Molise "G. Caporale" | Lorusso A, Marcacci M, Di Domenico M, Ancora M, Curini V, Mangone I, Rinaldi A, Scialabba S, Di Pasquale A, Cammà C, Puglia I, Calistri P, Savini G |
| EPI_ISL_965270 | SIESP CHIETI | Istituto Zooprofilattico Sperimentale dell'Abruzzo e Molise "G. Caporale" | Lorusso A, Marcacci M, Di Domenico M, Ancora M, Curini V, Mangone I, Rinaldi A, Scialabba S, Di Pasquale A, Cammà C, Puglia I, Calistri P, Savini G |
| EPI_ISL_965271 | SIESP CHIETI, DISTRETTO SAN CHIETI | Istituto Zooprofilattico Sperimentale dell'Abruzzo e Molise "G. Caporale" | Lorusso A, Marcacci M, Di Domenico M, Ancora M, Curini V, Mangone I, Rinaldi A, Scialabba S, Di Pasquale A, Cammà C, Puglia I, Calistri P, Savini G |
| EPI_ISL_965272 | SIESP CHIETI | Istituto Zooprofilattico Sperimentale dell'Abruzzo e Molise "G. Caporale" | Lorusso A, Marcacci M, Di Domenico M, Ancora M, Curini V, Mangone I, Rinaldi A, Scialabba S, Di Pasquale A, Cammà C, Puglia I, Calistri P, Savini G |
| EPI_ISL_965273, EPI_ISL_965274, EPI_ISL_965275 | SIESP CHIETI, DISTRETTO SAN CHIETI | Istituto Zooprofilattico Sperimentale dell'Abruzzo e Molise "G. Caporale" | Lorusso A, Marcacci M, Di Domenico M, Ancora M, Curini V, Mangone I, Rinaldi A, Scialabba S, Di Pasquale A, Cammà C, Puglia I, Calistri P, Savini G |

| | | | |
|---|---|---|---|
| EPI_ISL_965276 | OSP CIV ATRI | Istituto Zooprofilattico Sperimentale dell'Abruzzo e Molise "G. Caporale" | Lorusso A, Marcacci M, Di Domenico M, Ancora M, Curini V, Mangone I, Rinaldi A, Scialabba S, Di Pasquale A, Cammà C, Puglia I, Calistri P, Savini G |
| EPI_ISL_965278, EPI_ISL_965279, EPI_ISL_965280, EPI_ISL_965281, EPI_ISL_965282, EPI_ISL_965283, EPI_ISL_965284, EPI_ISL_965285, EPI_ISL_965286, EPI_ISL_965287, EPI_ISL_965288, EPI_ISL_965289, EPI_ISL_965290, EPI_ISL_965291, EPI_ISL_965292, EPI_ISL_965293, EPI_ISL_965294, EPI_ISL_965295, EPI_ISL_965296, EPI_ISL_965297, EPI_ISL_965298, EPI_ISL_965299, EPI_ISL_965300 | | | |
| see above | P.O.CARDARELLI | Istituto Zooprofilattico Sperimentale dell'Abruzzo e Molise "G. Caporale" | Scutellà M, Niro G, Lorusso A, Marcacci M, Di Domenico M, Ancora M, Curini V, Mangone I, Rinaldi A, Scialabba S, Di Pasquale A, Cammà C, Puglia I, Calistri P, Savini G |
| EPI_ISL_969131 | Laboratorio Microbiologia e Virologia P.O. Cotugno A.O. dei Colli | Laboratorio Microbiologia e Virologia P.O. Cotugno A.O. dei Colli | Luigi Atripaldi, Claudia Tiberio, Anna Perfetti, |
| EPI_ISL_969227 | Laboratorio Microbiologia e Virologia P.O. Cotugno A.O. dei Colli | Laboratorio Microbiologia e Virologia P.O. Cotugno A.O. dei Colli | Luigi Atripaldi, Claudia Tiberio, Anna Perfetti |
| EPI_ISL_969297 | Laboratorio Microbiologia e Virologia, P.O. Cotugno, A.O. dei Colli | Laboratorio Microbiologia e Virologia, P.O. Cotugno, A.O. dei Colli | Luigi Atripaldi, Claudia Tiberio, Anna Perfetti |
| EPI_ISL_969884 | Laboratorio Microbiologia e Virologia P.O. Cotugno A.O. dei Colli | Laboratorio Microbiologia e Virologia P.O. Cotugno A.O. dei Colli | Luigi Atripaldi,Claudia Tiberio, Anna Perfetti |
| EPI_ISL_970647 | Laboratorio Microbiologia e Virologia P.O. Cotugno A.O. dei Colli | Laboratorio Microbiologia e Virologia P.O. Cotugno A.O. dei Colli | Luigi Atripaldi, Claudia Tiberio,Anna Perfetti |
| EPI_ISL_974745 | Laboratorio Microbiologia e Virologia P.O. Cotugno A.O. dei Colli | Laboratorio Microbiologia e Virologia P.O. Cotugno A.O. dei Colli | Luigi Atripaldi, Claudia Tiberio, Anna Perfetti, |
| EPI_ISL_977495, EPI_ISL_977496, EPI_ISL_977497, EPI_ISL_977498 | University of Bari Biomedical Sciences and Human Oncology | University of Bari Biomedical Sciences and Human Oncology | Chironna M., Sallustio A., Loconsole D., Accogli M. |
| EPI_ISL_977598 | Laboratorio Microbiologia e Virologia P.O. Cotugno A.O. dei Colli | Laboratorio Microbiologia e Virologia P.O. Cotugno A.O. dei Colli | Luigi Atripaldi, Claudia Tiberio, Anna Perfetti |
| EPI_ISL_977601, EPI_ISL_977603 | Laboratorio Microbiologia e Virologia P.O. Cotugno A.O. dei Colli | Laboratorio Microbiologia e Virologia P.O. Cotugno A.O. dei Colli | Luigi Atripaldi, Claudia Tiberio, Anna Perfetti, |
| EPI_ISL_977604, EPI_ISL_977605, EPI_ISL_977606, EPI_ISL_977607, EPI_ISL_977608, EPI_ISL_977609, EPI_ISL_977610, EPI_ISL_977611 | SC (UCO) Igiene e Sanità Pubblica (funzione integrata con SC Microbiologia e Virologia) e Laboratory of Molecular Virology of the International Centre for Genetic Engineering and Biotechnology (ICGEB) | ARGO Laboratorio Genomica ed Epigenomica | Licastro D, Dal Monego S, Degasperi M, Marcello A, D'Agaro P, De Rosa R |
| EPI_ISL_977612, EPI_ISL_977613, EPI_ISL_977614, EPI_ISL_977615, EPI_ISL_977616, EPI_ISL_977617, EPI_ISL_977618, EPI_ISL_977619, EPI_ISL_977620 | SC (UCO) Igiene e Sanità Pubblica (funzione integrata con SC Microbiologia e Virologia) e Laboratory of Molecular Virology of the International Centre for Genetic Engineering and Biotechnology (ICGEB) | ARGO Laboratorio Genomica ed Epigenomica | Licastro D, Dal Monego S, Degasperi M, Marcello A, D'Agaro P, Pipan C |
| EPI_ISL_977621 | SC (UCO) Igiene e Sanità Pubblica (funzione integrata con SC Microbiologia e Virologia) e Laboratory of Molecular Virology of the International Centre for Genetic Engineering and Biotechnology (ICGEB) | ARGO Laboratorio Genomica ed Epigenomica | Licastro D, Dal Monego S, Degasperi M, Marcello A, D'Agaro P |
| EPI_ISL_977622, EPI_ISL_977623, EPI_ISL_977624, EPI_ISL_977625, EPI_ISL_977626, EPI_ISL_977627, EPI_ISL_977628, EPI_ISL_977629, EPI_ISL_977630, EPI_ISL_977631, EPI_ISL_977632, EPI_ISL_977633, EPI_ISL_977634, EPI_ISL_977635, EPI_ISL_977636, EPI_ISL_977637, EPI_ISL_977638, EPI_ISL_977639, EPI_ISL_977640 | | | |
| see above | SC (UCO) Igiene e Sanità Pubblica (funzione integrata con SC Microbiologia e Virologia) e Laboratory of Molecular Virology of the International Centre for Genetic Engineering and Biotechnology (ICGEB) | ARGO Laboratorio Genomica ed Epigenomica | Licastro D, Dal Monego S, Degasperi M, Marcello A, Segat L, Piscianz E, D'Agaro P |
| EPI_ISL_977641, EPI_ISL_977642, EPI_ISL_977643, EPI_ISL_977644, EPI_ISL_977645, EPI_ISL_977646, EPI_ISL_977647, EPI_ISL_977648, EPI_ISL_977649, EPI_ISL_977650 | SC (UCO) Igiene e Sanità Pubblica (funzione integrata con SC Microbiologia e Virologia) e Laboratory of Molecular Virology of the International Centre for Genetic Engineering and Biotechnology (ICGEB) | ARGO Laboratorio Genomica ed Epigenomica | Licastro D, Dal Monego S, Degasperi M, Marcello A, D'Agaro P, Lombardo F |
| EPI_ISL_977651 | Laboratorio Microbiologia e Virologia P.O. Cotugno A.O. dei Colli | Laboratorio Microbiologia e Virologia P.O. Cotugno A.O. dei Colli | Luigi Atripaldi, Claudia Tiberio, Anna Perfetti, |
| EPI_ISL_983096, EPI_ISL_983097, EPI_ISL_983098 | Microbiology and Virology Unit, Florence Careggi University Hospital | Microbiology and Virology Unit, Florence Careggi University Hospital | Vincenzo Di Pilato, Marco Coppi, Fabio Morecchiato, Noemi Aiezza, Ilaria Baccani, Alberto Antonelli, Emanuele Gori, Gian Maria Rossolini |
| EPI_ISL_983325 | INMI Lazzaro Spallanzani IRCCS | INMI Lazzaro Spallanzani IRCCS | M Rueca, O Butera, F Messina, CEM Gruber, B Bartolini, E Giombini, A Di Caro, MR Capobianchi |
| EPI_ISL_983326 | Laboratorio di Genetica Medica Ospedale Belcolle | INMI Lazzaro Spallanzani IRCCS | F Messina, C.E.M Gruber, B Bartolini, E Giombini, M Rueca, O Butera, F Natoni, G Pessina, A Di Caro, MR Capobianchi |
| EPI_ISL_983327 | Dipartimento di Prevenzione ASL Roma 4 | INMI Lazzaro Spallanzani IRCCS | O Butera, F Messina, CEM Gruber, B Bartolini, E Giombini, M Rueca, S Ursino, MR Capobianchi, A Di Caro |
| EPI_ISL_983328 | Azienda Ospedaliera San Camillo Forlanini | INMI Lazzaro Spallanzani IRCCS | E Giombini, M. Rueca, B Bartolini, O Butera, C.E.M Gruber, F Messina, G Parisi, ML Guarino, A Di Caro, MR Capobianchi |
| EPI_ISL_983329 | Istituto Zooprofilattico Sperimentale Lazio e Toscana "M. Aleandri" | INMI Lazzaro Spallanzani IRCCS | CEM Gruber, B Bartolini, E Giombini, M Rueca, O Butera, F Messina, MT Scicluna, G Manna, A Cersini, A Di Caro, MR Capobianchi |
| EPI_ISL_984995, EPI_ISL_984996 | U.O. Igiene, Ospedale Policlinico San Martino | U.O. Igiene, Ospedale Policlinico San Martino | Bruzzone Bianca, Caligiuri Patrizia, De Pace Vanessa, Domnich Alexander, Orsi Andrea, Ricucci Valentina, Icardi Giancarlo |
| EPI_ISL_984997, EPI_ISL_984998, EPI_ISL_984999 | S.C. Laboratorio Analisi, ASL 3 Liguria | U.O. Igiene, Ospedale Policlinico San Martino | Bruzzone Bianca, Caligiuri Patrizia, De Pace Vanessa, Domnich Alexander, Orsi Andrea, Ricucci Valentina, Spitaleri Antonino, Icardi Giancarlo |
| EPI_ISL_985000 | U.O. Igiene, Ospedale Policlinico San Martino | U.O. Igiene, Ospedale Policlinico San Martino | Bruzzone Bianca, Caligiuri Patrizia, De Pace Vanessa, Domnich Alexander, Orsi Andrea, Ricucci Valentina, Spitaleri Antonino, Icardi Giancarlo |
| EPI_ISL_985001, EPI_ISL_985002, EPI_ISL_985003, EPI_ISL_985004, EPI_ISL_985005, EPI_ISL_985006, EPI_ISL_985007, EPI_ISL_985008, EPI_ISL_985009, EPI_ISL_985010, EPI_ISL_985011, EPI_ISL_985012, EPI_ISL_985013, EPI_ISL_985014, EPI_ISL_985015, EPI_ISL_985016 | | | |
| see above | S.C. Laboratorio Analisi, ASL 3 Liguria | U.O. Igiene, Ospedale Policlinico San Martino | Bruzzone Bianca, Caligiuri Patrizia, De Pace Vanessa, Domnich Alexander, Orsi Andrea, Ricucci Valentina, Spitaleri Antonino, Icardi Giancarlo |
| EPI_ISL_985017 | S.C. Patologia Clinica, Ospedale Sant'Andrea, ASL 5 | U.O. Igiene, Ospedale Policlinico San Martino | Bruzzone Bianca, Battolla Enrico, Caligiuri Patrizia, De Pace Vanessa, Domnich Alexander, Orsi Andrea, Ricucci Valentina, Icardi Giancarlo |
| EPI_ISL_985018, EPI_ISL_985019, EPI_ISL_985020, EPI_ISL_985021, EPI_ISL_985022, EPI_ISL_985023, EPI_ISL_985024 | S.S.D. Microbiologia, Stabilimento ospedaliero di Sanremo, ASL 1 Liguria | U.O. Igiene, Ospedale Policlinico San Martino | Bruzzone Bianca, Caligiuri Patrizia, De Pace Vanessa, Domnich Alexander, Dusi Pier Andrea, Orsi Andrea, Ricucci Valentina, Icardi Giancarlo |
| EPI_ISL_985025, EPI_ISL_985026 | S.S.D. Microbiologia, Ospedale Santa Corona di Pietra Ligure, ASL 2 Liguria | U.O. Igiene, Ospedale Policlinico San Martino | Bruzzone Bianca, Caligiuri Patrizia, De Pace Vanessa, Domnich Alexander, Orsi Andrea, Ricucci Valentina, Valle Caterina, Icardi Giancarlo |

| | | | |
|---|---|---|---|
| EPI_ISL_985027, EPI_ISL_985028, EPI_ISL_985029, EPI_ISL_985030, EPI_ISL_985031 | Laboratorio di Patologia Clinica, Ospedale San Paolo in Valloria, ASL 2 Liguria | U.O. Igiene, Ospedale Policlinico San Martino | Bruzzone Bianca, Caligiuri Patrizia, De Pace Vanessa, Domnich Alexander, Lillo Flavia, Orsi Andrea, Ricucci Valentina, Icardi Giancarlo |
| EPI_ISL_985032 | U.O. Igiene, Ospedale Policlinico San Martino | U.O. Igiene, Ospedale Policlinico San Martino | Bruzzone Bianca, Caligiuri Patrizia, De Pace Vanessa, Domnich Alexander, Orsi Andrea, Ricucci Valentina, Icardi Giancarlo |
